# Identification and evaluation of 41 risk loci for juvenile idiopathic arthritis informs precision medicine: mechanistic implications of DNA topology and an HLA-A*02:01-ERAP2 interaction

**DOI:** 10.64898/2026.09.16.26363161

**Authors:** Hannah C. Ainsworth, Ekaterina S. Khvatkova, Marc Sudman, Miranda C. Marion, Kaiyu Jiang, John F. Bohnsack, Mary E. Comeau, Patrick M. Gaffney, Johannes-Peter Haas, Timothy D. Howard, Kimme Hyrich, Judith A. James, Peter A. Nigrovic, Ellen B. Nordal, Alan M. Rosenberg, Marite Rygg, Samantha L. Smith, Vibeke Videm, Lucy R. Wedderburn, Carol A. Wise, Rae S.M. Yeung, Susan D. Thompson, Grant S. Schulert, Sampath Prahalad, John Bowes, Sheila T. Angeles-Han, James N. Jarvis, Carl D. Langefeld

## Abstract

Juvenile idiopathic arthritis (JIA), the most common class of pediatric rheumatic diseases, can lead to joint damage and extra-articular features including uveitis. Early treatment can improve functional outcomes; but many patients do not respond to first-line treatments. We present a JIA Immunochip-based association analysis of rheumatoid factor-negative polyarticular JIA and oligoarticular JIA. This study spans 3,939 cases and 14,412 controls, including 1,123 cases and 1,356 controls not previously analyzed. We identify 41 (14 novel) JIA risk regions and report a JIA polygenic risk score that also associates with age of onset. Functional mechanisms presented include: a novel interaction between HLA-A*02:01 and *ERAP2*, cluster-based approach for functional amino acid discovery within HLA-DRB1, and sequence-dependent DNA topology for variant prioritization. We present a comprehensive list of curated gene-drug/small molecule interactions for clinically focused studies. These findings provide a deepened understanding of the genetics of JIA and potential mechanisms to inform precision medicine.

## INTRODUCTION

Juvenile Idiopathic Arthritis (JIA), the most common class of childhood rheumatic diseases^1^, comprises a heterogenous group of chronic arthritides and has an estimated pooled prevalence of 20.5 per 100,000, with wide regional variation^2^. JIA has the potential for long-term joint damage, functional impairment, and the development of extra-articular complications such as uveitis^1,3,4^. Rheumatoid factor (RF)-negative polyarticular JIA and oligoarticular JIA, are the two most common categories (herein, denoted ‘subtypes’), exhibit an age and sex bias (female:male=3:1)^5^, and are the focus of this manuscript. JIA has a strong genetic component with a sibling risk ratio of λS=11.6 (95% Confidence Interval: 4.9-27.5)^6^. Although previous works have identified multiple risk loci, including within the major histocompatibility complex (MHC) region^7–10^, significant heritability remains unexplained. Further, even with existing treatments, identifying effective therapeutics for patients remains challenging and emphasizes the need for additional pharmacological targets^11,12^.

Given the strong heritability, heterogenous presentation, and varied treatment response, JIA is well-poised for a largely untapped genetics-based precision medicine approach. Precision medicine initiatives require genetic-based mechanisms, not just identification of genetic risk^13^. Here, we present a significantly expanded JIA genetic association study, featuring 1,123 cases not previously published and report 41 regions of association (14 novel); but critically, we also present results beyond cataloging risk variants by exploring specific mechanisms of disease. One of the novel approaches is the use of sequence-dependent DNA topology (or DNA shape). Despite the established role of DNA shape features (e.g., minor groove width and propeller twist) in molecular interactions^14–18^, its application within the context of genetic association studies is limited in number and scope.^19–21^ Using DNA shape, we present a novel metric to predict allele-specific transcription factor binding and we highlight how this functional annotation can overcome challenges posed by linkage disequilibrium while providing functional information at the nucleotide level (e.g., as opposed to region-based annotations). We also present the first evidence of an interaction between the aminopeptidase *ERAP2* and an HLA-A Class I allele, highlighting the necessity of HLA-A genotyping for *ERAP2* functional studies. These and other presented findings provide meaningful insights into genetic risk for JIA and supportive evidence for interrogation of specific hypotheses within a clinical (e.g., drug-repositioning) lens.

## RESULTS

### Demographics and samples

In total, 3,939 JIA cases and 14,412 controls passed genotype quality control, including 1,123 cases and 1,356 controls of European ancestry not previously published (herein, denoted “Phase 3 data”)^9,10^. Across autosomes, 118,525 SNPs passed quality control metrics. Based on population structure analyses, three admixture variables were included in all analyses (**Figure S1**), resulting in a scaled inflation factor of λ1000=1.04. Of the 3,939 JIA cases, 1,525 (38.72%) were RF-polyarticular cases and 2,414 (61.28%) were oligoarticular cases; 75.1% were female (**Table S1**). Age of Onset (AOO) was available for 3,000 cases with an average and standard deviation of 5.27 ± 4.02 years.

### JIA SNP associations

Across regions of strong JIA association (**Figure 1a**), a majority (61%) featured multiple independent signals, as identified by stepwise logistic regression procedure or Bayesian fine-mapping via SuSiE^22^ (**Figure 1b**). As such, a multi-SNP likelihood ratio test (RegionalLRT) was computed using the independently associated SNPs within each region. Genomic regions are categorized as Tier 1 (T1; P<5x10^-8^) or Tier 2 (T2; P<1x10^-6^) based on either the single-SNP association or the RegionalLRT (i.e., multiple independent signals). Thirty regions met T1 and eleven met T2 status (**Table 1; Figure S2; Tables S2-S3**). Tier 3 (T3) SNPs met PFDR<0.05 (**Table S4)**. Across T1 and T2 regions, 14 (34%) are novel to JIA with genome-wide level evidence (P<1x10^-6^). Ten of these novel regions were identified via the RegionalLRT, highlighting the value of multi-SNP modeling. These regions included: *CD28-CTLA4, IL7, TNFSF15-TNFSF8, ZNF365-EGR2, SMPD3-ZFP90-CHD3, NOS2-LYRM9, EIF2-DYRK3*, *PRDM1*, *IKZF4-ERBB3*, *IFNG-AS1-IFNG.* The remaining four novel regions, *FASLG*-*TNFSF18*, *TRAFD1*-*PTPN11*, *RAC2*, and *CNNM4* exhibited a single SNP association signal. Compared to non-significant (NS) SNPs (P>0.5), T1 and T2 SNPs exhibited greater overlap with ENCODE-defined credible cis-regulatory elements (cCREs)^23^ and were significantly enriched for cCREs in blood (**Figure 1c; Table S5**). The majority of T1 and T2 regions (n=23) contained eQTLs in either a gene expression dataset of JIA cases and controls^24^ or a subset of GTEx^25^ tissues prioritized based on biological relevance to JIA (EBV-transformed lymphocytes, cultured fibroblasts, whole blood, and spleen) **(Figure 1c; Figure S3; Table S6)**, highlighting the regulatory role of identified regions. To evaluate the potential role of transcription factor binding sites (TFBS), T1 and T2 SNPs were queried against a curated database of combined evidence from JASPAR^26^ predicted TFBS and matched ChIP-seq data from REMAP^27^ (see METHODS). In total, 83 high-confidence TFBS overlapped 49 SNPs, spanning 18 genomic regions (**Table S7**). The majority of the associated SNPs mapped to non-coding regions (**Table S8)**. T1 and T2 SNPs were enriched for higher regulatory rankings (ranks 1 and 2) via RegulomeDb^28^ (T1 and T2= 71.8%; NS=52.6%; P=3.00x10^-26^) (**Table S9**). Many of the T1 and T2 regions have been previously implicated in autoimmune diseases^23^ (**Table S10; Figure S4**), such as rheumatoid arthritis, inflammatory bowel disease, Crohn’s Disease, multiple sclerosis, and systemic lupus erythematosus (SLE).

**Figure 1:**
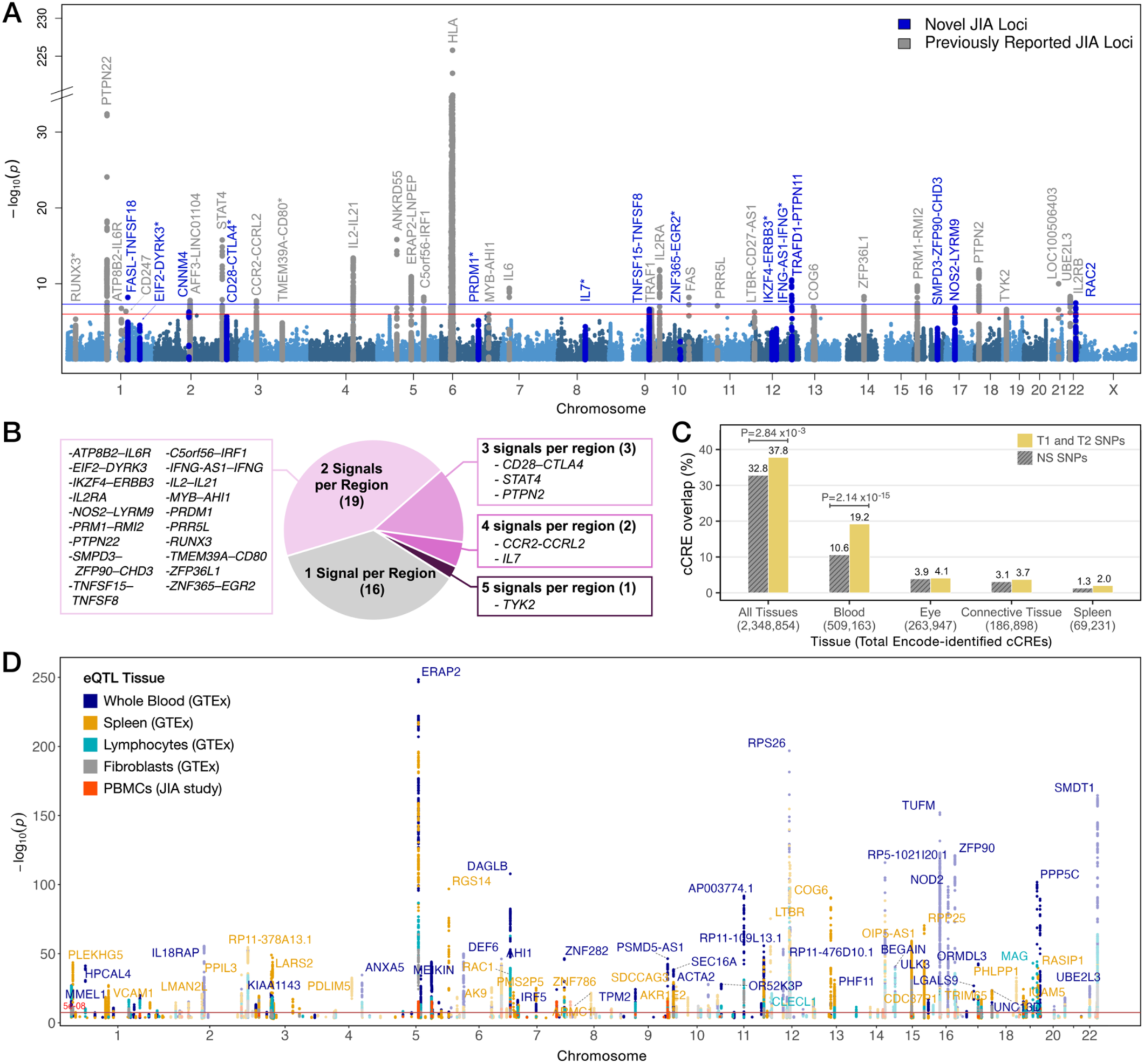
Genome-wide associations with juvenile idiopathic arthritis (JIA). **A)** Manhattan plot of SNP associations. Loci identified by single-SNP associations or the RegionalLRT are labeled. Tier 1 (T1: P<5x10^-8^) and Tier 2 (T2: P<1x10^-6^) significance thresholds are denoted by blue and red horizontal lines, respectively. Novel JIA genome-wide associations are highlighted in blue with an asterisk. **B)** Over 60 percent of associated regions (P<1x10^-6^) exhibited multiple independent SNP signals. The number of independent signals is denoted in parenthesis in the pie chart. **C)** JIA-associated SNPs (T1, T2, or selected by the stepwise procedure within T1 or T2 regions n=837 SNPs) show increased overlap with ENCODE credible cis regulatory elements (cCREs) compared to non-significant SNPs (P>0.5; n= 44,075). Tissue-specific cCREs were selected based on tissue relevance to JIA and are ordered by the total number of cCREs detected by ENCODE. **D)** Manhattan plot of eQTL associations among JIA T1 and T2 SNPs. Data are from GTEx and a published JIA study^24^. GTEx tissues were selected based on disease relevance. Genes associated with the eQTL (eGene) are labeled next to the most significant eQTL for that eGENE.

**Table 1.**
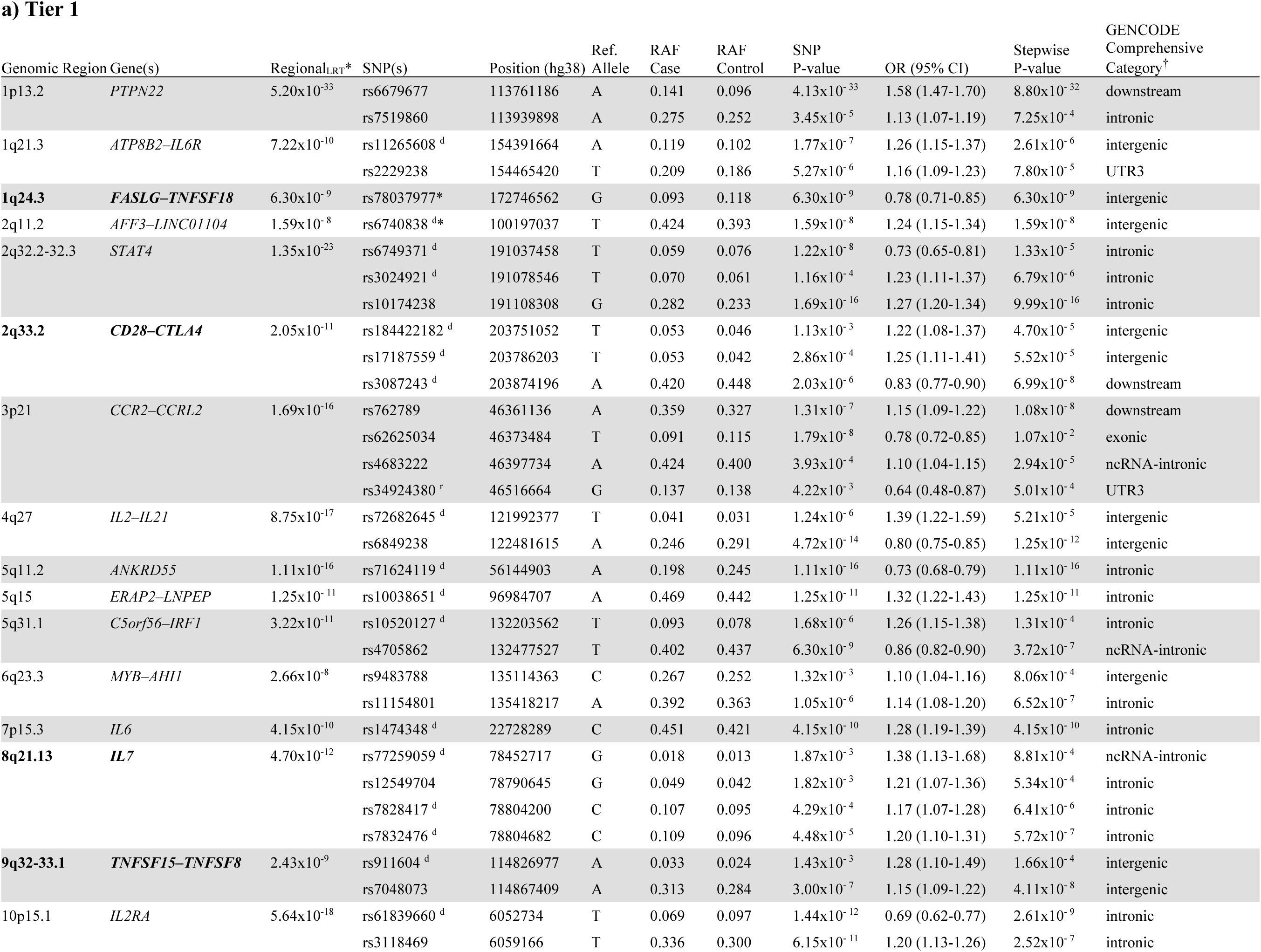
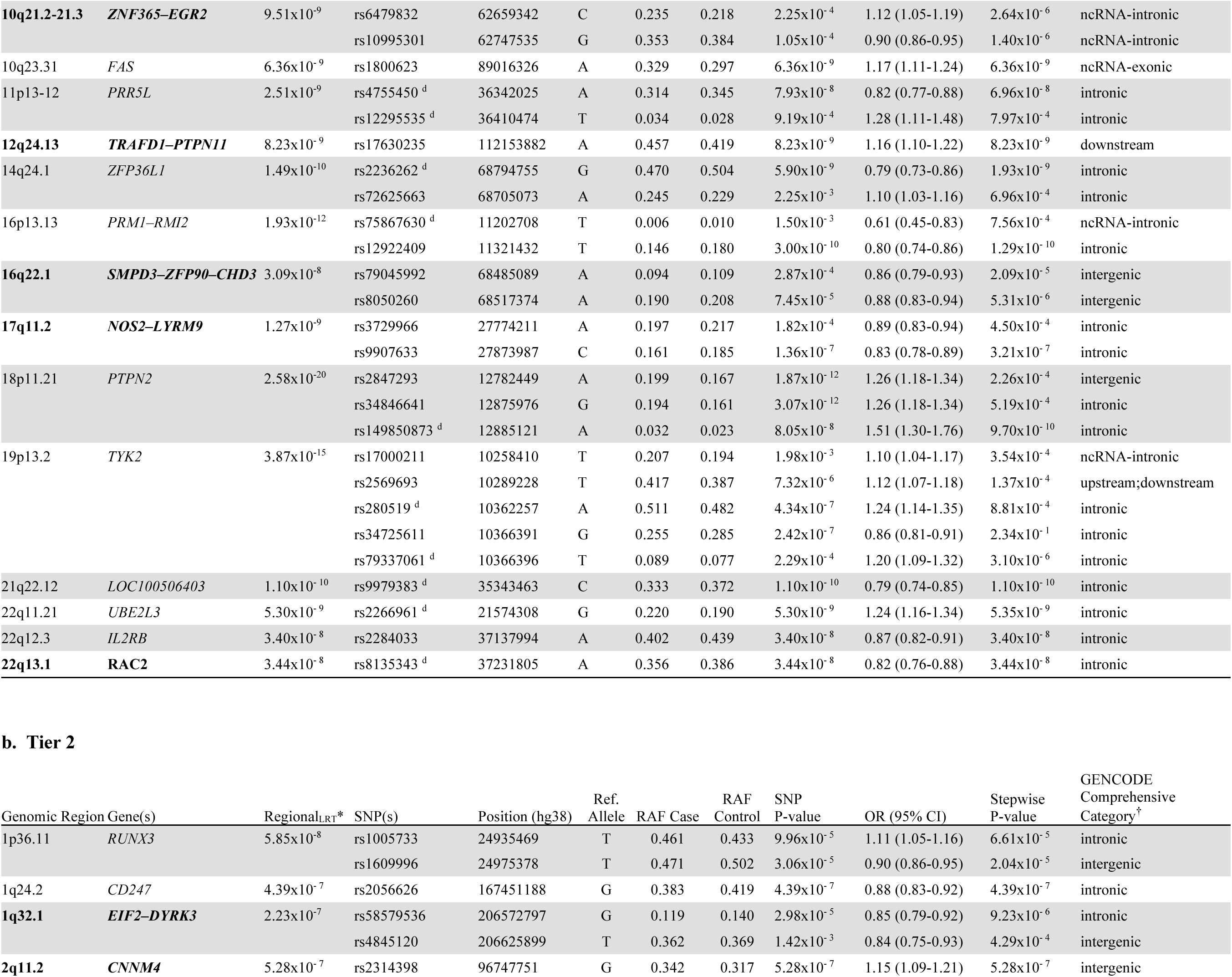
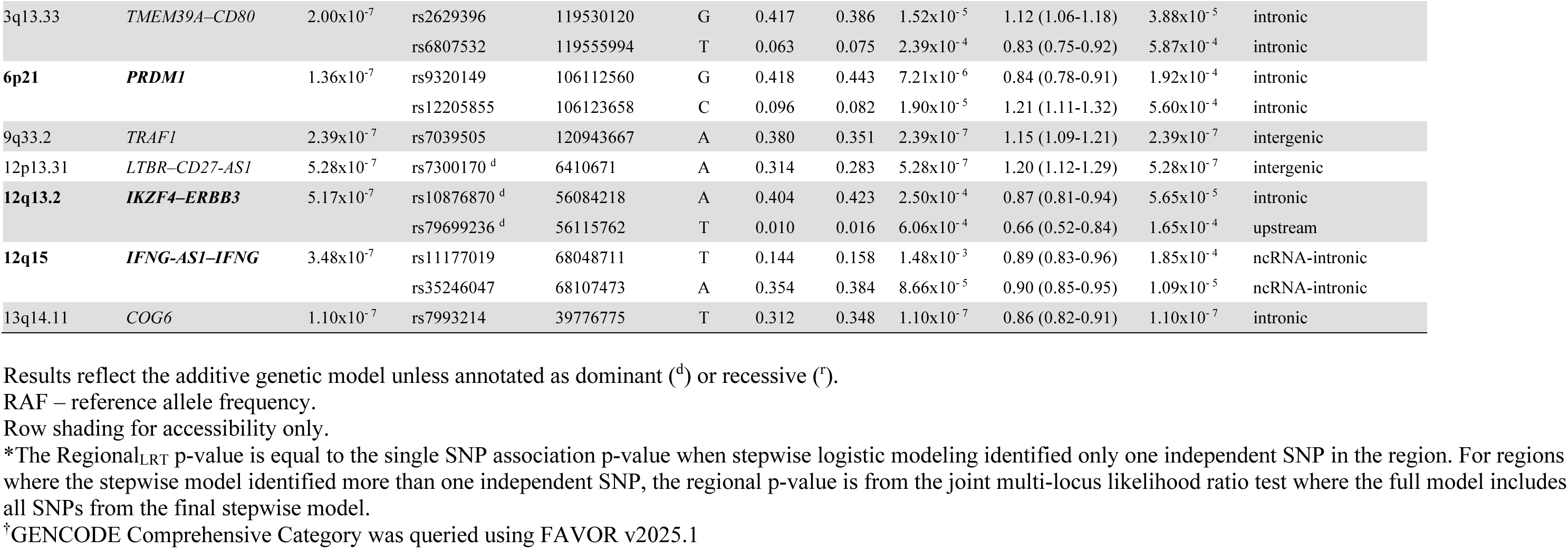
Genomic regions associated with JIA at the Tier 1 and Tier 2 significance at either individual SNP or regional levels (novel regions are bolded).

### Human Leukocyte Antigen (HLA) Associations

MHC Class I (HLA-A, -B, -C) and Class II (HLA-DPB1, -DQA1, -DQB1, -DRB1) two-field alleles were imputed (median posterior probabilities >0.90; **Table S11**) and tested for association (**Table S12; Figure S5**). Stepwise logistic regression identified 18 two-field alleles independently associated with JIA (2 Class I; 16 Class II). These alleles accounted for the majority of evidence of associations in the MHC region (**Figure S6**). The final multilocus HLA model had an area under the ROC curve of c=0.763 (**Table S13**).

Of the 18 alleles in the multilocus model, 10 were from DRB1, suggesting that multiple DRB1 alleles confer JIA risk. To determine if these DRB1 alleles captured distinct or overlapping biological profiles, amino acid sequence similarity and clustering was completed across all (risk and non-risk) two-field DRB1 alleles that passed quality control metrics (**Figure 2a**). The approach was also applied to the other imputed HLA genes (**Figure S7**). In DRB1, sequence similarity yields two primary clusters^29^, herein labeled Cluster 1 and Cluster 2. Annotation of odds ratios from single-allele analyses revealed that Cluster 2 is predominantly comprised of risk alleles, spanning DRB1*11, DRB1*08 and DRB1*13. Only three amino acid positions: 10, 12, and 149 (**Figure 2b; Figure S8**) perfectly differentiate between the two clusters. Positions 10 and 12 are encoded by exon 2 and are within the DRB1 peptide binding region. While Cluster 1 features a polar glutamine (position 10) and positively-charged lysine (position 12), Cluster 2 (JIA risk cluster) features a hydrophobic tyrosine and a polar threonine at these two positions, respectively. Such dissimilar substitutions likely translate to distinct binding profiles between these two clusters. At the third position (149, encoded by exon 3), another dissimilar substitution is found: Cluster 1 features a polar glutamine while the risk-cluster features a positively charged histidine.

**Figure 2:**
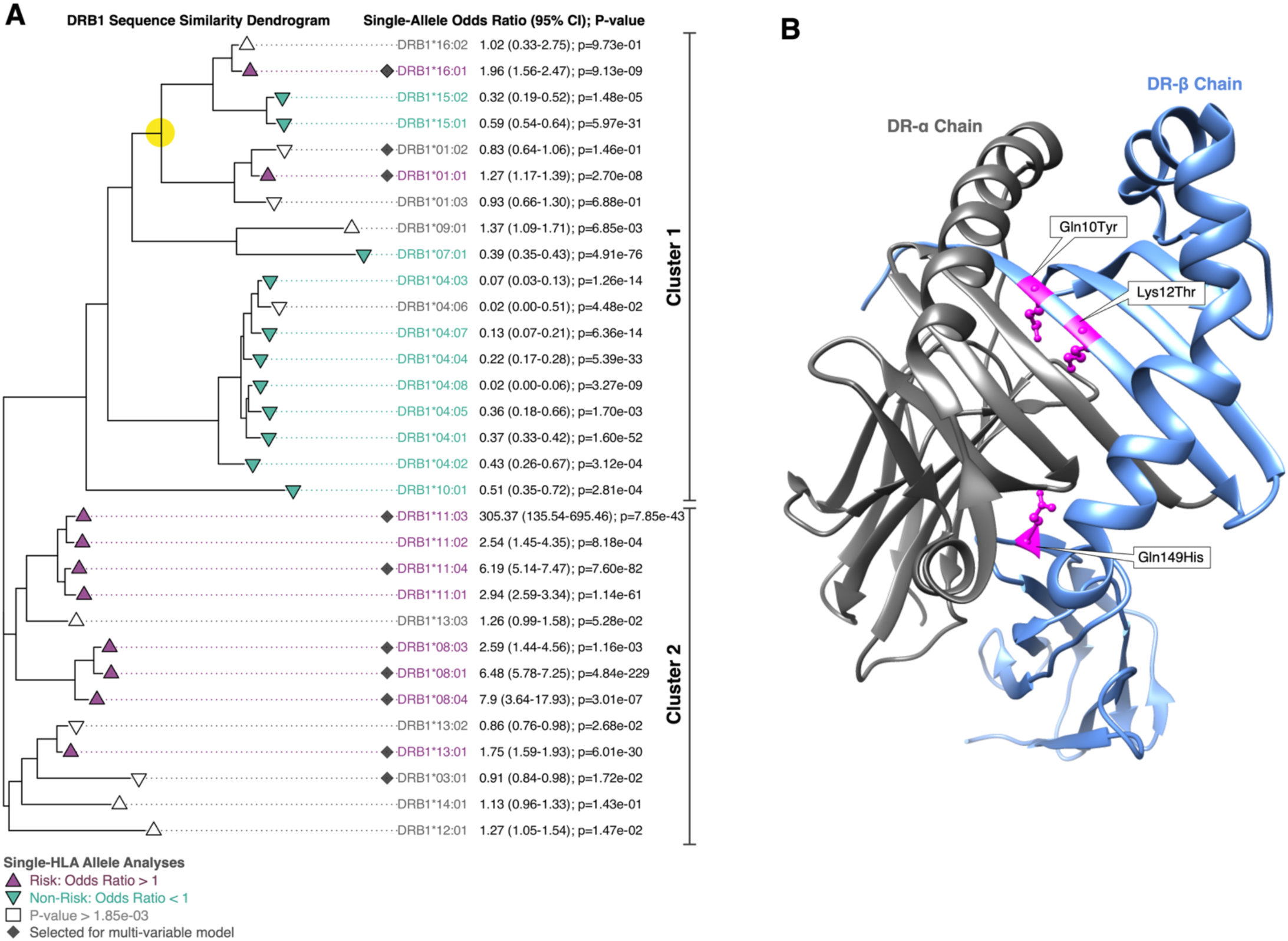
Sequence similarity of DRB1 alleles highlights JIA-risk cluster with distinct amino acid profile within the peptide binding region. **A)** Amino acid sequence similarity of all analyzed DRB1 alleles distinguishes two primary clusters (Cluster 1 and Cluster 2). Notably, overlay with JIA risk from single-allele analyses reveals that Cluster 2 is largely comprised of risk alleles, spanning DRB1*11, DRB1*08, and DRB1*13. Three amino acid substitutions consistently differentiate alleles between Cluster 1 and Cluster 2: Gln10Tyr, Lys12Thr, and Gln149His (**Figure S8**). The two risk alleles in Cluster 1 (DRB1*16:01 and DRB1*01:01) have 95% sequence similarity (node highlighted in yellow) with JIA non-risk with DRB1*15:01 and DRB1*15:02. Sequence similarity across this subcluster identifies 4 amino acids (positions -1, 47, 67, and 71) differing between these risk and non-risk alleles (**Figure S9**). **B)** Structural positioning of the three amino acids differentiating Clusters 1 and 2 are illustrated using PDB:5LAX (DRB1*04:01) as the model. Two amino acids changes (Gln10Tyr) and Lys12Thr) reside within peptide binding region (exon 2); Gln149His (exon 3) resides within an extracellular structure.

Cluster 1 contained only two associated risk alleles, DRB1*16:01 and DRB*01:01. These alleles exhibited 95% similarity with non-risk alleles DRB1*15:01 and DRB1*15:02. Here, four amino acid positions differed between these risk and non-risk alleles (**Figure S9)**. One position is within the leader peptide which is cleaved during protein folding. The remaining three positions, 47, 67, and 71 (exon 3), are within the binding region and are in physical proximity within the folded protein. These substitutions (Phe47Tyr; Ile67Leu/Ile67Phe; Ala71Thr) may not preserve the physico-chemical properties of the protein and thus impact peptide presentation. Having risk alleles from both Clusters 1 and 2 did not exhibit greater than additive risk.

For AOO, only SNPs within the HLA region met genome-wide significance (**Figure S10**), corresponding to Class I and Class II HLA associations (**Tables S14**). Identified HLA alleles were largely consistent with the JIA analyses, albeit fewer alleles reached significance (nAOO=39; nJIA=86); this was also reflected in the multilocus model which included eight alleles (**Table S15**).

### JIA Interactions

A genome-wide association scan for SNP-by-HLA interactions was computed using a case-only analysis^30^. Across the 18 HLA alleles selected in the multilocus model, only a single interaction signal surpassed T2 significance: *ERAP2-LNPEP* with A*02:01 (Padd=6.56x10^-7^) (**Figure S11**). Because *ERAP2-LNPEP* was strongly associated within the JIA case-control analysis (lead SNP: rs10038651; Pdom= 1.25x10^-11^; ORdom=1.32), the genomic region was re-analyzed, stratified by the presence of A*02:01. The original association signal of rs10038651 was enhanced with A*02:01 and absent without A*02:01 (**Figure 3a,b**; A*02:01^+^ rs10038651 Padd=3.93x10^-12^; ORadd= 1.26; A*02:01^-^Padd=0.68, OR = 0.98). SNP rs10038651 is a strong eQTL for *ERAP2* in multiple tissues (**Figure 3c**); the JIA-risk allele is associated with increased *ERAP2* expression, residing on the haplotype which encodes the non-truncated ERAP2 protein and implicated across autoimmune diseases^31^.

**Figure 3:**
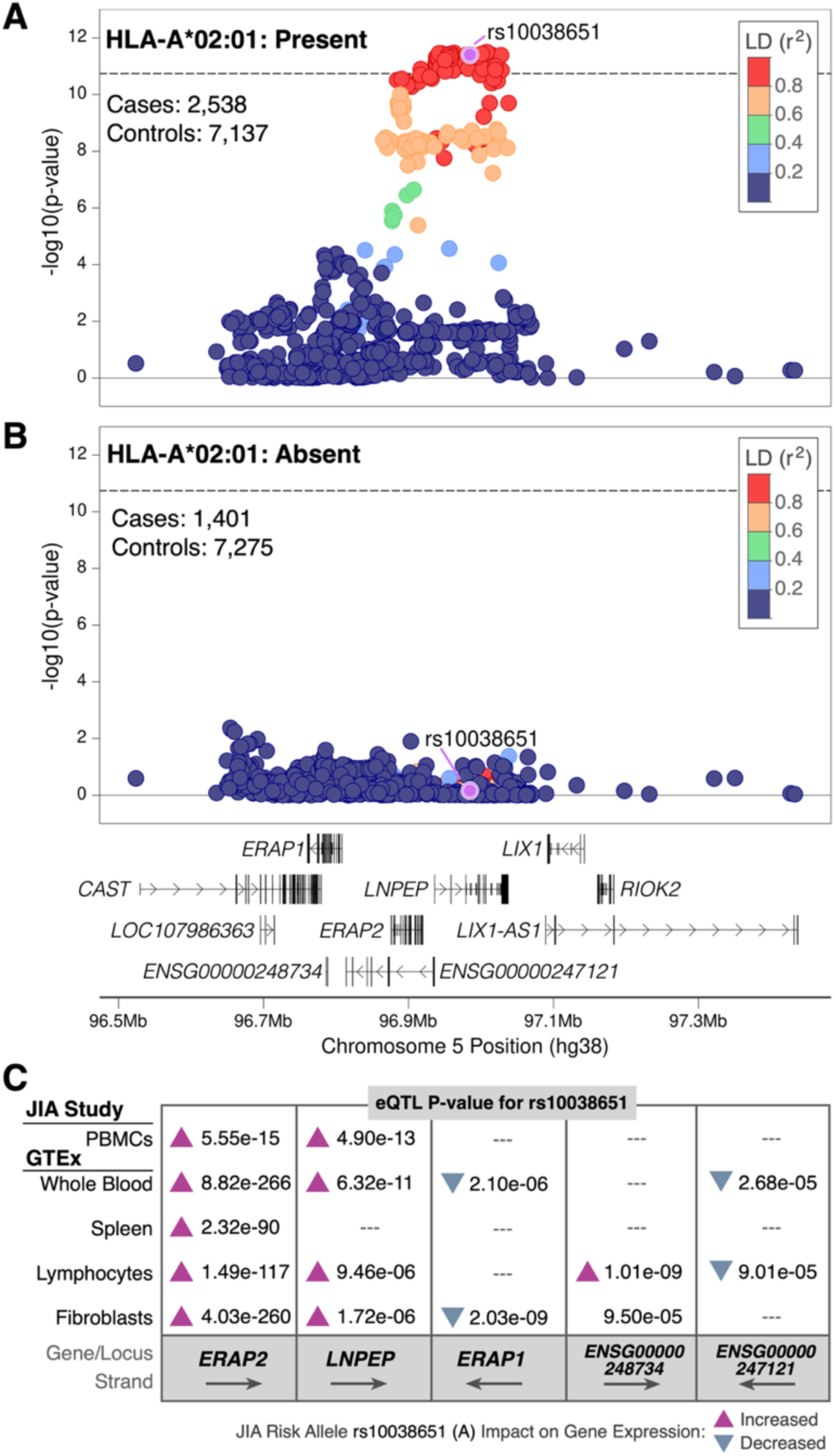
JIA SNP associations in ERAP2-LNPEP region are dependent on presence of HLA-A*02:01. When restricting the association analysis to subjects having at least one copy of HLA-A*02:01 (2,538 cases, 7,137 controls), SNP associations in the region become stronger than the original association signal, agnostic to A*02:01 status (dashed line). Top SNP rs10038651 from the non-stratified analysis is highlighted. **B)** In subjects without HLA-A*02:01 (1,401 cases, 7,275 controls), the region no longer exhibits JIA associations. **C)** eQTLs in disease-relevant tissues using rs10038651 as a proxy for the JIA-association signal. *ERAP2* exhibits the largest eQTL signal with the risk allele (A) associated with increased expression.

Outside of the HLA, there were no significant SNP-by-SNP interactions meeting genome-wide multiple-comparisons thresholds.

### Intrinsic DNA Topology

We hypothesized that a subset of causal SNPs are functional due to their impact on intrinsic DNA topology. We tested whether changes in minor groove width (ΔMGW) and propeller twist (ΔProT) differed between highest-associated SNPs (T1) and non-significant (NS P>0.5) SNPs, excluding the HLA region (**Table S16**). T1 SNPs showed a greater mean±SD ΔMGW 0.71±0.44 compared to NS SNPs 0.65±0.38 (p=0.0016); we note that for bi-allelic SNPs across the genome reported in dbSNP150 (n=199,022,898), the mean^19^ was 0.68±0.43. ΔProT trended towards greater values for T1 SNPs (mean±SD=5.67±3.33) compared to 5.39±3.16 for NS SNPs (P=0.048); this compares to a mean of 5.53±3.14 across bi-allelic SNPs in the genome. These patterns held across minor allele frequencies (MAF) (**Table S17**). Quantile regression also identified a shift towards larger ΔMGW but less for ΔProT (**Table S17**). Because DNA shape is computed within a rolling window, ΔMGW and ΔProT can differentiate among SNPs in linkage disequilibrium (LD). For instance, within *STAT4*, ΔMGW and ΔProT shift prioritization from rs10174238 (lead SNP) to rs11889341 (**Figure S12**) which was among the largest ‘shape disrupting SNPs’ (**Table S16**) and has been identified as a putative causal variant in SLE^32^.

Transcription factor (TF) binding site sequences are highly conserved and changes in native DNA topology can impact TF binding^15,18^. Thus, we hypothesized that even modest changes in DNA topology might alter TF binding. We computed SNP-TFBS scores for MGW and ProT for 46 JIA-associated SNP-TFBS combinations, corresponding to 30 SNPs that overlapped high-confidence TFBS for 36 TFs with experimental binding data (see METHODS; **Figure S13; Table S18**). Compared to all T1 and T2 SNPs, this prioritized subset exhibited greater overlap with cCREs (66.7% versus 37.8%) and was enriched for higher ranks (ranks 1 or 2) by RegulomeDb (96.7% vs 71.8%). While cCRE overlap and highly-ranked RegulomeDb scores support the functional relevance of the genomic coordinates for these 30 SNPs, our TFBS-SNP scores aimed to predict the presence of allele-specific effects (unhindered by LD), via TF binding. Larger TFBS-SNP scores correspond to stronger predictions of allele-specific events. Isoclines based on an L2-norm were computed to facilitate comparison across TFs (**Table S19**).

Globally, there was an inverse trend between TFBS-SNP scores and MAF (rSpearman’s = - 0.36; P=0.053) (**Figure S14**). Approximately half (24/46) of analyzed (**Figure S15)** SNP-TFBS sites were predicted to have allele-specific effects (Isoclines 2-6), positing these SNPs as potentially functional (**Figure 4a**). Across these 24 SNP-TFBS combinations, the JIA risk allele was predicted to facilitate binding at 13 sites and disrupt binding at 11. Among the highest prioritized SNP-TFBS combinations was an intronic *PTPN2* SNP (rs7234029) overlapping a GATA1 (MA0035.4) binding site. The JIA-risk allele, C, showed disruption for both MGW and ProT (**Figure 4b**). This SNP has been previously implicated across autoimmune diseases and *in silico* predictions suggesting potential TFBS effects^33^. We further evaluated GATA1 binding in K562 cells using oligos based on rs7234029. Results from the DNA pulldown assay corroborated our TF-binding prediction, showing decreased GATA1 binding for the risk allele (**Figure 4c; Figure S16a**). Other TF-SNP combinations in the highest prioritized isocline 6 included MZF1-(rs79699236) *IKZ4-ERBB3,* which is a novel JIA locus; CDX2-(rs2358995) *PTPN22;* and ZEB1-(rs75867630) *PRM1-RMI2*. Interestingly, ZEB1, for which the JIA-risk allele is predicted to facilitate ZEB1 binding, has several drug-gene interactions, including Oxaliplatin which has published evidence for ameliorating synovial inflammation^34^.

**Figure 4.**
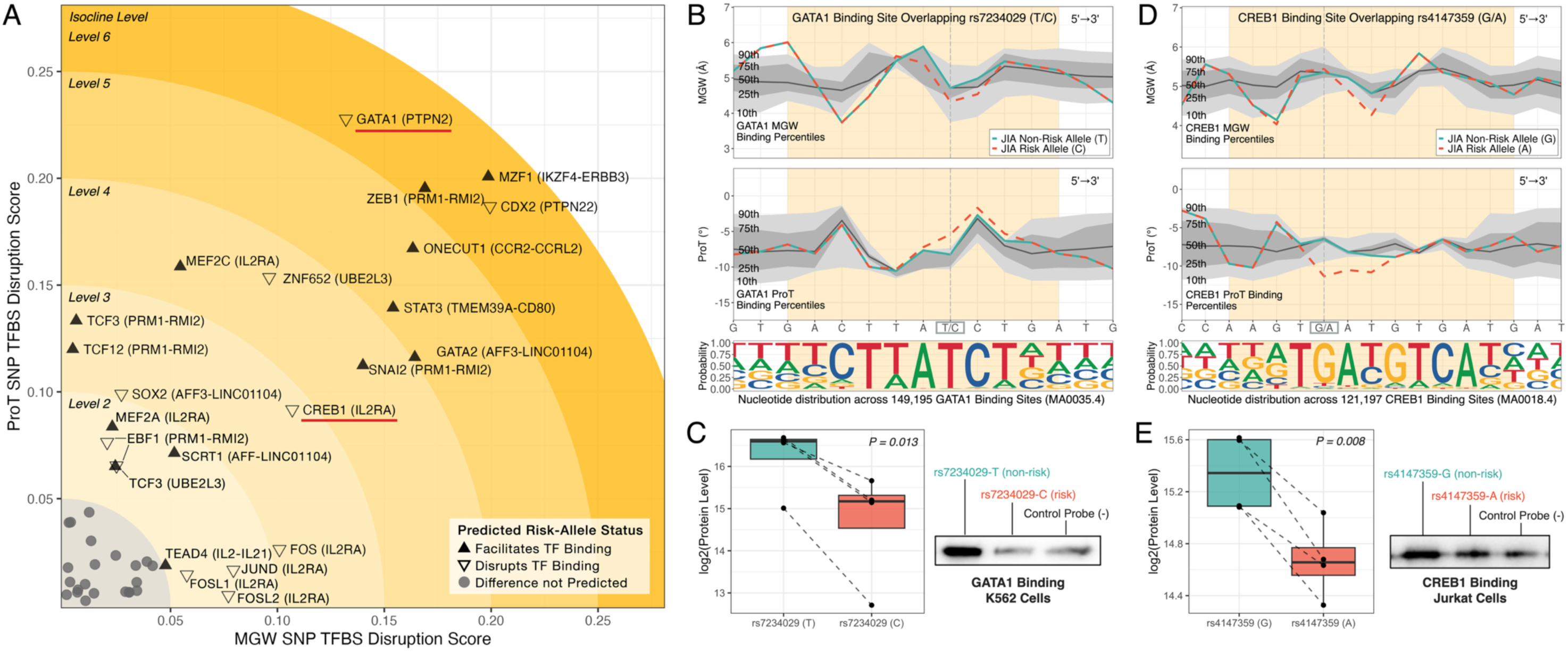
Predicted impact of sequence-dependent DNA topology on transcription factor binding among 30 JIA-associated SNPs. **A:** Predicted allele-specific TF binding effects across 46 SNP-TFBS combinations. Larger disruption scores, based on MGW (x-axis) and ProT (y-axis), correspond to stronger predictions of allele-specific effects. Points are labeled by the TF and the JIA-associated region (based on proximal location) in parenthesis. Relative to the non-risk allele, open downward triangles indicate that the risk allele is predicted to disrupt binding, while solid upward triangles indicate the risk allele is predicted to facilitate binding. Gray circles within isocline level 1 were not predicted to have allele-specific effects. TFs with red underline are highlighted in parts **B-E**. **B** and **D:** Comparison of TF topological profiles with allelic profiles at a SNP of interest: (**B**) GATA1-rs7234029. (**D**) CREB1-rs4147359. MGW and ProT TF topological profiles are based on JASPAR 2024 experimental binding motifs and are visualized as percentiles (10th / 90^th^: light gray regions; 25th / 75^th^: medium gray regions; 50^th^: dark gray line). The predicted TFBS is shown in yellow, overlapping the SNP of interest (vertical dotted line). Topological profiles of the risk (orange dashed line) and non-risk (solid blue line) alleles are also plotted. For both rs7234029 and rs4147359, the risk alleles exhibit topological profiles that deviate even beyond the 10^th^-90^th^ TF percentile range, yielding a predicted function of disruptive GATA1 and CREB1 binding, respectively. **C-E:** Laboratory identification of allele-specific binding activity summarized as boxplots for (**C**) GATA1-rs7234029 in K562 cells and (**E**) CREB1-rs4147359 in Jurkat cells via DNA pull-down. Paired experiments (n=4) are denoted with dashed lines. Reduced binding was observed for both risk alleles, corroborating topology-based predictions. Western blot verification of DNA pull-down assay is also shown.

Among the twenty SNP-TFBS in isoclines 2-5, seven were derived from three SNPs within the *IL2RA* region (rs61839660, rs41295065, and rs4147359), spanning both of the region’s independent JIA-association signals (**Table 1**). Intronic SNP rs61839660 was the top JIA-association signal and overlapped bindings sites for MEF2A and MEF2C. The risk allele was predicted to facilitate binding for both TFs. Interestingly, within the MEF2C (MA0497.1) binding site, this G/A SNP overlaps the 13^th^ position which features both G and A at this location within the MEF2C motif (**Figure S15u**); yet the topology predicts allelic differences in binding. Allele-specific binding of MEF2C and MEF2A at rs61839660 is supported by previously published literature^35,36^ and highlights utility of considering beyond nucleotide sequence alone. In LD with rs61839660 is rs41295065 which resides within an intergenic region between *IL2RA* and *RBM17*; multiple TFBS (JUND, FOS, FOSL1, FOSL2) overlap this SNP. The JIA-risk allele’s topological profile was predicted to disrupt binding across all four TFs. Finally, SNP rs4147359 was in high LD with the lead SNP in the secondary *IL2RA* association signal and located within the *IL2RA-RBM17* intergenic region. This SNP overlaps a CREB1 (MA0018.4) binding site with the risk allele predicted to hinder CREB1 binding (**Figure 4d**). The predicted dampening of CREB1 binding was corroborated by our DNA pulldown assay in Jurkat T cells, where the risk allele reduced CREB1 binding by approximately two-fold (**Figure 4e; Figure S16b**).

### Polygenic Risk Score

We constructed a polygenic risk score (PRS) using the region-specific stepwise results previously reported^9,10^ using 341 SNPs weighted by the absolute value of the corresponding log(OR) for the additive genetic model. We also computed a PRS that included the HLA A*02:01-by-rs10038651 (ERAP2) interaction, identified within our current study. We applied these PRS to an independent subset of Phase 3 data (1001 JIA cases, 1100 controls (**Figure 5a**). In this external testing dataset, the PRS for non-HLA SNPs alone was highly significant (β=0.18±0.034, P=1.08x10^-7^) with an area under the ROC curve of C=0.77. Including the multilocus-selected HLA alleles and the A*02:01-by-rs10038651 interaction improved the PRS predictive value (β=0.375±0.029, P=1.96x10^-38^, C=0.80). Including a quadratic term for the PRS was also significant (P=2.6x10^-5^) with an inflection point at ∼52.5, but only modestly improved prediction (C=0.81) (**Figure 5b)**.

**Figure 5:**
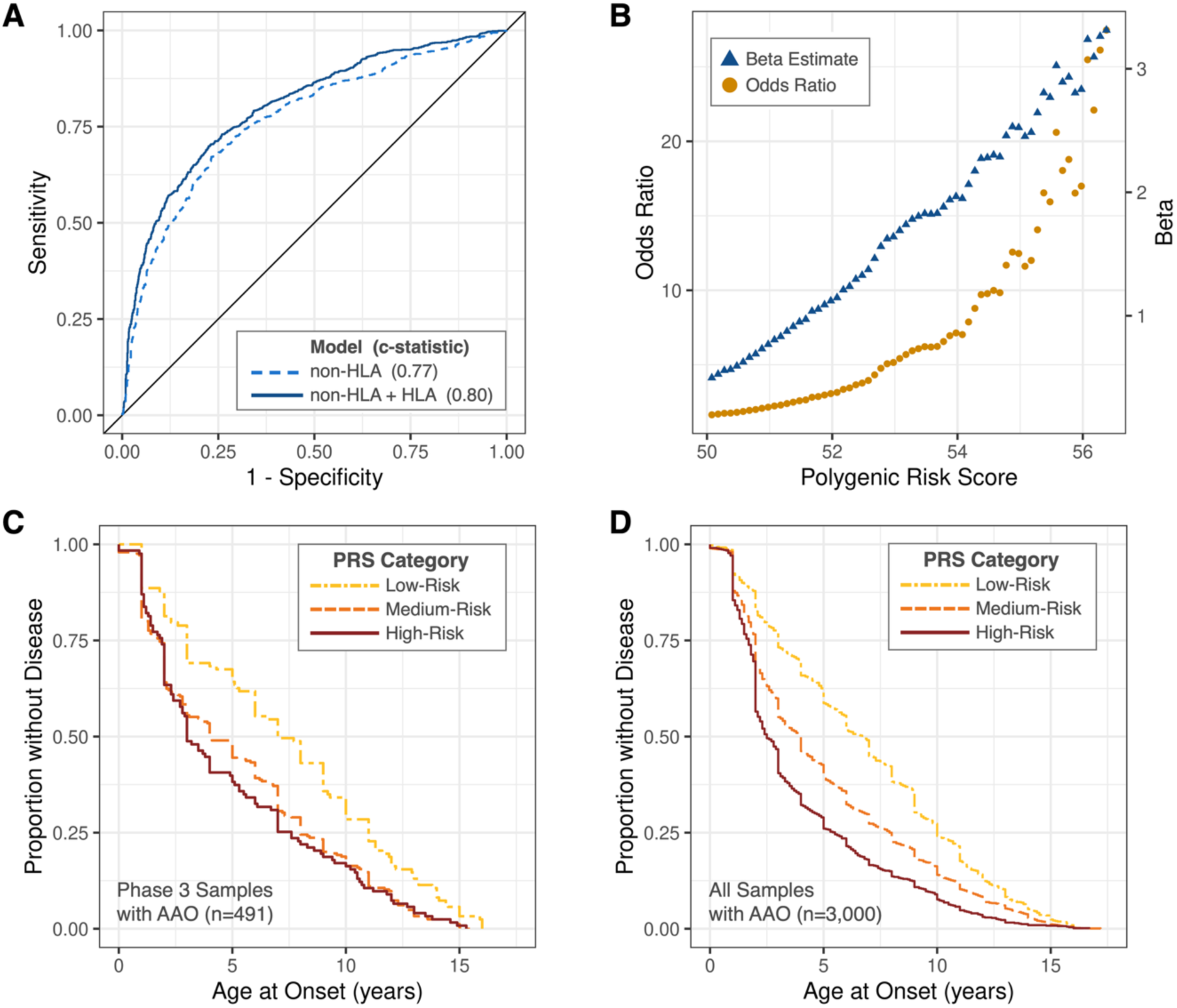
Performance of polygenic risk score (PRS) models for JIA and Age at Onset (AAO). A) Comparison of area under the receiver operator curve for two PRS models in independent Phase 3 samples. Non-HLA model included 341 SNPs weighted by the absolute value of the corresponding log of the odds ratio derived from Phase 2 samples (c=0.77). The ‘non-HLA + HLA’ model also included HLA alleles that were selected by the multivariable model and a jointly dominant (presence of *ERAP2* risk SNP and A*02:01) model for the *ERAP2*-A*02:01 interaction (c=0.80). B) Using a sliding window of polygenic risk scores (‘Non-HLA + HLA’ model), JIA risk was computed for independent Phase 3 samples (1001 JIA cases; 1100 controls) within a given PRS window. The beta estimates largely exhibited an additive effect, except for a slight inflection at 52.5. C) Kaplan-Meier curves for independent Phase 3 samples (with AAO, n=491) illustrate AAO differences by PRS category (polygenic risk for JIA relative to control). (Hazard Ratio=1.34; PCox=5.8x10-6). D) AAO Kaplan-Meier curves for the full JIA cohort (with AAO, n=3,000) by PRS category further illustrate significant earlier AAO for JIA cases with highest PRS (PLogrank = 1.1x10-43 ; Hazard Ratio=1.39 ; PCox=7.7x10-36).

We hypothesized that a greater PRS would be associated with an earlier AOO. **Figure 5c** shows strong differences among the Kaplan Meier curves stratified by the lower quartile, intermediate two quartiles, and the upper quartile of the HLA-inclusive PRS (Logrank test P=8.9x10^-6^) in the Phase 3 data of 491 JIA cases. The ordinal test under a Cox proportional hazards model, yielded a hazard ratio of HR=1.34 (PCox=5.8x10^-6^) **Figure 5c**; analyzing the PRS as a continuous variable, yielded an HR=1.13 (PCox=2.14x10^-7^). Given the PRS was derived from a case-control analysis, we applied the PRS to AOO for the entire cohort of 3,000 cases (**Figure 5d**) (Logrank test P=1.1x10^-43^ ; HR=1.39; PCox=7.7x10^-36^); analyzing the PRS as a continuous variable, yielded an HR=1.13 (PCox=6.76x10^-41^). The Kaplan-Meier estimated median time to AOO for the lower and upper PRS quartiles were 6.7 and 2.4 years, respectively; a 4.3-year difference. Finally, a PRS based on all association data within this study is also reported (**Table S20)**.

### Pathway Analysis and Drug-Gene interactions

We queried 583 eGenes from T1, T2, and T3 associations for potential drug-gene interactions (DI) using the Drug-Gene Interactions database^37^ (DGIdb), resulting in 94 eGenes linked to unique 582 drugs/small molecules (**Table S21**). Three eGenes were from the cytochrome P450 superfamily (*CYP1B1*, *CYP27B1*, *CYP2D6*) which are involved in drug metabolism and linked to 339 drugs. Reviewing the 582 drugs for links to autoimmune or rheumatic diseases (i.e., existing indications or peer-reviewed publications), identified 126 drugs (91 had non-CYP linkages) (**Figure S17**; **Table S22**).

Cognizant that the Immunochip was designed to fine-map 187 regions implicated in early autoimmune association studies, we completed a pathway analysis on 583 eGenes and identified 11 clusters, spanning 108 eGenes (**Figure S18; Table S23**). Cluster 1 (**Figure S18a**) is comprised of seven genes, related to chemokine receptors and features multiple DI with autoimmune and rheumatic (A&R) implications, including: plerixafor^38^, acetylcysteine^39^, cyclophosphamide anhydrous^40^, and genistein^41^. Cluster 5 (**Figure S18e**) was comprised of 22 eGenes, 9 with DI. This included *MAPK3* (8 DI) and *RAC1* (3 DI) as central nodes with links to multiple immune-important genes like *IL6* (28 DI), *TYK2* (9 DI), and *CD247* (3 DI). Thus, this cluster represents multiple pathways highly relevant for drug-targeting, including: *JAK-STAT, MAPK3*, *NFkB*, *PI3K-AKT-mTOR*, and *Keap1-Nrf2-ARE* pathways^42^.

## DISCUSSION

In this study, forty-one regions were associated with JIA (30 T1, 11 T2), including 14 novel regions. Similar to other complex diseases, >60% of these regions exhibited evidence for multiple, independent SNP signals, highlighting the importance of multi-SNP modeling approaches^43–45^. We leveraged the concept of multi-SNP signals by applying a regional-prioritization approach through the RegionalLRT. This process required at least one SNP meeting FDR significance; thus, the RegionalLRT adheres to a similar logic as a Fisher’s protected least significant differences multiple comparisons procedure^46^. Using the RegionalLRT, several regions increased in significance, such as *TYK2* with five independent signals. TYK2 inhibitors, such as deucravacitinib, have pharmacological interest and are approved for adult plaque psoriasis and are undergoing trials for psoriatic arthritis. Ranking by SNP-association would have classified *TYK2* as a T2 region (p=2.32 x10^-7^) but the RegionalLRT strengthened the association (p=3.77x10^-15^). This method identified 10 of the 14 novel regions. Generally, we observed consensus in the number independent signals between the multilocus stepwise modeling and the Bayesian approach, SuSiE^22^. SuSie is only validated for additive genetic models, whereas the stepwise modeling is applicable to any genetic model; an important distinction since many regions showed evidence of non-additive models. Exploring non-additive associations is well-motivated in complex genetic diseases as illustrated by renal disease and *APOL1*^47^.

Consistent with past studies, the HLA region showed the strongest association signal. Within the multilocus model, 10 of the 18 two-field HLA alleles were from DRB1, suggesting multiple, independent associations from this single gene. We sought to link association to biological mechanism via amino acid profiles. To avoid the challenges imposed by high-order interactions of a folded protein model^48,49^, we applied a clustering approach to narrow the amino acid drivers of risk. DRB1 allelic sequence similarity yielded two primary clusters, one which was predominantly comprised of JIA-risk alleles. Only three amino acids differentiated between these two (predominantly) non-risk and risk clusters. Two of these amino acids were encoded by exon 2 and fell within the binding pocket of DRB1, the third amino acid was encoded by exon 3, within an exogenous protein structure. This provides critical information for designing future binding experiments for JIA risk alleles which could help identify potential sources of immune activation or therapeutics.

Both A*02:01 and *ERAP2* are strongly associated with JIA risk^7,9,50^; however, this study presents the first population-based evidence of a statistical interaction in JIA. MHC Class I and aminopeptidase interactions have been observed in a handful of other immune-mediated diseases but those studies implicated other alleles, including: B*27 (Ankylosing Spondylitis), C*06:02 (Psoriasis), B*51 (Behçet’s Disease), and A*29 (birdshot chorioretinopathy)^51–54^. The disease-specificity of implicated Class I alleles suggests disease-specific mechanisms of risk mediated by peptide presentation. For most of these diseases, excluding birdshot chorioretinopathy, the strongest interactions were with *ERAP1*, not *ERAP2*. Although ERAP1 and ERAP2 share high sequence similarity and are involved in peptide processing, there are mechanistic differences, including differential preference of residues (e.g., ERAP2 favoring N-terminal based residues and shorter peptides)^51,55,56^. The greatest JIA risk was observed for having at least one copy of HLA-A*02:01 and the *ERAP2* haplotype corresponding to the non-truncated protein and increased *ERAP2* expression. These findings suggest that for some patients, JIA risk is potentially moderated through *ERAP2*-enabled peptide presentation to HLA-A*02:01. The role of ERAP2 for HLA-A*02:01 peptide presentation has been observed in cancer immunotherapy studies^57^ and highlights ERAP2 as a potential drug target. Critically, these results inform study design (e.g., presence/absence of A*02:01) for functional studies in JIA.

We applied DNA shape as a novel method for large-scale association studies to prioritize plausibly functional variants. This methodology complements existing resources (e.g., Regulome, cCREs, eQTLs) that identify functional regions, but on their own, do not decipher causal variants, either because the experiments are not allele-specific or LD creates correlated signals. Because DNA shape is an intrinsic property of DNA, it circumvents potential biases imposed by functional resources lacking large sample sizes or underrepresented genetic backgrounds/sequences. The biological relevance of DNA shape is well-documented^58–60^, but its application to large genetic association studies is largely untapped. We observed enrichment for large ΔMGW and ΔProT among JIA-associated SNPs and developed a SNP disruption score to quantify predicted allele-specific effects on TF binding, which has strong support for DNA shape relevance^15,18,61,62^. Half of the evaluated SNP-TFBS combinations did not have predicted allele-specific effects, emphasizing moving beyond mere proximity with functional features. Because a nucleotide exerts topological effects within the context of neighboring nucleotides^16^, DNA shape provides more information about a substitution within a TFBS. Thus, we posit topological TFBS profiles may be more biologically representative than traditional TF sequence motif representations which summarize information (nucleotide distribution) at each position, independently^63^. Supporting our *in silico* findings, several examples of functional evidence were reported, either from our own assessments or existing literature. Our identification of multiple SNPs within *IL2RA* linked to changes in TF binding, including our lab-validated CREB1 binding, highlights exciting opportunities for functional studies on combinations of co-inherited alleles. Again, DNA topology is posited as an additional tool as large changes in these metrics do not guarantee the causal-nature of a SNP.

The polygenetic risk score based on associations from our previous publication and applied to an external test set of independent set of cases and controls showed good predictive performance (C=0.77 for non-HLA region; C=0.80 combined HLA+non-HLA) in this European ancestral cohort. The combined HLA+non-HLA score incorporated the novel A*02:01-ERAP2 interaction. A higher genetic load of risk alleles was associated with an earlier age of JIA onset, with a median 4.3-year difference between lower and upper PRS quartiles.

A precision medicine paradigm seeks to link genetic associations to genes and then to drugs to identify drug targets and sources of heterogeneity in drug response. For linking eGenes to drugs with phenotype relevance, AI and LLM hold great potential given the complexity of integrating multiple names, sources, and indications. However, evaluating LLM results requires transparency of the process and careful manual validation. Of the 610 drugs identified by DGIdb, our prompts of Copilot identified 76 with A&R (autoimmune and rheumatic) relevance. Manual curation did not identify any false positives within this list but did yield an additional 50 drugs, resulting a total of 126 drugs with A&R relevance. Some of the drugs added via manual curation had established A&R indications (e.g., Filgotinib^64^), but others emerged from peer-reviewed research. Thus, AI tools are helpful but not sufficient; all lists require vetting by experts for the potential of pragmatic or traditional clinical trials.

This study has several limitations. The Immunochip interrogates immune-relevant regions and does not have genome-wide coverage. Our application of a TFBS disruption metric was limited to TFBS identified by REMAP2022 and JASPAR 2024, biasing towards more frequently studied TFs. For instance, previous work^32^ identified rs11889341 as a putative causal variant based on HMGA1 binding, and our prior assessment^19^ highlighted this variant for its MGW disruption. However, because this HMGA1 TFBS was not within the leveraged public resources, it was not analyzed with the TFBS disruption pipeline described here. However, this SNP was flagged for its strong effects on MGW and ProT, highlighting the utility of delta shape features for regions lacking robust functional annotations. Finally, the available cohorts were of European ancestry and examination of these results and new discoveries in other ancestral groups should be a priority.

In conclusion, in addition to the 14 novel loci, these results motivate multiple hypotheses such as interrogation of ERAP2 peptide presentation to A*02:01, epitope binding of the JIA DRB1 risk cluster, and the influence of identified allele-specific TF binding events. The novel approaches in this study are broadly applicable to complex genetic traits.

## ACKNOWLEDGEMENTS

In remembrance of Professor Susan D. Thompson of Cincinnati Children’s Hospital Medical Center for her decades of work in genetics and immunology of childhood autoimmune diseases, especially JIA. We gratefully acknowledge the contribution of members of the JIA Immunochip consortium^10^.

This work was supported by the National Institutes of Health: R01 EY030521, R01 AR057106, P01 AR048929, R01 AR078785-02, 2R01 AR073201, 2P30 AR070253, RC1 AR058587, P30 AR073750, and P20-AR070549. We also acknowledge the Daryl and Marguerite Errett Discovery Award (H.C.A.); the Val Browning Foundation (J.F.B.); Unrestricted funding of the Garmisch-Partenkirchen Biobank by the "Hilfe für das rheumakranke Kind e.V."(J-P.H.); the Scottish Rite for Children Research Fund (C.A.W.); Arthritis Society Canada —Grant #: 07-86 (A.M.R.); UM1AI144291 (J.A.J.); the OMRF Biorepository (J.A.J.); NIHR Senior Investigator award (L.R.W.); the NIHR-GOSH Biomedical Research Centre (L.R.W.); NIHR Manchester Biomedical Research Centre (K.L.H.); a Canada Research Chair in Precision Medicine in Childhood Arthritis and Rheumatic Diseases (R.S.M.Y.); the Centre for Centre for Genetics and Genomics Versus Arthritis —grant: 21754 (J.B.); Liaison Committee between the Central Norway Regional Health Authority and NTNU - Norwegian University of Science and Technology (grant 5056/46051000 to V.V.); the Northern Norway Regional Health Authority (Grant HNF1450-19) (V.V.); and the NIHR Manchester Biomedical Research Centre—NIHR203308 (S.L.S.);

We acknowledge Nils Thomas Songstad and Nina Moe for patient recruitment in the Norwegian subcohort of the Nordic JIA study and Kristin Rian for technical support. The HUNT study is a collaboration between the HUNT Research Centre (Faculty of Medicine and Health Sciences, NTNU - Norwegian University of Science and Technology), Nord-Trøndelag County Council, Central Norway Health Authority, and the Norwegian Institute of Public Health. Johanna Hadler, Katie Cremin, Karena Pryce, and Jessica Harris are acknowledged for excellent technical assistance (E.B.N., M.R., V.V.).

This study acknowledges the use of the following UK JIA cohort collections: The British Society for Paediatric and Adolescent Rheumatology (BSPAR) (funded by a research grant from the British Society for Rheumatology (BSR); The UK Childhood Arthritis Prospective Study (CAPS) was funded by Arthritis UK (grant number 20542). United Kingdom Juvenile Idiopathic Arthritis Genetics Consortium (UKJIAGC). This work is supported by the NIHR Biomedical Research Centre: Manchester. The views expressed in this publication are those of the authors and not necessarily those of the NIHR, NHS or the UK Department of Health and Social Care.

We also acknowledge that the Childhood Arthritis Response to Medication Study was funded by Sparks UK (08ICH09) Big Lottery Fund, UK (RG/1/010135231), Great Ormond Street Children’s Charity (VS0518) Versus Arthritis (22084) and MRC (CLUSTER, MR/R013926/1). L.R.W. was supported by an NIHR Senior Investigator award and the NIHR-GOSH Biomedical Research Centre.

Primary analyses were computed using the Wake Forest University (WFU) High-Performance Computing Facility, a centrally managed computational resource available to WFU researchers including faculty, staff, students, and collaborators. Additional analyses were completed using the Wake Forest University Health Sciences computing cluster.

## DATA AVAILABILITY

Summary statistics will be submitted to the GWAS catalog. The polygenic risk score model is available in supplementary tables and will be submitted to the PGS catalog upon publication per PGS requirements. Individual genotype data will be made available via dbGaP consistent with individual consent/assent. Annotation data was pulled from public repositories or previously published data, including: Drug-Gene Interaction Database (DGIdb) v5.0 (https://dgidb.org); Functional Annotation of Variants Online Resource (FAVOR) v2.0 (https://favor.genohub.org); Genotype-Tissue Expression (GTEx) v8 (https://gtexportal.org); GWAS Catalog (https://www.ebi.ac.uk/gwas/); IPD-IMGT/HLA FTP Directory Release 3.56.0 (http://www.ebi.ac.uk/ipd/imgt/hla/); JASPAR 2022 Transcription Factor Binding Data -as available at the UCSC Table browser (https://www.genome.ucsc.edu/cgi-bin/hgTables); JASPAR CORE 2024 Collection (https://jaspar.elixir.no); ReMap 2022 Non-Redundant Peaks (https://remap.univ-amu.fr); JIA PBMC eQTL Dataset (GEO Accession: GSE13501, https://www.ncbi.nlm.nih.gov/geo/query/acc.cgi?acc=GSE13501); RegulomeDB v2.2; SCREEN (ENCODE cCREs) (https://screen.wenglab.org). Additional information is available in the Key Resources list, found in Table S24.

## CODE AVAILABILITY

For this study, publicly available software was utilized for all analyses, as described in METHODS and the Key Resources List, found in Table S24.

## AUTHOR CONTRIBUTIONS

Study was conceptualized by H.C.A. and C.D.L. Investigations were completed by H.C.A., E.S.K., M.S., M.C.M., K.J., M.E.C., J.N.J., and C.D.L. Funding was acquired by H.C.A., J.A.J., P.A.N., S.T.A-H., J.N.J., and C.D.L. Original manuscript was drafted by H.C.A., E.S.K., M.C.M., M.E.C., T.D.H., and C.D.L. Methodology was developed by H.C.A., E.S.K., K.J., M.E.C., J.N.J., and C.D.L. Validation was performed by K.J., M.E.C., and C.D.L. Visualizations were created by H.C.A., E.S.K., M.C.M., K.J., M.E.C. Formal analysis was conducted by H.C.A., E.S.K., M.C.M., M.E.C., and C.D.L. Data was curated by H.C.A., E.S.K., M.S., M.C.M., K.J., and C.D.L. Resources were provided by J.F.B., P.M.G., J.A.J., M.S., P.A.N., E.B.N., A.M.R., M.R., V.V., C.A.W., R.S-M.Y., G.S.S, S.P., J.B., S.T.A-H., J.N.J., and C.D. L. Reviewing and editing of the manuscript was conducted by H.C.A., E.S.K., M.S., M.C.M., K.J., J.F.B., M.E.C., P.M.G., J-P.H., T.D.H., K.H., J.A.J., P.A.N., E.B.N., A.M.R., M.R., S.L.S., V.V., L.R.W., C.A.W., R.S-M.Y., S.D.T., G.S.S., S.P., J.B., S.T.A-H., J.N.J., and C.D.L.

## ONLINE METHODS

### Study cohort

Samples were obtained from previously published studies^1,2^ and 1,356 controls and 1,123 cases were from newly acquired samples. Data were contributed by groups in the United States of America, United Kingdom, Germany, Norway, and Canada. All samples were provided with approval from respective institutional review boards (IRB) or ethics committees. JIA cases were assigned to subtypes according to the ILAR classification criteria^3^. For all analyses described in this study, cases were limited to the two most common JIA categories (herein denoted as ‘subtypes’), polyarticular RF- and oligoarticular (persistent and extended).

### Genotyping and quality controls

All samples were genotyped using the Illumina Infinium Immunoassay V1 (Immunochip). Genotyping was performed according to Illumina’s protocols at labs at the University of Manchester (UK), Cincinnati Children’s Hospital (USA), Feinstein Institutes for Medical Research (USA), University of Utah (USA), the Hospital for Sick Children (Canada), and University of Toronto (Canada). The Illumina GenomeStudio GenTrain V.2.0 algorithm was used to recluster all samples, together, for allele calling, and was completed at Cincinnati Children’s Hospital.

Single-nucleotide polymorphisms (SNPs) were initially excluded if they had a call rate <98% and a cluster separation score of <0.4. A SNP was subsequently removed for differential missingness between cases and controls (p<0.05), exhibited departure from Hardy-Weinberg equilibrium (p<0.000001 in cases or p<0.01 in controls), or had a minor allele frequency (MAF) <0.01 in controls.

Population substructure was computed for all samples using high-quality, linkage disequilibrium (LD)-pruned SNPs and the software ADMIXTURE^4^. Population estimates were computed for 2, 3, and 4 postulated ancestral populations (*k*). Reference genotypes from CEU, YRI, and CHB populations from Phase II HapMap^5^ were used as anchoring populations. The appropriate *k* was identified by comparing the cross-validation error for the three runs. Population outliers were excluded from further analyses. Additional samples were removed based on the following quality control features. Samples with a call rate <98% or excessive heterozygosity (f > 0.1) were excluded. Duplicates were removed (dropping the sample with the lowest call rate). KING^6^ was used to identify sample pairs with a kinship coefficient > 0.25. The sample with the most complete phenotypic data and/or genotypic data was retained from each related pair. Self-reported sex was not confirmed genetically due to the Immunochip’s low number of SNPs on the X chromosome.

### Statistical analysis of SNP associations

Reported regions are named based on proximal genes that spanned or bound the association signal, unless existing literature supported usage of a gene name more relevant to JIA or autoimmune phenotypes. Three tiers of statistical significance were defined for this study: Tier 1 (P< 5x10^-8^) meeting standard genome-wide association significance, Tier 2 (P<1x10^-6^) meeting significance based on the number of independent (LD-pruned) tests across the Immunochip^7^, and Tier 3 using the Benjamini-Hochberg False Discovery Rate (FDR) across all associations (PFDR<0.05).

JIA case-control SNP associations were computed using a logistic regression model via SNPGWA version 4.0 module of SNPLASH (see Table S24). For each model, three admixture proportions were included as covariates to account for population substructure. SNP associations are reported for the additive genetic model unless the lack-of-fit test for the additive model was significant (P<0.05). In this scenario, inference was based on the most significant genetic model (dominant, recessive, or additive model). Consideration of the additive and recessive models required at least 10 and 30 individuals homozygous for the minor allele, respectively. The genomic control inflation factor (λGC) was calculated using SNPs that met the quality control criteria described above. As recommended for large studies^8^, the λGC was scaled to 1,000 cases and 1,000 controls for standardization and comparison with other studies.

Stepwise logistic regression was used to identify the presence of multiple, independent signals within each JIA-associated region, adjusting for population substructure. SNPs entering the stepwise analysis were selected by taking a 1Mb window around a lead SNP based on the single-SNP analyses. A stepwise logistic regression model was then computed using forward selection (entry P<0.001) with backward elimination (exit P>0.001). For regions with only one SNP retained by the stepwise model, a “regional p-value” was defined by the single-SNP association p-value for that SNP. For regions with two or more independent signals, a regional-p-value was computed using the likelihood ratio test for the SNPs in the multilocus model (regional likelihood ratio test; RegionalLRT), which included all SNPs identified by the stepwise modeling and admixture estimates.

In addition to the stepwise logistic regression and RegionalLRT, we applied the Sum of Single Effects Model (SuSiE)^9^, which implements a Bayesian method for prioritizing SNPs in each region by quantifying the uncertainty of each selection. Because SuSiE’s algorithm is only validated for the additive model, SuSiE was not computed for regions with a large proportion of SNPs exhibiting non-additive modes of inheritance.

SNP associations with Age of Onset were analyzed with a linear regression model adjusting for admixture estimates (QSNPGWA, SNPLASH v4).

Manhattan plots were created using topr^10^, and regional association plots were created using the Bioconductor package karyoploteR^11^. For regional association plots, linkage disequilibrium (LD) values were computed using control data from this study via PLINK v1.9^12^. Gene track annotations were constructed using the Bioconductor packages org.Hs.eg.db and TxDb.Hsapiens.UCSC.hg38.knownGene which contain gene mappings based on the UCSC knownGene table.

Polygenic risk scores (PRS) were computed using regional-stepwise association signals from our previously published study samples^2^ and were applied to the independent cases and controls that were not included in the previous publication; that is, it is an external test based on 1,001 JIA cases and 1,100 controls. Tagging SNPs for association signals were summed by using the absolute value of the log of the odds ratio for the additive genetic model. Given its strong effect, we also included the A*02:01-*ERAP2* interaction reported here, where the included term is defined as a jointly dominant model for A*02:01 and rs10038651 minor allele (i.e., assigned ‘1’ if both A*02:01 is present and the *ERAP2* rs10038651 T allele is present), weighted by its log(OR) from the previous publication samples^2^. PRS performance was assessed within 2,101 independent (“Phase 3”) samples by using a logistic regression model, adjusting for population substructure. A logistic regression model was also computed including a quadratic term for the PRS to test for potential non-linear effects. The same PRS, which was derived from the previously published JIA risk associations, was applied to both the case/control data and to the age of onset data. We hypothesized that higher genetic risk would be associated with earlier age of onset. The relationship between PRS and age of onset was visualized using Kaplan-Meier curves by splitting the cohort of independent samples into three categories based on PRS quartiles (lower quartile, inter-quartiles, and upper quartile). The ordinal test using a Cox proportional hazards model was computed to test for an association with the PRS, adjusting for population substructure. This analysis was repeated using the full set of JIA cases with age of onset data (n=3,000). We also computed a PRS based on regional-stepwise association results from entire sample quality-control passed samples reported in the present study.

### Functional annotations

#### The GWAS Catalog

was used to explore whether the JIA-associated regions are also associated with other autoimmune diseases. SNPs from the NHGRI-EBI GWAS catalog^13^ were downloaded on December 6, 2024 via the EBI hosted downloads link (ebi.ac.uk/gwas/docs/file-downloads). The SNPs associated with autoimmune-related traits were identified and examined for overlap (based on genomic coordinates) with JIA Tier 1 and Tier 2 associated regions in our study. To determine if there were differences in the representation of diseases across established and novel loci, the counts for the most represented diseases within established loci (rheumatoid arthritis, inflammatory bowel disease, Crohn’s disease, multiple sclerosis, systemic lupus erythematosus, type I diabetes, celiac’s disease, ulcerative colitis, psoriasis, and ankylosing spondylitis) were compared using a chi-squared test for independence.

#### Transcription Factor Binding Site Overlap

We assembled a dataset of high-confidence transcription factor binding sites (TFBS) by intersecting JASPAR’s 2022 sequence-based TFBS predictions^14^ and REMAP 2022, a database of non-redundant ChIP-seq peaks^15^. Using BEDTools^16^, the 678,565,535 JASPAR 2022 TFBS were intersected with the 67,058,536 ChIP-seq peaks of REMAP 2022 (based on human data) and only retained if the intersection was also matched by transcription factor (TF). This process yielded a database of 13,539,090 autosomal TFBS spanning 460 TFs which had supporting evidence by both sequence-based predictions and ChIP-seq data. JIA SNP associations (excluding the HLA region) were intersected with this dataset to identify SNP-TFBS overlap. UniProt annotations^17^ were retrieved from uniprot.org via the UniProt ID listed on JASPAR website.

#### Expression quantitative trait loci (eQTLs)

JIA-associated SNPs were queried for evidence of being an eQTL in JIA-relevant tissues (see below). The list of JIA-associated SNPs (Tiers 1-3) was expanded to include SNPs in high LD based on 1000G EUR population (r^2^> 0.5) by using the LDlink R-package v1.4.0^18^. These SNPs were then queried for association with gene expression in whole blood, cultured fibroblasts, EBV-transformed lymphocytes, and spleen in the Genotype-Tissue Expression database (GTEx version 8)^19^. Here, statistical significance of an eQTL was defined by GTEx: https://gtexportal.org/home/methods. The JIA-associated SNPs (plus proxies) were also tested for association with gene expression in peripheral blood mononuclear cells (PBMCs) from a subset of the JIA patients (n=63) and controls (n=20) who were genotyped on the Immunochip. The gene expression study was previously described^20^. Statistical significance for the PBMC-based eQTL analysis was defined by p < 0.0001. Genome-wide plots of eQTLs were created using topr^10^. Genes identified via eQTL searches (eGenes) were used for pathways analyses and drug target queries (see *Biological Pathways*, below).

#### Functional Databases

JIA associated SNPs were interrogated for functional evidence using: RegulomeDb^21^, Search Candidate Regulatory Elements by ENCODE (SCREEN)^22^, Functional Annotation of Variants (FAVOR)^23^, and the Drug-Gene Interaction database (DGIdb)^24^, Regulomedb was searched by rsID, using the pre-calculated scores based on dbSNP v153 common SNPs in GRCh38, as available for download at https://regulomedb.org. Comparison of RegulomeDb rankings between T1/T2 SNPs and non-significant (NS p>0.5) SNPs was computed based on a two-sample test of the proportions of SNPs within highest evidence ranks (Regulome Ranks 1-2). SCREEN candidate cis-Regulatory elements (cCREs) based on human GRCh38 data were downloaded. Overlap for cCREs was considered for five categories: any tissue, whole blood, eye (selected due to the high proportion of JIA patients that develop uveitis^25^), connective tissue, and spleen. A two-sample test of the proportions was used to evaluate enrichment of cCREs present in T1/T2 SNPs compared to non-significant (NS p>0.5) SNPs. FAVOR annotations using GRCh38 were obtained using the online resource at https://favor.genohub.org. FAVOR was also used to annotate the SNPs’ functional attributes (e.g., intronic, intergenic, ncRNA). Potential drug-gene interactions were identified using the drug-gene database (DGIdb) v.5.0.11^24^. Queried genes included those associated with eQTLs (eGenes) or those which exhibited predicted allele-specific binding events in the DNA topology TFBS analysis (described, below). The drug/small molecule-interactions identified by the DGIdb are only intended for research purposes. Only drugs/small molecules that were labeled as ‘approved’ by the DGI are presented in results. We note that the ‘approved’ label is an aggregate label created by DGIdb, across regulatory bodies^24^. Thus, a DGIdb ‘approved’ status for a particular drug is not synonymous with ‘FDA approval’. Drugs retrieved from DGIdb queries were searched for potential autoimmune and rheumatic (A&R) implications. As a first pass, all drugs were submitted to Microsoft CoPilot with a prompt to identify evidence of A&R relevance with ‘slow thinking’ selected, with prompt instruction to return reference links for each provided piece of evidence. Subsequently, all provided links and evidence were manually evaluated. All remaining drugs that were not identified via LLM queries were then manually searched for evidence of A&R relevance using the American College of Rheumatology (rheumatology.org) and PubMed searches. For drugs only identified with A&R evidence via peer-reviewed research, two PMIDs are provided as examples.

### Sequence-dependent DNA topology

DNA shape features of minor groove width (MGW) and propeller twist (ProT) were computed using the Bioconductor package DNAshapeR^26^. MGW and ProT were predicted using a rolling pentameter window; thus, for each SNP, the flanking (± 4) nucleotides were extracted from the hg38 reference genome using BEDTools^16^. For each SNP, MGW and ProT were computed for both alleles, and changes in MGW (ΔMGW) and ProT (ΔProT) were computed as a Euclidean distance at the SNP ± 1bp^27^. ΔMGW and ΔProT were compared between JIA-associated (Tiers 1 or 2) and non-significant (P>0.5) SNPs using a two-sample t-test. Quantile logistic regression using the R package quantreg (<u>10.32614/CRAN.package.quantreg</u>) was used to assess differences at the 10^th^, 25^th^, 50^th^, 75^th^, and 90^th^ percentiles.

To quantify a SNP’s impact on TF binding, we constructed TF topological profiles using experimental binding sites from JASPAR 2024’s website^28^. Using these curated sequences, which contain experimentally-derived binding events, reduced the likelihood that our topological profiles would include false positives (non-binding event sequences). MGW and ProT profiles were computed for TFs that had experimental binding sites available and had at least one TFBS that overlapped a JIA-associated SNP (previously described JASPAR-REMAP intersection) (**Figure S12**). For these TFs, sequences from curated bindings sites were extracted from the hg38 reference genome, including ±4bp to account for the sliding pentameter window of DNA shape prediction. DNAshapeR was again used for MGW and ProT computation for each sequence. It is important to recognize that the length of a binding site varies based on the TF. For each TF, each position in the n-length binding sequence was summarized for MGW and ProT values at the 90^th^, 75^th^, 50^th^, 25^th^, and 10^th^ percentiles.

TFBS that overlapped a JIA-associated SNP were aligned to the TF topological profile described above. For a given SNP, each allele’s topological profile was compared to the TF profile by constructing a disruption metric (**Equation 1**). The metric begins with an *n-mer* sequence (indexed by *j*) that represents a TFBS (± 2 flanking nucleotides). For *i=1,…,n* positions in this sequence, *μi,j* represent the shape feature prediction (MGW or ProT) of the sequence at position *i*. The disruption metric incorporates the median and interquartile range of the TF topological profile, thereby accounting for the varied conservation of specific positions within a TFBS. Thus, the topological binding disruption metric *d* is given by the following equation:

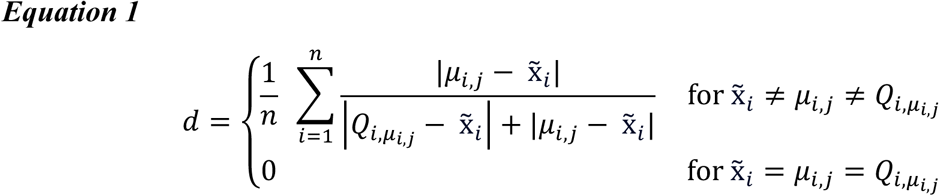

*Where*,

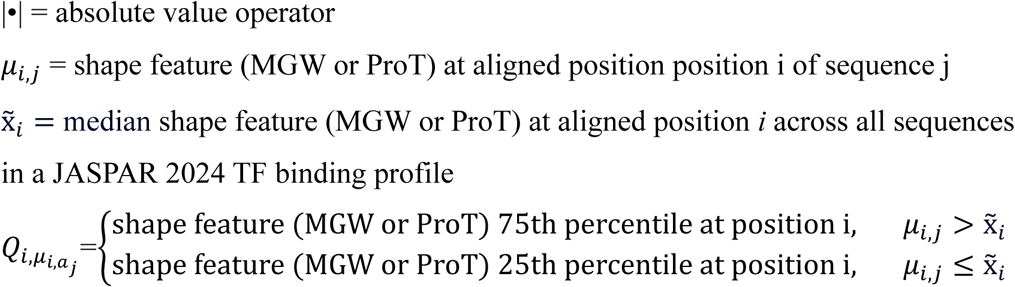

The numerator is the absolute difference between the shape feature of the input sequence and the median of the TF topological profile. The denominator also includes the absolute difference between the shape feature of the input sequence and the nearest inner quartile (e.g., 25^th^ quartile if the sequence value is below the median; 75^th^ quartile if the sequence value is above the median). Thus, *d* represents the average across the individual measures at each position across the binding site and is bounded between 0 and 1. This captures the standardized deviation of a given sequence from a TF topological profile.

The absolute change in *d* as calculated for two alleles at a SNP provides our SNP-TFBS Score (**Eq 2**).

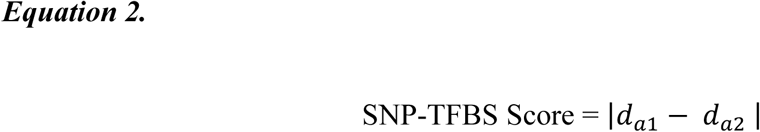

Where,

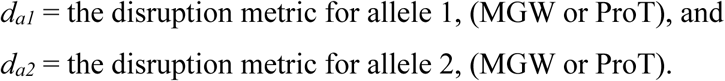

Greater SNP-TFBS scores indicate a greater likelihood of allele-specific TF binding effects. To facilitate ranking using both MGW and ProT SNP-TFBS scores, an l2-norm, which measured the Euclidean distance of the two scores from a (0,0) reference point, was computed. To categorize levels of predicted binding disruption, isoclines were defined based on 0.05 intervals.

### Experimental validation of identified allele-specific TFBS events

K562 and Jurkat were selected for their expression of GATA1 and CREB1, respectively. Cells were cultured in RPMI 1640 medium supplemented with 10% FBS and Penicillin and streptomycin maintaining a cell density of 1-8 × 10^5^ cells per mL at 37 °C and 5% CO2. Nuclei were isolated using Nuclear Extract Kit (Active Motif, Carlsbad, CA 92008) following manufacture’s instruction. The protein concentrations of the nuclear extracts were determined by Pierce™ BCA Protein Assay Kit (Thermo Fisher Scientific).

#### Preparation of Biotinylated Double-Stranded DNA Probes

The fragment to be analyzed was generated by PCR using genomic DNA from human CD4+ T cells as template. The PCR amplicon was cloned into a PGL4.23 vector (Promega, Madison, MI, USA). The mutations were introduced with the QuikChange II Site-Directed Mutagenesis Kit (Agilent, Santa Clara, CA) following the protocol from Agilent. All recombinant plasmid sequences were confirmed by Sanger sequencing. PCR primers for probes were designed to cover the region to be analyzed and to have an amplicon length between 200bp to 230bp. A 5^’^-biotin-modified nucleotide was added to the reverse primers. As a negative control, a fragment of the vector backbone was amplified. The probes were amplified by standard PCR and were then purified from agarose gels using Wizard® SV Gel and PCR Clean-Up System (Promega) following the manufacturer’s instructions. DNA concentration was determined using NanoDrop (Thermo fisher Scientific).

#### DNA Pull-down Assay

To block non-specific DNA–protein interactions, nuclear extracts (2mg/ml) were incubated with salmon sperm DNA (a final concentration of 100 μg/ml. Thermo Fisher Scientific) on a rotator for 15 min at room temperature. After incubation, samples were centrifuged at 16,000xg for 1 min to remove precipitates. One milligram of nuclear extract was transferred to microcentrifuge tubes containing 1ug of DNA probe and rotated for 40 min at room temperature, after which 30 ul of Streptavidin MicroBeads (Miltenyi Biotec, Inc. Auburn, CA, USA) were added to the tube and incubated for another 10 min. The biotinylated probes/protein complex were isolated using the μMACS Separator in combination with μColumns following the manufacturer’s instructions (Miltenyi Biotec, Inc.). The column was washed 8 times with 200 ul washing buffer (20mM Tris-HCl, (pH8.0), 1 mM EDTA, 10% Glycerol, 1 mM DTT, 50 mM NaCl). To elute the proteins, 20 μl pre-heated (95C) elution buffer (1x Sample buffer. Bio-Rad, Hercules, CA, USA) were added to the columns incubating for 5 min, then 110 ul pre-heated elution buffer were added to the columns and collected eluates in 1.5 ml microcentrifuge tubes. Samples (30 ul each) were analyzed by 10% SDS-PAGE/Western blot. The protein levels were quantified with the iBright Analysis System (Thermo Fisher Scientific). Statistical significance was based on P<0.05 using a paired t-test between the reference and alternate alleles (n=4 replicates) for each SNP.

### HLA imputation and associations

#### Imputation and association testing

Two-field HLA alleles were imputed using HiBAG and its European ancestry reference panel^29^. For single-allele association analyses, a logistic regression model was computed for JIA cases vs. controls and a linear regression model was computed for age of onset. Allelic dosages were used in the models to account for imputation uncertainty. Single-allele analyses were limited to HLA alleles with a best guess count of at least 10 in either cases or controls. To build the multi-allelic models, stepwise (logistic or linear, depending on outcome) regression with forward selection and backwards elimination was computed, with a significance threshold of P<1x10^-4^. To assess impact of HLA alleles on JIA SNP associations in the major histocompatibility (MHC) region, SNP associations were re-analyzed adjusting for HLA alleles selected by the stepwise regression model. To account for population substructure, admixture proportions were included in all analyses.

#### Clustering analyses

To circumvent challenges from using stepwise modeling for high-order interactions among amino acids^30,31^, a clustering approach was used to isolate potentially functional amino acids relevant to JIA. Amino acid FASTA files for HLA-A, B, C, DRB1, DQA1, DQB1, and DPB1 (e.g., DRB1_prot.fasta) were retrieved from the IPD-IMGT/HLA GitHub account. Sequences for the two-field alleles analyzed in the single-allele analyses were extracted using biopython^32^. Amino acid sequences for each gene were clustered using the EBI web-hosted version of CLUSTAL OMEGA with standard input options^33^. The resulting phylogenetic trees were visualized using the Bioconductor package ggtree^34^, and sequence alignments were visualized using Jalview v2.11.5.1^35^. Comparisons of consensus sequences (e.g., between clades or within a clade) were determined by relative JIA risk patterns across the tree. The Protein Data Bank (PDB) was queried for relevant molecular structures to illustrate the position of highlighted amino acids within an HLA molecule. Images using PDB structures were created using Chimera v1.15^36^.

### Analysis of interactions

SNP-by-SNP interactions were computed using logistic regression models which included the two SNPs, their centered cross-product, and three admixture proportions. The likelihood ratio test for the centered cross product was implemented using the Intertwolog module in SNPLASH.

SNP-by-HLA interactions were computed across the genome using each two-field HLA allele selected by step-wise modeling (previously described). Interactions were tested using a case-only logistic regression model^37^, where the outcome was the binary presence of the two-field allele (based on best-guess), the SNP was the predictor, and admixture proportions were included to account for population substructure. Based on the results of those analyses, two additional models (JIA cases versus controls —adjusting for admixture proportions) were computed in the *ERAP2* region, with the samples stratified into two subsets based on presence/absence of A*02:01.

### Biological pathways

Genes associated with the eQTLs identified by GTEx and JIA PBMCs (eGenes, described above) were used to construct biological pathways. Genes were linked to their protein products and protein-protein interactions were identified using STRING (v12.0)^38^. MCODE clustering and visualization was completed using the clusterMaker2 application in Cytoscape (v3.8.2)^39–41^. Clusters with at least three proteins were retained. Clusters were labeled by summarizing the ten pathways in Reactome (v92) for which the cluster was most enriched (top ten enrichment p-values)^42^.

### Software and Databases

Software and publicly resourced databases have been described throughout the methods. A compilation of all software and databases utilized within this study are presented in **Table S24**.

