## Supplementary Figures for "Identification and evaluation of 41 risk loci for juvenile idiopathic arthritis informs precision medicine: mechanistic implications of DNA topology and an HLA-A*02:01-ERAP2 interaction"

**Supplementary Figure 1: Admixture proportions plots.**

Population structure of study subjects, motivating usage of three admixture proportions (for substructure) as covariates in association testing. Observations corresponding to JIA cases and controls are coded in green and blue, respectively. Observations corresponding to subjects with ancestry from Han Chinese in Beijing, China (CHB), Utah residents with Northern and Western European ancestry (CEU), and Yoruba in Ibadan, Nigeria (YRI) are coded in purple, yellow, and black, respectively. **A**) admixture proportions 1 versus 2. **B**) admixture proportions 1 versus 3. **C**) admixture proportions 2 versus 3.

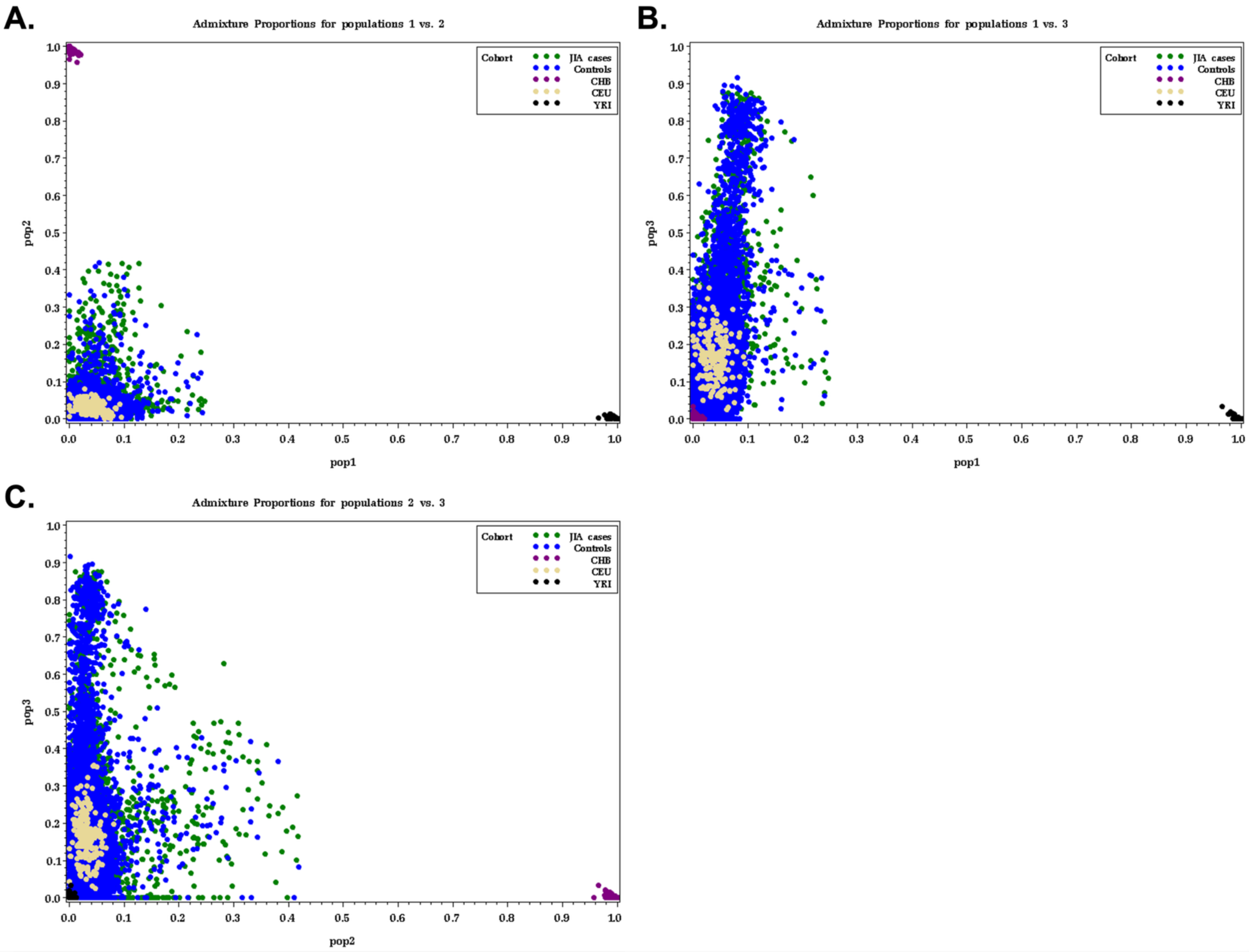

**Supplementary Figure 2: Regional plots of association for Tier 1 and Tier 2 loci associated with JIA.**

**(a)-(ao)** For each region, SNPs identified through the stepwise procedure are labeled in pink. T1 ($P< 5x10^{-8}$) and T2 ($P < 1x10^{-6}$) significance thresholds are plotted as red horizonal lines (solid and dashed, respectively). Because regions with more than one independent SNP identified through stepwise modeling were deemed significant by the joint multi-locus likelihood ratio test, individual SNP associations may fall below these thresholds in regions that met significance via the likelihood ratio test. Plots depicting the 'posterior Inclusion probabilities' (PIP) were created for loci where SuSiE fine-mapping could be computed, and SNPs contained in SuSiE credible sets are displayed in saturated colors. The number of independent signals identified by SuSiE and the stepwise procedure are indicated in the table below each plot. Linkage disequilibrium (LD) was calculated relative to the top SNP in each region using genotype data from JIA controls. SNPs overlapping a JASPAR 2022 transcription factor binding site (TFBS) supported by ReMap 2022 non-redundant peak data are annotated with white asterisks.

### (a) *PTPN22* Region (Tier 1)

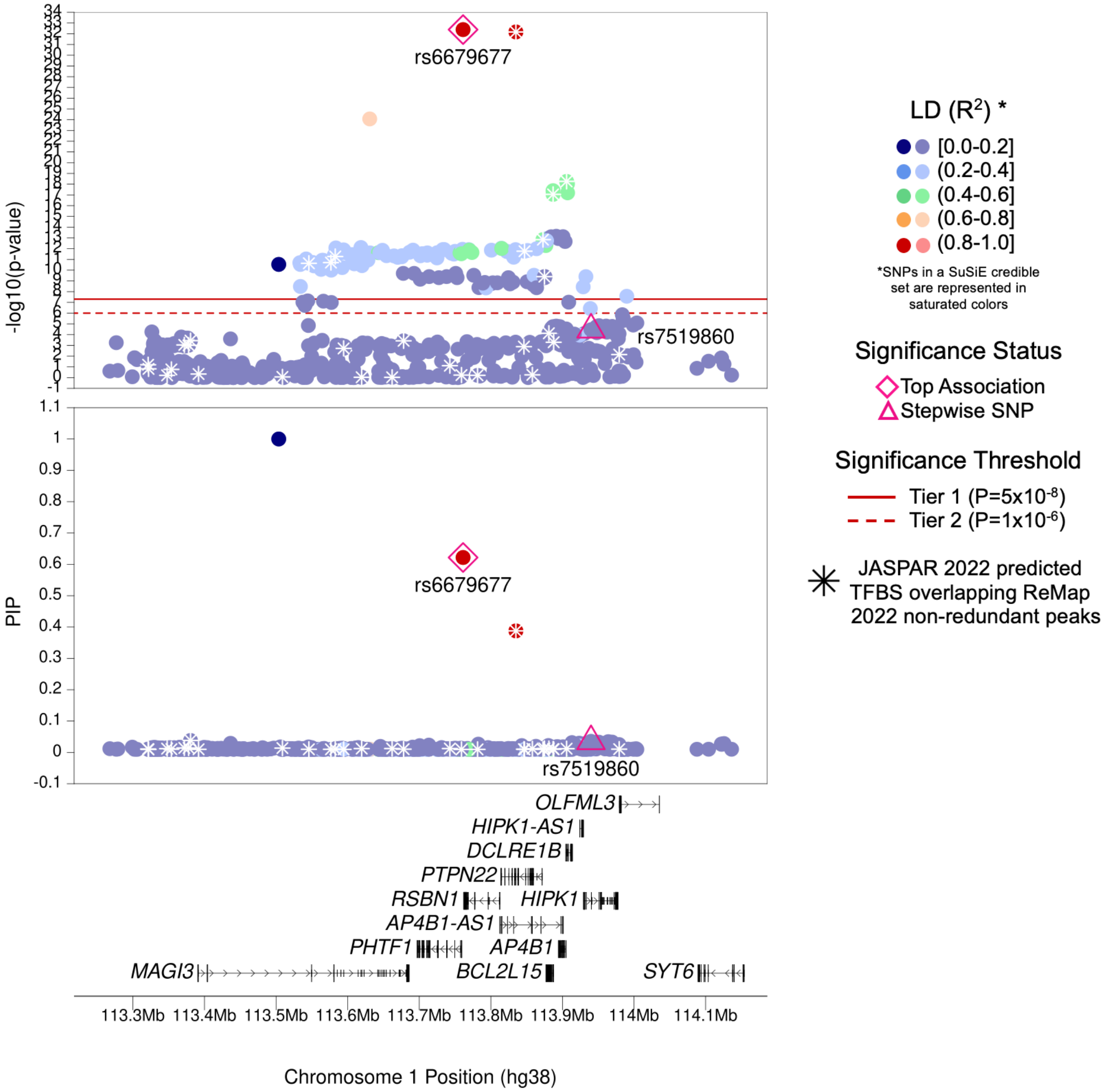

| | |
|---|---|
| **Region Name** | *PTPN22* |
| **Stepwise Region Window (hg38)** | chr1:113261186-114261186 |
| **Regional P-Value (df)** | P = $5.20\times10^{-33}$ (df = 2) |
| **Novel for JIA (Yes/No)** | No |
| **Number of Signals: Stepwise** | 2 |
| **Number of Credible Sets: SuSiE (95% CS)** | 2 |
| **T1/T2 + Stepwise TF Overlap (Yes/No)** | Yes (4 TFBS with TFs CDX2, CEBPG, FOXA2, and POU2F3) |
| **Implicated GTEx eGenes* and JIA PBMC eGenes† linked to JIA significant SNPs‡ or their proxies** | *AP4B1; BCL2L15; PHTF1* |
| **Implicated GTEx eGenes (All Tissues) linked to JIA significant SNPs‡ or their proxies** | *AP4B1; AP4B1-AS1; BCL2L15; DCLRE1B; HIPK1; HIPK1-AS1; OLFML3; PHTF1; PTPN22; RSBN1; SYT6; TRIM33* |
| **DGIdb has drug-gene interactions between GTEx eGenes* and JIA PBMC eGenes†** | No |
| **Region Implicated in Autoimmune Disease (Yes/No)** | Yes |

*Limited to EBV-transformed lymphocytes, cultured fibroblasts, whole blood, and spleen tissues

†JIA eGenes for peripheral blood mononuclear cells (PBMCs) from Barnes et al. (PMID: 19565513) were defined as genes with $P_{eQTL} < 1\times10^{-4}$

‡JIA significant SNPs defined as SNPs with Best $P_{FDR} < 0.05$

### (b) *ATP8B2–IL6R* Region (Tier 1)

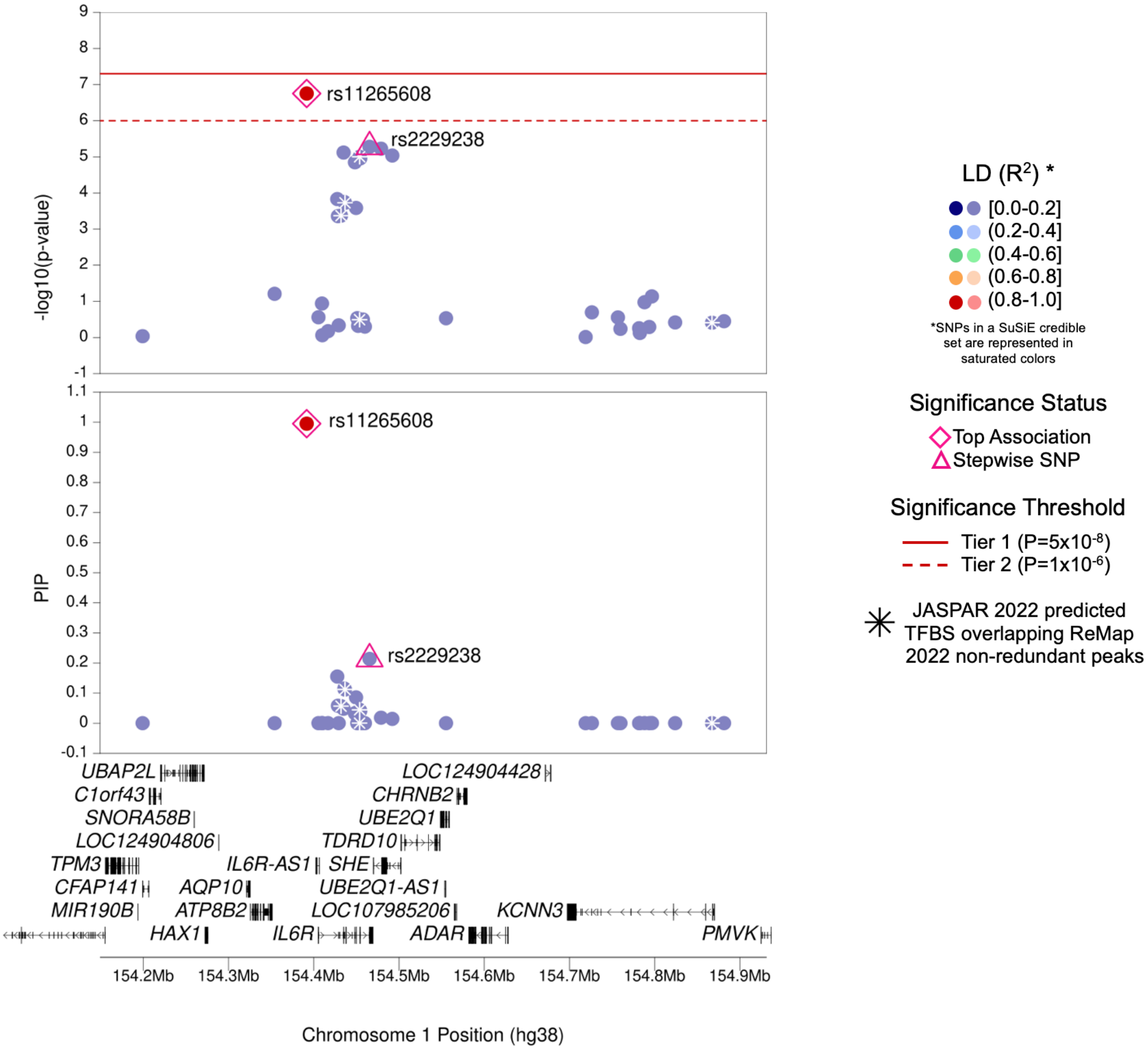

| | |
|---|---|
| **Region Name** | *ATP8B2–IL6R* |
| **Stepwise Region Window (hg38)** | chr1:153891664-154891664 |
| **Regional P-Value (df)** | P = $7.22 \times 10^{-10}$ (df = 2) |
| **Novel for JIA (Yes/No)** | No |
| **Number of Signals: Stepwise** | 2 |
| **Number of Credible Sets: SuSiE (95% CS)** | 1 |
| **T1/T2 + Stepwise TF Overlap (Yes/No)** | No |
| **Implicated GTEx eGenes* and JIA PBMC eGenes† linked to JIA significant SNPs‡ or their proxies** | *ATP8B2* |
| **Implicated GTEx eGenes (All Tissues) linked to JIA significant SNPs‡ or their proxies** | *ATP8B2; CHTOP; IL6R; NUP210L; RAB13; RP1-178F15.4; RP11-350G8.5; RP11-422P24.12; S100A1; S100A13; S100A14; SHE; TDRD10; UBE2Q1-AS1* |
| **DGIdb has drug-gene interactions between GTEx eGenes* and JIA PBMC eGenes†** | No |
| **Region Implicated in Autoimmune Disease (Yes/No)** | Yes |

*Limited to EBV-transformed lymphocytes, cultured fibroblasts, whole blood, and spleen tissues

†JIA eGenes for peripheral blood mononuclear cells (PBMCs) from Barnes et al. (PMID: 19565513) were defined as genes with $P_{eQTL} < 1 \times 10^{-4}$

‡JIA significant SNPs defined as SNPs with Best $P_{FDR} < 0.05$

### (c) *FASLG–TNFSF18* Region (Tier 1)

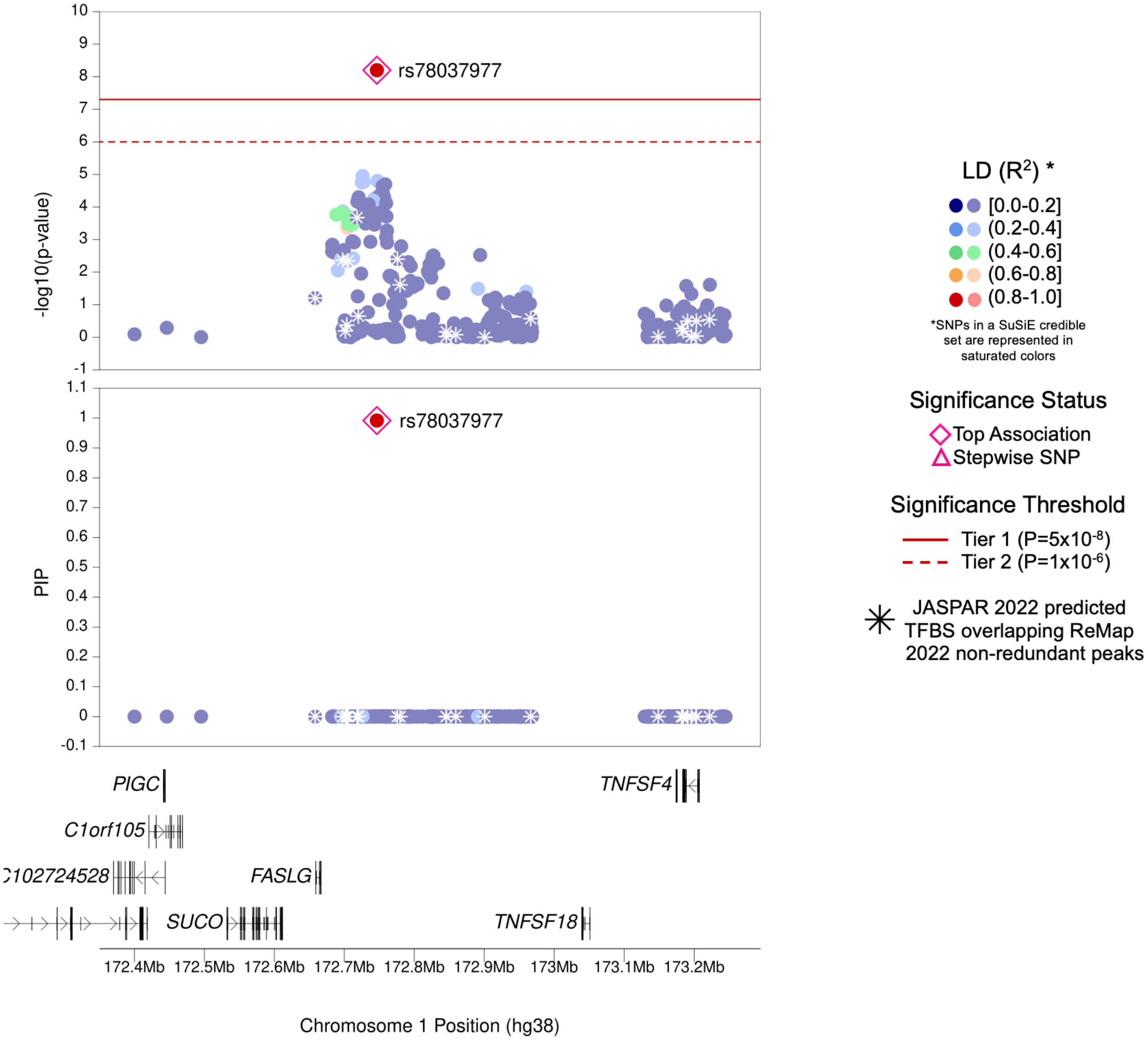

| | |
|---|---|
| **Region Name** | *FASLG–TNFSF18* |
| **Stepwise Region Window (hg38)** | chr1:172246562-173246562 |
| **Regional P-Value (df)** | P = 6.30x10$^{-9}$ (df = 1) |
| **Novel for JIA (Yes/No)** | **Yes** |
| **Number of Signals: Stepwise** | 1 |
| **Number of Credible Sets: SuSiE (95% CS)** | 1 |
| **T1/T2 + Stepwise TF Overlap (Yes/No)** | No |
| **Implicated GTEx eGenes* and JIA PBMC eGenes† linked to JIA significant SNPs‡ or their proxies** | *SUCO* |
| **Implicated GTEx eGenes (All Tissues) linked to JIA significant SNPs‡ or their proxies** | *RP1-15D23.2; SUCO* |
| **DGIdb has drug-gene interactions between GTEx eGenes* and JIA PBMC eGenes†** | No |
| **Region Implicated in Autoimmune Disease (Yes/No)** | Yes |

*Limited to EBV-transformed lymphocytes, cultured fibroblasts, whole blood, and spleen tissues

†JIA eGenes for peripheral blood mononuclear cells (PBMCs) from Barnes et al. (PMID: 19565513) were defined as genes with $P_{eQTL} < 1x10^{-4}$

‡JIA significant SNPs defined as SNPs with Best $P_{FDR} < 0.05$

### (d) *AFF3–LINC01104* Region (Tier 1)

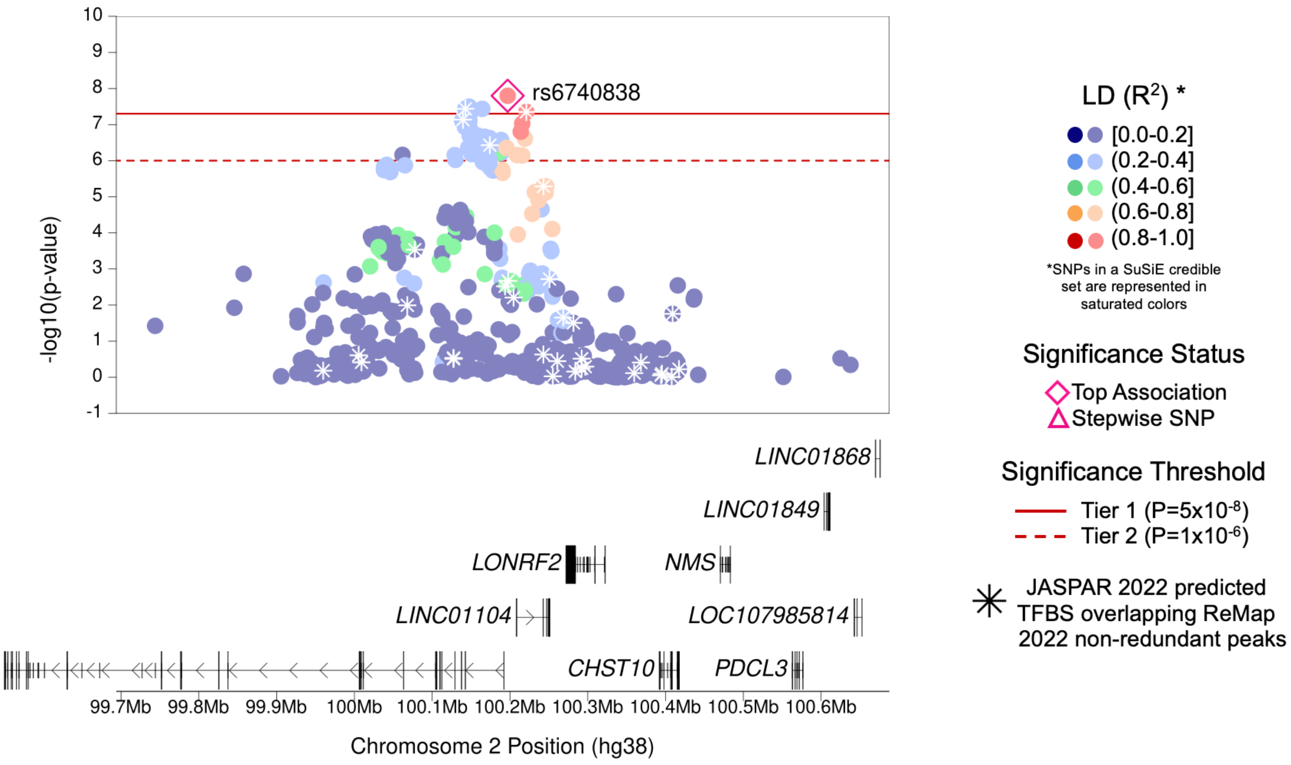

| | |
|---|---|
| **Region Name** | *AFF3–LINC01104* |
| **Stepwise Region Window (hg38)** | chr2:99697037-100697037 |
| **Regional P-Value (df)** | $P = 1.59 \times 10^{-8}$ (df = 1) |
| **Novel for JIA (Yes/No)** | No |
| **Number of Signals: Stepwise** | 1 |
| **Number of Credible Sets: SuSiE (95% CS)** | 0 (Large number of non-additive SNPs in region; SuSiE not computed) |
| **T1/T2 + Stepwise TF Overlap (Yes/No)** | Yes (4 TFBS with TFs SOX2, SPIB, SCRT1, and GATA2) |
| **Implicated GTEx eGenes* and JIA PBMC eGenes† linked to JIA significant SNPs‡ or their proxies** | *AFF3* |
| **Implicated GTEx eGenes (All Tissues) linked to JIA significant SNPs‡ or their proxies** | *AC092667.2; AFF3; EIF5B; LONRF2* |
| **DGIdb has drug-gene interactions between GTEx eGenes* and JIA PBMC eGenes†** | No |
| **Region Implicated in Autoimmune Disease (Yes/No)** | Yes |

*Limited to EBV-transformed lymphocytes, cultured fibroblasts, whole blood, and spleen tissues

†JIA eGenes for peripheral blood mononuclear cells (PBMCs) from Barnes et al. (PMID: 19565513) were defined as genes with $P_{eQTL} < 1 \times 10^{-4}$

‡JIA significant SNPs defined as SNPs with Best $P_{FDR} < 0.05$

### (e) *STAT4* Region (Tier 1)

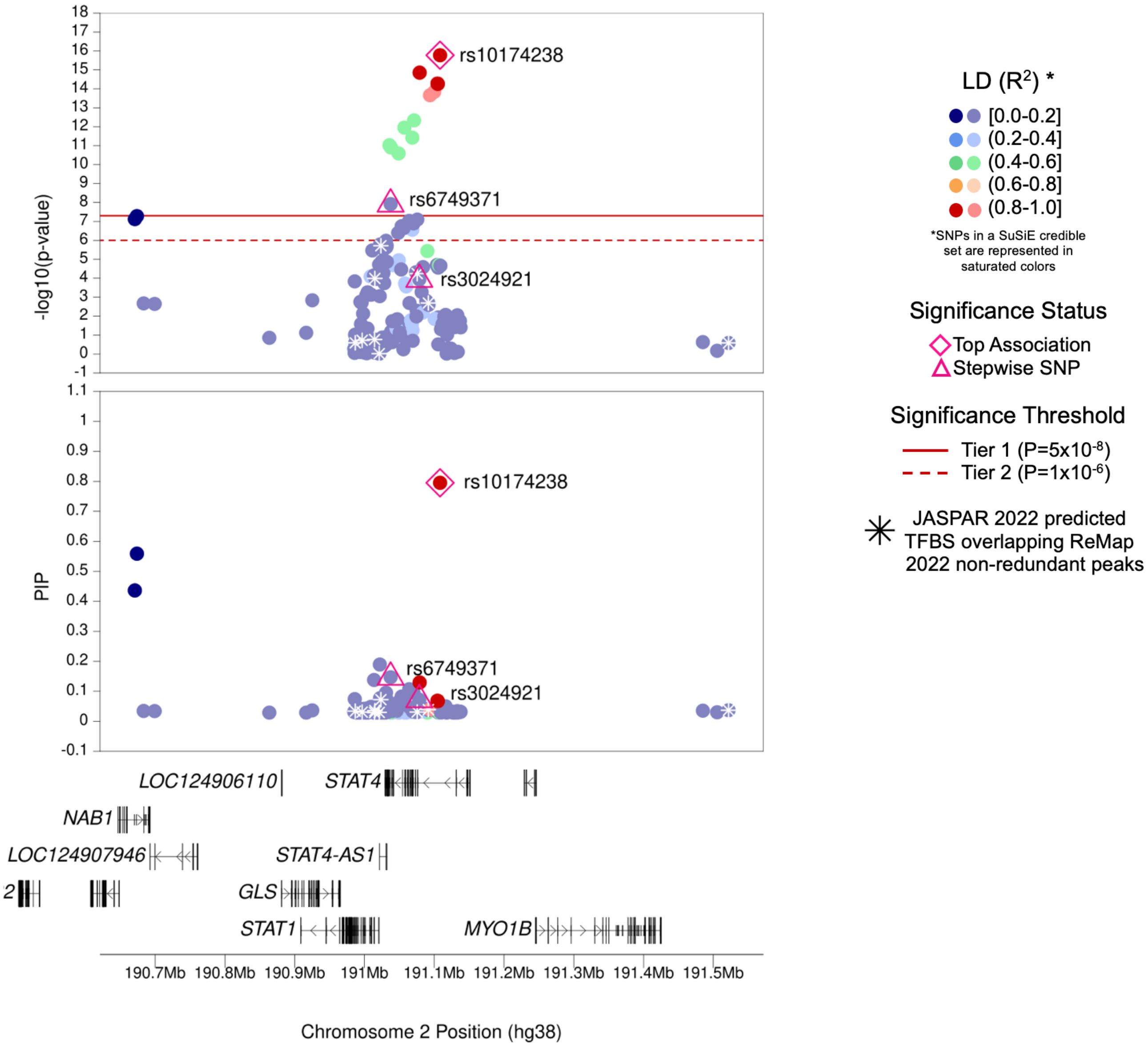

| | |
|---|---|
| **Region Name** | *STAT4* |
| **Stepwise Region Window (hg38)** | chr2:190608308-191608308 |
| **Regional P-Value (df)** | P = $1.35x10^{-23}$ (df = 3) |
| **Novel for JIA (Yes/No)** | No |
| **Number of Signals: Stepwise** | 3 |
| **Number of Credible Sets: SuSiE (95% CS)** | 2 |
| **T1/T2 + Stepwise TF Overlap (Yes/No)** | No |
| **Implicated GTEx eGenes* and JIA PBMC eGenes† linked to JIA significant SNPs‡ or their proxies** | *HIBCH* |
| **Implicated GTEx eGenes (All Tissues) linked to JIA significant SNPs‡ or their proxies** | *AC067945.4; AC093388.3; GLS; HIBCH; NEMP2; RP11-647K16.1; STAT1* |
| **DGIdb has drug-gene interactions between GTEx eGenes* and JIA PBMC eGenes†** | No |
| **Region Implicated in Autoimmune Disease (Yes/No)** | Yes |

*Limited to EBV-transformed lymphocytes, cultured fibroblasts, whole blood, and spleen tissues

†JIA eGenes for peripheral blood mononuclear cells (PBMCs) from Barnes et al. (PMID: 19565513) were defined as genes with $P_{eQTL} < 1x10^{-4}$

‡JIA significant SNPs defined as SNPs with Best $P_{FDR} < 0.05$

### (f) *CD28–CTLA4* Region (Tier 1)

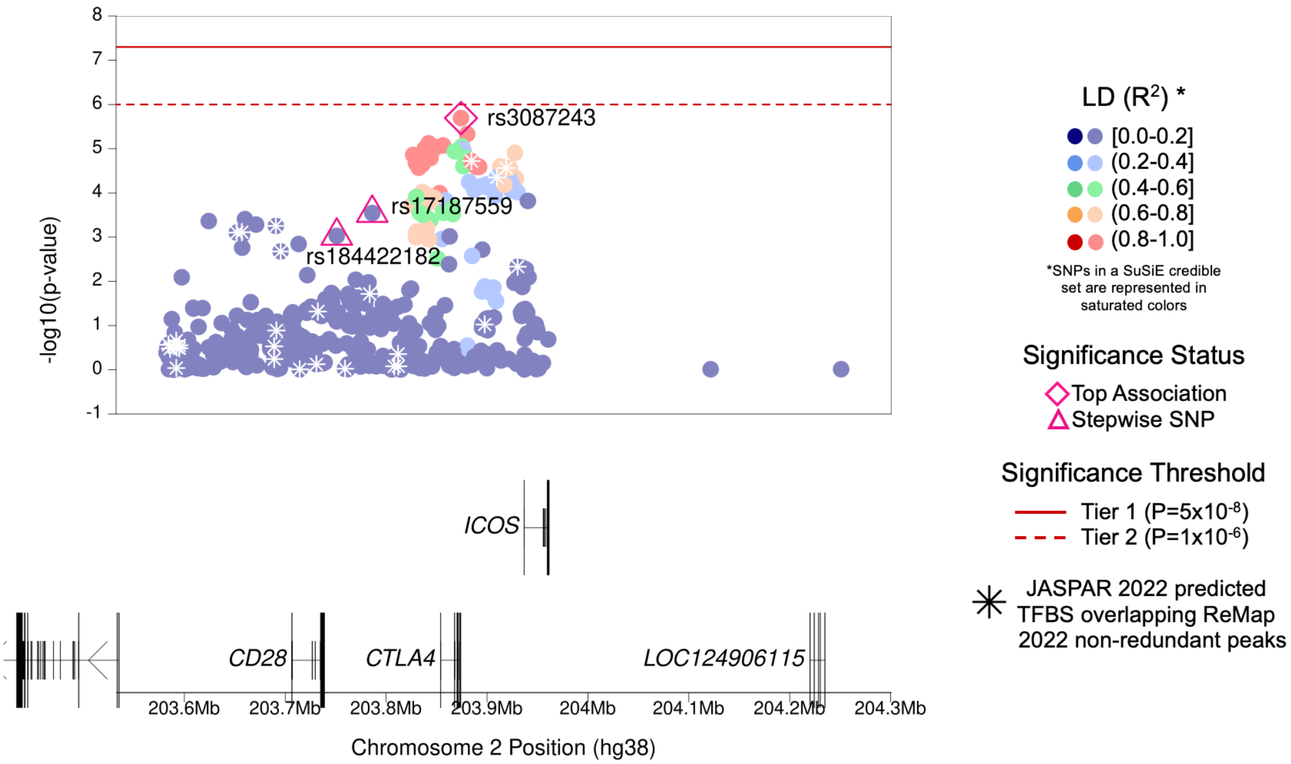

| | |
|---|---|
| **Region Name** | *CD28–CTLA4* |
| **Stepwise Region Window (hg38)** | chr2:203374196-204374196 |
| **Regional P-Value (df)** | $P = 2.05 \times 10^{-11}$ (df = 3) |
| **Novel for JIA (Yes/No)** | **Yes** |
| **Number of Signals: Stepwise** | 3 |
| **Number of Credible Sets: SuSiE (95% CS)** | 0 (Large number of non-additive SNPs in region; SuSiE not computed) |
| **T1/T2 + Stepwise TF Overlap (Yes/No)** | No |
| **Implicated GTEx eGenes* and JIA PBMC eGenes† linked to JIA significant SNPs‡ or their proxies** | NA (no eGenes) |
| **Implicated GTEx eGenes (All Tissues) linked to JIA significant SNPs‡ or their proxies** | *CARF; CTLA4; RAPH1* |
| **DGIdb has drug-gene interactions between GTEx eGenes* and JIA PBMC eGenes†** | No |
| **Region Implicated in Autoimmune Disease (Yes/No)** | Yes |

*Limited to EBV-transformed lymphocytes, cultured fibroblasts, whole blood, and spleen tissues

†JIA eGenes for peripheral blood mononuclear cells (PBMCs) from Barnes et al. (PMID: 19565513) were defined as genes with $P_{eQTL} < 1 \times 10^{-4}$

‡JIA significant SNPs defined as SNPs with Best $P_{FDR} < 0.05$

### (g) *CCR2–CCL2* Region (Tier 1)

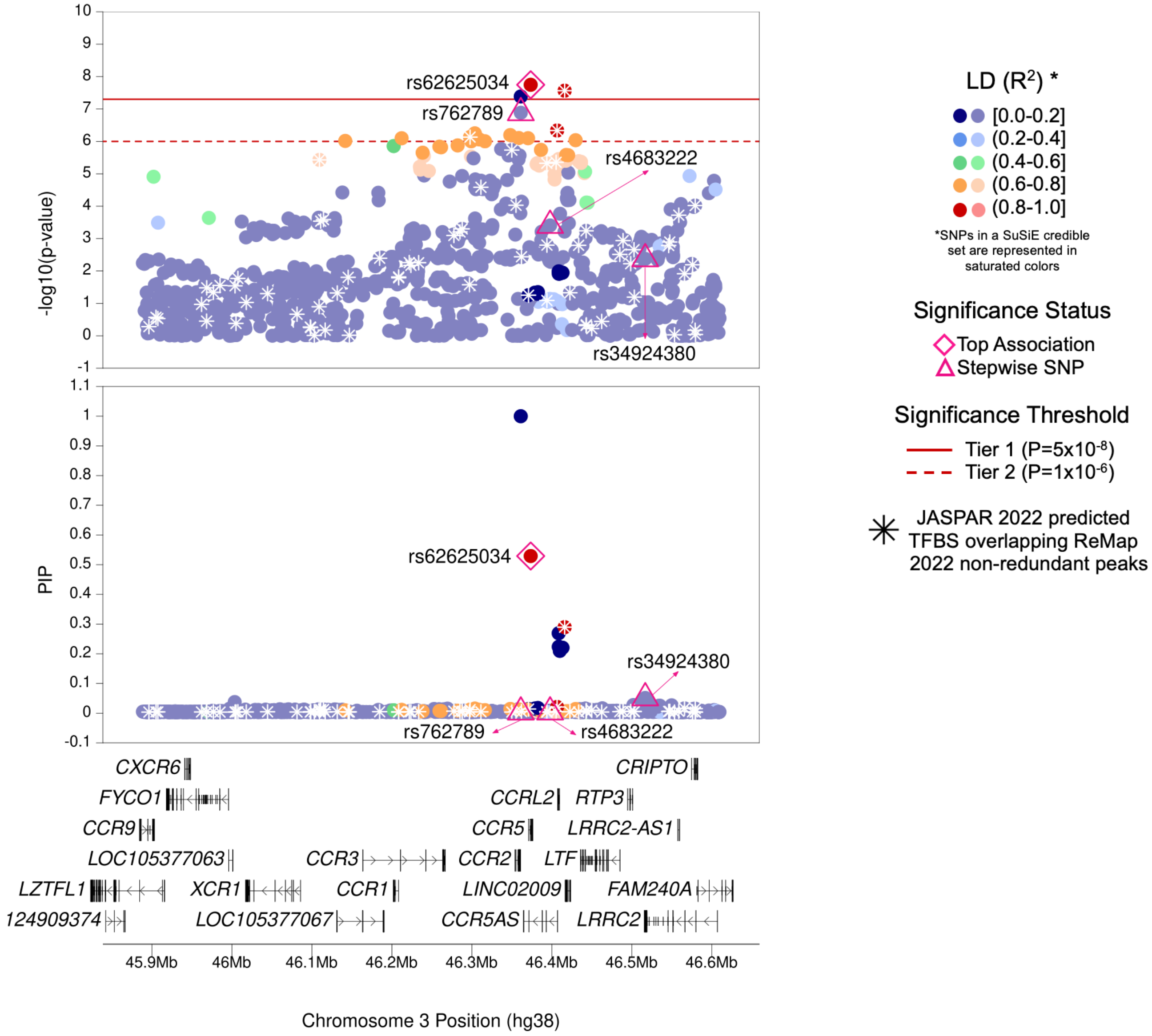

| | |
|---|---|
| **Region Name** | *CCR2–CCL2* |
| **Stepwise Region Window (hg38)** | chr3:45873484-46873484 |
| **Regional P-Value (df)** | $P = 1.69 \times 10^{-16}$ (df = 4) |
| **Novel for JIA (Yes/No)** | No |
| **Number of Signals: Stepwise** | 4 |
| **Number of Credible Sets: SuSiE (95% CS)** | 3 |
| **T1/T2 + Stepwise TF Overlap (Yes/No)** | Yes (1 TFBS with TF ONECUT1) |
| **Implicated GTEx eGenes* and JIA PBMC eGenes† linked to JIA significant SNPs‡ or their proxies** | *CCR1; CCR2; CCR3; LRRC2* |
| **Implicated GTEx eGenes (All Tissues) linked to JIA significant SNPs‡ or their proxies** | *ALS2CL; CCR1; CCR2; CCR3; CCR5; CCRL2; CXCR6; FYCO1; KIF9-AS1; LARS2; LIMD1; LINC02009; LRRC2; LRRC2-AS1; LTF; PRSS42; PTH1R; RP11-24F11.2; RP11-509I21.2; RTP3; TDGF1* |
| **DGIdb has drug-gene interactions between GTEx eGenes* and JIA PBMC eGenes†** | Yes |
| **Region Implicated in Autoimmune Disease (Yes/No)** | Yes |

*Limited to EBV-transformed lymphocytes, cultured fibroblasts, whole blood, and spleen tissues

†JIA eGenes for peripheral blood mononuclear cells (PBMCs) from Barnes et al. (PMID: 19565513) were defined as genes with $P_{eQTL} < 1 \times 10^{-4}$

‡JIA significant SNPs defined as SNPs with Best $P_{FDR} < 0.05$

### (h) *IL2–IL21* Region (Tier 1)

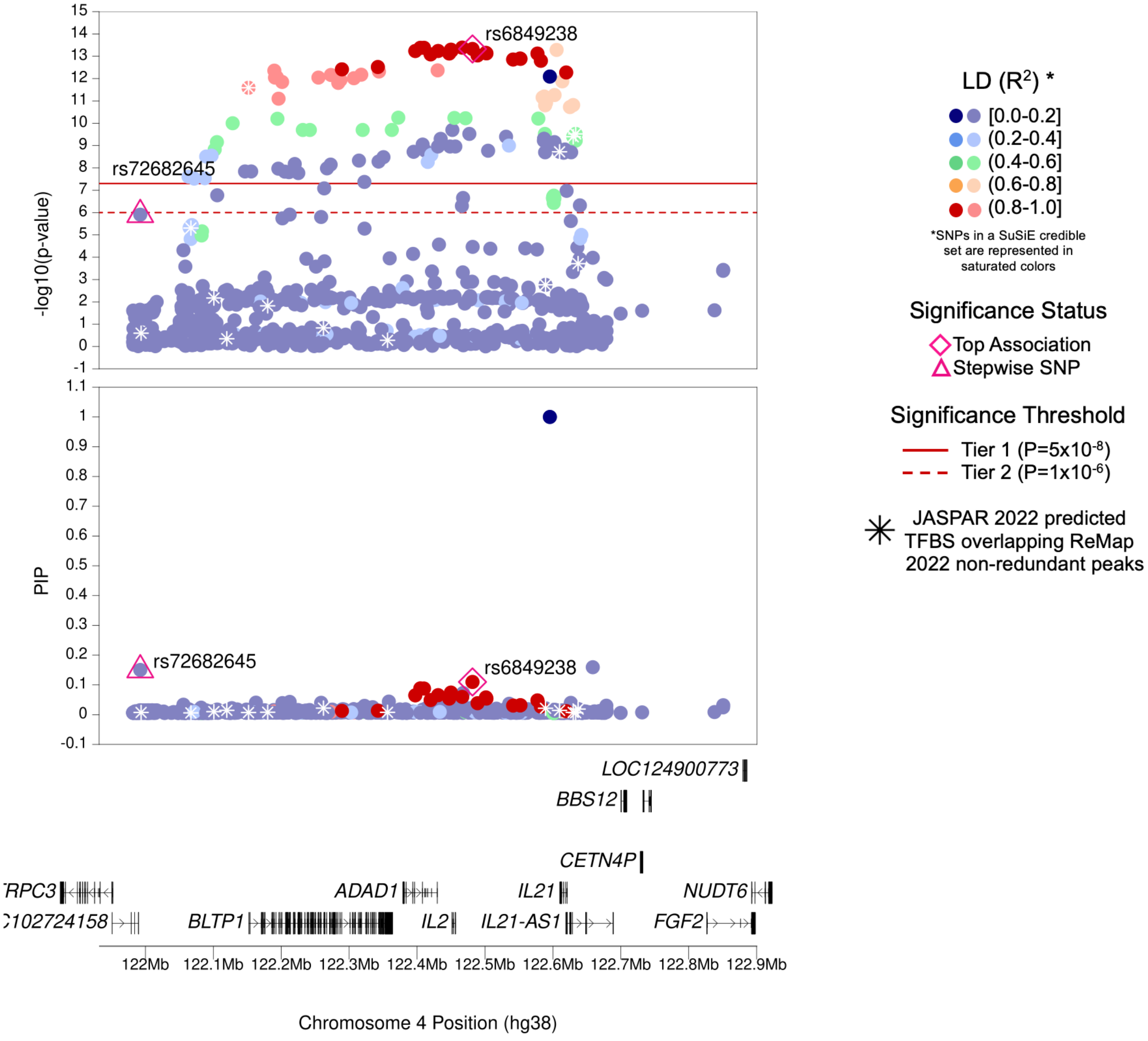

| | |
|---|---|
| **Region Name** | *IL2—IL21* |
| **Stepwise Region Window (hg38)** | chr4:121981615-122981615 |
| **Regional P-Value (df)** | $P = 8.75x10^{-17}$ (df = 2) |
| **Novel for JIA (Yes/No)** | No |
| **Number of Signals: Stepwise** | 2 |
| **Number of Credible Sets: SuSiE (95% CS)** | 2 |
| **T1/T2 + Stepwise TF Overlap (Yes/No)** | Yes (2 TFBS with TFs SPIB and TEAD4) |
| **Implicated GTEx eGenes* and JIA PBMC eGenes† linked to JIA significant SNPs‡ or their proxies** | NA (No eGenes) |
| **Implicated GTEx eGenes (All Tissues) linked to JIA significant SNPs‡ or their proxies** | *CETN4P; FGF2; IL21-AS1; KIAA1109* |
| **DGIdb has drug-gene interactions between GTEx eGenes* and JIA PBMC eGenes†** | No |
| **Region Implicated in Autoimmune Disease (Yes/No)** | Yes |

*Limited to EBV-transformed lymphocytes, cultured fibroblasts, whole blood, and spleen tissues

†JIA eGenes for peripheral blood mononuclear cells (PBMCs) from Barnes et al. (PMID: 19565513) were defined as genes with $P_{eQTL} < 1x10^{-4}$

‡JIA significant SNPs defined as SNPs with Best $P_{FDR} < 0.05$

### (i) *ANKRD55* Region (Tier 1)

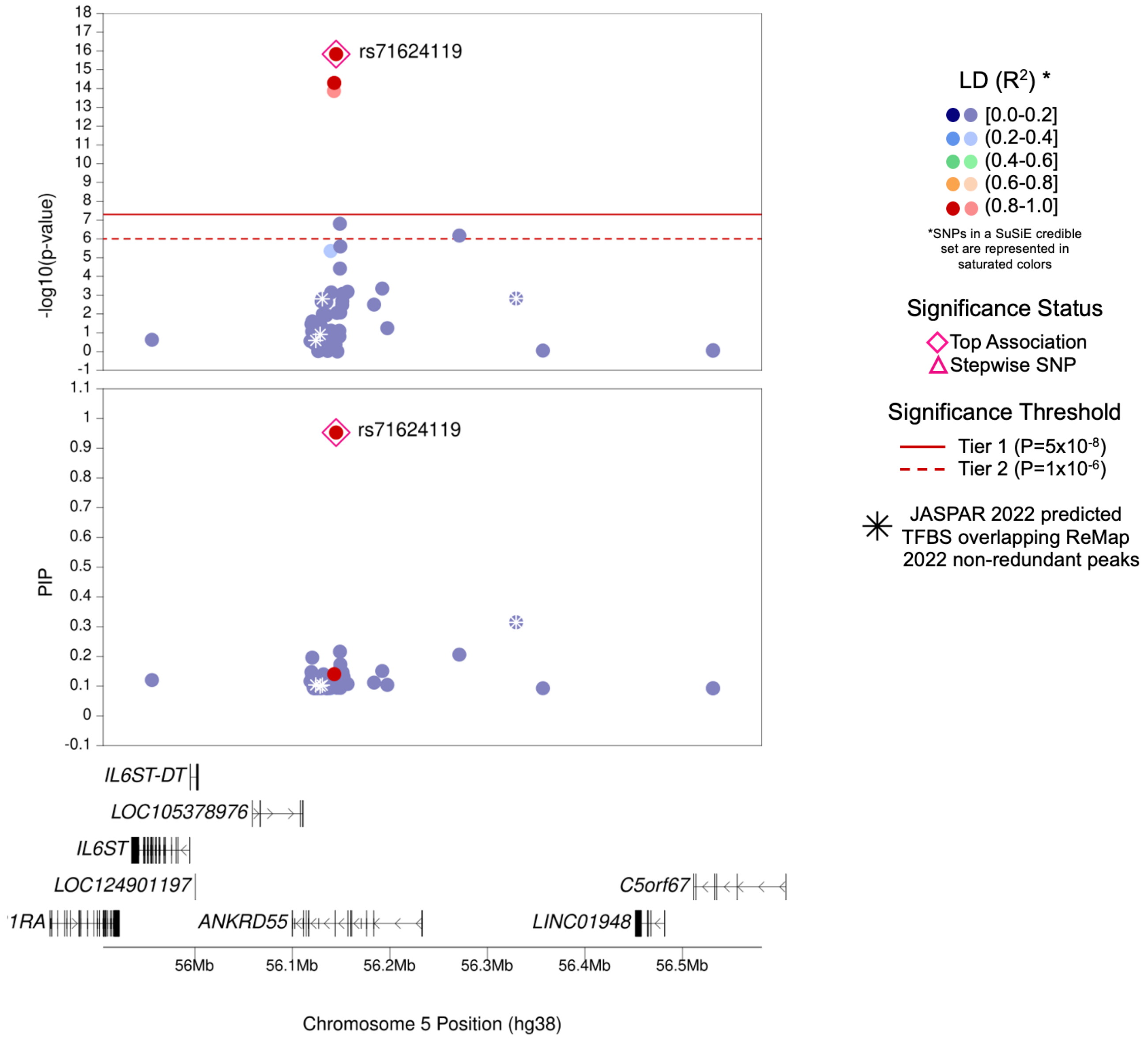

| | |
|---|---|
| **Region Name** | *ANKRD55* |
| **Stepwise Region Window (hg38)** | chr5:55644903-56644903 |
| **Regional P-Value (df)** | P = $1.11 \times 10^{-16}$ (df = 1) |
| **Novel for JIA (Yes/No)** | No |
| **Number of Signals: Stepwise** | 1 |
| **Number of Credible Sets: SuSiE (95% CS)** | 1 |
| **T1/T2 + Stepwise TF Overlap (Yes/No)** | No |
| **Implicated GTEx eGenes* and JIA PBMC eGenes† linked to JIA significant SNPs‡ or their proxies** | *ANKRD55* |
| **Implicated GTEx eGenes (All Tissues) linked to JIA significant SNPs‡ or their proxies** | *ANKRD55; IL6ST* |
| **DGIdb has drug-gene interactions between GTEx eGenes* and JIA PBMC eGenes†** | No |
| **Region Implicated in Autoimmune Disease (Yes/No)** | Yes |

*Limited to EBV-transformed lymphocytes, cultured fibroblasts, whole blood, and spleen tissues

†JIA eGenes for peripheral blood mononuclear cells (PBMCs) from Barnes et al. (PMID: 19565513) were defined as genes with $P_{eQTL} < 1 \times 10^{-4}$

‡JIA significant SNPs defined as SNPs with Best $P_{FDR} < 0.05$

### (j) *ERAP2–LNPEP* Region (Tier 1)

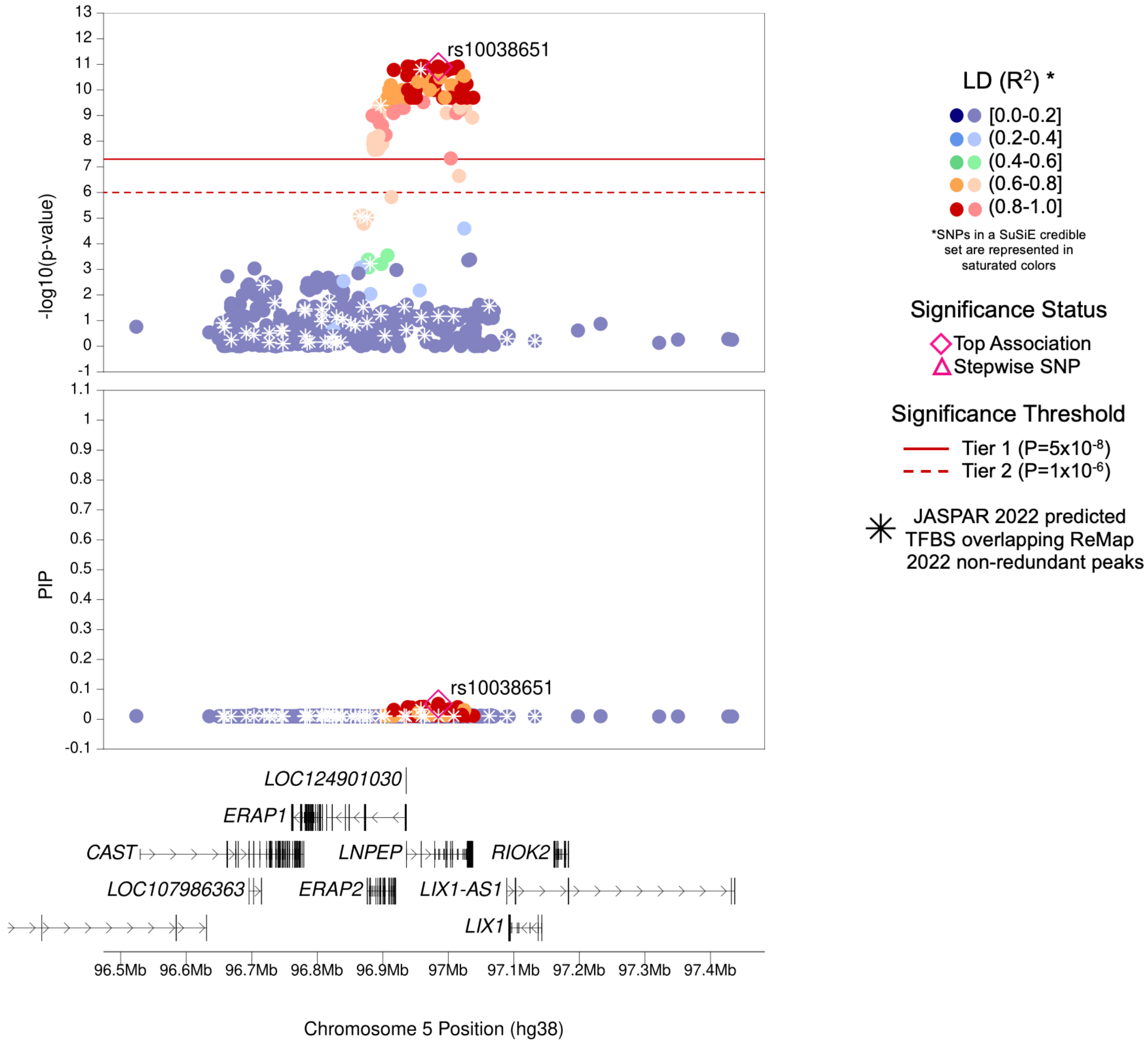

| | |
|---|---|
| **Region Name** | *ERAP2–LNPEP* |
| **Stepwise Region Window (hg38)** | chr5:96484707-97484707 |
| **Regional P-Value (df)** | $P = 1.25 \times 10^{-11}$ (df = 1) |
| **Novel for JIA (Yes/No)** | No |
| **Number of Signals: Stepwise** | 1 |
| **Number of Credible Sets: SuSiE (95% CS)** | 1 |
| **T1/T2 + Stepwise TF Overlap (Yes/No)** | No |
| **Implicated GTEx eGenes* and JIA PBMC eGenes† linked to JIA significant SNPs‡ or their proxies** | *ERAP1; ERAP2; LNPEP* |
| **Implicated GTEx eGenes (All Tissues) linked to JIA significant SNPs‡ or their proxies** | *CAST; CTD-2260A17.1; CTD-2260A17.2; CTD-2260A17.3; ERAP1; ERAP2; LIX1; LNPEP* |
| **DGIdb has drug-gene interactions between GTEx eGenes* and JIA PBMC eGenes†** | No |
| **Region Implicated in Autoimmune Disease (Yes/No)** | Yes |

*Limited to EBV-transformed lymphocytes, cultured fibroblasts, whole blood, and spleen tissues

†JIA eGenes for peripheral blood mononuclear cells (PBMCs) from Barnes et al. (PMID: 19565513) were defined as genes with $P_{eQTL} < 1 \times 10^{-4}$

‡JIA significant SNPs defined as SNPs with Best $P_{FDR} < 0.05$

### (k) *C5orf56–IRF1* Region (Tier 1)

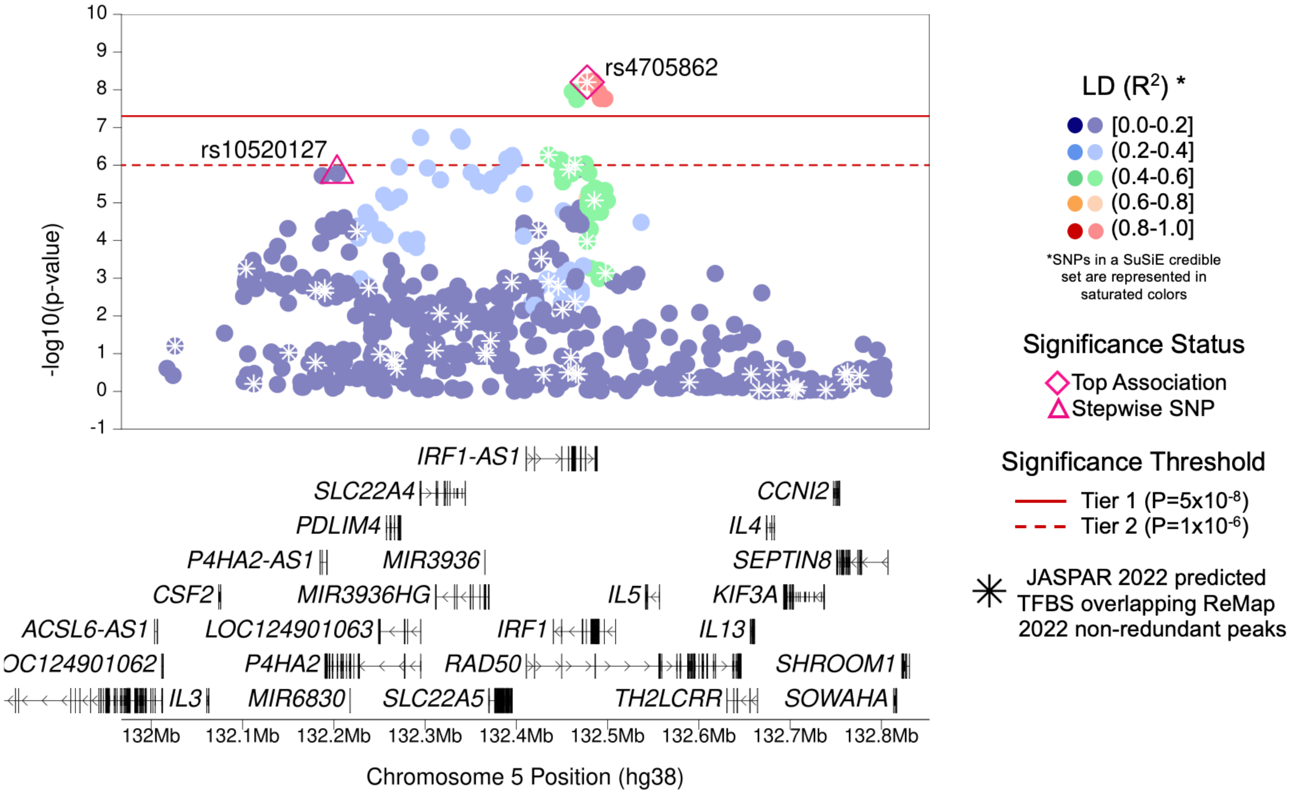

| | |
|---|---|
| **Region Name** | *C5orf56–IRF1* |
| **Stepwise Region Window (hg38)** | chr5:131977527-132977527 |
| **Regional P-Value (df)** | $P = 3.22x10^{-11}$ (df = 2) |
| **Novel for JIA (Yes/No)** | No |
| **Number of Signals: Stepwise** | 2 |
| **Number of Credible Sets: SuSiE (95% CS)** | 0 (Large number of non-additive SNPs in region; SuSiE not computed) |
| **T1/T2 + Stepwise TF Overlap (Yes/No)** | Yes (1 TFBS with TF NR2C2) |
| **Implicated GTEx eGenes* and JIA PBMC eGenes† linked to JIA significant SNPs‡ or their proxies** | *AC116366.6; C5orf56; MEIKIN; PDLIM4; RAD50; SLC22A4; SLC22A5* |
| **Implicated GTEx eGenes (All Tissues) linked to JIA significant SNPs‡ or their proxies** | *AC034220.3; AC116366.6; ACSL6; C5orf56; IL13; IRF1; MEIKIN; P4HA2; PDLIM4; RAD50; RAPGEF6; SEPT8; SLC22A4; SLC22A5* |
| **DGIdb has drug-gene interactions between GTEx eGenes* and JIA PBMC eGenes†** | Yes |
| **Region Implicated in Autoimmune Disease (Yes/No)** | Yes |

*Limited to EBV-transformed lymphocytes, cultured fibroblasts, whole blood, and spleen tissues

†JIA eGenes for peripheral blood mononuclear cells (PBMCs) from Barnes et al. (PMID: 19565513) were defined as genes with $P_{eQTL} < 1x10^{-4}$

‡JIA significant SNPs defined as SNPs with Best $P_{FDR} < 0.05$

#### (I) *MYB–AHI1* Region (Tier 1)

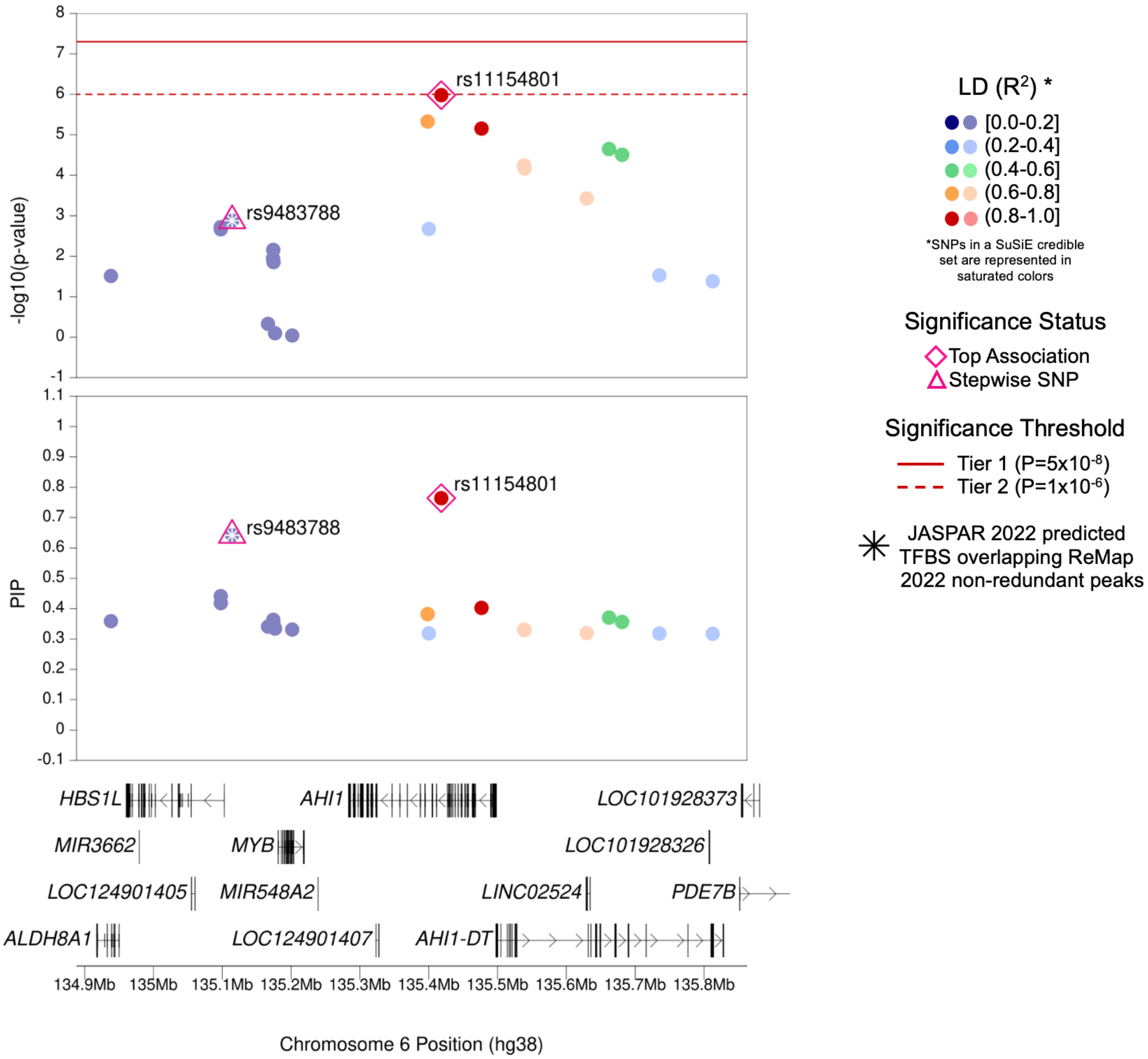

| | |
|---|---|
| **Region Name** | *MYB–AHI1* |
| **Stepwise Region Window (hg38)** | chr6:134918217-135918217 |
| **Regional P-Value (df)** | $P = 2.66x10^{-8}$ (df = 2) |
| **Novel for JIA (Yes/No)** | No |
| **Number of Signals: Stepwise** | 2 |
| **Number of Credible Sets: SuSiE (95% CS)** | 1 |
| **T1/T2 + Stepwise TF Overlap (Yes/No)** | Yes (2 TFBS with TFs MEF2A and MEF2C) |
| **Implicated GTEx eGenes* and JIA PBMC eGenes† linked to JIA significant SNPs‡ or their proxies** | *AHI1; ALDH8A1; HBS1L; RP3-388E23.2* |
| **Implicated GTEx eGenes (All Tissues) linked to JIA significant SNPs‡ or their proxies** | *AHI1; ALDH8A1; CTA-212D2.2; HBS1L; LINC00271; RP1-38C16.2; RP3-388E23.2* |
| **DGIdb has drug-gene interactions between GTEx eGenes* and JIA PBMC eGenes†** | No |
| **Region Implicated in Autoimmune Disease (Yes/No)** | Yes |

*Limited to EBV-transformed lymphocytes, cultured fibroblasts, whole blood, and spleen tissues

†JIA eGenes for peripheral blood mononuclear cells (PBMCs) from Barnes et al. (PMID: 19565513) were defined as genes with $P_{eQTL} < 1x10^{-4}$

‡JIA significant SNPs defined as SNPs with Best $P_{FDR} < 0.05$

### (m) *IL6* Region (Tier 1)

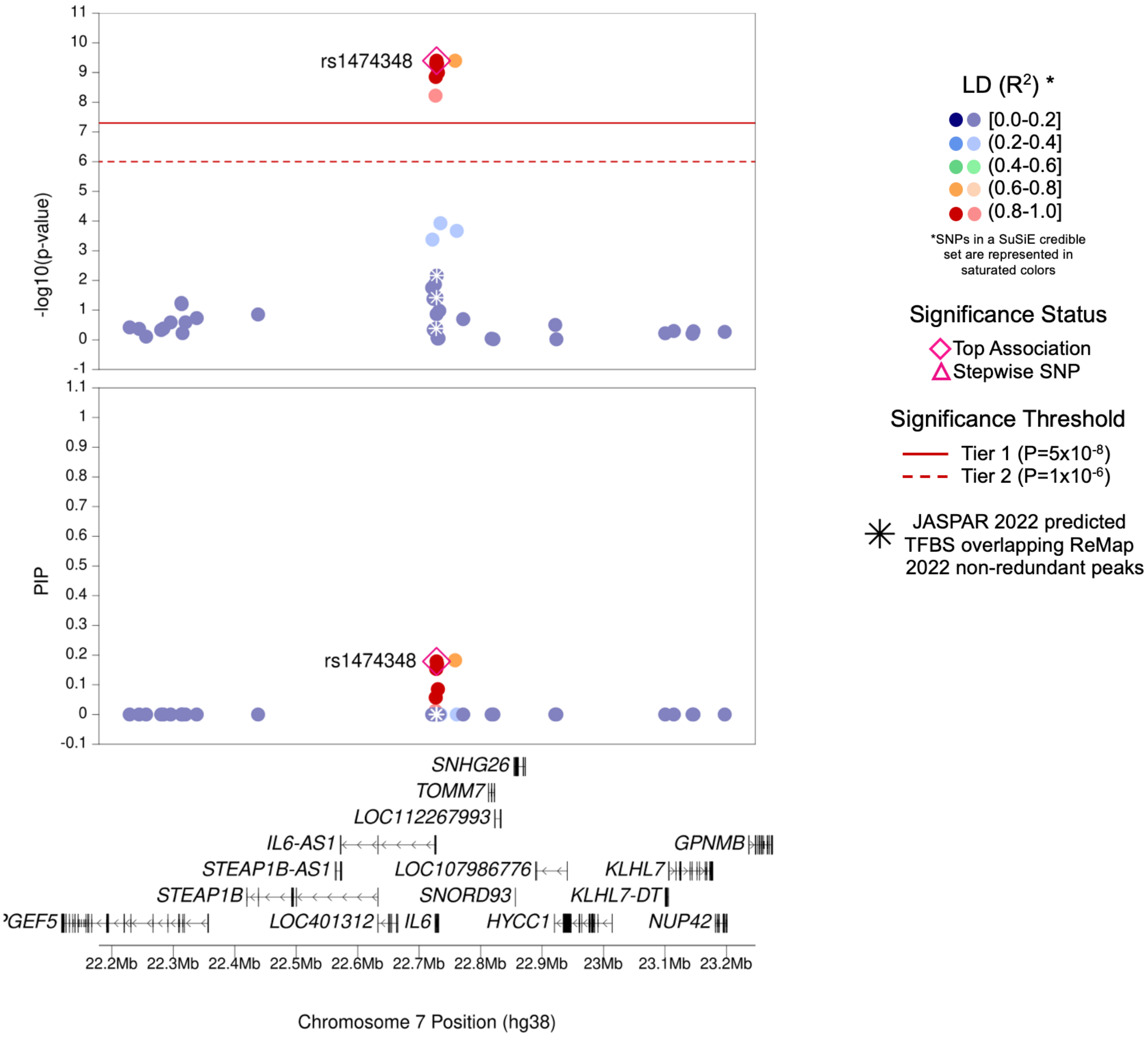

| | |
|---|---|
| **Region Name** | *IL6* |
| **Stepwise Region Window (hg38)** | chr7:22228289-23228289 |
| **Regional P-Value (df)** | P = $4.15 \times 10^{-10}$ (df = 1) |
| **Novel for JIA (Yes/No)** | No |
| **Number of Signals: Stepwise** | 1 |
| **Number of Credible Sets: SuSiE (95% CS)** | 1 |
| **T1/T2 + Stepwise TF Overlap (Yes/No)** | No |
| **Implicated GTEx eGenes* and JIA PBMC eGenes† linked to JIA significant SNPs‡ or their proxies** | *AC073072.5; IL6* |
| **Implicated GTEx eGenes (All Tissues) linked to JIA significant SNPs‡ or their proxies** | *AC002480.3; AC073072.5; CCDC126; FAM126A; IL6; STEAP1B* |
| **DGIdb has drug-gene interactions between GTEx eGenes* and JIA PBMC eGenes†** | Yes |
| **Region Implicated in Autoimmune Disease (Yes/No)** | Yes |

*Limited to EBV-transformed lymphocytes, cultured fibroblasts, whole blood, and spleen tissues

†JIA eGenes for peripheral blood mononuclear cells (PBMCs) from Barnes et al. (PMID: 19565513) were defined as genes with $P_{eQTL} < 1x10^{-4}$

‡JIA significant SNPs defined as SNPs with Best $P_{FDR} < 0.05$

### (n) *IL7* Region (Tier 1)

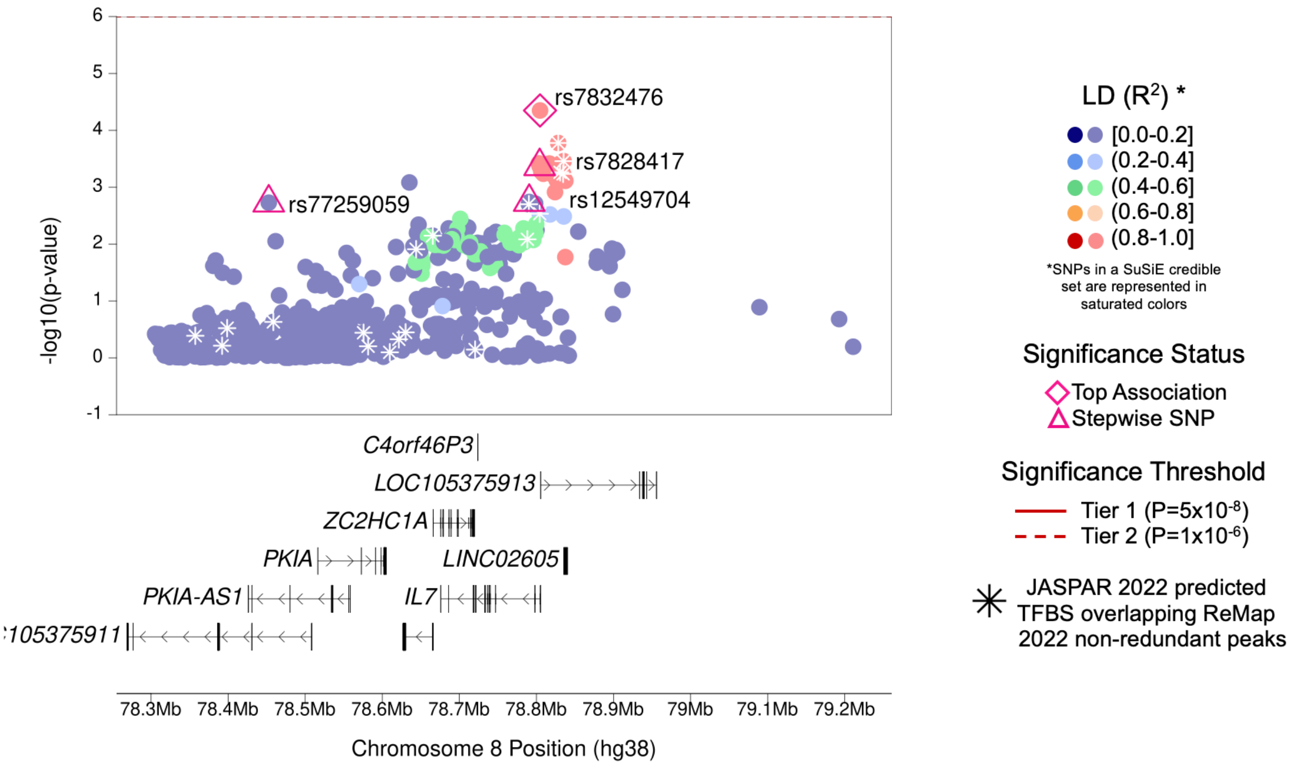

| | |
|---|---|
| **Region Name** | *IL7* |
| **Stepwise Region Window (hg38)** | chr8:78304682-79304682 |
| **Regional P-Value (df)** | $P = 4.70 \times 10^{-12}$ (df = 4) |
| **Novel for JIA (Yes/No)** | **Yes** |
| **Number of Signals: Stepwise** | 4 |
| **Number of Credible Sets: SuSiE (95% CS)** | 0 (Large number of non-additive SNPs in region; SuSiE not computed) |
| **T1/T2 + Stepwise TF Overlap (Yes/No)** | No |
| **Implicated GTEx eGenes* and JIA PBMC eGenes† linked to JIA significant SNPs‡ or their proxies** | *RP11-578O24.2; ZC2HC1A* |
| **Implicated GTEx eGenes (All Tissues) linked to JIA significant SNPs‡ or their proxies** | *IL7; RP11-578O24.2; THAP12P7; ZC2HC1A* |
| **DGIdb has drug-gene interactions between GTEx eGenes* and JIA PBMC eGenes†** | No |
| **Region Implicated in Autoimmune Disease (Yes/No)** | Yes |

*Limited to EBV-transformed lymphocytes, cultured fibroblasts, whole blood, and spleen tissues

†JIA eGenes for peripheral blood mononuclear cells (PBMCs) from Barnes et al. (PMID: 19565513) were defined as genes with $P_{eQTL} < 1 \times 10^{-4}$

‡JIA significant SNPs defined as SNPs with Best $P_{FDR} < 0.05$

### (o) *TNFSF15–TNFSF8* Region (Tier 1)

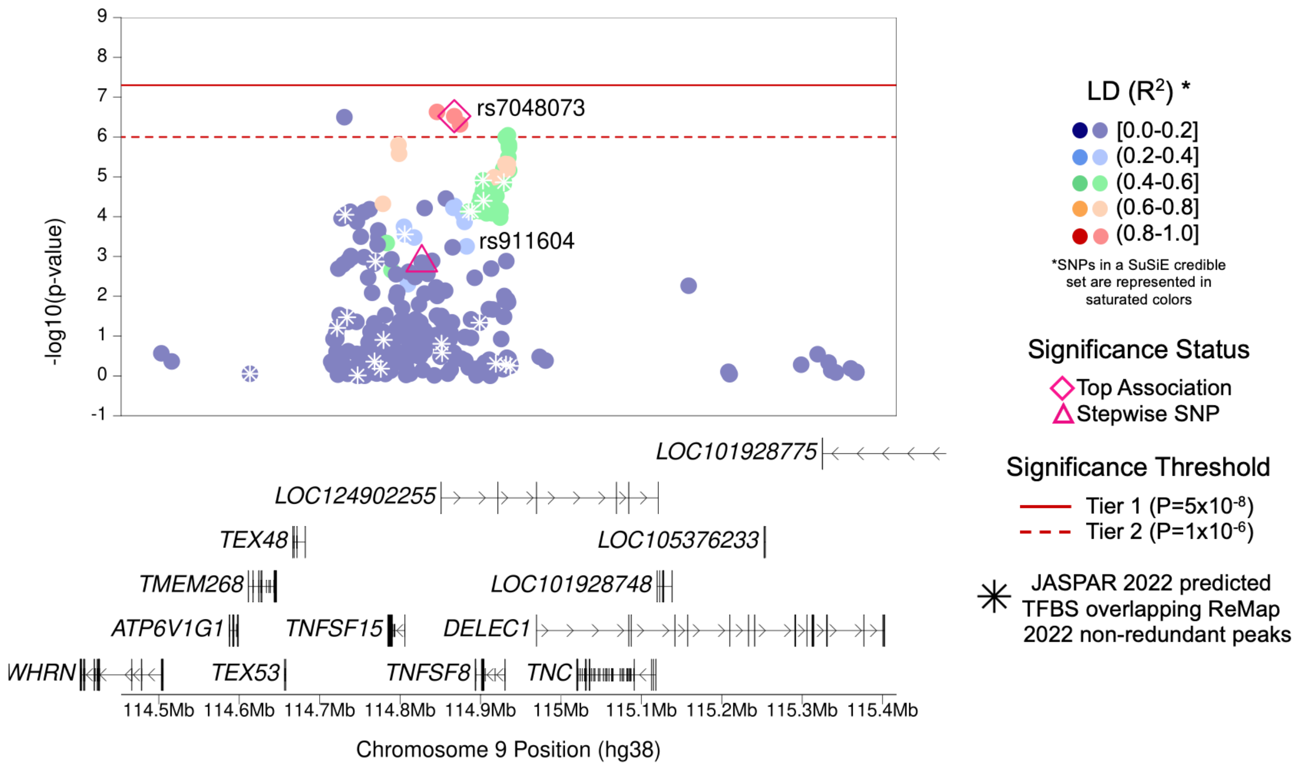

| | |
|---|---|
| **Region Name** | *TNFSF15–TNFSF8* |
| **Stepwise Region Window (hg38)** | chr9:114367409-115367409 |
| **Regional P-Value (df)** | P = $2.43 \times 10^{-9}$ (df = 2) |
| **Novel for JIA (Yes/No)** | **Yes** |
| **Number of Signals: Stepwise** | 2 |
| **Number of Credible Sets: SuSiE (95% CS)** | 0 (Large number of non-additive SNPs in region; SuSiE not computed) |
| **T1/T2 + Stepwise TF Overlap (Yes/No)** | No |
| **Implicated GTEx eGenes* and JIA PBMC eGenes† linked to JIA significant SNPs‡ or their proxies** | *TNFSF15* |
| **Implicated GTEx eGenes (All Tissues) linked to JIA significant SNPs‡ or their proxies** | *AMBP; TNFSF15* |
| **DGIdb has drug-gene interactions between GTEx eGenes* and JIA PBMC eGenes†** | No |
| **Region Implicated in Autoimmune Disease (Yes/No)** | Yes |

*Limited to EBV-transformed lymphocytes, cultured fibroblasts, whole blood, and spleen tissues

†JIA eGenes for peripheral blood mononuclear cells (PBMCs) from Barnes et al. (PMID: 19565513) were defined as genes with $P_{eQTL} < 1 \times 10^{-4}$

‡JIA significant SNPs defined as SNPs with Best $P_{FDR} < 0.05$

### (p) *IL2RA* Region (Tier 1)

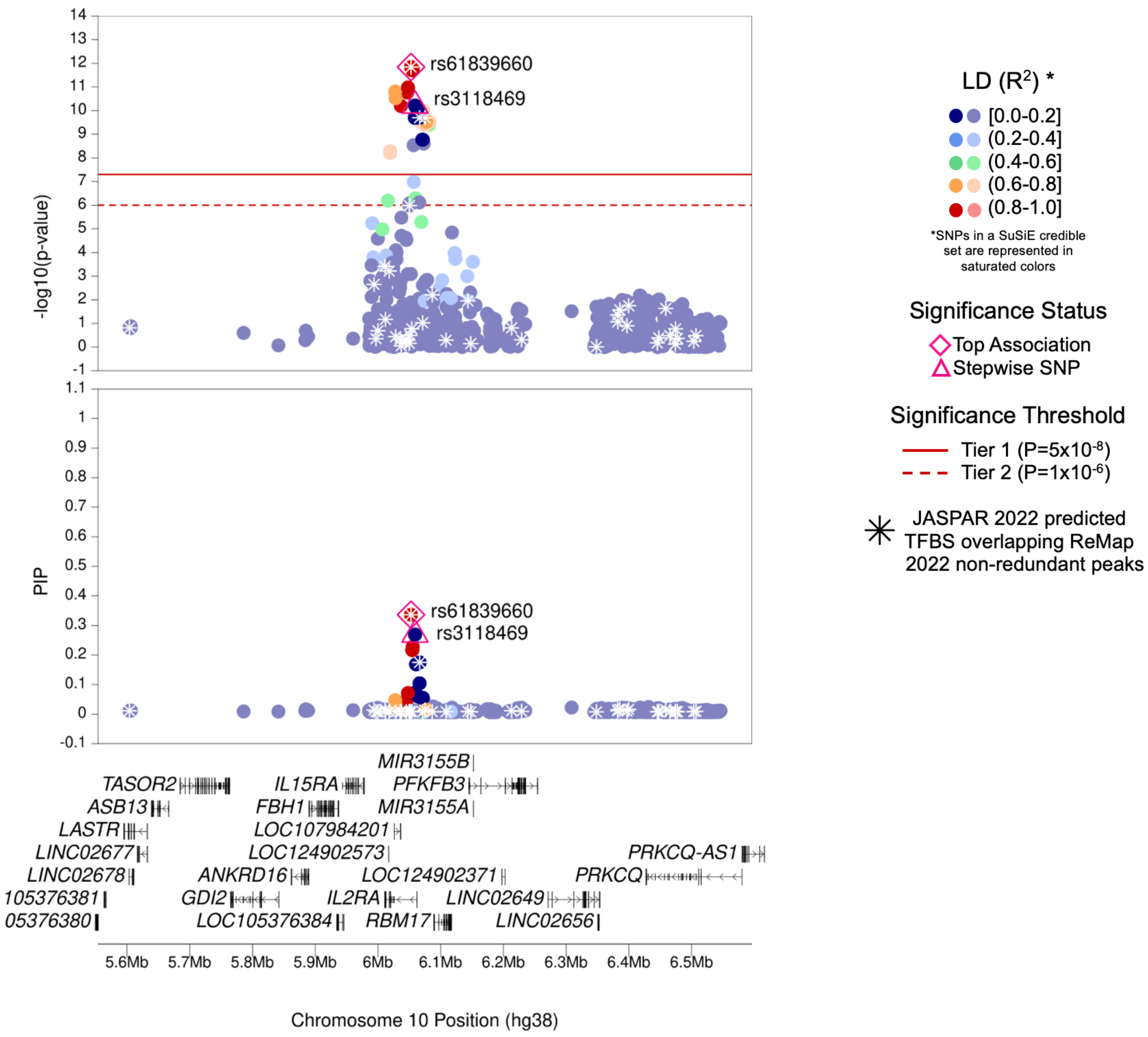

| | |
|---|---|
| **Region Name** | *IL2RA* |
| **Stepwise Region Window (hg38)** | chr10:5552734-6552734 |
| **Regional P-Value (df)** | P = $5.64 \times 10^{-18}$ (df = 2) |
| **Novel for JIA (Yes/No)** | No |
| **Number of Signals: Stepwise** | 2 |
| **Number of Credible Sets: SuSiE (95% CS)** | 2 |
| **T1/T2 + Stepwise TF Overlap (Yes/No)** | Yes (7 TFBS with TFs CREB1, FOSL1, FOSL2, FOS, JUND, MEF2A, MEF2C) |
| **Implicated GTEx eGenes* and JIA PBMC eGenes† linked to JIA significant SNPs‡ or their proxies** | NA (No eGenes) |
| **Implicated GTEx eGenes (All Tissues) linked to JIA significant SNPs‡ or their proxies** | *PFKFB3; RBM17* |
| **DGIdb has drug-gene interactions between GTEx eGenes* and JIA PBMC eGenes†** | No |
| **Region Implicated in Autoimmune Disease (Yes/No)** | Yes |

*Limited to EBV-transformed lymphocytes, cultured fibroblasts, whole blood, and spleen tissues

†JIA eGenes for peripheral blood mononuclear cells (PBMCs) from Barnes et al. (PMID: 19565513) were defined as genes with $P_{eQTL} < 1 \times 10^{-4}$

‡JIA significant SNPs defined as SNPs with Best $P_{FDR} < 0.05$

### (q) *ZNF365–EGR2* Region (Tier 1)

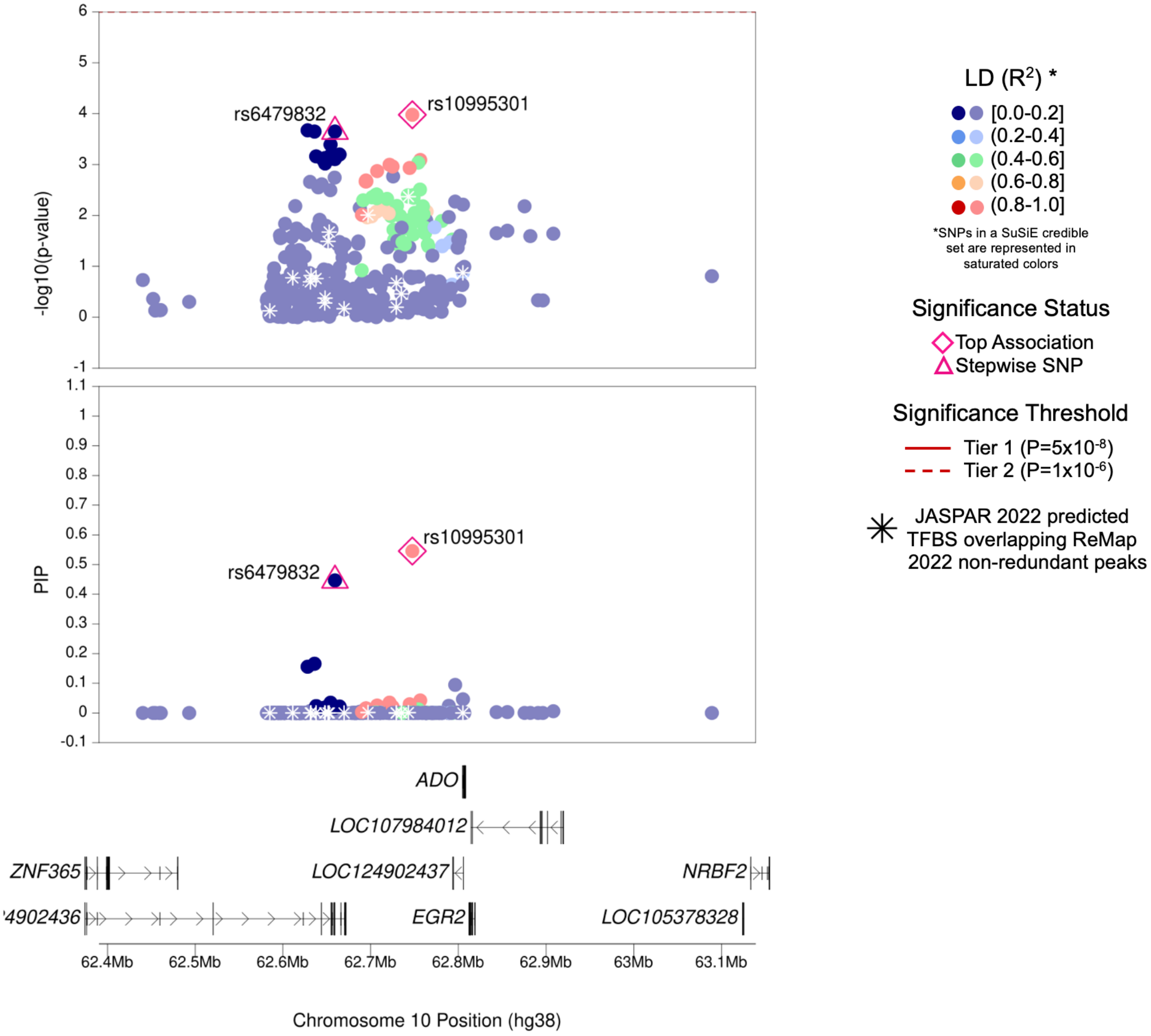

| | |
|---|---|
| **Region Name** | *ZNF365–EGR2* |
| **Stepwise Region Window (hg38)** | chr10:62247535-63247535 |
| **Regional P-Value (df)** | $P = 9.51 \times 10^{-9}$ (df = 2) |
| **Novel for JIA (Yes/No)** | **Yes** |
| **Number of Signals: Stepwise** | 2 |
| **Number of Credible Sets: SuSiE (95% CS)** | 1 |
| **T1/T2 + Stepwise TF Overlap (Yes/No)** | No |
| **Implicated GTEx eGenes* and JIA PBMC eGenes† linked to JIA significant SNPs‡ or their proxies** | *EGR2* |
| **Implicated GTEx eGenes (All Tissues) linked to JIA significant SNPs‡ or their proxies** | *ADO; EGR2; RP11-436D10.3; ZNF365* |
| **DGIdb has drug-gene interactions between GTEx eGenes* and JIA PBMC eGenes†** | Yes |
| **Region Implicated in Autoimmune Disease (Yes/No)** | Yes |

*Limited to EBV-transformed lymphocytes, cultured fibroblasts, whole blood, and spleen tissues

†JIA eGenes for peripheral blood mononuclear cells (PBMCs) from Barnes et al. (PMID: 19565513) were defined as genes with $P_{eQTL} < 1x10^{-4}$

‡JIA significant SNPs defined as SNPs with Best $P_{FDR} < 0.05$

### (r) *FAS* Region (Tier 1)

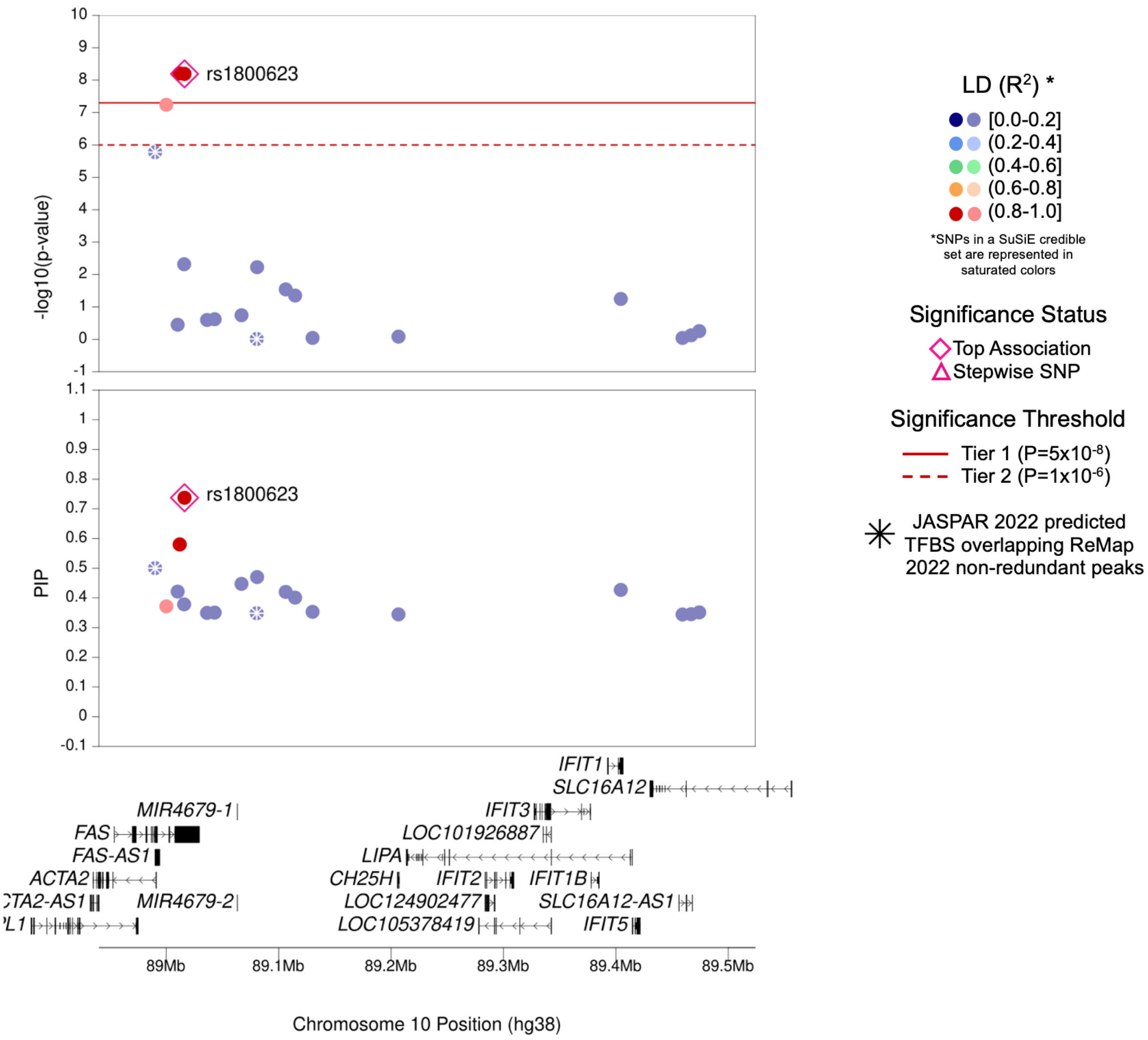

| | |
|---|---|
| **Region Name** | *FAS* |
| **Stepwise Region Window (hg38)** | chr10:88516326-89516326 |
| **Regional P-Value (df)** | $P = 6.36 \times 10^{-9}$ (df = 1) |
| **Novel for JIA (Yes/No)** | No |
| **Number of Signals: Stepwise** | 1 |
| **Number of Credible Sets: SuSiE (95% CS)** | 1 |
| **T1/T2 + Stepwise TF Overlap (Yes/No)** | No |
| **Implicated GTEx eGenes* and JIA PBMC eGenes† linked to JIA significant SNPs‡ or their proxies** | *ACTA2; FAS* |
| **Implicated GTEx eGenes (All Tissues) linked to JIA significant SNPs‡ or their proxies** | *ACTA2; FAS; FAS-AS1; RP11-399O19.9* |
| **DGIdb has drug-gene interactions between GTEx eGenes* and JIA PBMC eGenes†** | Yes |
| **Region Implicated in Autoimmune Disease (Yes/No)** | Yes |

*Limited to EBV-transformed lymphocytes, cultured fibroblasts, whole blood, and spleen tissues

†JIA eGenes for peripheral blood mononuclear cells (PBMCs) from Barnes et al. (PMID: 19565513) were defined as genes with $P_{eQTL} < 1\text{x}10^{-4}$

‡JIA significant SNPs defined as SNPs with Best $P_{FDR} < 0.05$

### (s) *PRR5L* Region (Tier 1)

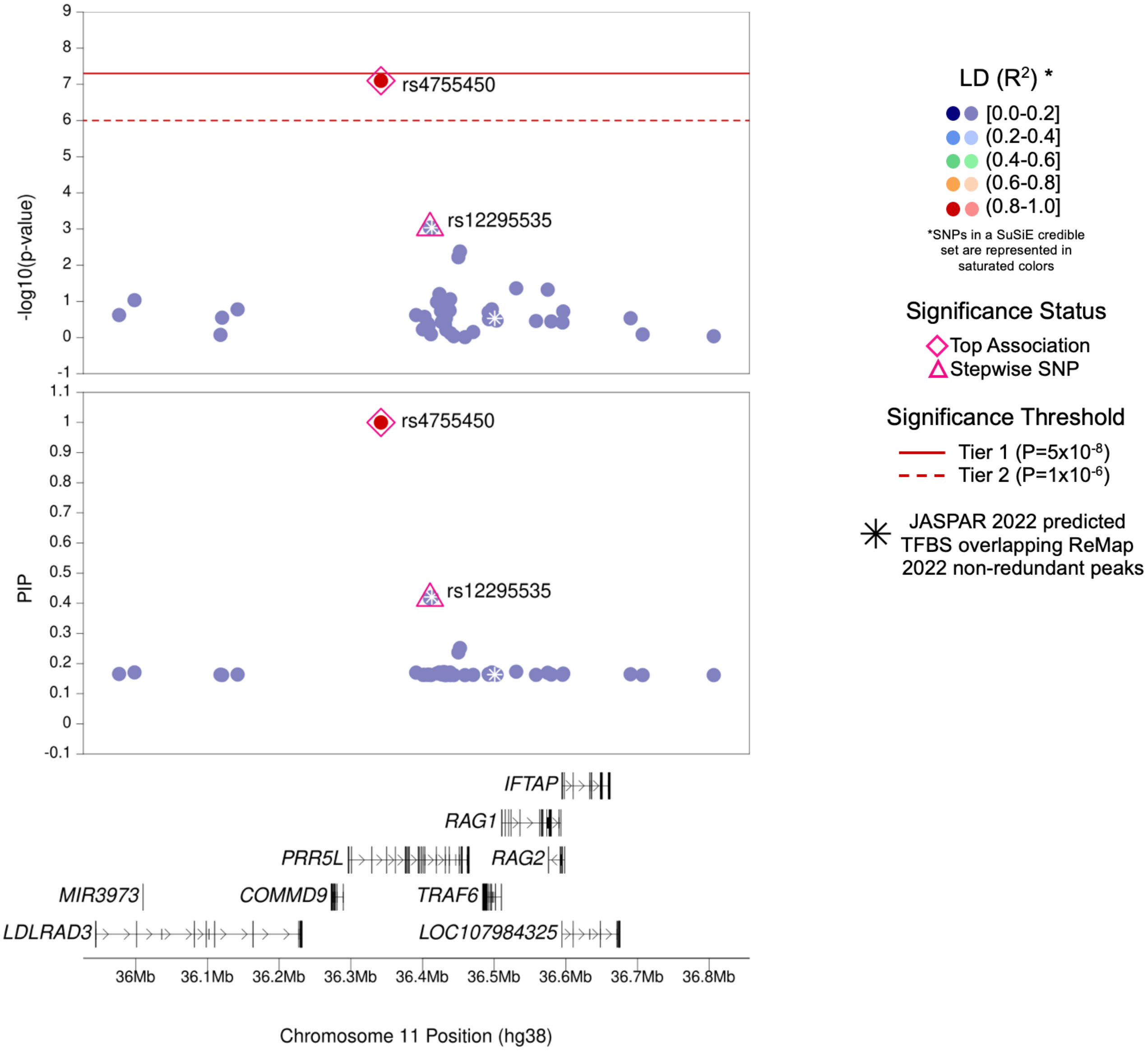

| | |
|---|---|
| **Region Name** | *PRR5L* |
| **Stepwise Region Window (hg38)** | chr11:35842025-36842025 |
| **Regional P-Value (df)** | $P = 2.51x10^{-9}$ (df = 2) |
| **Novel for JIA (Yes/No)** | No |
| **Number of Signals: Stepwise** | 2 |
| **Number of Credible Sets: SuSiE (95% CS)** | 1 |
| **T1/T2 + Stepwise TF Overlap (Yes/No)** | No |
| **Implicated GTEx eGenes* and JIA PBMC eGenes† linked to JIA significant SNPs‡ or their proxies** | NA (No eGenes) |
| **Implicated GTEx eGenes (All Tissues) linked to JIA significant SNPs‡ or their proxies** | *PRR5L* |
| **DGIdb has drug-gene interactions between GTEx eGenes* and JIA PBMC eGenes†** | No |
| **Region Implicated in Autoimmune Disease (Yes/No)** | Yes |

*Limited to EBV-transformed lymphocytes, cultured fibroblasts, whole blood, and spleen tissues

†JIA eGenes for peripheral blood mononuclear cells (PBMCs) from Barnes et al. (PMID: 19565513) were defined as genes with $P_{eQTL} < 1x10^{-4}$

‡JIA significant SNPs defined as SNPs with Best $P_{FDR} < 0.05$

### (t) *TRAFD1–PTPN11* Region (Tier 1)

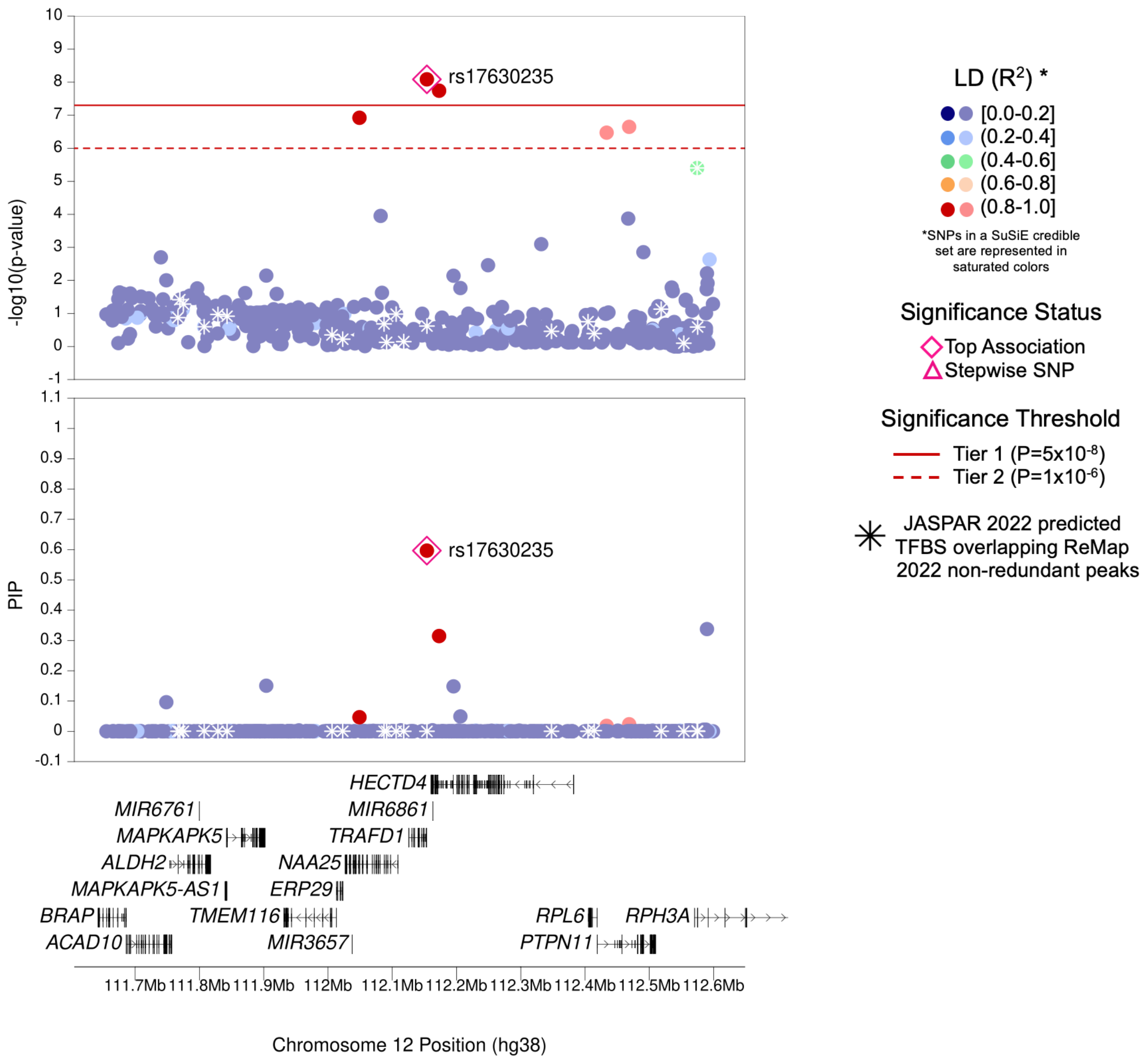

| | |
|---|---|
| **Region Name** | *TRAFD1–PTPN11* |
| **Stepwise Region Window (hg38)** | chr12:111653882-112653882 |
| **Regional P-Value (df)** | P = $8.23 \times 10^{-9}$ (df = 1) |
| **Novel for JIA (Yes/No)** | **Yes** |
| **Number of Signals: Stepwise** | 1 |
| **Number of Credible Sets: SuSiE (95% CS)** | 1 |
| **T1/T2 + Stepwise TF Overlap (Yes/No)** | No |
| **Implicated GTEx eGenes* and JIA PBMC eGenes† linked to JIA significant SNPs‡ or their proxies** | NA (No eGenes) |
| **Implicated GTEx eGenes (All Tissues) linked to JIA significant SNPs‡ or their proxies** | *ADAM1B; ALDH2; BRAP; HECTD4; LINC01405; MAPKAPK5; MAPKAPK5-AS1; NAA25; TMEM116* |
| **DGIdb has drug-gene interactions between GTEx eGenes* and JIA PBMC eGenes†** | No |
| **Region Implicated in Autoimmune Disease (Yes/No)** | Yes |

*Limited to EBV-transformed lymphocytes, cultured fibroblasts, whole blood, and spleen tissues

†JIA eGenes for peripheral blood mononuclear cells (PBMCs) from Barnes et al. (PMID: 19565513) were defined as genes with $P_{eQTL} < 1 \times 10^{-4}$

‡JIA significant SNPs defined as SNPs with Best $P_{FDR} < 0.05$

### (u) *ZFP36L1* Region (Tier 1)

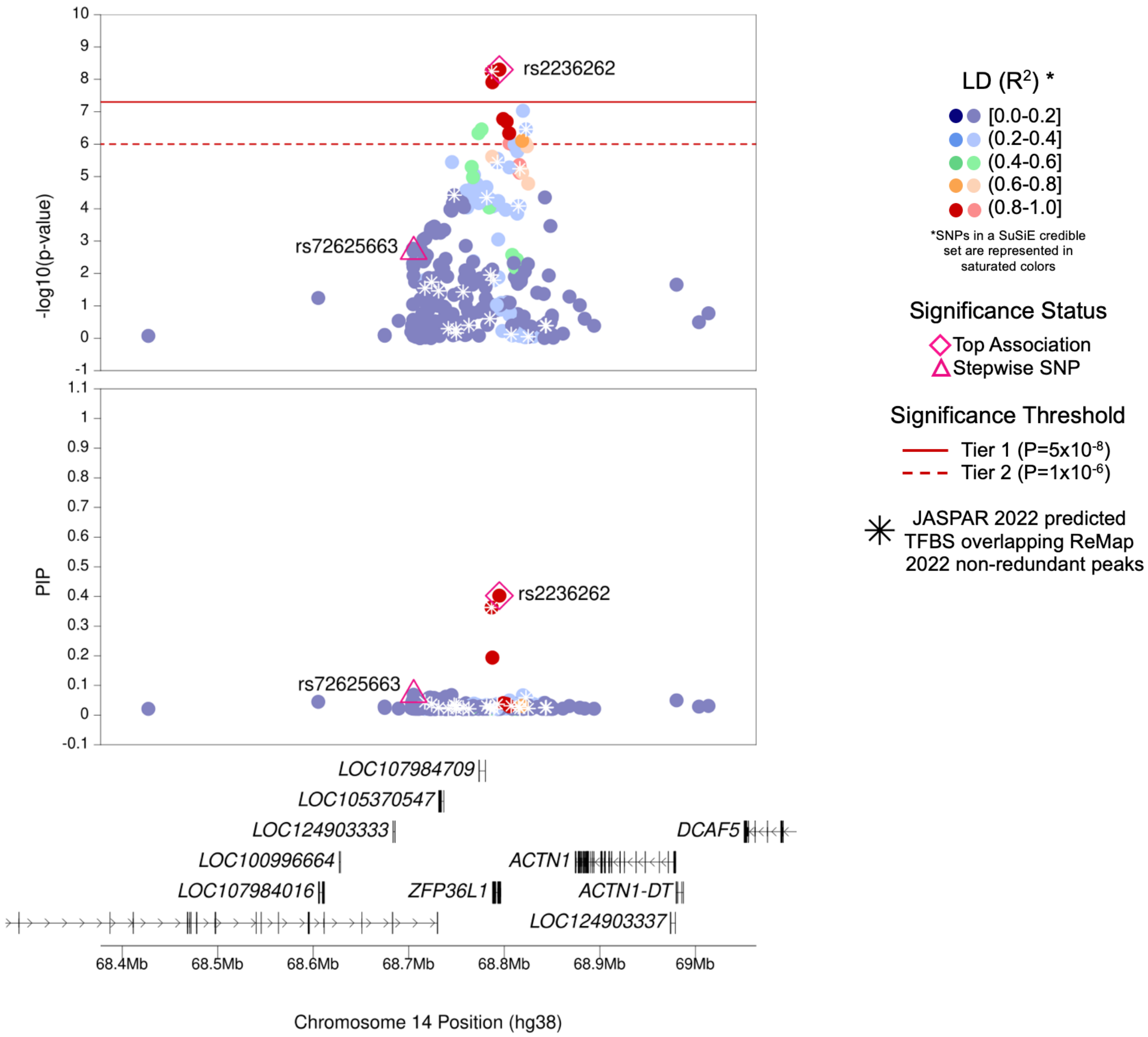

| | |
|---|---|
| **Region Name** | *ZFP36L1* |
| **Stepwise Region Window (hg38)** | chr14:68294755-69294755 |
| **Regional P-Value (df)** | P = $1.49x10^{-10}$ (df = 2) |
| **Novel for JIA (Yes/No)** | No |
| **Number of Signals: Stepwise** | 2 |
| **Number of Credible Sets: SuSiE (95% CS)** | 1 |
| **T1/T2 + Stepwise TF Overlap (Yes/No)** | Yes (1 TFBS with TF EBF1) |
| **Implicated GTEx eGenes* and JIA PBMC eGenes† linked to JIA significant SNPs‡ or their proxies** | *ZFP36L1* |
| **Implicated GTEx eGenes (All Tissues) linked to JIA significant SNPs‡ or their proxies** | *CTD-2325P2.4; ZFP36L1* |
| **DGIdb has drug-gene interactions between GTEx eGenes* and JIA PBMC eGenes†** | No |
| **Region Implicated in Autoimmune Disease (Yes/No)** | Yes |

*Limited to EBV-transformed lymphocytes, cultured fibroblasts, whole blood, and spleen tissues

†JIA eGenes for peripheral blood mononuclear cells (PBMCs) from Barnes et al. (PMID: 19565513) were defined as genes with $P_{eQTL} < 1x10^{-4}$

‡JIA significant SNPs defined as SNPs with Best $P_{FDR} < 0.05$

### (v) *PRM1–RMI2* Region (Tier 1)

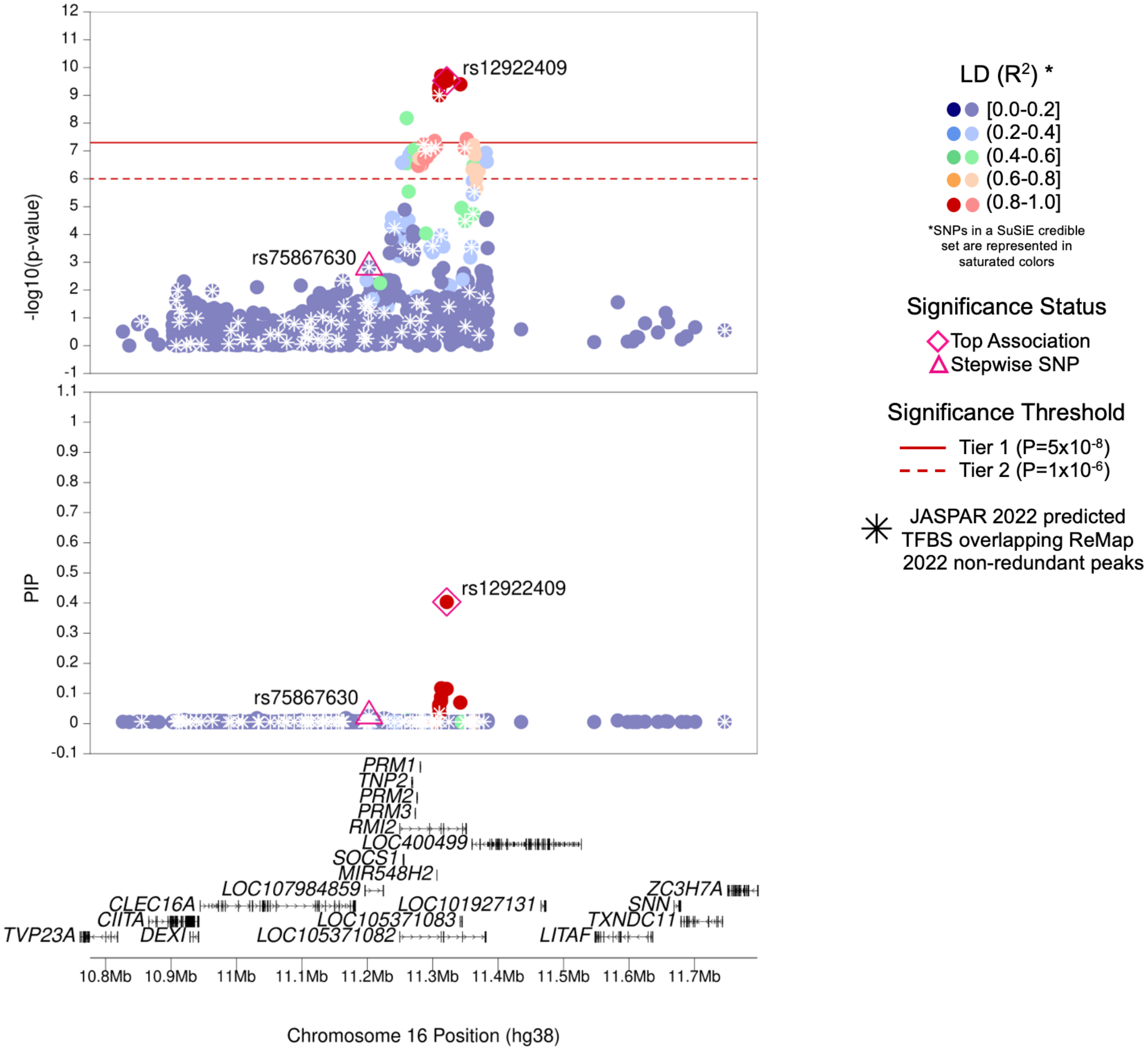

| | |
|---|---|
| **Region Name** | *PRM1–RMI2* |
| **Stepwise Region Window (hg38)** | chr16:10821432-11821432 |
| **Regional P-Value (df)** | $P = 1.93 \times 10^{-12}$ (df = 2) |
| **Novel for JIA (Yes/No)** | No |
| **Number of Signals: Stepwise** | 2 |
| **Number of Credible Sets: SuSiE (95% CS)** | 1 |
| **T1/T2 + Stepwise TF Overlap (Yes/No)** | Yes (11 TFBS with TFs EBF1, EBF3, MYOD1, SIX2, SNAI2, TCF12, TCF3, ZEB1, ZNF148) |
| **Implicated GTEx eGenes* and JIA PBMC eGenes† linked to JIA significant SNPs‡ or their proxies** | *RMI2* |
| **Implicated GTEx eGenes (All Tissues) linked to JIA significant SNPs‡ or their proxies** | *CTD-3088G3.4; DEXI; RMI2; RP11-485G7.5; RP11-485G7.6; SOCS1* |
| **DGIdb has drug-gene interactions between GTEx eGenes* and JIA PBMC eGenes†** | No |
| **Region Implicated in Autoimmune Disease (Yes/No)** | Yes |

*Limited to EBV-transformed lymphocytes, cultured fibroblasts, whole blood, and spleen tissues

†JIA eGenes for peripheral blood mononuclear cells (PBMCs) from Barnes et al. (PMID: 19565513) were defined as genes with $P_{eQTL} < 1 \times 10^{-4}$

‡JIA significant SNPs defined as SNPs with Best $P_{FDR} < 0.05$

### (w) *SMPD3–ZFP90–CDH3* Region (Tier 1)

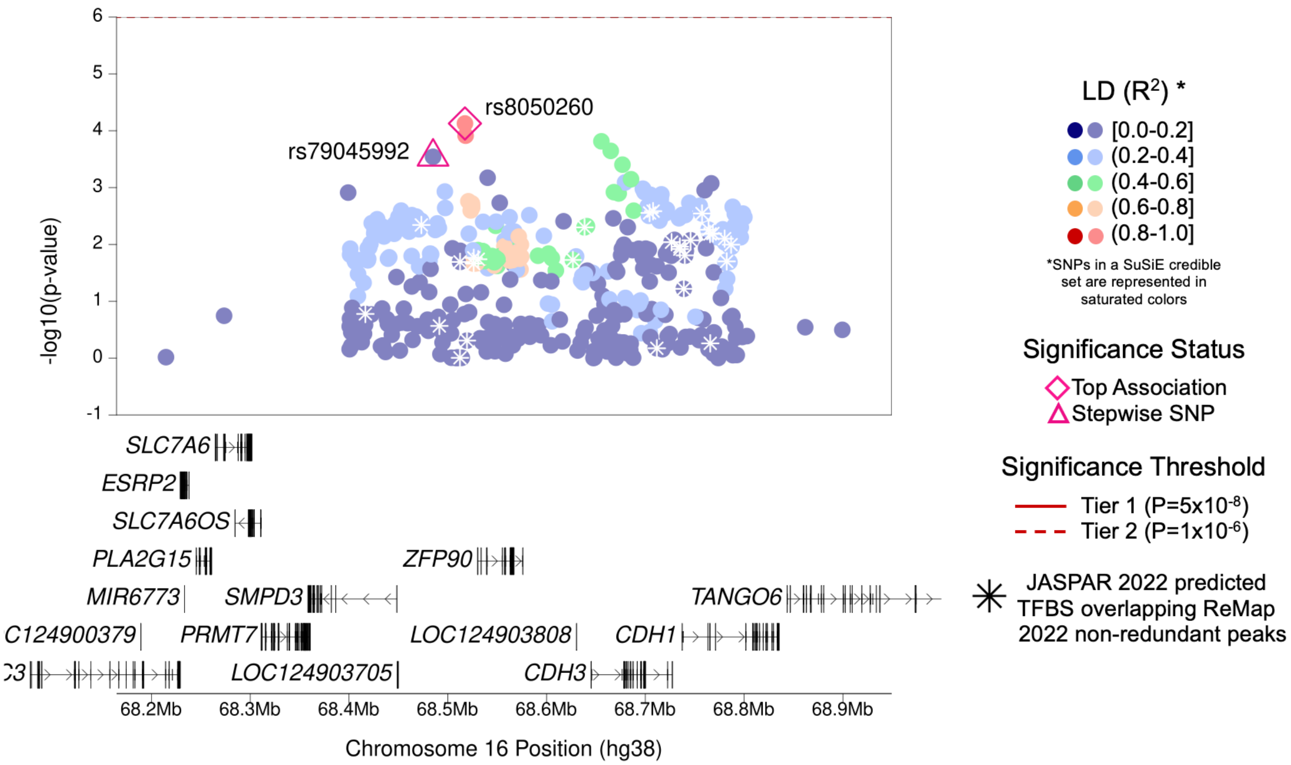

| | |
|---|---|
| **Region Name** | *SMPD3–ZFP90–CDH3* |
| **Stepwise Region Window (hg38)** | chr16:68017374-69017374 |
| **Regional P-Value (df)** | $P = 3.09x10^{-8}$ (df = 2) |
| **Novel for JIA (Yes/No)** | **Yes** |
| **Number of Signals: Stepwise** | 2 |
| **Number of Credible Sets: SuSiE (95% CS)** | 0 (Large number of non-additive SNPs in region; SuSiE not computed) |
| **T1/T2 + Stepwise TF Overlap (Yes/No)** | No |
| **Implicated GTEx eGenes* and JIA PBMC eGenes† linked to JIA significant SNPs‡ or their proxies** | *CDH1; DUS2; PRMT7; RP11-615I2.6; SLC7A6; SMDP3; SNTB2; ZFP90* |
| **Implicated GTEx eGenes (All Tissues) linked to JIA significant SNPs‡ or their proxies** | *CDH1; CDH3; CTD-2012K14.6; DDX28; DPEP2; DUS2; FTLP14; NFATC3; PLA2G15; PRMT7; RP11-615I2.1; RP11-615I2.6; RP11-96D1.8; SLC7A6; SMDP3; SNTB2; UTP4; ZFP90* |
| **DGIdb has drug-gene interactions between GTEx eGenes* and JIA PBMC eGenes†** | Yes |
| **Region Implicated in Autoimmune Disease (Yes/No)** | Yes |

*Limited to EBV-transformed lymphocytes, cultured fibroblasts, whole blood, and spleen tissues

†JIA eGenes for peripheral blood mononuclear cells (PBMCs) from Barnes et al. (PMID: 19565513) were defined as genes with $P_{eQTL} < 1x10^{-4}$

‡JIA significant SNPs defined as SNPs with Best $P_{FDR} < 0.05$

### (x) *NOS2–LYRM9* Region (Tier 1)

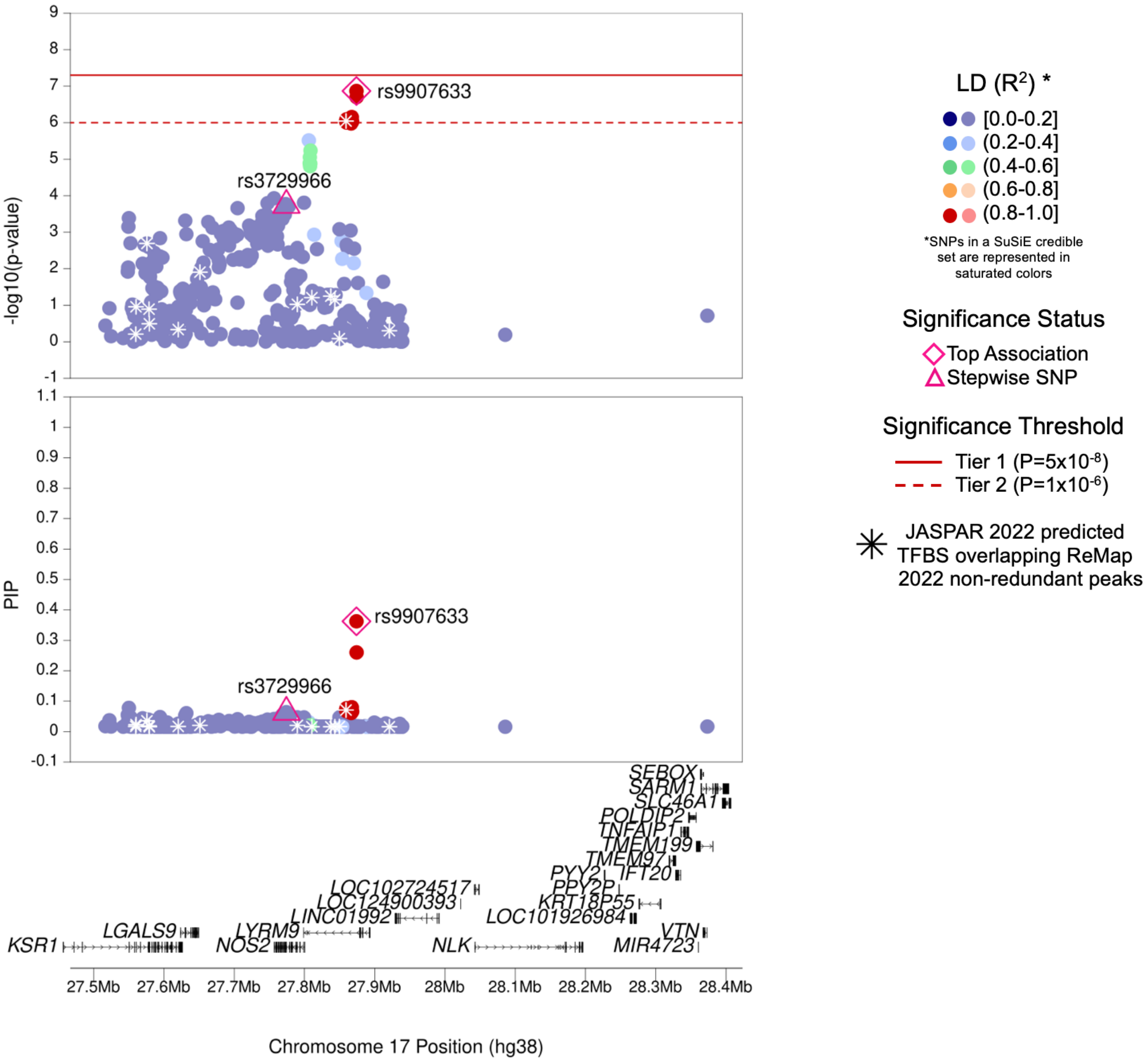

| | |
|---|---|
| **Region Name** | *NOS2–LYRM9* |
| **Stepwise Region Window (hg38)** | chr17:27373987-28373987 |
| **Regional P-Value (df)** | $P = 1.27x10^{-9}$ (df = 2) |
| **Novel for JIA (Yes/No)** | **Yes** |
| **Number of Signals: Stepwise** | 2 |
| **Number of Credible Sets: SuSiE (95% CS)** | 1 |
| **T1/T2 + Stepwise TF Overlap (Yes/No)** | Yes (2 TFBS for TF MITF) |
| **Implicated GTEx eGenes* and JIA PBMC eGenes† linked to JIA significant SNPs‡ or their proxies** | *LGALS9* |
| **Implicated GTEx eGenes (All Tissues) linked to JIA significant SNPs‡ or their proxies** | *LGALS9; LYRM9; NLK; NOS2; NOS2P1; RP11-138P22.1* |
| **DGIdb has drug-gene interactions between GTEx eGenes* and JIA PBMC eGenes†** | No |
| **Region Implicated in Autoimmune Disease (Yes/No)** | Yes |

*Limited to EBV-transformed lymphocytes, cultured fibroblasts, whole blood, and spleen tissues

†JIA eGenes for peripheral blood mononuclear cells (PBMCs) from Barnes et al. (PMID: 19565513) were defined as genes with $P_{eQTL} < 1x10^{-4}$

‡JIA significant SNPs defined as SNPs with Best $P_{FDR} < 0.05$

### (y) *PTPN2* Region (Tier 1)

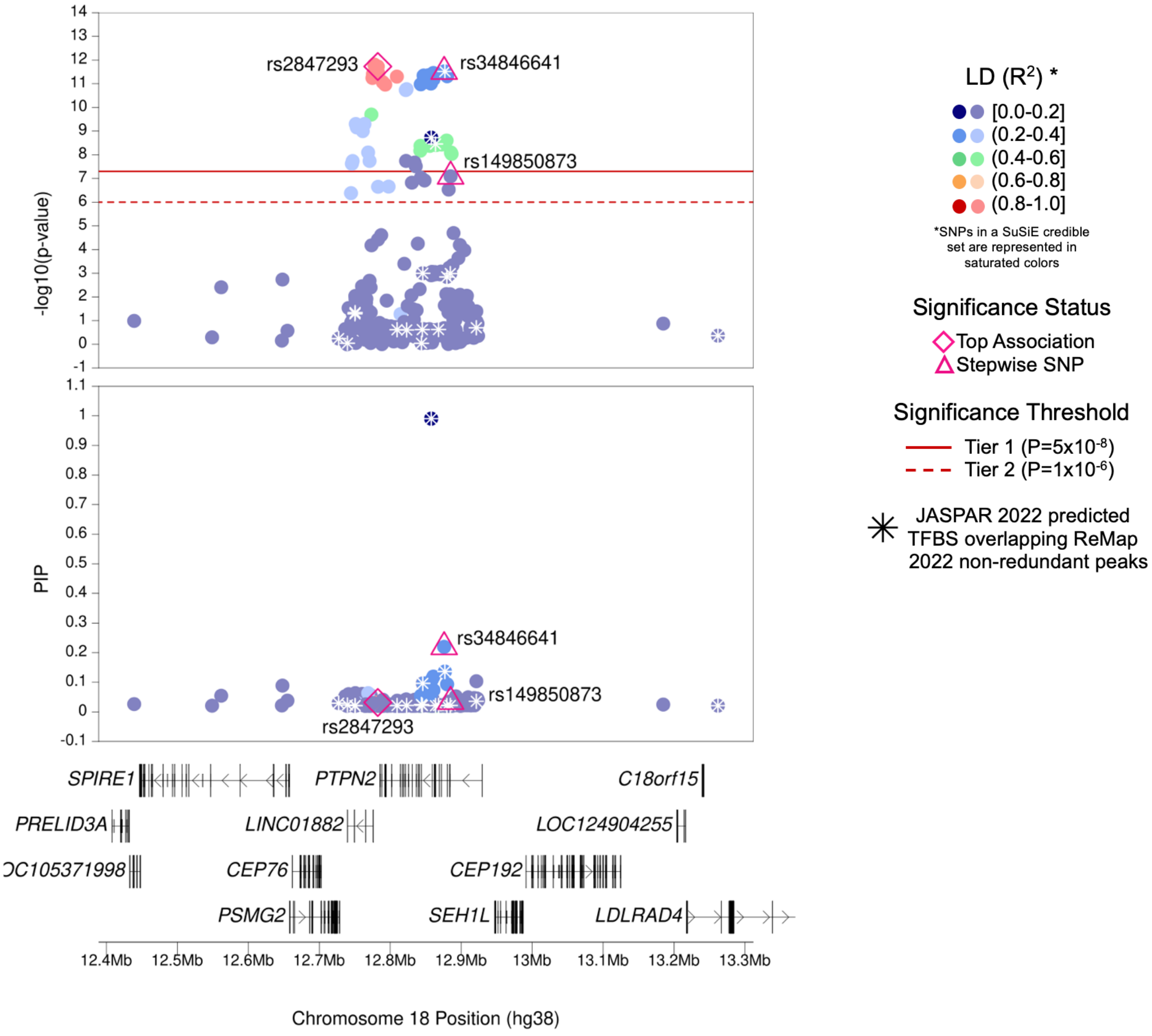

| | |
|---|---|
| **Region Name** | *PTPN2* |
| **Stepwise Region Window (hg38)** | chr18:12282449-13282449 |
| **Regional P-Value (df)** | P = $2.58 \times 10^{-20}$ (df = 3) |
| **Novel for JIA (Yes/No)** | No |
| **Number of Signals: Stepwise** | 3 |
| **Number of Credible Sets: SuSiE (95% CS)** | 2 |
| **T1/T2 + Stepwise TF Overlap (Yes/No)** | Yes (3 TFBS with GATA1, PHOX2B, SPIB) |
| **Implicated GTEx eGenes* and JIA PBMC eGenes† linked to JIA significant SNPs‡ or their proxies** | *CEP192; LINC01882; PTPN2; RP11-973H7.1* |
| **Implicated GTEx eGenes (All Tissues) linked to JIA significant SNPs‡ or their proxies** | *CEP192; CEP76; LINC01882; PTPN2; RP11-861E21.2; RP11-973H7.1* |
| **DGIdb has drug-gene interactions between GTEx eGenes* and JIA PBMC eGenes†** | No |
| **Region Implicated in Autoimmune Disease (Yes/No)** | Yes |

*Limited to EBV-transformed lymphocytes, cultured fibroblasts, whole blood, and spleen tissues

†JIA eGenes for peripheral blood mononuclear cells (PBMCs) from Barnes et al. (PMID: 19565513) were defined as genes with $P_{eQTL} < 1 \times 10^{-4}$

‡JIA significant SNPs defined as SNPs with Best $P_{FDR} < 0.05$

### (z) *TYK2* Region (Tier 1)

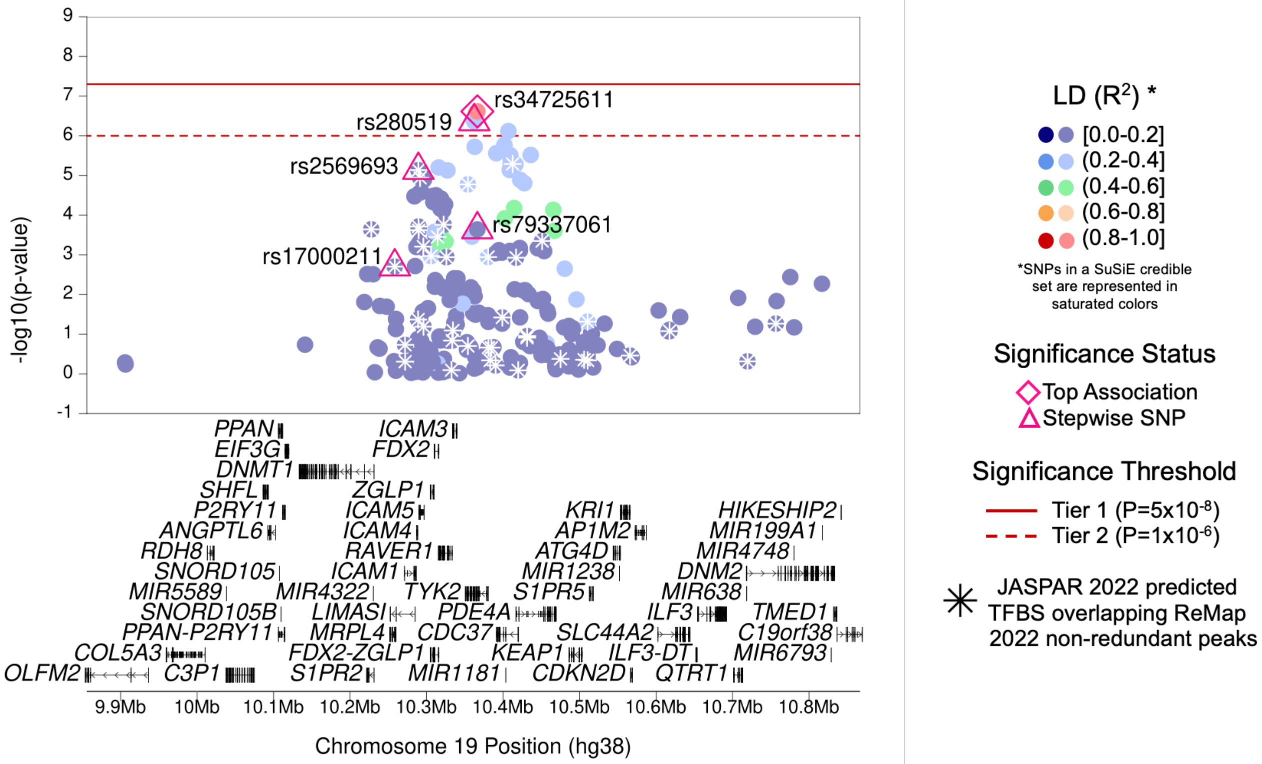

| | |
|---|---|
| **Region Name** | *TYK2* |
| **Stepwise Region Window (hg38)** | chr19:9866391-10866391 |
| **Regional P-Value (df)** | P = $3.87x10^{-15}$ (df = 5) |
| **Novel for JIA (Yes/No)** | No |
| **Number of Signals: Stepwise** | 5 |
| **Number of Credible Sets: SuSiE (95% CS)** | 0 (Large number of non-additive SNPs in region; SuSiE not computed) |
| **T1/T2 + Stepwise TF Overlap (Yes/No)** | Yes (2 TFBS for TFs KLF4 and KLF9) |
| **Implicated GTEx eGenes* and JIA PBMC eGenes† linked to JIA significant SNPs‡ or their proxies** | *AC114271.2; AP1M2; C19orf66; COL5A3; EIF3G; ICAM4; KEAP1; P2RY11; S1PR2; TYK2* |
| **Implicated GTEx eGenes (All Tissues) linked to JIA significant SNPs‡ or their proxies** | *AC114271.2; AP1M2; C19orf66; CDC37; COL5A3; CTC-510F12.2; CTD-2369P2.5; CTD-2369P2.8; EIF3G; FDX2; ICAM5; KEAP1; MRPL4; P2RY11; PDE4A; S1PR2; S1PR5; TYK2; ZGLP1; ZNF266; ZNF561* |
| **DGIdb has drug-gene interactions between GTEx eGenes* and JIA PBMC eGenes†** | Yes |
| **Region Implicated in Autoimmune Disease (Yes/No)** | Yes |

*Limited to EBV-transformed lymphocytes, cultured fibroblasts, whole blood, and spleen tissues

†JIA eGenes for peripheral blood mononuclear cells (PBMCs) from Barnes et al. (PMID: 19565513) were defined as genes with $P_{eQTL} < 1x10^{-4}$

‡JIA significant SNPs defined as SNPs with Best $P_{FDR} < 0.05$

### (aa) *LOC100506403* Region (Tier 1)

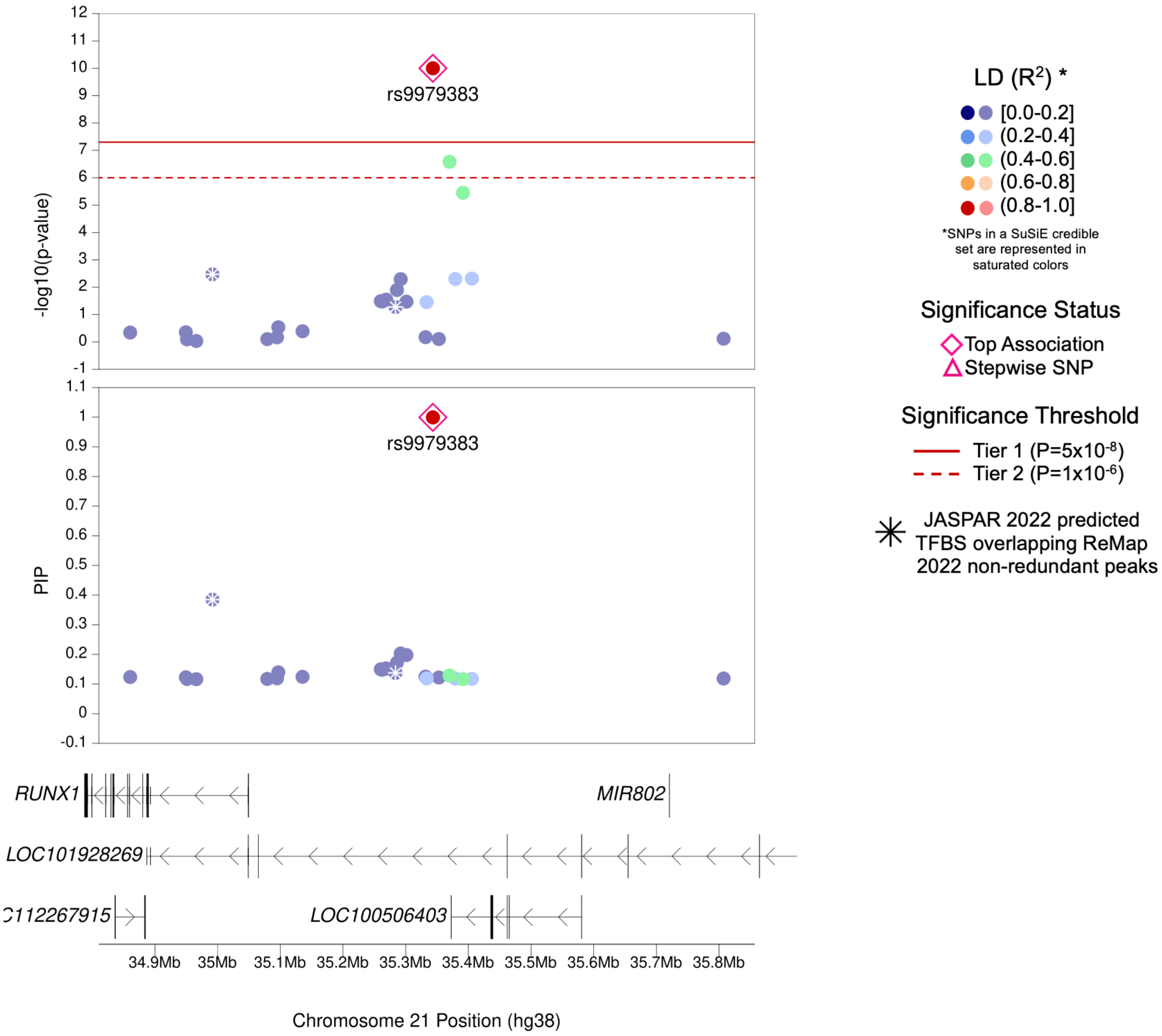

| | |
|---|---|
| **Region Name** | *LOC100506403* |
| **Stepwise Region Window (hg38)** | chr21:34843463-35843463 |
| **Regional P-Value (df)** | P = $1.10\times10^{-10}$ (df = 1) |
| **Novel for JIA (Yes/No)** | No |
| **Number of Signals: Stepwise** | 1 |
| **Number of Credible Sets: SuSiE (95% CS)** | 1 |
| **T1/T2 + Stepwise TF Overlap (Yes/No)** | No |
| **Implicated GTEx eGenes* and JIA PBMC eGenes† linked to JIA significant SNPs‡ or their proxies** | NA (No eGenes) |
| **Implicated GTEx eGenes (All Tissues) linked to JIA significant SNPs‡ or their proxies** | NA (No eGenes) |
| **DGIdb has drug-gene interactions between GTEx eGenes* and JIA PBMC eGenes†** | No |
| **Region Implicated in Autoimmune Disease (Yes/No)** | Yes |

*Limited to EBV-transformed lymphocytes, cultured fibroblasts, whole blood, and spleen tissues

†JIA eGenes for peripheral blood mononuclear cells (PBMCs) from Barnes et al. (PMID: 19565513) were defined as genes with $P_{eQTL} < 1\times10^{-4}$

‡JIA significant SNPs defined as SNPs with Best $P_{FDR} < 0.05$

### (ab) *UBE2L3* Region (Tier 1)

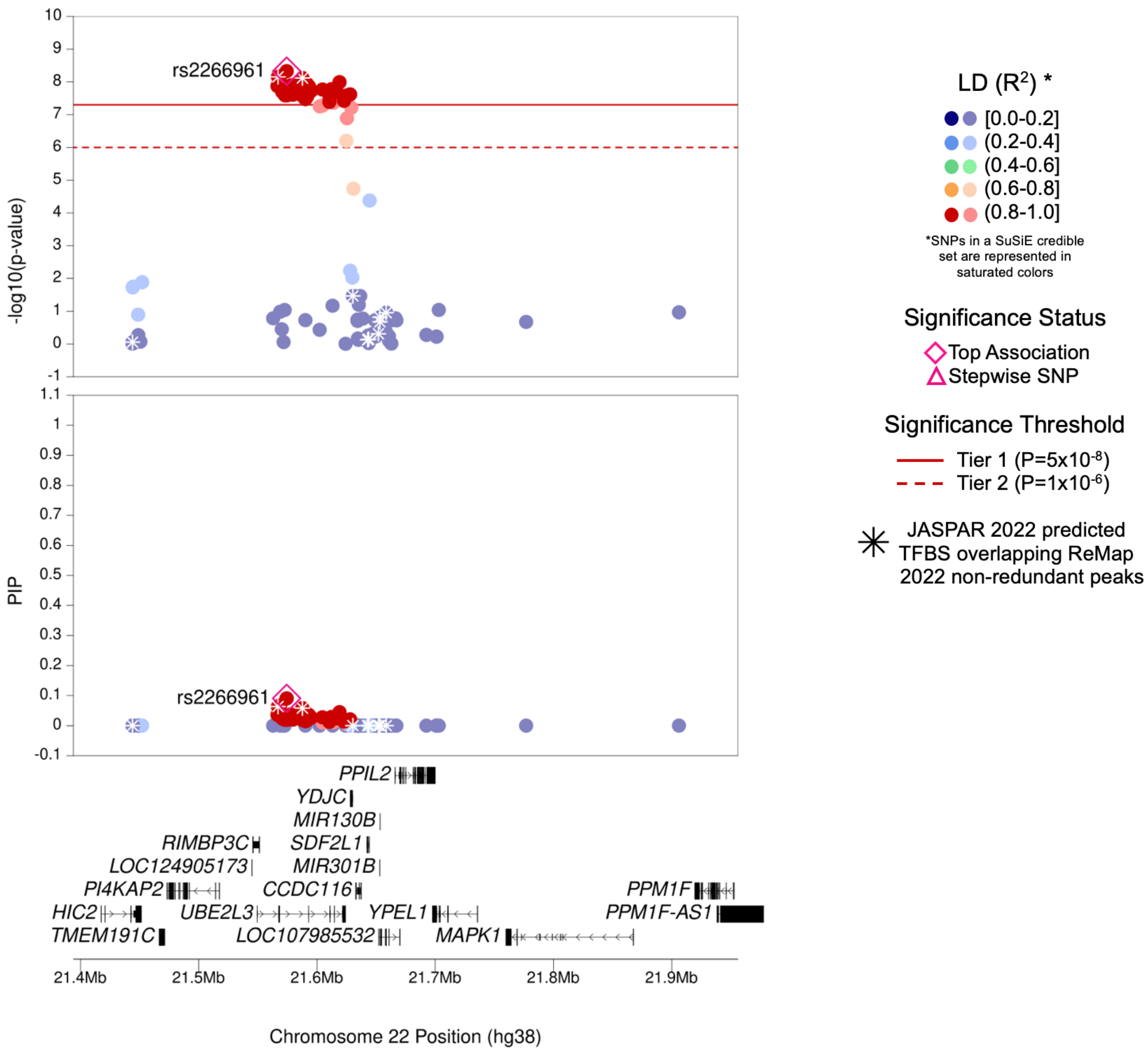

| | |
|---|---|
| **Region Name** | *UBE2L3* |
| **Stepwise Region Window (hg38)** | chr22:21074308-22074308 |
| **Regional P-Value (df)** | P = $5.30 \times 10^{-9}$ (df = 1) |
| **Novel for JIA (Yes/No)** | No |
| **Number of Signals: Stepwise** | 1 |
| **Number of Credible Sets: SuSiE (95% CS)** | 1 |
| **T1/T2 + Stepwise TF Overlap (Yes/No)** | Yes (3 TFBS with TFs TCF3, TCF4, and ZNF652) |
| **Implicated GTEx eGenes* and JIA PBMC eGenes† linked to JIA significant SNPs‡ or their proxies** | *CCDC116; UBE2L3* |
| **Implicated GTEx eGenes (All Tissues) linked to JIA significant SNPs‡ or their proxies** | *BCRP6; CCDC116; HIC2; KB-1183D5.13; KB-1440D3.14; PPIL2; TOP3BP1; UBE2L3; YDJC* |
| **DGIdb has drug-gene interactions between GTEx eGenes* and JIA PBMC eGenes†** | No |
| **Region Implicated in Autoimmune Disease (Yes/No)** | Yes |

*Limited to EBV-transformed lymphocytes, cultured fibroblasts, whole blood, and spleen tissues

†JIA eGenes for peripheral blood mononuclear cells (PBMCs) from Barnes et al. (PMID: 19565513) were defined as genes with $P_{eQTL} < 1 \times 10^{-4}$

‡JIA significant SNPs defined as SNPs with Best $P_{FDR} < 0.05$

### (ac) *IL2RB* Region (Tier 1)

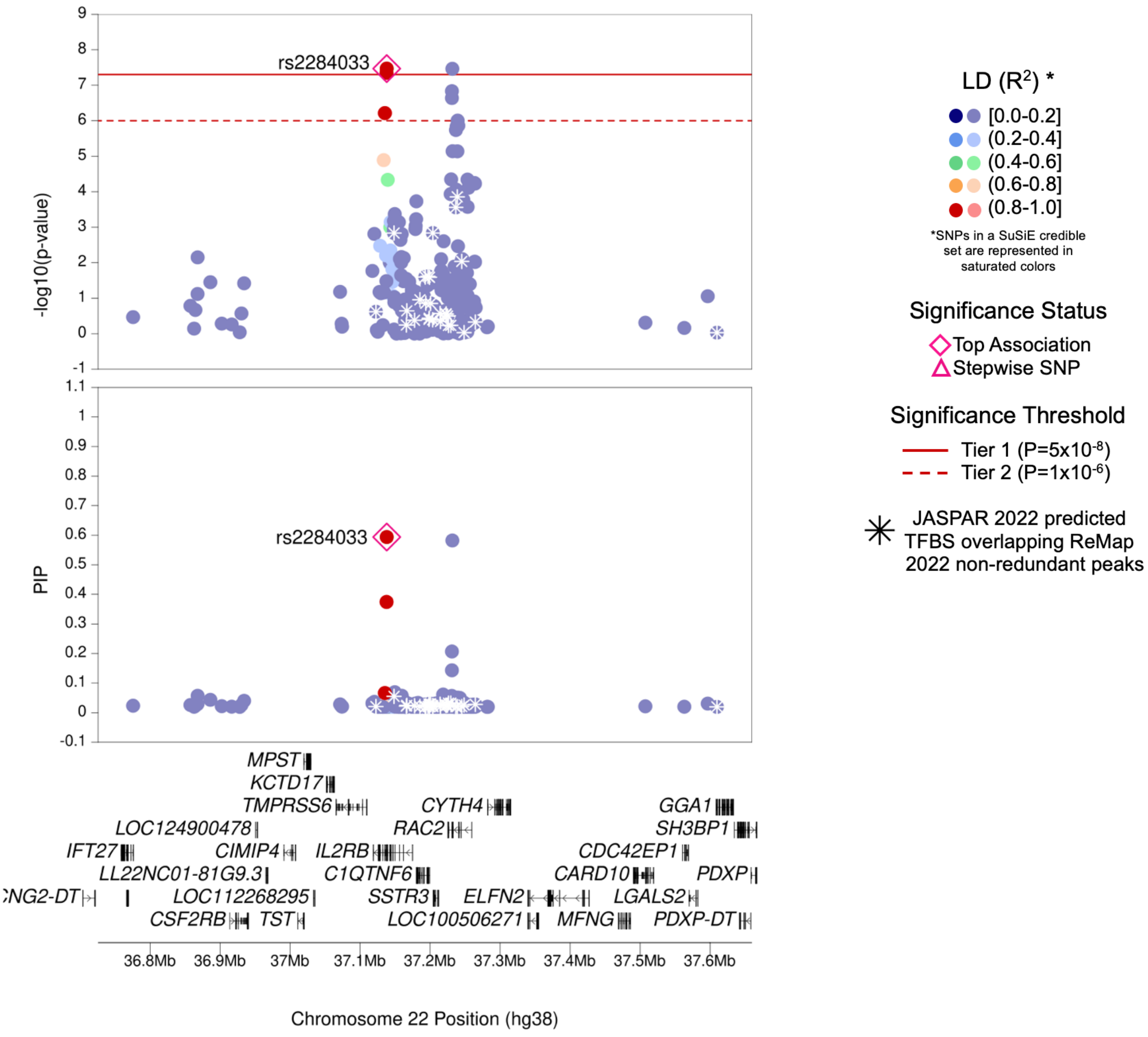

| | |
|---|---|
| **Region Name** | *IL2RB* |
| **Stepwise Region Window (hg38)** | chr22:37128579-37146722 |
| **Regional P-Value (df)** | P = $3.40 \times 10^{-8}$ (df = 1) |
| **Novel for JIA (Yes/No)** | No |
| **Number of Signals: Stepwise** | 1 |
| **Number of Credible Sets: SuSiE (95% CS)** | 1 |
| **T1/T2 + Stepwise TF Overlap (Yes/No)** | No |
| **Implicated GTEx eGenes* and JIA PBMC eGenes† linked to JIA significant SNPs‡ or their proxies** | *C1QTNF6* |
| **Implicated GTEx eGenes (All Tissues) linked to JIA significant SNPs‡ or their proxies** | *C1QTNF6; ELFN2; IL2RB; RAC2; RP1-151B14.9; SSTR3; TMPRSS6* |
| **DGIdb has drug-gene interactions between GTEx eGenes* and JIA PBMC eGenes†** | No |
| **Region Implicated in Autoimmune Disease (Yes/No)** | Yes |

*Limited to EBV-transformed lymphocytes, cultured fibroblasts, whole blood, and spleen tissues

†JIA eGenes for peripheral blood mononuclear cells (PBMCs) from Barnes et al. (PMID: 19565513) were defined as genes with $P_{eQTL} < 1 \times 10^{-4}$

‡JIA significant SNPs defined as SNPs with Best $P_{FDR} < 0.05$

### (ad) *RAC2* Region (Tier 1)

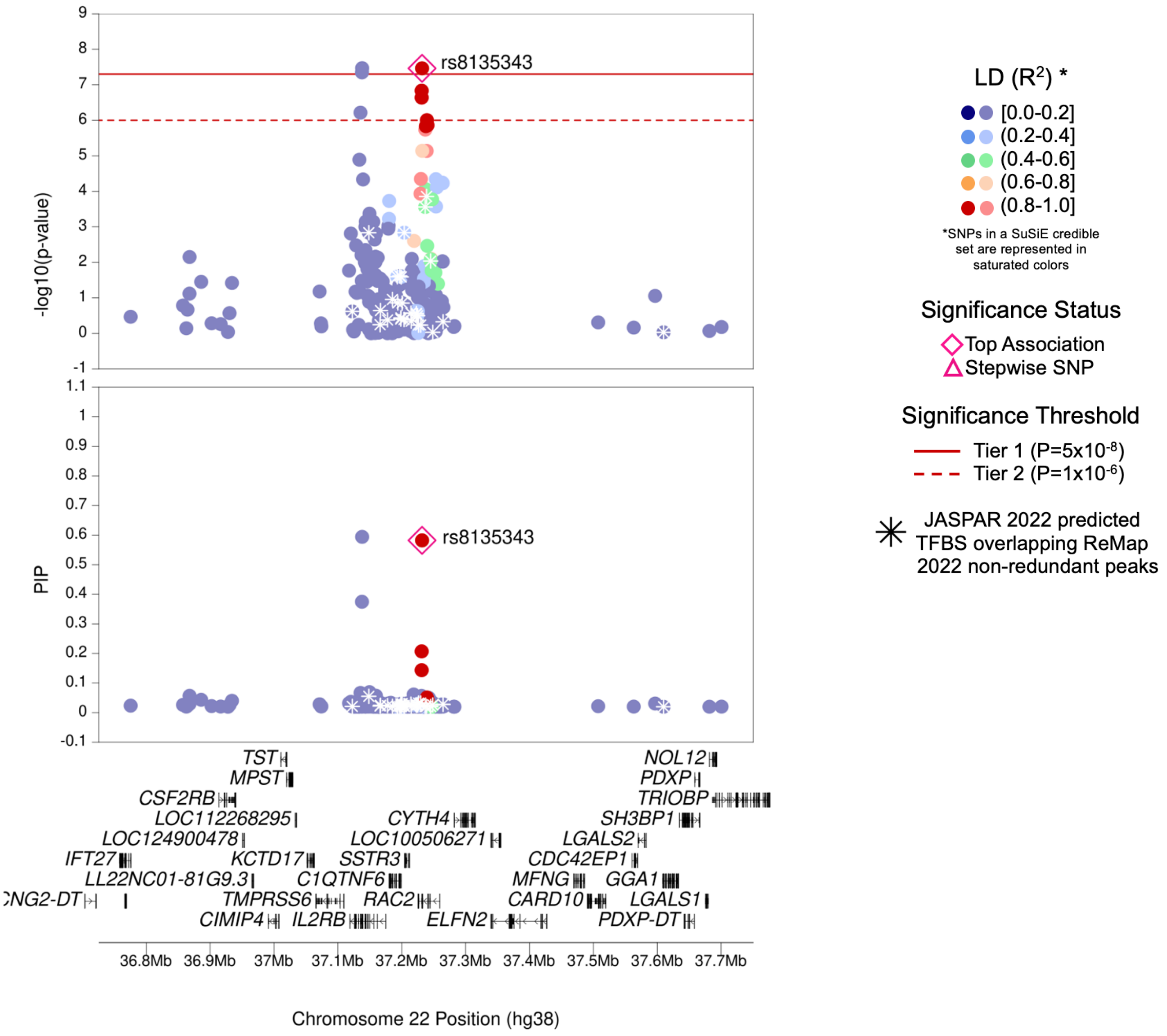

| | |
|---|---|
| **Region Name** | *RAC2* |
| **Stepwise Region Window (hg38)** | chr22:37180019-37264137 |
| **Regional P-Value (df)** | P = $3.44 \times 10^{-8}$ (df = 1) |
| **Novel for JIA (Yes/No)** | **Yes** |
| **Number of Signals: Stepwise** | 1 |
| **Number of Credible Sets: SuSiE (95% CS)** | 1 |
| **T1/T2 + Stepwise TF Overlap (Yes/No)** | No |
| **Implicated GTEx eGenes* and JIA PBMC eGenes† linked to JIA significant SNPs‡ or their proxies** | *C1QTNF6* |
| **Implicated GTEx eGenes (All Tissues) linked to JIA significant SNPs‡ or their proxies** | *C1QTNF6; ELFN2; IL2RB; RAC2; RP1-151B14.9; SSTR3; TMPRSS6* |
| **DGIdb has drug-gene interactions between GTEx eGenes* and JIA PBMC eGenes†** | No |
| **Region Implicated in Autoimmune Disease (Yes/No)** | Yes |

*Limited to EBV-transformed lymphocytes, cultured fibroblasts, whole blood, and spleen tissues

†JIA eGenes for peripheral blood mononuclear cells (PBMCs) from Barnes et al. (PMID: 19565513) were defined as genes with $P_{eQTL} < 1 \times 10^{-4}$

‡JIA significant SNPs defined as SNPs with Best $P_{FDR} < 0.05$

### (ae) *RUNX3* Region (Tier 2)

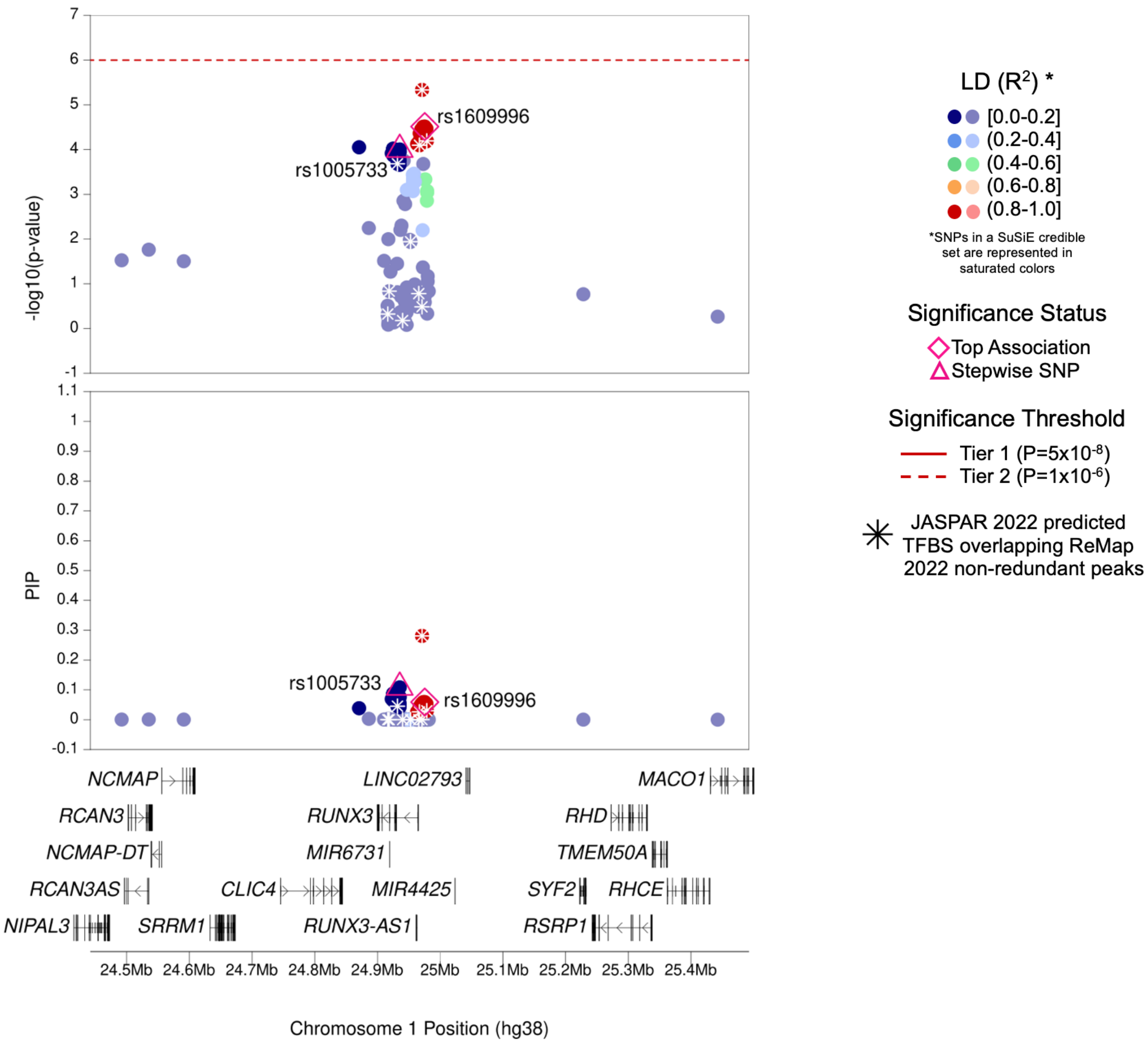

| | |
|---|---|
| **Region Name** | *RUNX3* |
| **Stepwise Region Window (hg38)** | chr1:24475378-25475378 |
| **Regional P-Value (df)** | P = $5.85 \times 10^{-8}$ (df = 2) |
| **Novel for JIA (Yes/No)** | No |
| **Number of Signals: Stepwise** | 2 |
| **Number of Credible Sets: SuSiE (95% CS)** | 2 |
| **T1/T2 + Stepwise TF Overlap (Yes/No)** | No |
| **Implicated GTEx eGenes* and JIA PBMC eGenes† linked to JIA significant SNPs‡ or their proxies** | NA (No eGenes) |
| **Implicated GTEx eGenes (All Tissues) linked to JIA significant SNPs‡ or their proxies** | *RHD; RUNX3; SELENON* |
| **DGIdb has drug-gene interactions between GTEx eGenes* and JIA PBMC eGenes†** | No |
| **Region Implicated in Autoimmune Disease (Yes/No)** | Yes |

*Limited to EBV-transformed lymphocytes, cultured fibroblasts, whole blood, and spleen tissues

†JIA eGenes for peripheral blood mononuclear cells (PBMCs) from Barnes et al. (PMID: 19565513) were defined as genes with $P_{eQTL} < 1 \times 10^{-4}$

‡JIA significant SNPs defined as SNPs with Best $P_{FDR} < 0.05$

### (af) *CD247* Region (Tier 2)

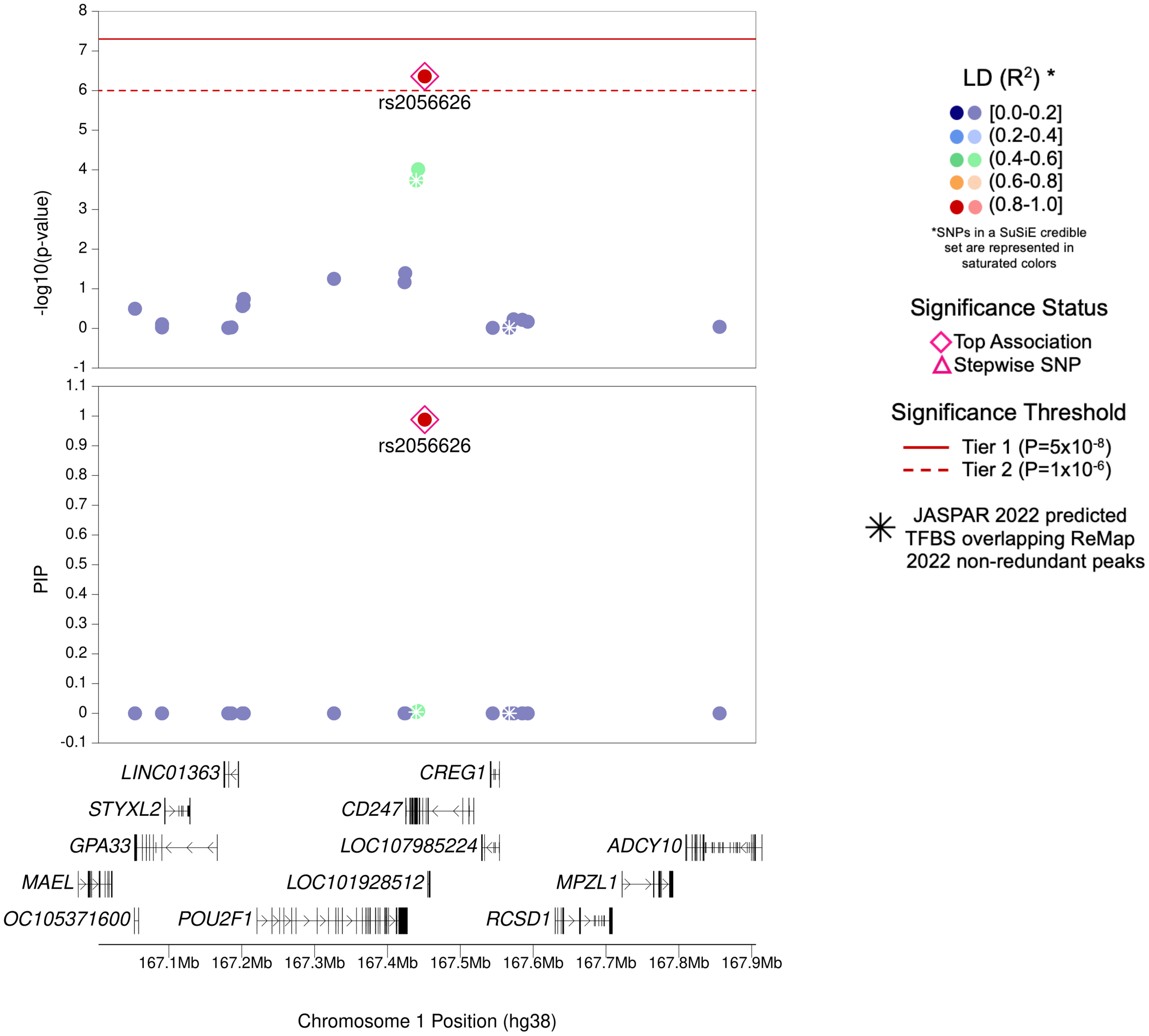

| | |
|---|---|
| **Region Name** | *CD247* |
| **Stepwise Region Window (hg38)** | chr1:166951188-167951188 |
| **Regional P-Value (df)** | $P = 4.39x10^{-7}$ (df = 1) |
| **Novel for JIA (Yes/No)** | No |
| **Number of Signals: Stepwise** | 1 |
| **Number of Credible Sets: SuSiE (95% CS)** | 1 |
| **T1/T2 + Stepwise TF Overlap (Yes/No)** | No |
| **Implicated GTEx eGenes* and JIA PBMC eGenes† linked to JIA significant SNPs‡ or their proxies** | *CD247; RP11-104L21.2* |
| **Implicated GTEx eGenes (All Tissues) linked to JIA significant SNPs‡ or their proxies** | *CD247; MPC2; RP11-104L21.2; RP11-104L21.3* |
| **DGIdb has drug-gene interactions between GTEx eGenes* and JIA PBMC eGenes†** | Yes |
| **Region Implicated in Autoimmune Disease (Yes/No)** | Yes |

*Limited to EBV-transformed lymphocytes, cultured fibroblasts, whole blood, and spleen tissues

†JIA eGenes for peripheral blood mononuclear cells (PBMCs) from Barnes et al. (PMID: 19565513) were defined as genes with $P_{eQTL} < 1x10^{-4}$

‡JIA significant SNPs defined as SNPs with Best $P_{FDR} < 0.05$

### (ag) *EIF2–DYRK3* Region (Tier 2)

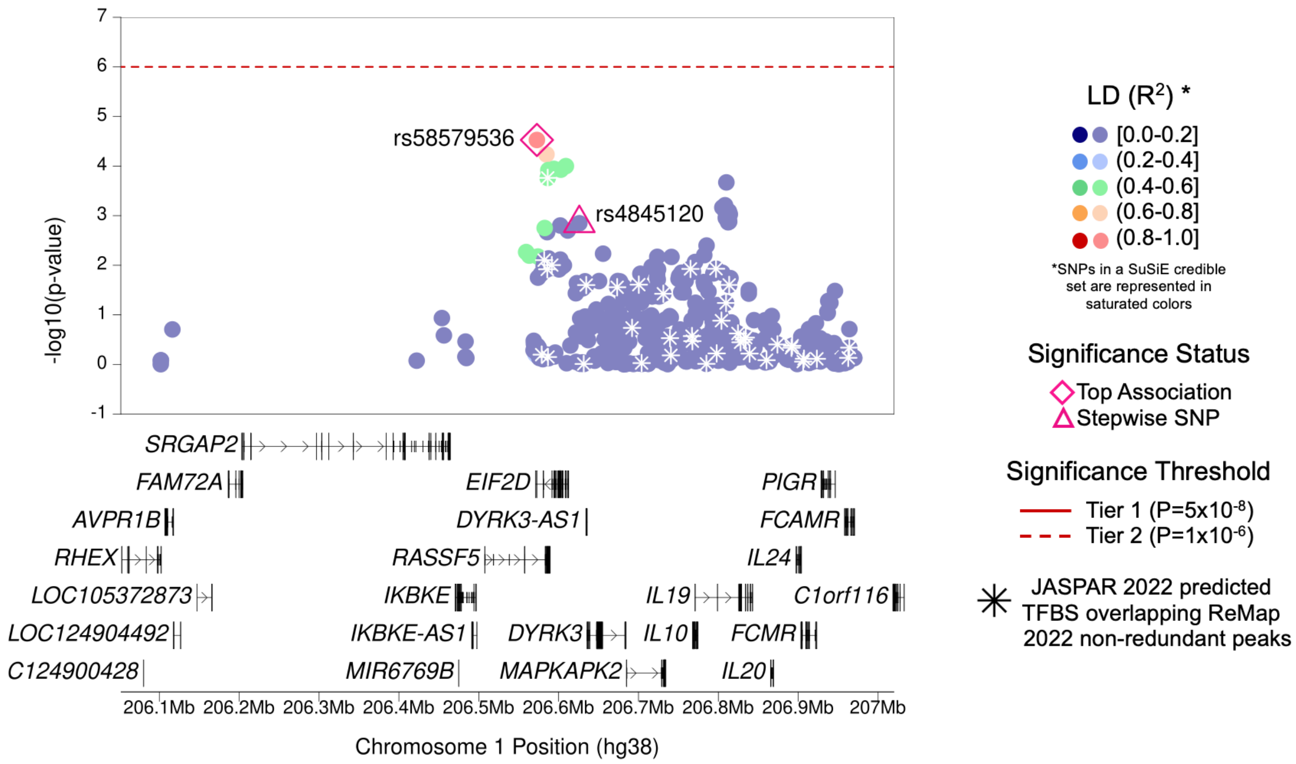

| | |
|---|---|
| **Region Name** | *EIF2–DYRK3* |
| **Stepwise Region Window (hg38)** | chr1:206072797-207072797 |
| **Regional P-Value (df)** | P = $2.23\times10^{-7}$ (df = 2) |
| **Novel for JIA (Yes/No)** | **Yes** |
| **Number of Signals: Stepwise** | 2 |
| **Number of Credible Sets: SuSiE (95% CS)** | 0 (Large number of non-additive SNPs in region; SuSiE not computed) |
| **T1/T2 + Stepwise TF Overlap (Yes/No)** | No |
| **Implicated GTEx eGenes* and JIA PBMC eGenes† linked to JIA significant SNPs‡ or their proxies** | *RP11-534L20.4* |
| **Implicated GTEx eGenes (All Tissues) linked to JIA significant SNPs‡ or their proxies** | *DYRK3; EIF2D; RASSF5; RP11-534L20.4; SRGAP2* |
| **DGIdb has drug-gene interactions between GTEx eGenes* and JIA PBMC eGenes†** | No |
| **Region Implicated in Autoimmune Disease (Yes/No)** | Yes |

*Limited to EBV-transformed lymphocytes, cultured fibroblasts, whole blood, and spleen tissues

†JIA eGenes for peripheral blood mononuclear cells (PBMCs) from Barnes et al. (PMID: 19565513) were defined as genes with $P_{eQTL} < 1\times10^{-4}$

‡JIA significant SNPs defined as SNPs with Best $P_{FDR} < 0.05$

### (ah) *CNNM4* Region (Tier 2)

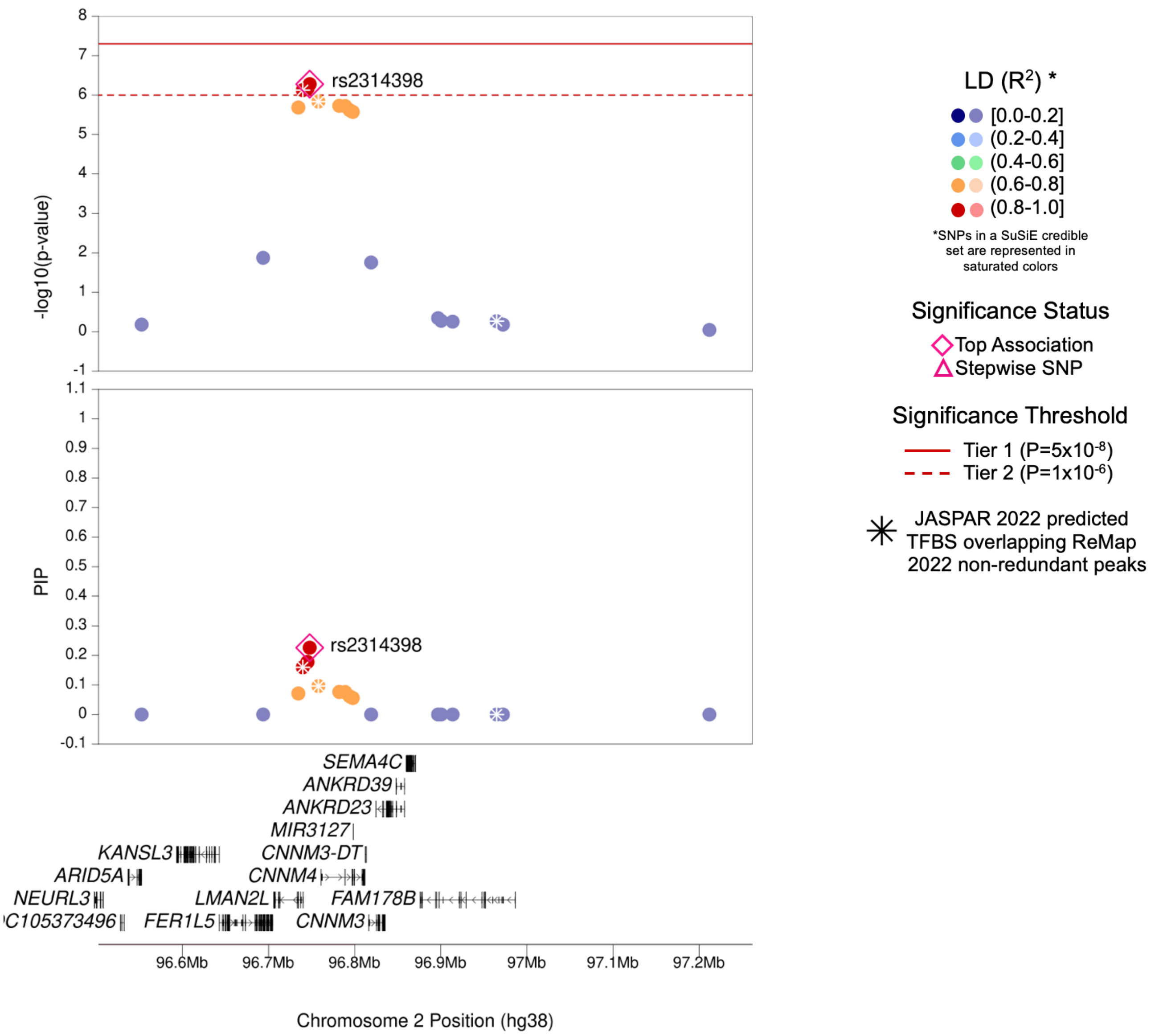

| | |
|---|---|
| **Region Name** | *CNNM4* |
| **Stepwise Region Window (hg38)** | chr2:96247751-97247751 |
| **Regional P-Value (df)** | $P = 5.28 \times 10^{-7}$ (df = 1) |
| **Novel for JIA (Yes/No)** | **Yes** |
| **Number of Signals: Stepwise** | 1 |
| **Number of Credible Sets: SuSiE (95% CS)** | 1 |
| **T1/T2 + Stepwise TF Overlap (Yes/No)** | Yes (1 TFBS for TF SCRT1) |
| **Implicated GTEx eGenes* and JIA PBMC eGenes† linked to JIA significant SNPs‡ or their proxies** | *AC159540.1; ACTR1B; ARID5A; CIAO1; CNNM4; LMAN2L* |
| **Implicated GTEx eGenes (All Tissues) linked to JIA significant SNPs‡ or their proxies** | *AC159540.1; ACTR1B; ADRA2B; ANKRD36; ANKRD36B; ANKRD39; ARID5A; CIAO1; CNNM3; CNNM4; FAHD2B; FAHD2CP; FAM178B; GPAT2; GPAT2P1; ITPRIPL1; LINC00342; LMAN2L; TMEM127* |
| **DGIdb has drug-gene interactions between GTEx eGenes* and JIA PBMC eGenes†** | No |
| **Region Implicated in Autoimmune Disease (Yes/No)** | **No (As of 2025-02-05)** |

*Limited to EBV-transformed lymphocytes, cultured fibroblasts, whole blood, and spleen tissues

†JIA eGenes for peripheral blood mononuclear cells (PBMCs) from Barnes et al. (PMID: 19565513) were defined as genes with $P_{eQTL} < 1 \times 10^{-4}$

‡JIA significant SNPs defined as SNPs with Best $P_{FDR} < 0.05$

### (ai) *TMEM39A–CD80* Region (Tier 2)

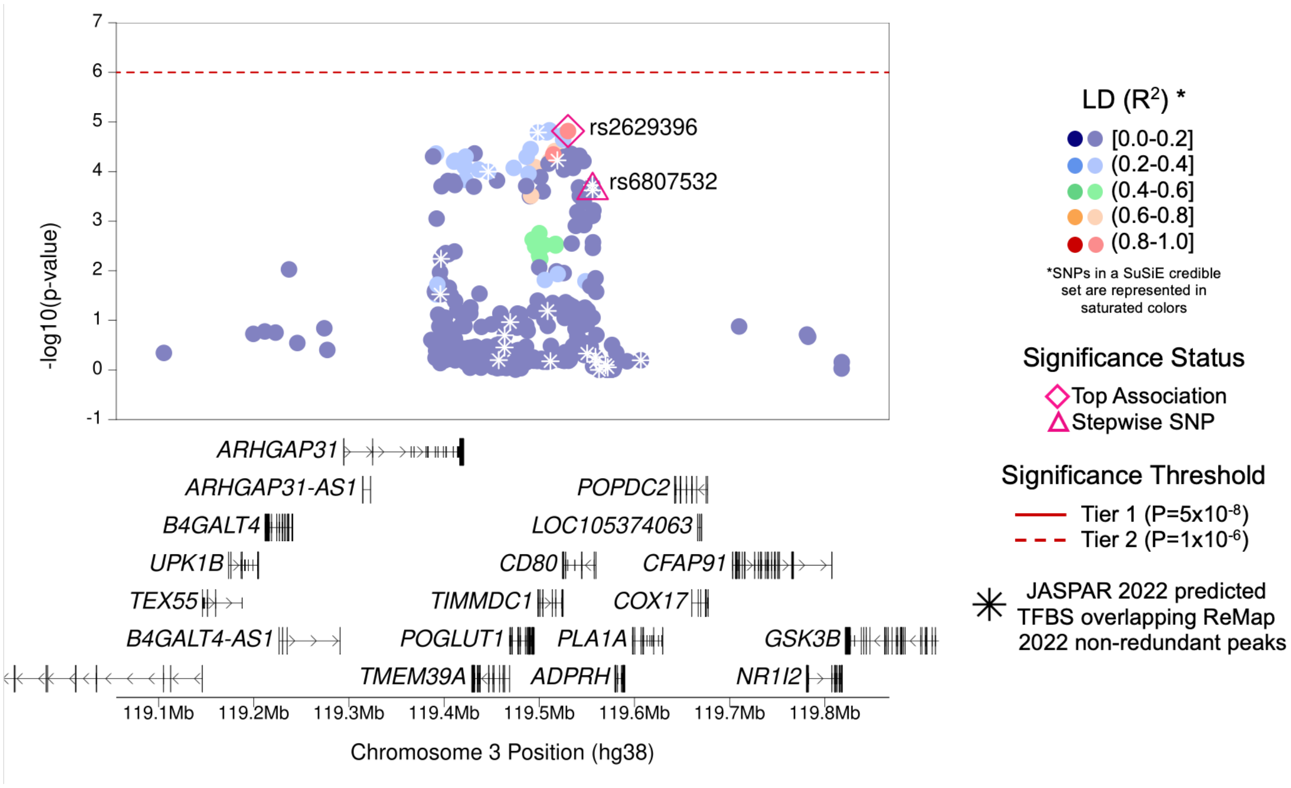

| | |
|---|---|
| **Region Name** | *TMEM39A–CD80* |
| **Stepwise Region Window (hg38)** | chr3:119030120-120030120 |
| **Regional P-Value (df)** | P = $2.00 \times 10^{-7}$ (df = 2) |
| **Novel for JIA (Yes/No)** | No |
| **Number of Signals: Stepwise** | 2 |
| **Number of Credible Sets: SuSiE (95% CS)** | 0 (Large number of non-additive SNPs in region; SuSiE not computed) |
| **T1/T2 + Stepwise TF Overlap (Yes/No)** | Yes (1 TFBS with TF STAT3) |
| **Implicated GTEx eGenes* and JIA PBMC eGenes† linked to JIA significant SNPs‡ or their proxies** | *POGLUT1* |
| **Implicated GTEx eGenes (All Tissues) linked to JIA significant SNPs‡ or their proxies** | *ADPRH; POGLUT1; TIMMDC1; TMEM39A* |
| **DGIdb has drug-gene interactions between GTEx eGenes* and JIA PBMC eGenes†** | No |
| **Region Implicated in Autoimmune Disease (Yes/No)** | Yes |

*Limited to EBV-transformed lymphocytes, cultured fibroblasts, whole blood, and spleen tissues

†JIA eGenes for peripheral blood mononuclear cells (PBMCs) from Barnes et al. (PMID: 19565513) were defined as genes with $P_{eQTL} < 1 \times 10^{-4}$

‡JIA significant SNPs defined as SNPs with Best $P_{FDR} < 0.05$

### (aj) *PRDM1* Region (Tier 2)

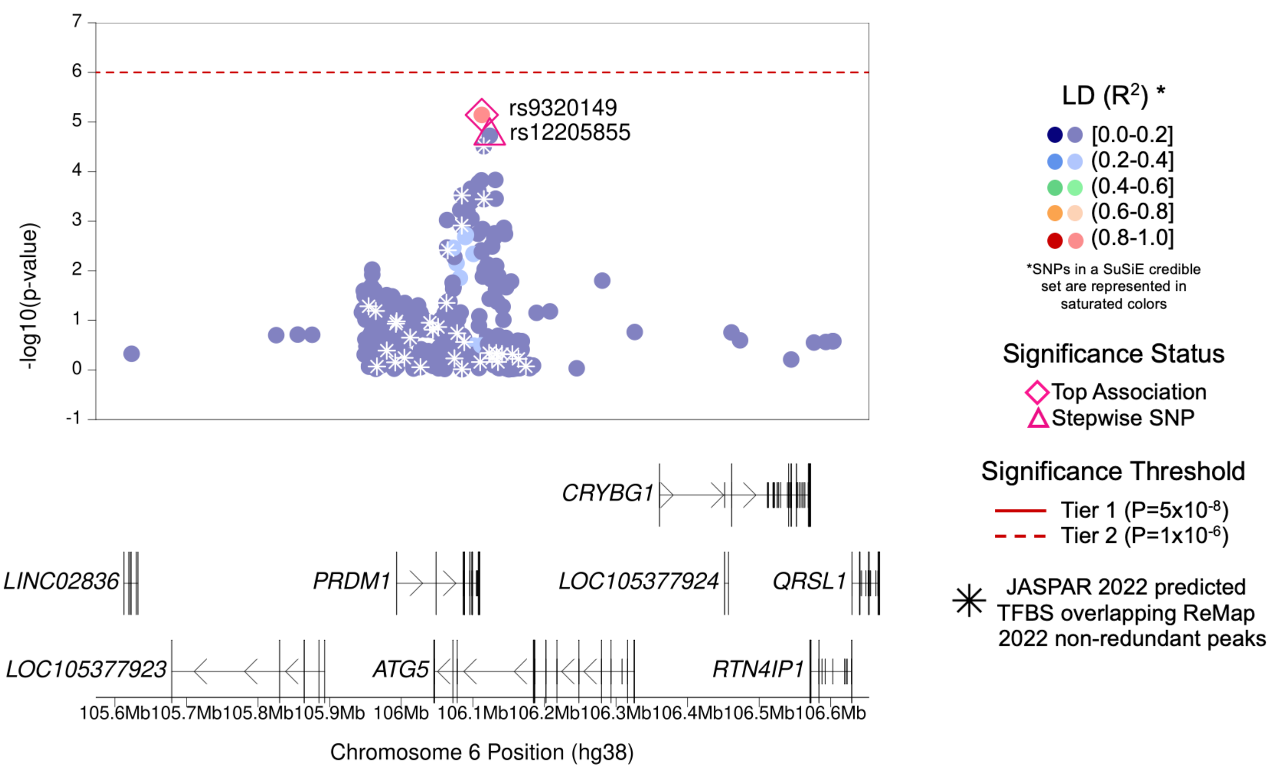

| | |
|---|---|
| **Region Name** | *PRDM1* |
| **Stepwise Region Window (hg38)** | chr6:105612560-106612560 |
| **Regional P-Value (df)** | P = $1.36 \times 10^{-7}$ (df = 2) |
| **Novel for JIA (Yes/No)** | **Yes** |
| **Number of Signals: Stepwise** | 2 |
| **Number of Credible Sets: SuSiE (95% CS)** | 0 (Large number of non-additive SNPs in region; SuSiE not computed) |
| **T1/T2 + Stepwise TF Overlap (Yes/No)** | No |
| **Implicated GTEx eGenes* and JIA PBMC eGenes† linked to JIA significant SNPs‡ or their proxies** | *ATG5* |
| **Implicated GTEx eGenes (All Tissues) linked to JIA significant SNPs‡ or their proxies** | NA (No eGenes) |
| **DGIdb has drug-gene interactions between GTEx eGenes* and JIA PBMC eGenes†** | Yes |
| **Region Implicated in Autoimmune Disease (Yes/No)** | Yes |

*Limited to EBV-transformed lymphocytes, cultured fibroblasts, whole blood, and spleen tissues

†JIA eGenes for peripheral blood mononuclear cells (PBMCs) from Barnes et al. (PMID: 19565513) were defined as genes with $P_{eQTL} < 1 \times 10^{-4}$

‡JIA significant SNPs defined as SNPs with Best $P_{FDR} < 0.05$

### (ak) *TRAF1* Region (Tier 2)

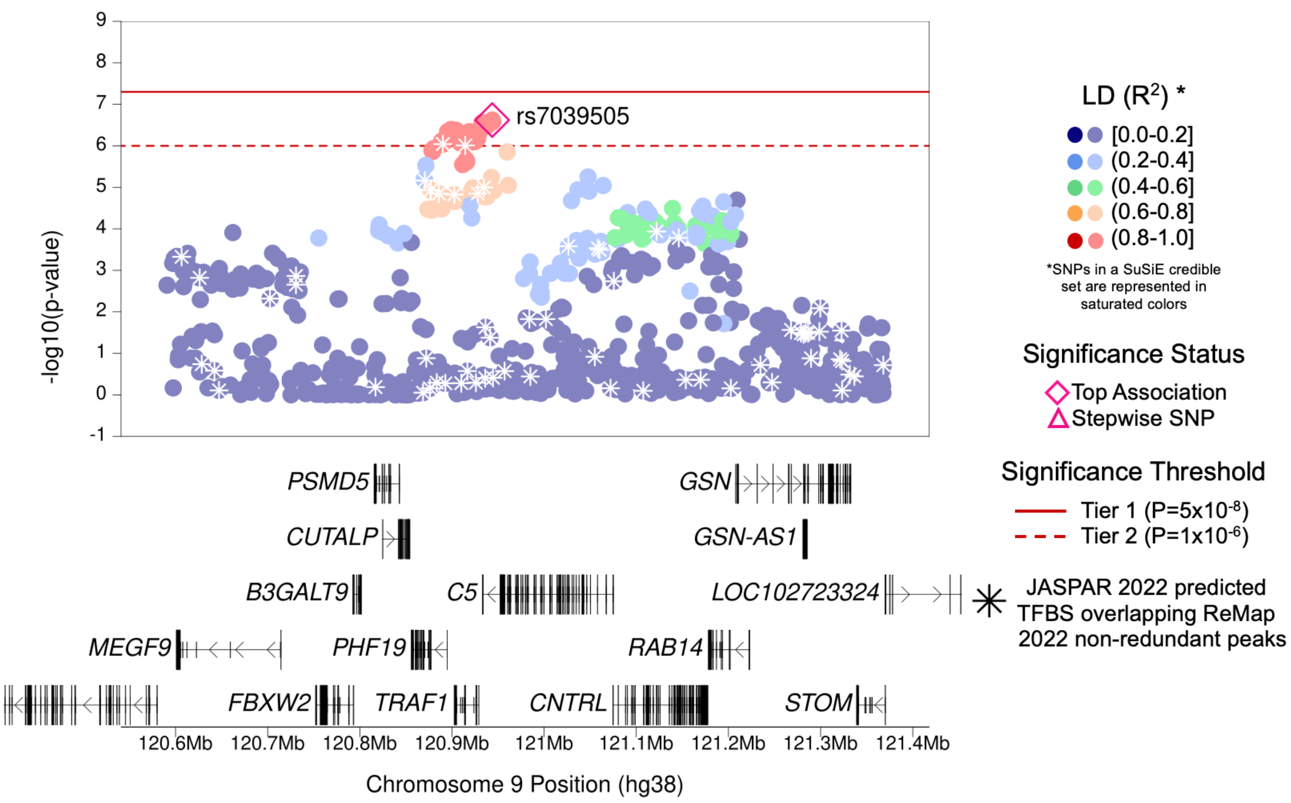

| | |
|---|---|
| **Region Name** | *TRAF1* |
| **Stepwise Region Window (hg38)** | chr9:120443667-121443667 |
| **Regional P-Value (df)** | P = $2.39x10^{-7}$ (df = 1) |
| **Novel for JIA (Yes/No)** | No |
| **Number of Signals: Stepwise** | 1 |
| **Number of Credible Sets: SuSiE (95% CS)** | 0 (Large number of non-additive SNPs in region; SuSiE not computed) |
| **T1/T2 + Stepwise TF Overlap (Yes/No)** | No |
| **Implicated GTEx eGenes* and JIA PBMC eGenes† linked to JIA significant SNPs‡ or their proxies** | *C5; CNTRL; FBXW2; GGTA1P; LOC253039; PHF19; PSMD5; PSMD5-AS1; TRAF1* |
| **Implicated GTEx eGenes (All Tissues) linked to JIA significant SNPs‡ or their proxies** | *AHCYP2; C5; CNTRL; FBXW2; GGTA1P; MEGF9; PHF19; PSMD5; PSMD5-AS1; RAB14; RP11-27I1.6; TRAF1* |
| **DGIdb has drug-gene interactions between GTEx eGenes* and JIA PBMC eGenes†** | Yes |
| **Region Implicated in Autoimmune Disease (Yes/No)** | Yes |

*Limited to EBV-transformed lymphocytes, cultured fibroblasts, whole blood, and spleen tissues

†JIA eGenes for peripheral blood mononuclear cells (PBMCs) from Barnes et al. (PMID: 19565513) were defined as genes with $P_{eQTL} < 1x10^{-4}$

‡JIA significant SNPs defined as SNPs with Best $P_{FDR} < 0.05$

### (al) *LTBR–CD27-AS1* Region (Tier 2)

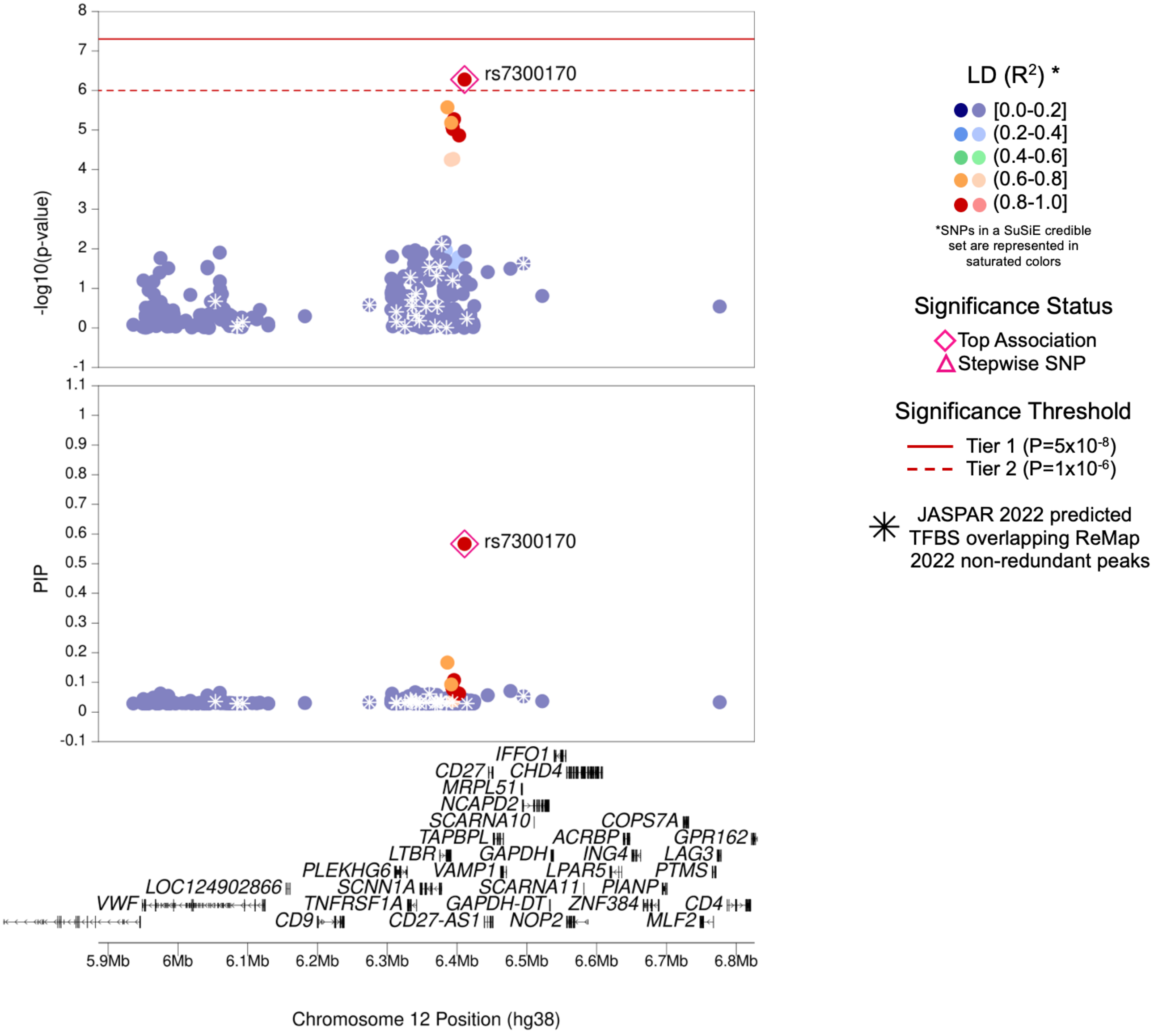

| Region Name | *LTBR–CD27-AS1* |
|---|---|
| **Stepwise Region Window (hg38)** | chr12:5910671-6910671 |
| **Regional P-Value (df)** | $P = 5.28x10^{-7}$ (df = 1) |
| **Novel for JIA (Yes/No)** | No |
| **Number of Signals: Stepwise** | 1 |
| **Number of Credible Sets: SuSiE (95% CS)** | 1 |
| **T1/T2 + Stepwise TF Overlap (Yes/No)** | No |
| **Implicated GTEx eGenes* and JIA PBMC eGenes† linked to JIA significant SNPs‡ or their proxies** | *LTBR* |
| **Implicated GTEx eGenes (All Tissues) linked to JIA significant SNPs‡ or their proxies** | *LTBR; RP1-102E24.8* |
| **DGIdb has drug-gene interactions between GTEx eGenes* and JIA PBMC eGenes†** | No |
| **Region Implicated in Autoimmune Disease (Yes/No)** | Yes |

*Limited to EBV-transformed lymphocytes, cultured fibroblasts, whole blood, and spleen tissues

†JIA eGenes for peripheral blood mononuclear cells (PBMCs) from Barnes et al. (PMID: 19565513) were defined as genes with $P_{eQTL} < 1x10^{-4}$

‡JIA significant SNPs defined as SNPs with Best $P_{FDR} < 0.05$

### (am) *IKZF4–ERBB3* Region (Tier 2)

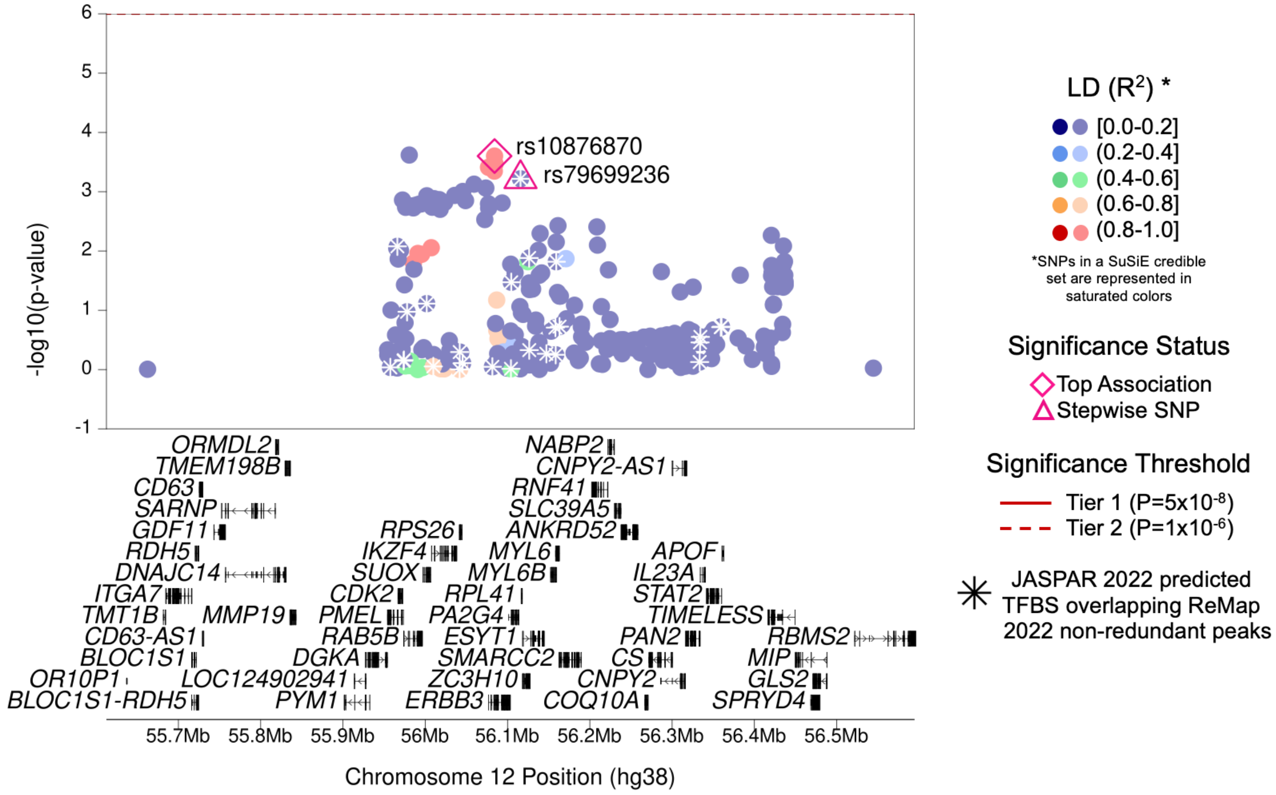

| | |
|---|---|
| **Region Name** | *IKZF4–ERBB3* |
| **Stepwise Region Window (hg38)** | chr12:55584218-56584218 |
| **Regional P-Value (df)** | P = $5.17 \times 10^{-7}$ (df = 1) |
| **Novel for JIA (Yes/No)** | **Yes** |
| **Number of Signals: Stepwise** | 1 |
| **Number of Credible Sets: SuSiE (95% CS)** | 0 (Large number of non-additive SNPs in region; SuSiE not computed) |
| **T1/T2 + Stepwise TF Overlap (Yes/No)** | Yes (1 TFBS with TF MZF1) |
| **Implicated GTEx eGenes* and JIA PBMC eGenes† linked to JIA significant SNPs‡ or their proxies** | *PAN2; RPS26; SUOX* |
| **Implicated GTEx eGenes (All Tissues) linked to JIA significant SNPs‡ or their proxies** | *CNPY2; ERBB3; GDF11; IKZF4; IL23A; MYL6B; PAN2; PMEL; RAB5B; RP11-977G19.11; RPS26; SPRYD4; STAT2; SUOX; ZC3H10* |
| **DGIdb has drug-gene interactions between GTEx eGenes* and JIA PBMC eGenes†** | Yes |
| **Region Implicated in Autoimmune Disease (Yes/No)** | Yes |

*Limited to EBV-transformed lymphocytes, cultured fibroblasts, whole blood, and spleen tissues

†JIA eGenes for peripheral blood mononuclear cells (PBMCs) from Barnes et al. (PMID: 19565513) were defined as genes with $P_{eQTL} < 1 \times 10^{-4}$

‡JIA significant SNPs defined as SNPs with Best $P_{FDR} < 0.05$

### (an) *IFNG-AS1–IFNG* Region (Tier 2)

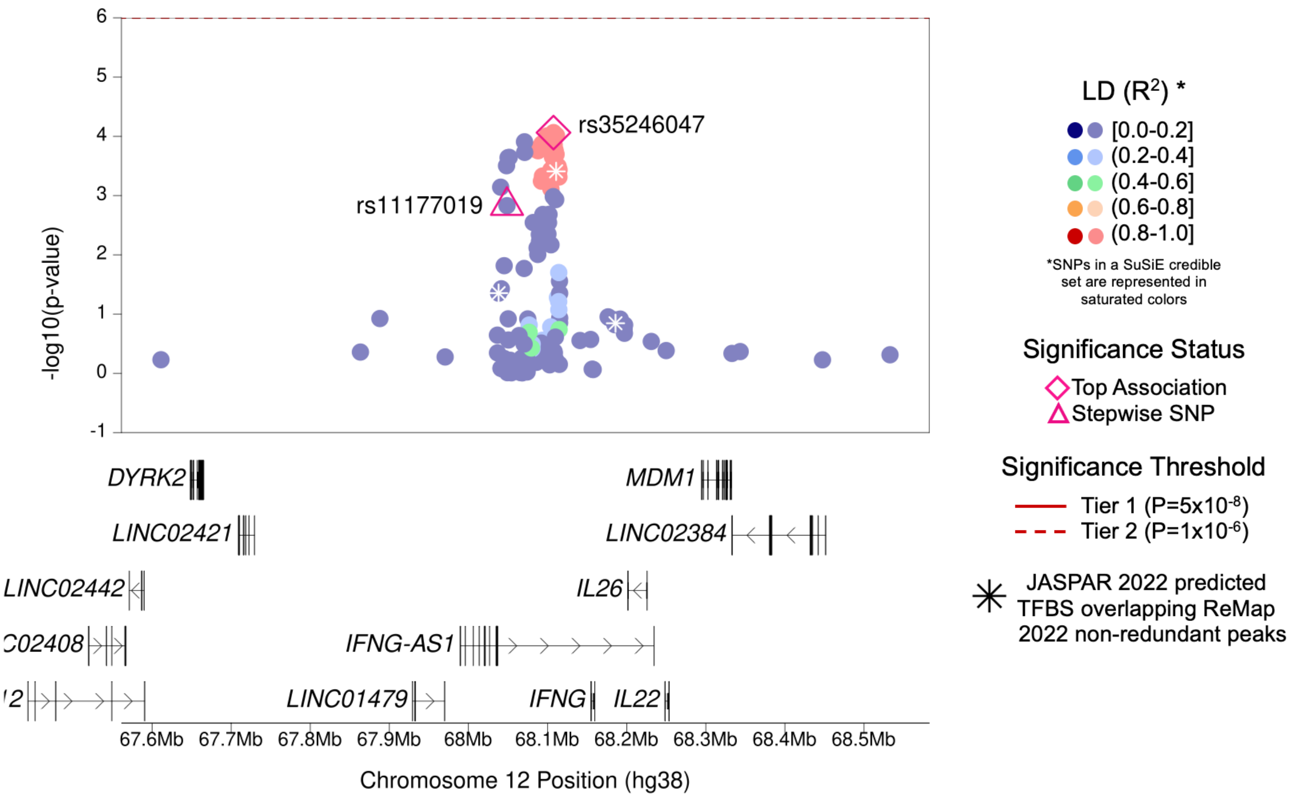

| | |
|---|---|
| **Region Name** | *IFNG-AS1–IFNG* |
| **Stepwise Region Window (hg38)** | chr12:67607473-68607473 |
| **Regional P-Value (df)** | $P = 3.48x10^{-7}$ (df = 1) |
| **Novel for JIA (Yes/No)** | **Yes** |
| **Number of Signals: Stepwise** | 1 |
| **Number of Credible Sets: SuSiE (95% CS)** | 0 (Large number of non-additive SNPs in region; SuSiE not computed) |
| **T1/T2 + Stepwise TF Overlap (Yes/No)** | No |
| **Implicated GTEx eGenes* and JIA PBMC eGenes† linked to JIA significant SNPs‡ or their proxies** | *IFNG-AS1* |
| **Implicated GTEx eGenes (All Tissues) linked to JIA significant SNPs‡ or their proxies** | *IFNG-AS1* |
| **DGIdb has drug-gene interactions between GTEx eGenes* and JIA PBMC eGenes†** | No |
| **Region Implicated in Autoimmune Disease (Yes/No)** | Yes |

*Limited to EBV-transformed lymphocytes, cultured fibroblasts, whole blood, and spleen tissues

†JIA eGenes for peripheral blood mononuclear cells (PBMCs) from Barnes et al. (PMID: 19565513) were defined as genes with $P_{eQTL} < 1x10^{-4}$

‡JIA significant SNPs defined as SNPs with Best $P_{FDR} < 0.05$

### (ao) *COG6* Region (Tier 2)

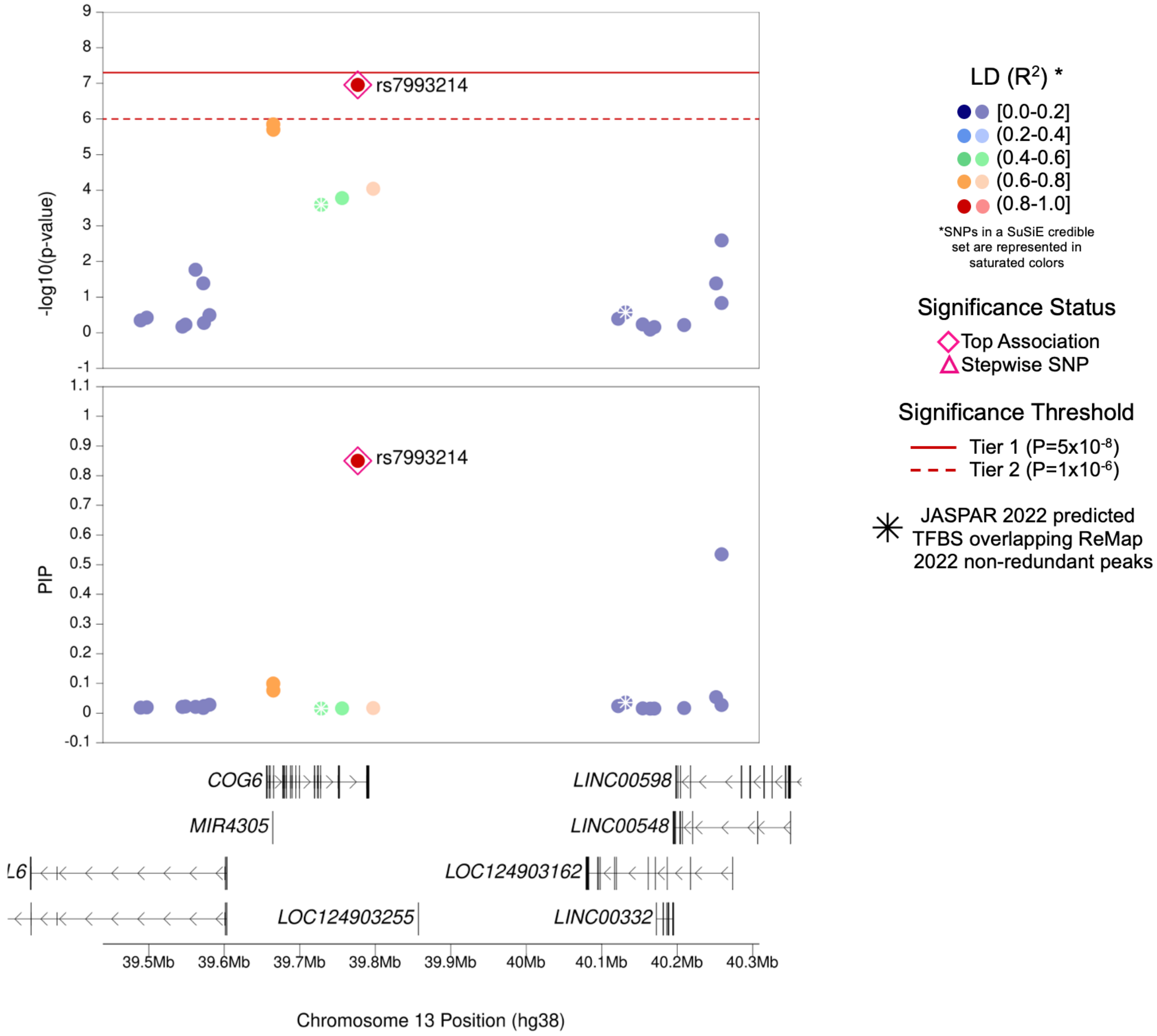

| | |
|---|---|
| **Region Name** | *COG6* |
| **Stepwise Region Window (hg38)** | chr13:39276775-40276775 |
| **Regional P-Value (df)** | P = $1.10 \times 10^{-7}$ (df = 1) |
| **Novel for JIA (Yes/No)** | No |
| **Number of Signals: Stepwise** | 1 |
| **Number of Credible Sets: SuSiE (95% CS)** | 1 |
| **T1/T2 + Stepwise TF Overlap (Yes/No)** | No |
| **Implicated GTEx eGenes* and JIA PBMC eGenes† linked to JIA significant SNPs‡ or their proxies** | *COG6; NHLRC3* |
| **Implicated GTEx eGenes (All Tissues) linked to JIA significant SNPs‡ or their proxies** | *COG6; NHLRC3* |
| **DGIdb has drug-gene interactions between GTEx eGenes* and JIA PBMC eGenes†** | No |
| **Region Implicated in Autoimmune Disease (Yes/No)** | Yes |

*Limited to EBV-transformed lymphocytes, cultured fibroblasts, whole blood, and spleen tissues

†JIA eGenes for peripheral blood mononuclear cells (PBMCs) from Barnes et al. (PMID: 19565513) were defined as genes with $P_{eQTL} < 1 \times 10^{-4}$

‡JIA significant SNPs defined as SNPs with Best $P_{FDR} < 0.05$

**Supplementary Figure 3. eQTLs of JIA-associated SNPs and LD proxies.**

JIA-associated SNPs meeting Tier 1, 2, or 3 ($p_{FDR} < 0.05$) and proxies selected by linkage disequilibrium ($r^2 \geq 0.5$ in 1000 EUR population) were queried for eQTL status in GTEx-relevant tissues (whole blood, Cells-Cultured Fibroblasts, Cells-EBV-Transformed Lymphocytes and Spleen) and a dataset of eQTLs in PBMCs within previously published JIA study [20]. Only eQTLs meeting p-value < $1x10^{-4}$ (red dashed line) are plotted. Peaks are labeled by associated eGENE. Note: Y-axis maximum is dependent on tissue.

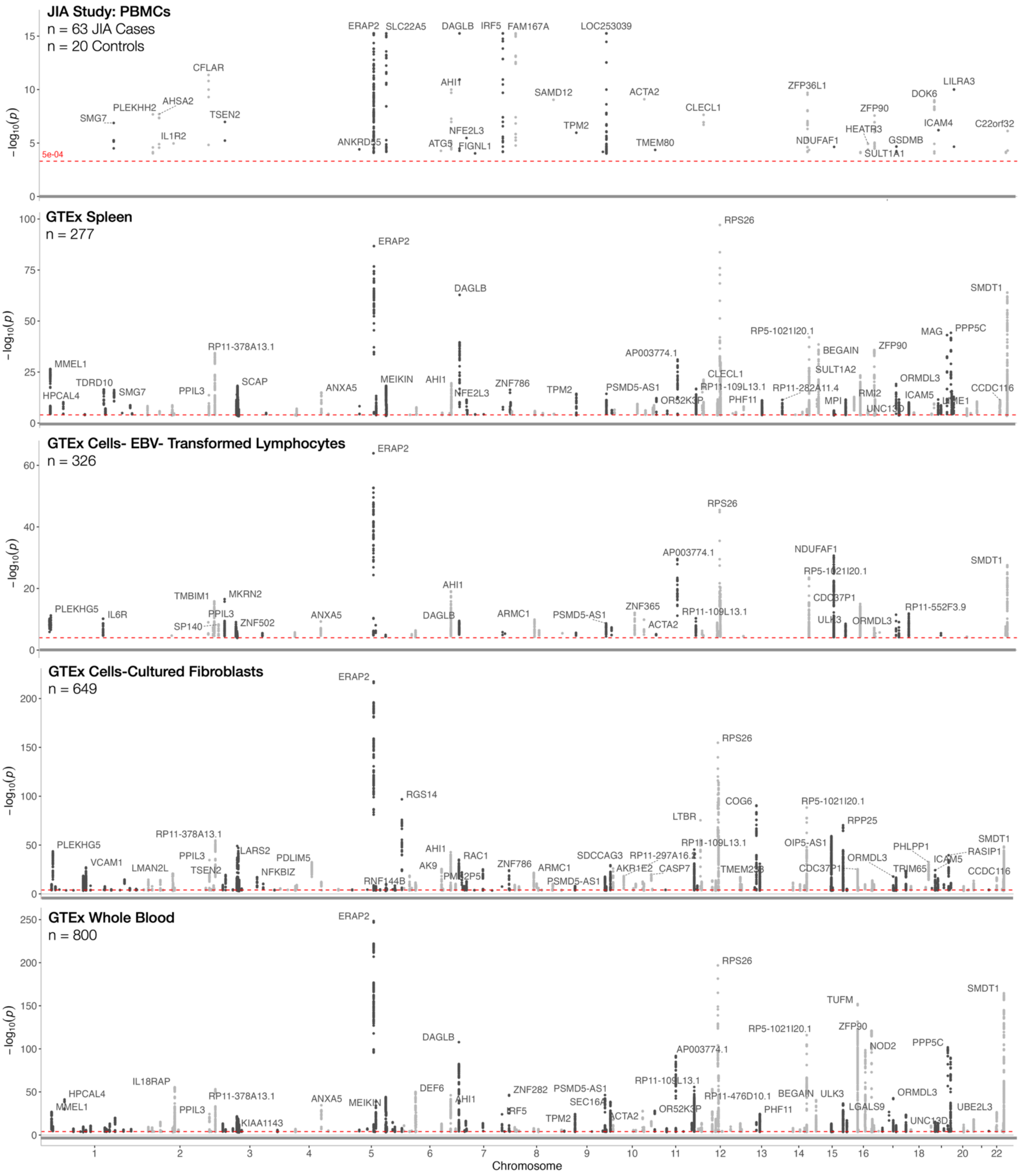

**Supplementary Figure 4: JIA Associated Genes implicated in autoimmune diseases as designated by the GWAS Catalog.**
Many of the established and novel genomic regions to JIA have been implicated across other autoimmune diseases. **A**. Number of shared loci with established JIA-associated regions (n=27 regions). **B**. Number of shared loci with regions that are novel to JIA (n=14 regions). The relative frequency of the diseases implicated by the genomic regions tended to differ between established and novel loci (chi-squared test: p=0.0033), with the novel regions exhibiting an overrepresentation of inflammatory bowel disease, Crohn's disease, systemic lupus erythematosus, and ulcerative colitis; and an underrepresentation of Celiac's diseases, ankylosing spondylitis.

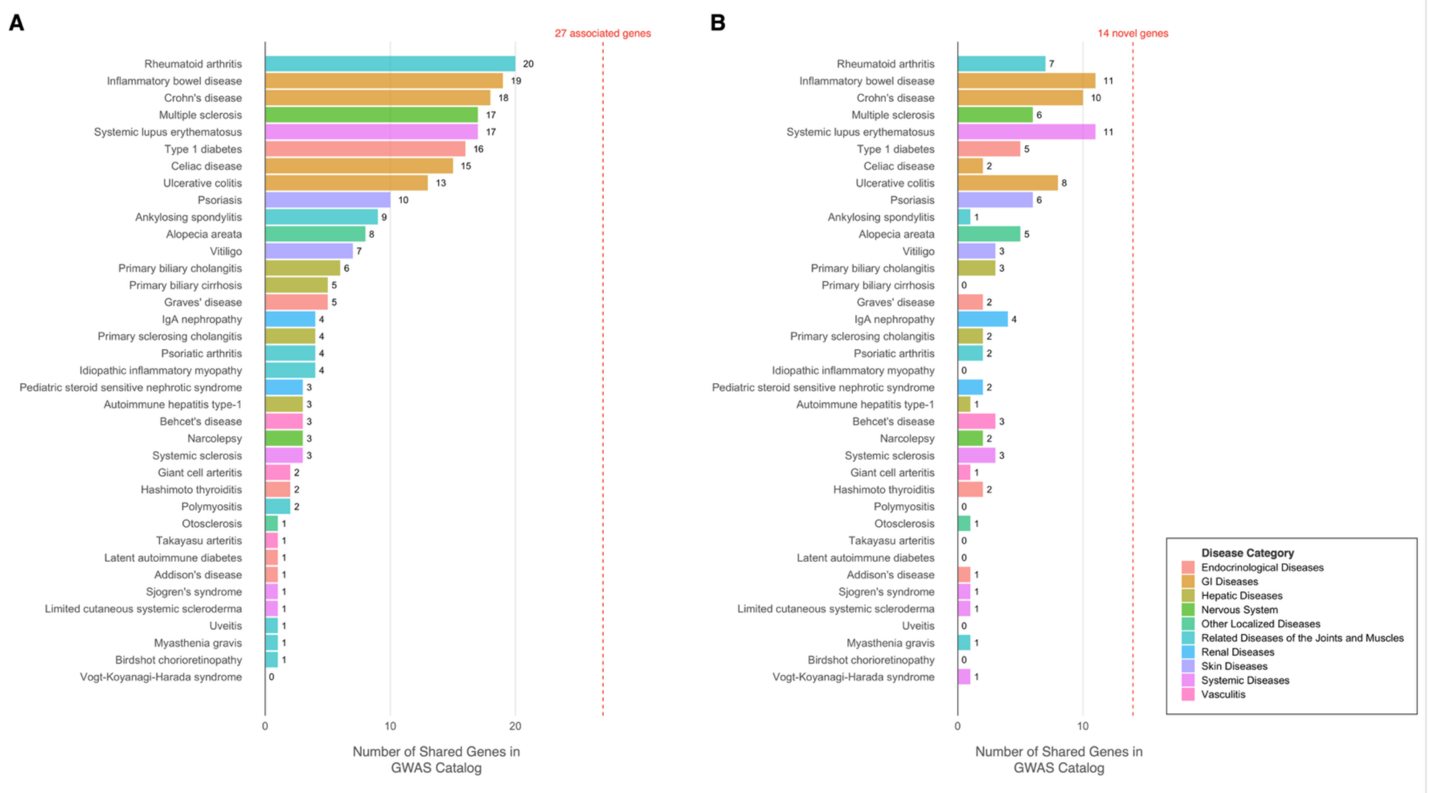

**Supplementary Figure 5: HLA Associations compared to risk allele frequencies.**

JIA associations with Class I (top panel) and Class II (bottom panel) alleles by JIA case dosage frequency. Colors are differentiated across HLA genes. Dashed line denotes JIA risk alleles (odds ratio > 1; above line) or non-risk alleles (odds ratio < 1; below line). Alleles with comparable p-values and allele frequencies are likely co-inherited; diamond symbol denotes alleles selected for the multi-allelic model via stepwise modeling procedure. Among Class I alleles, A*02:01 exhibited the largest association signal and was the most frequent (0.38)among cases. Of the 18 Class I and Class II alleles selected for the multi-allelic model, 10 were from DRB1.

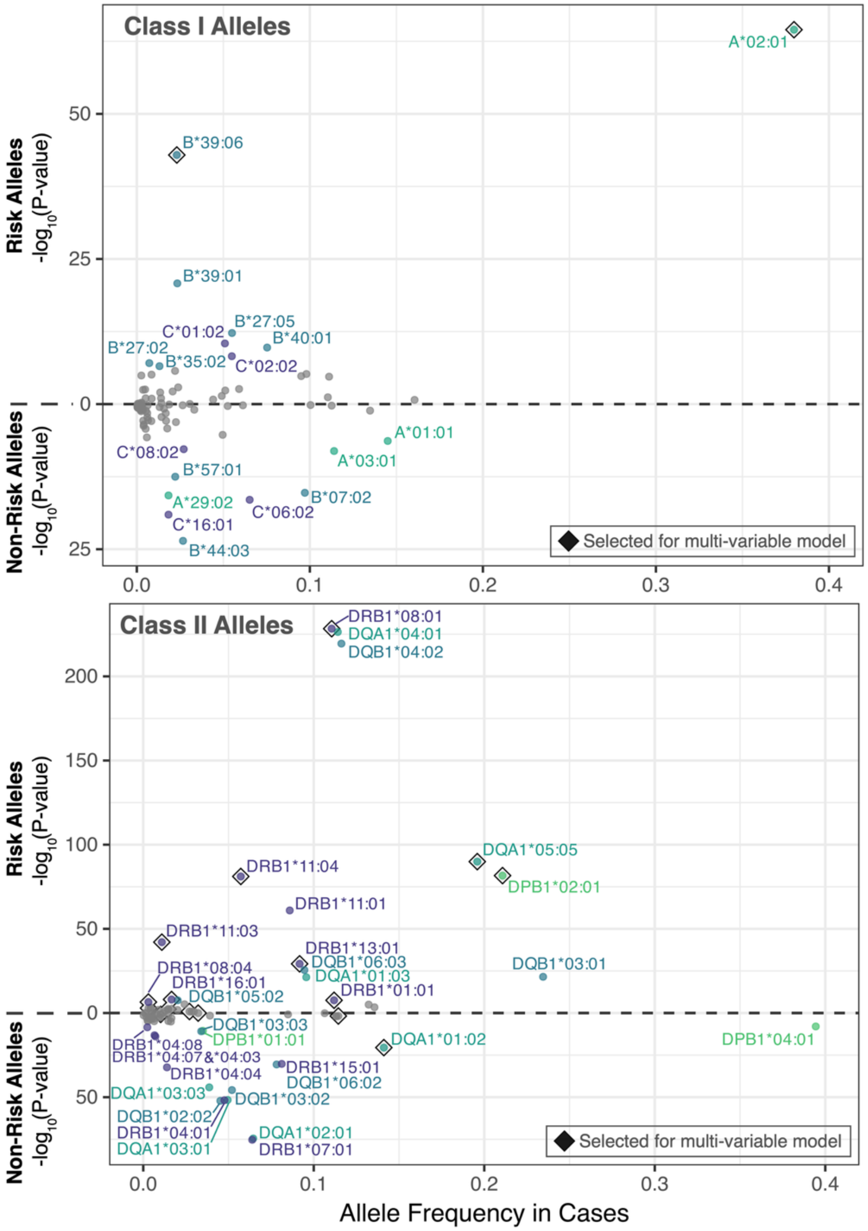

**Supplementary Figure 6: SNP associations across Major Histocompatibility (MHC), with and without adjustment for significant two-field HLA alleles.**
Mirror plot illustrates SNP associations across MHC. Dashed red lines indicates genome-wide significance $P=5x10^{-8}$. Gray data points (top half of plot) illustrate adjustment for population substructure. Purple data points (bottom half of plot) illustrate SNP associations when including the 18 two-field alleles selected by the stepwise procedure. Majority of SNP associations are diminished with inclusion of stepwise-selected two-field alleles.

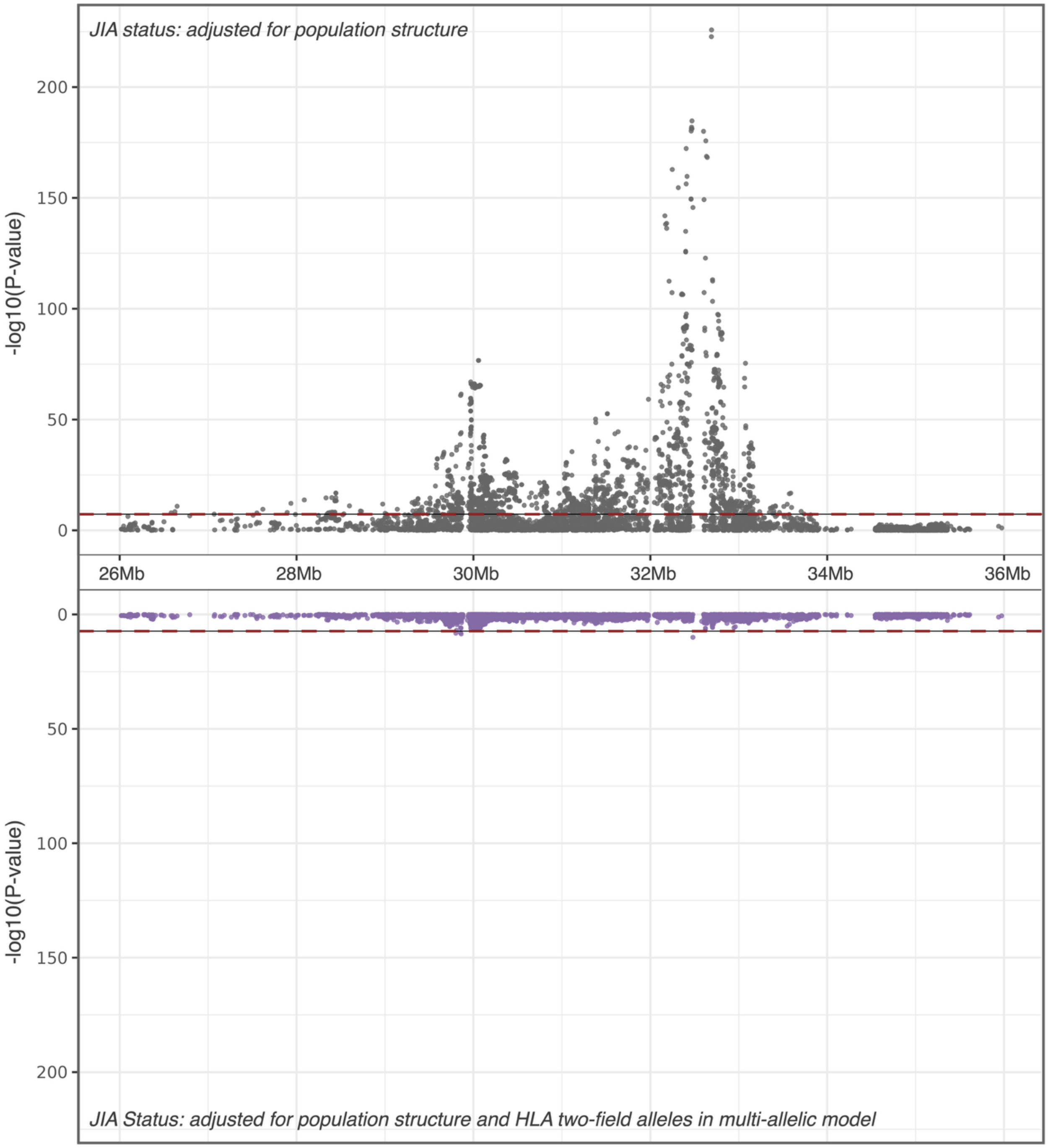

**Supplementary Figure 7: Dendrograms of HLA alleles by gene, based on sequence similarity.** Protein sequences for two-field HLA Alleles were analyzed for sequence similarity using Clustal Omega and the resulting dendrograms are shown. For each allele, association results from the single-allele analyses are included, as well as whether the allele was selected for the multi-allelic model (diamond symbol).

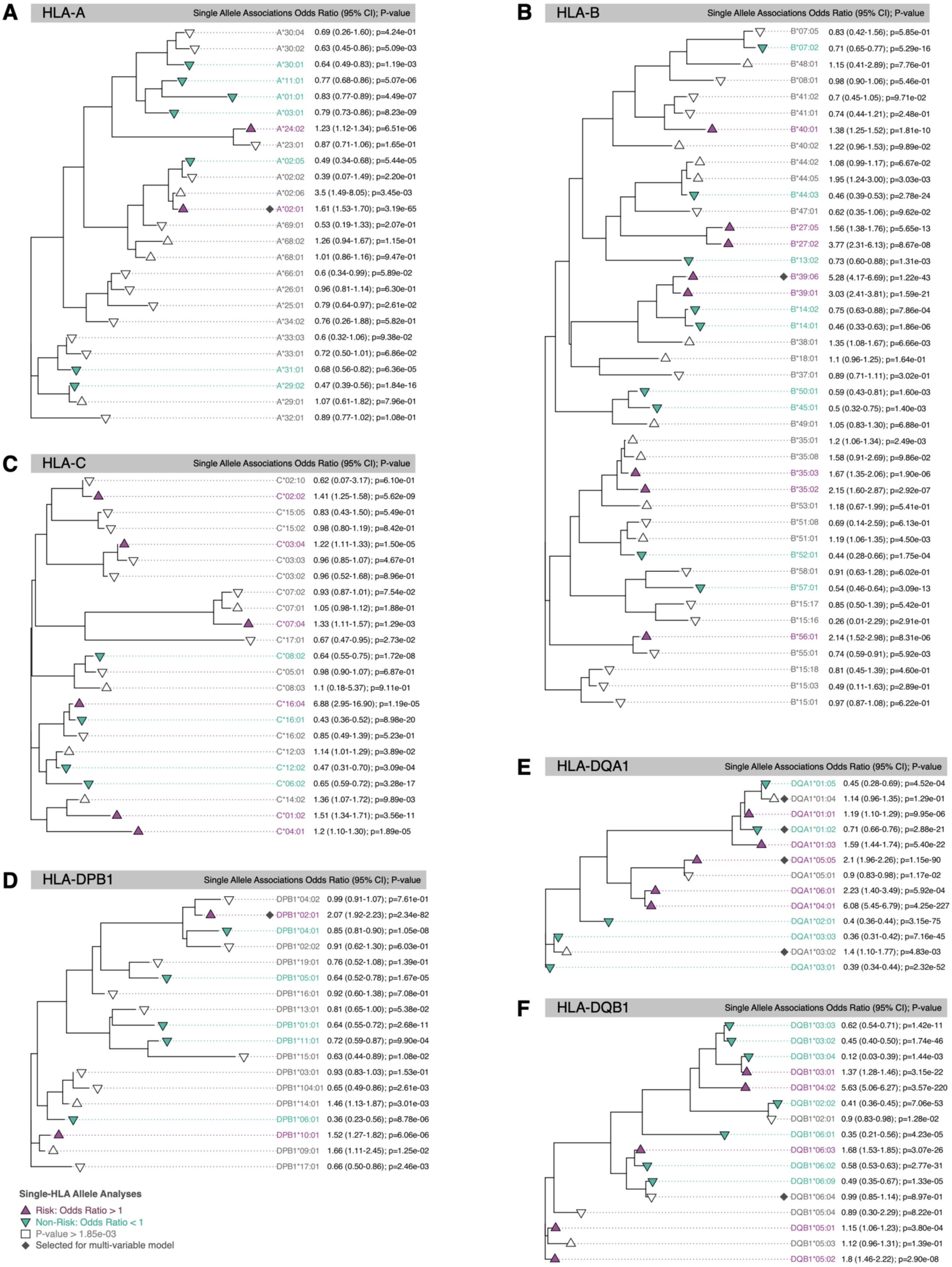

**Supplementary Figure 8: Sequence alignment of DRB1 alleles illustrates amino acid differentiation of two primary clusters based on sequence similarity.**

**A.** Amino acid sequence similarity of DRB1 alleles (depicted as dendrogram; primary figure 2) illustrates two main clusters. Notably, within “Cluster 2”, all alleles meeting significance exhibit consistent JIA risk. **B.** Using cluster 1 as reference, non-consensus amino acids for cluster 2 are highlighted in pink and annotated with arrows. Only three amino acids consistently differentiate between the two clusters.

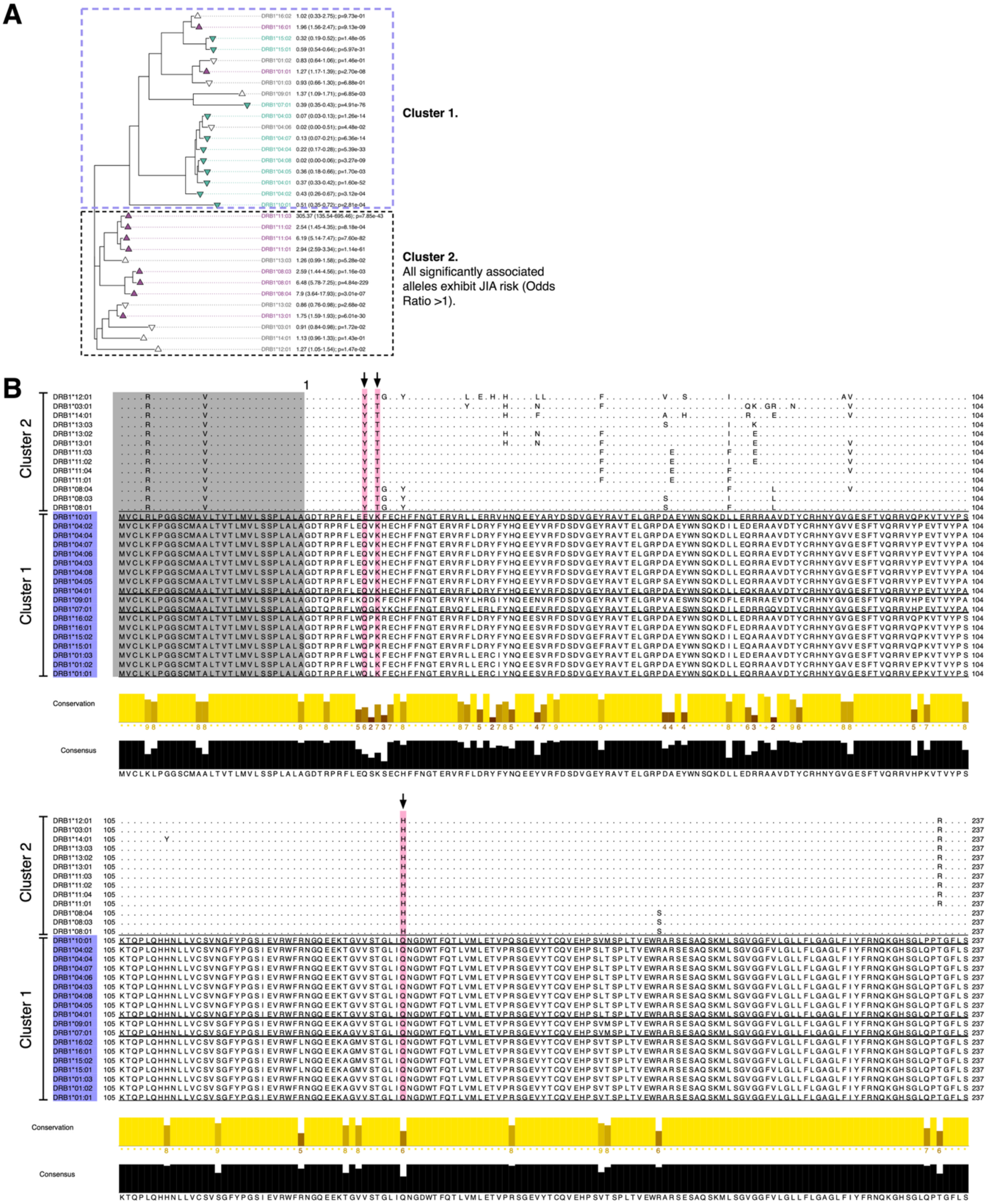

**Supplementary Figure 9: Sequence alignment of DRB1*01:01, DRB1*16:01, DRB1*15:02, and DRB1*15:01.**

**A.** Amino acid sequence similarity of DRB1 alleles (depicted as dendrogram; primary figure 2) identifies subcluster of alleles with 95% sequence similarity but differing odds ratios for JIA risk (DRB1*01:01 and DRB1*16:01 as risk; DRB1*15:02 and DRB1*15:01 as non-risk). **B.** Using the two non-risk alleles as reference, non-consensus amino acids for DRB1*01:01 and DRB1*16:01 are highlighted in pink and annotated with arrows. Only four amino acids (spanning Exons 1 and 2) differentiate between the two sets of alleles. **C.** Using PDB:5V4M (DR15) as a structural model shows that three of the amino acids (positions 47, 67, and 71) are in close proximity within the binding region. The fourth amino acid is within the leader peptide (not shown within 3D model).

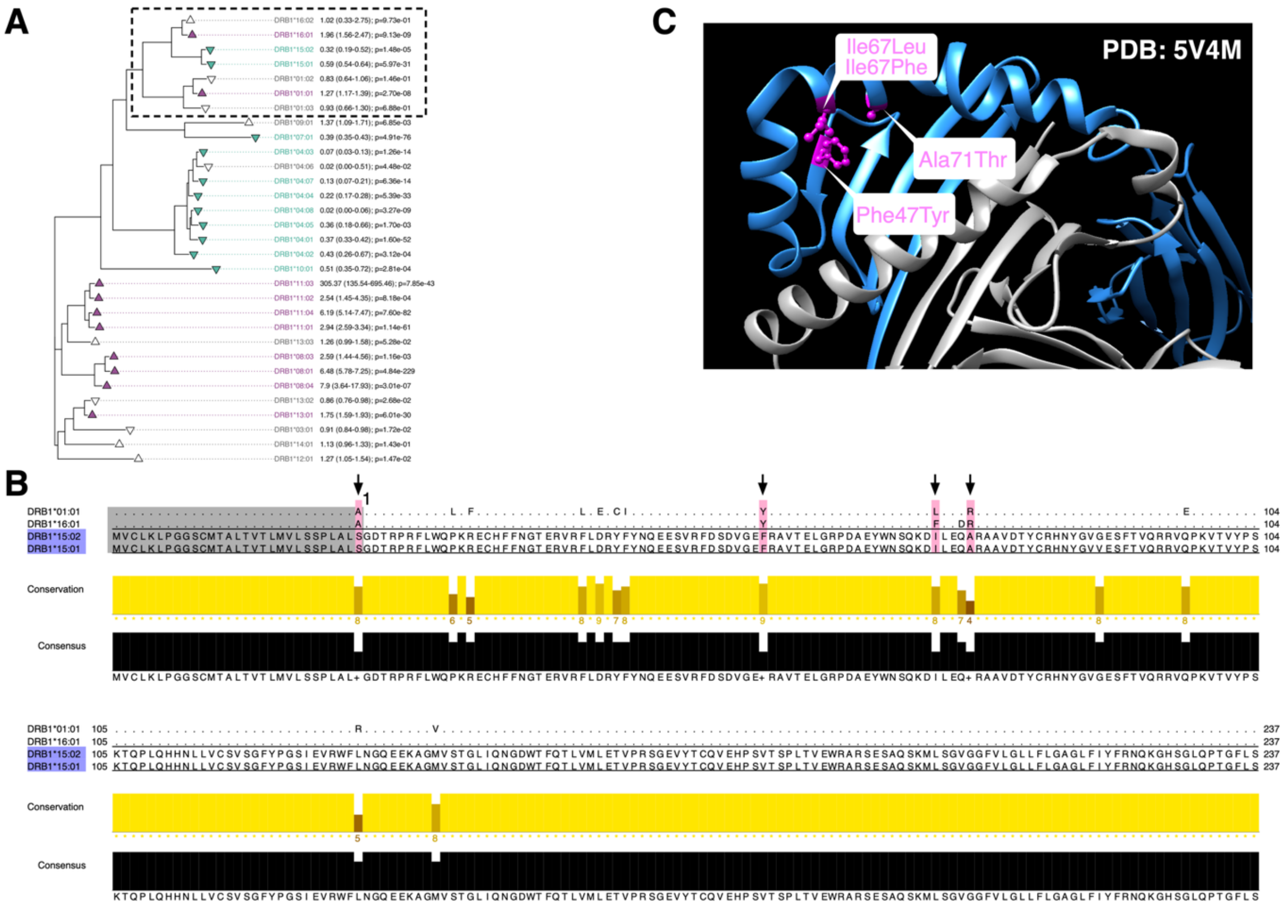

**Supplementary Figure 10: Genome-wide associations for age of onset only exhibits robust SNP associations within the HLA region on chromosome 6.**

Genome-wide associations for age of onset across 3000 JIA cases, adjusted for population substructure.

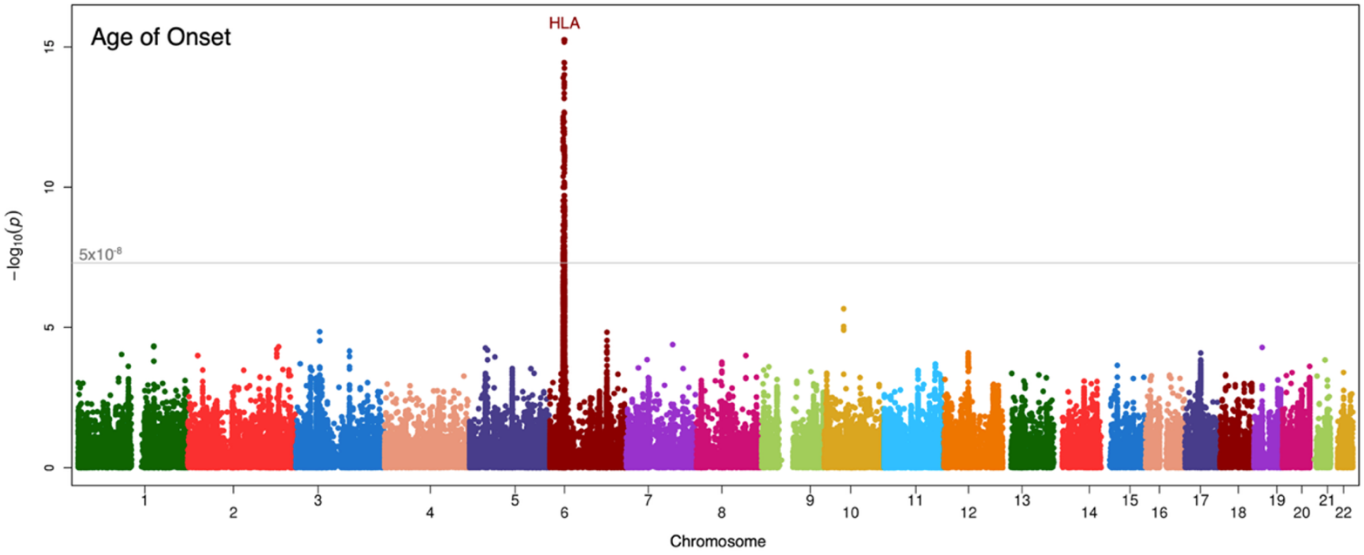

**Supplementary Figure 11: Genome-wide associations for interactions with A*02:01 (case-only analysis).**

For each of the HLA alleles selected by the stepwise modelling procedure, a case-only analysis was computed to a SNP-by-HLA interaction. A*02:01 was the only allele to exhibit an HLA-by-SNP interaction.

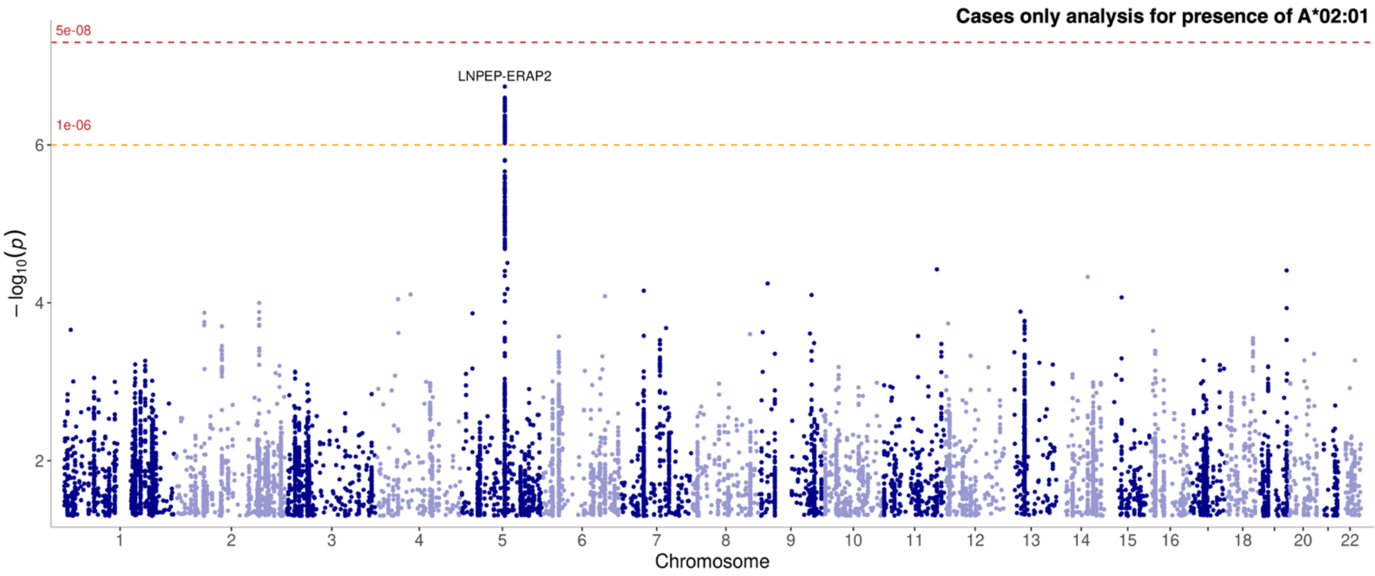

**Supplementary Figure 12: Differentiation of SNPs by DNA topology breaks up linkage disequilibrium.**

Within the STAT4 region, multiple SNPs reach T1 ($P<5x10^{-8}$) significance (red solid line) and are in LD. Evaluation of ΔMGW and ΔProT provides alternative prioritization, highlighting SNP rs11889341 over the initially labeled lead SNP.

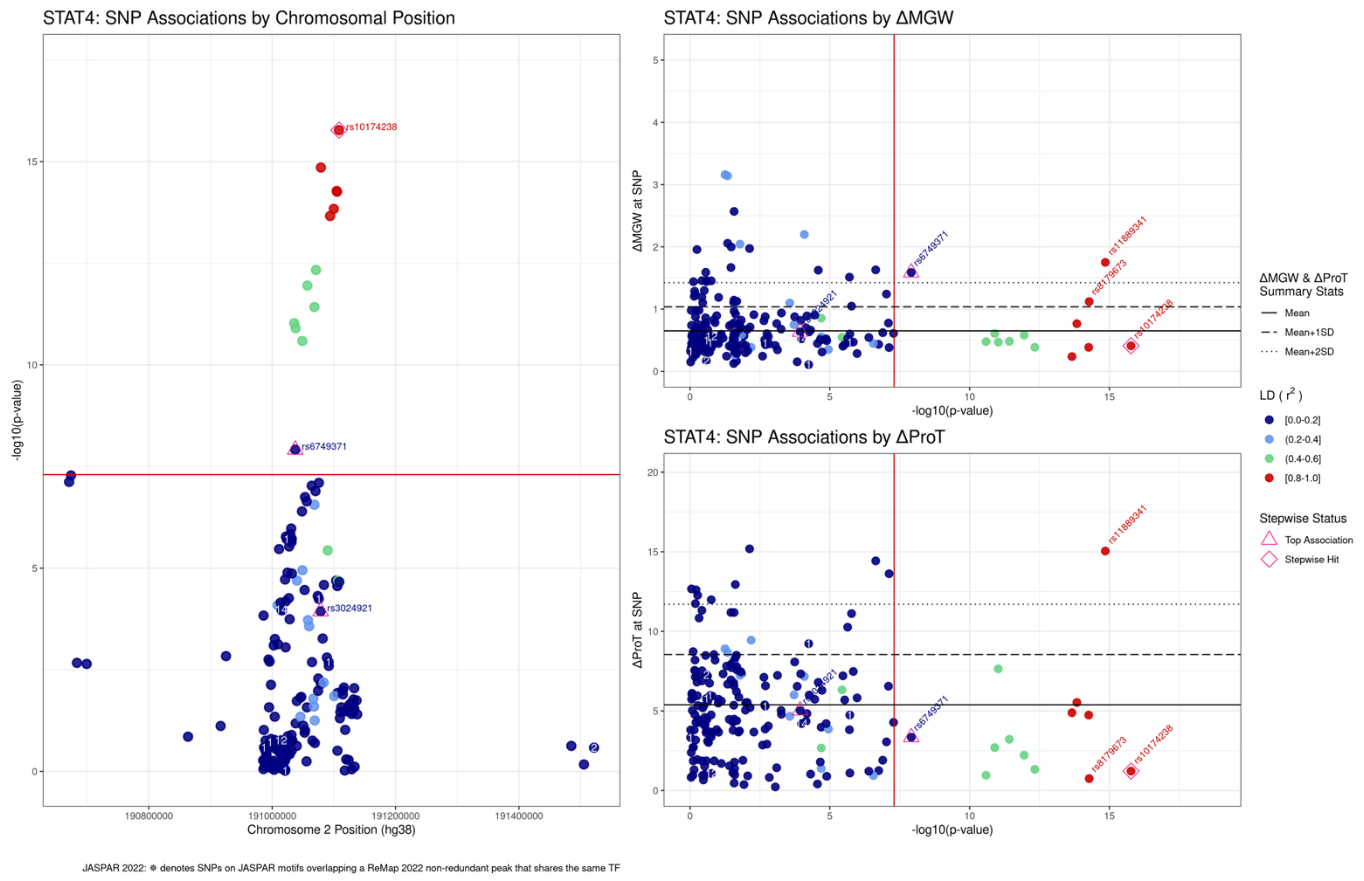

**Supplementary Figure 13: Study-level flow diagram describing the derivation of novel functional TFBS database and topological profiles.**

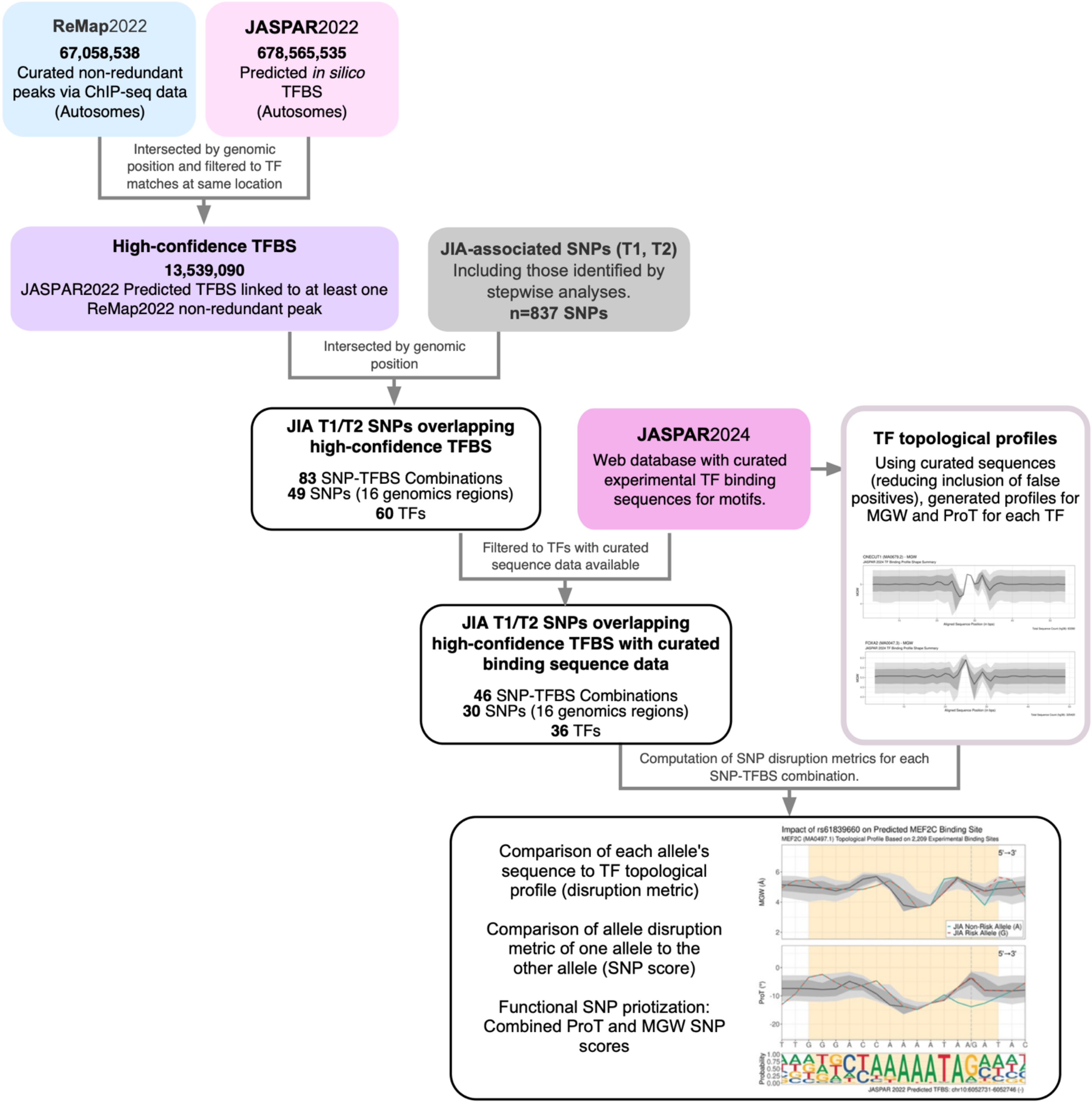

**Supplementary Figure 14: Inverse trend between minor allele frequency and SNP-TFBS disruption scores for 30 unique JIA-associated SNPs.**

A) Minor allele frequency (MAF) in controls compared to SNP-TFBS disruption scores. SNPs with larger MAF are correlated with smaller SNP-TFB disruption scores. Pearson's correlation shown. B) MAF in controls and SNP-TFBS are shown as ranks with Spearman's correlation. For both plots, SNPs with multiple TFBS were pruned to retain the largest SNP-TFBS score.

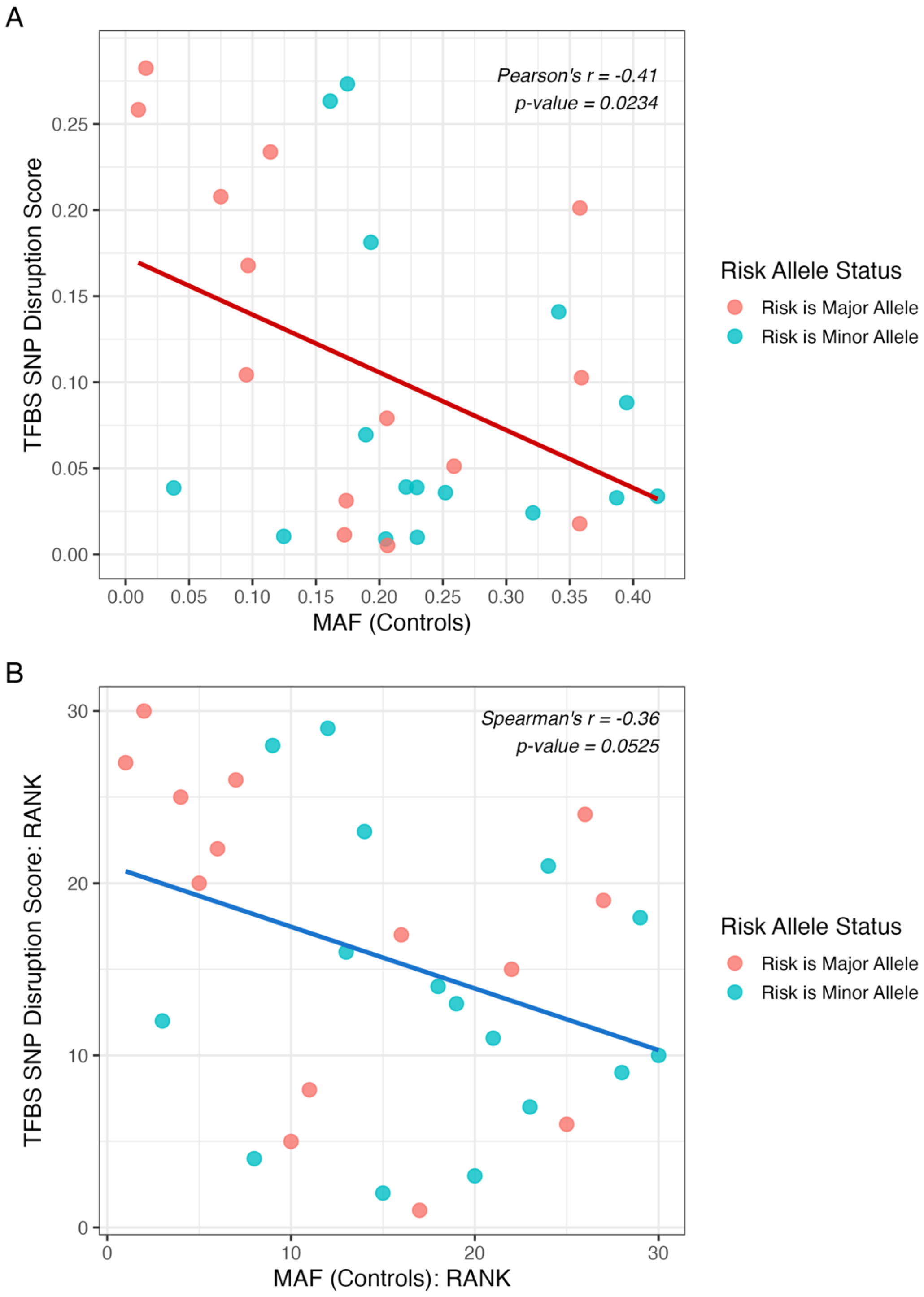

**Supplementary Figure 15: Impact of JIA-associated SNPs on predicted transcription factor binding sites (TFBS).**

**(a)-(at)** Minor groove width (MGW) and propellor twist (ProT) profiles for JASPAR 2022 predicted transcription factor binding sites (TFBS) containing SNPs that are associated with JIA (Tier 1 or Tier 2 regions). Predicted transcription factor binding sites (highlighted in yellow) are oriented from 5'→3'. Sequence logos along the x-axis are produced from JASPAR 2024 experimental binding motifs supporting these TFBS predictions, and distributions of MGW and ProT within these experimental binding motifs are captured through percentiles (10th / 90th = light gray regions; 25th / 75th = medium gray regions; 50th = dark gray line). MGW and ProT predictions for the sequences containing the JIA risk allele (dashed orange line) and non-risk allele (solid blue line) are superimposed. All 46 SNP-TFBS combinations that were tested are presented. Only those labeled as Isoclines 2-6 are predicted to have allele-specific binding effects (**Table S19**).

**(a) Impact of rs2358995 on predicted CDX2 binding site (positive strand).**

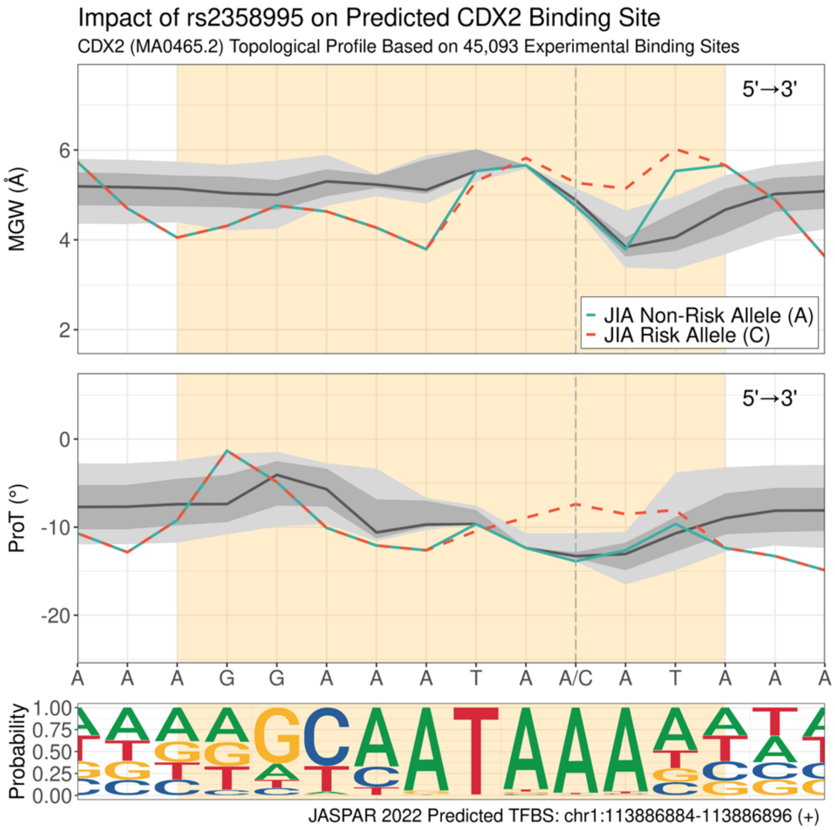

**(b) Impact of rs2488457 on predicted CEBPG binding site (negative strand).**

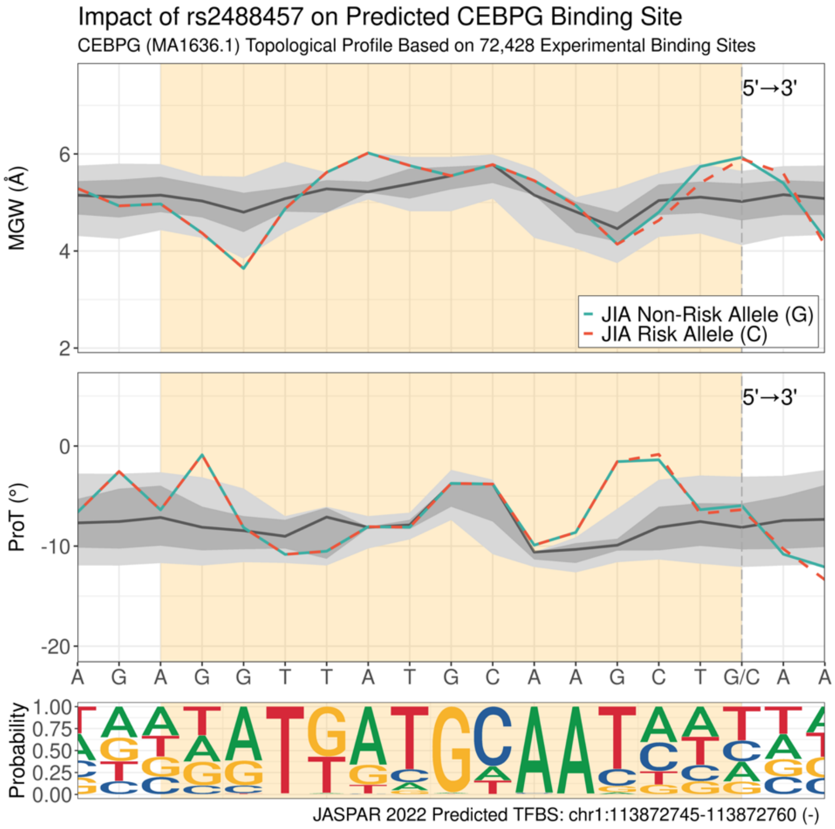

**(c) Impact of rs4147359 on predicted CREB1 binding site (positive strand).**

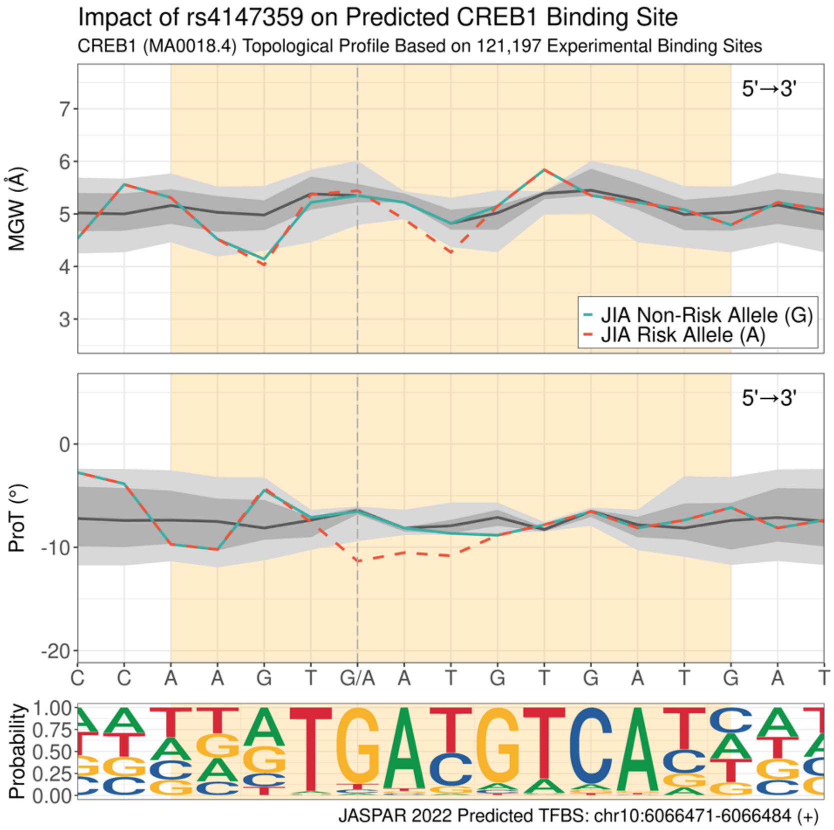

**(d) Impact of rs1274955 on predicted EBF1 binding site (negative strand).**

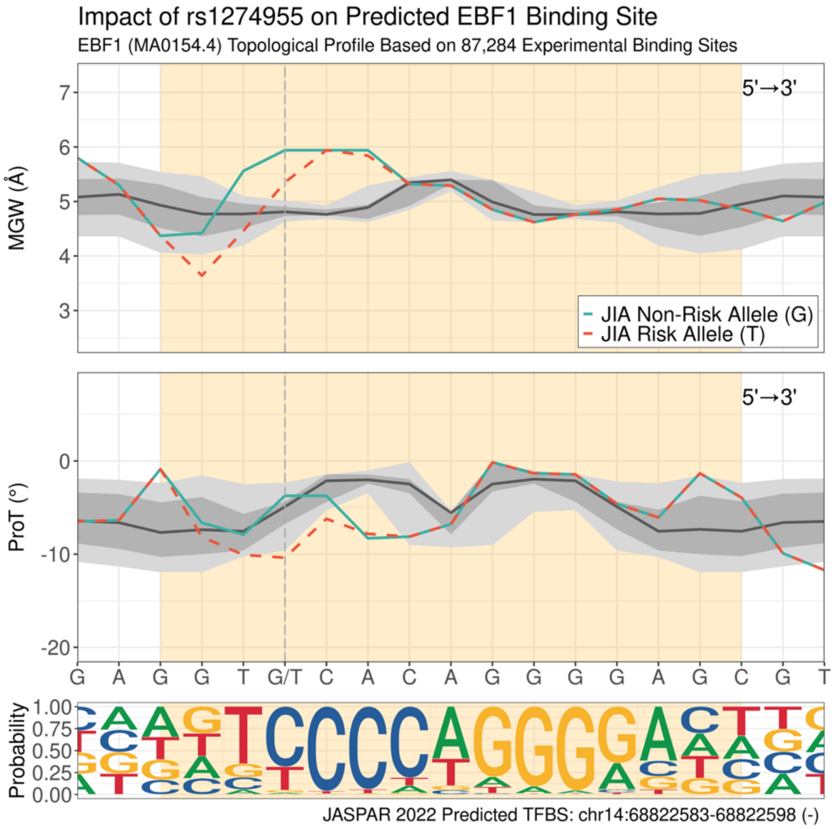

**(e) Impact of rs34437200 on predicted EBF1 binding site (negative strand).**

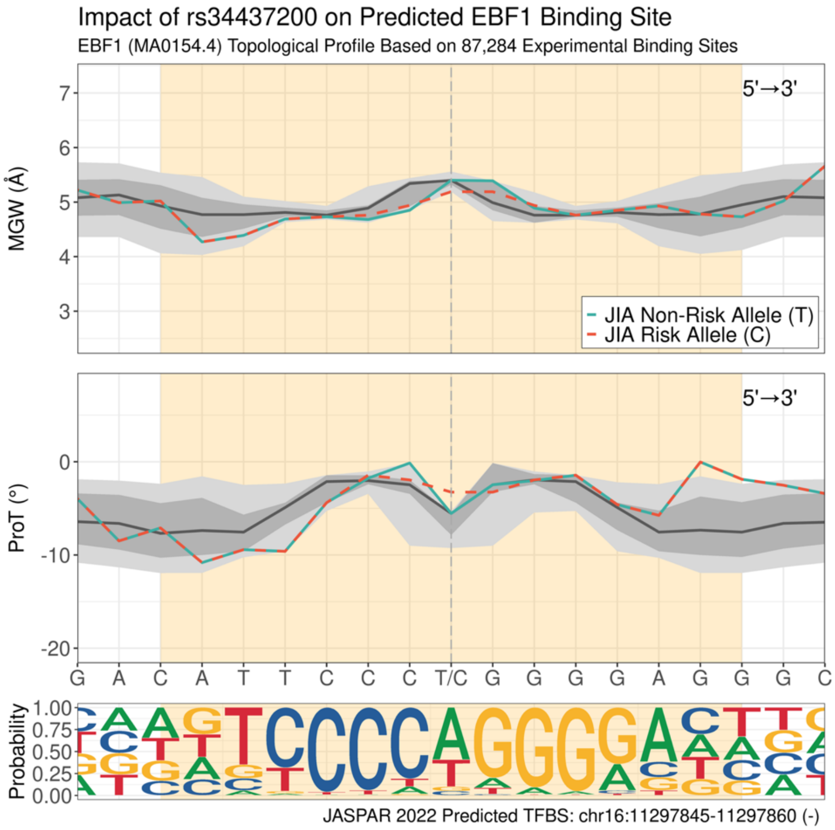

**(f) Impact of rs34437200 on predicted EBF1 binding site (positive strand).**

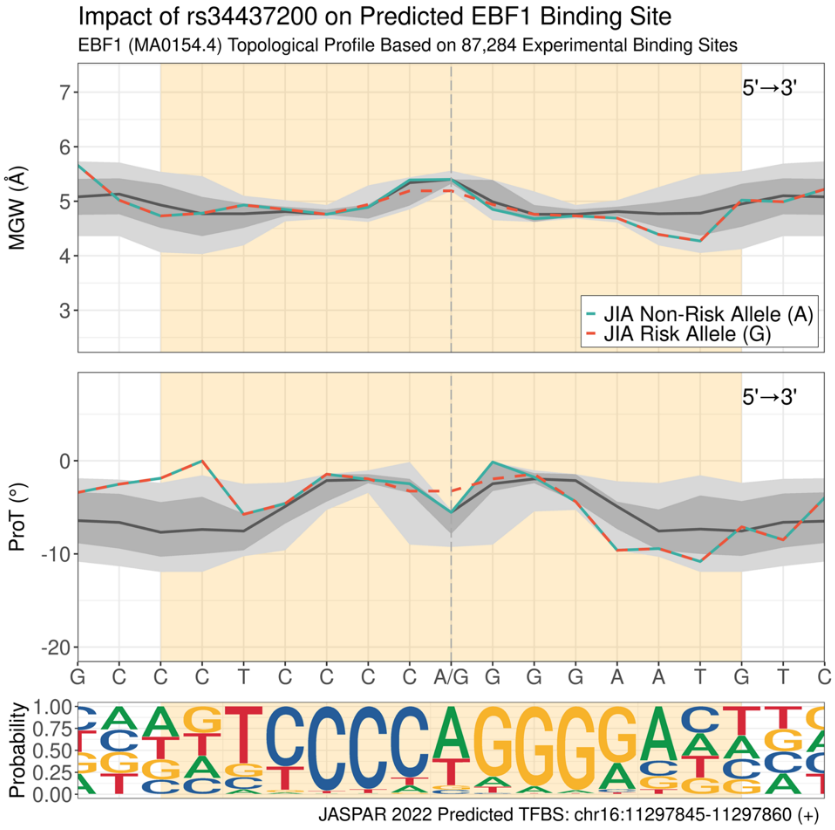

**(g) Impact of rs34437200 on predicted EBF3 binding site (negative strand).**

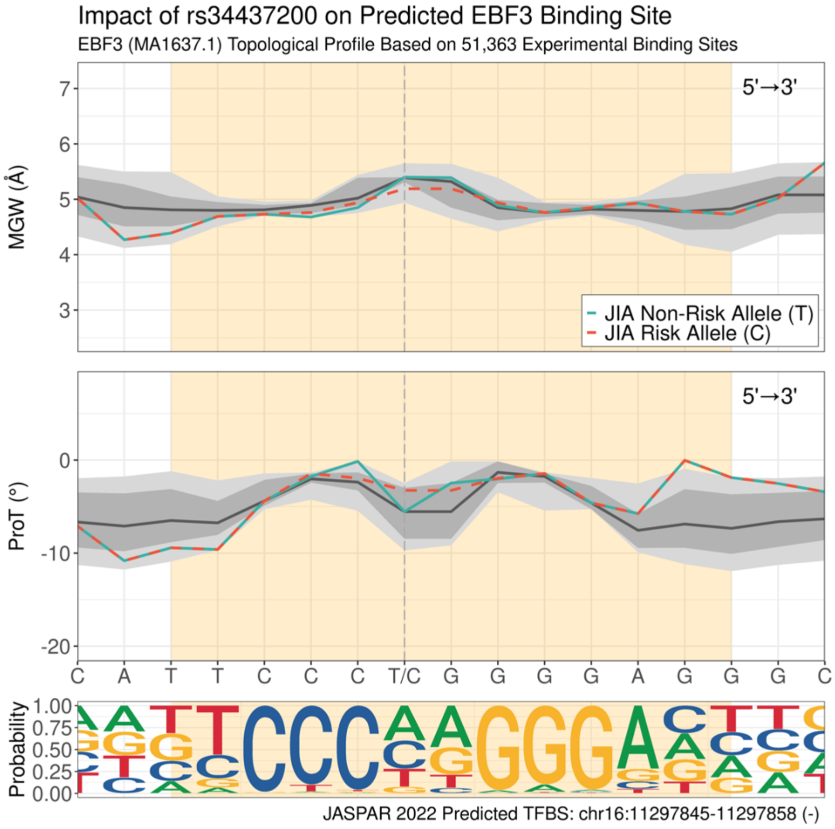

**(h) Impact of rs34437200 on predicted EBF3 binding site (positive strand).**

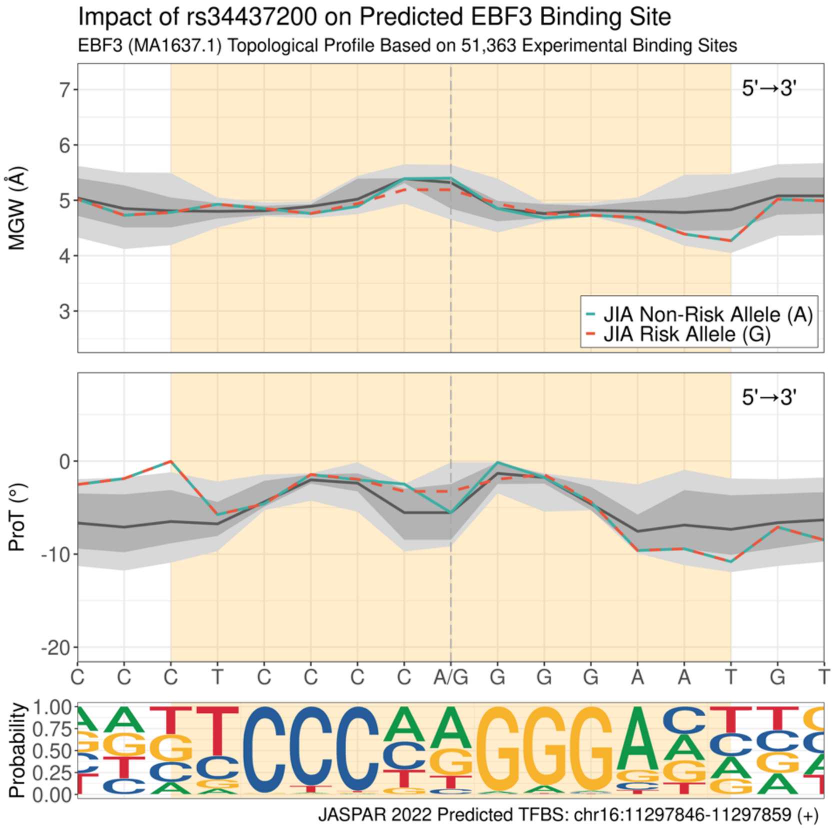

**(i) Impact of rs41295065 on predicted FOS binding site (negative strand).**

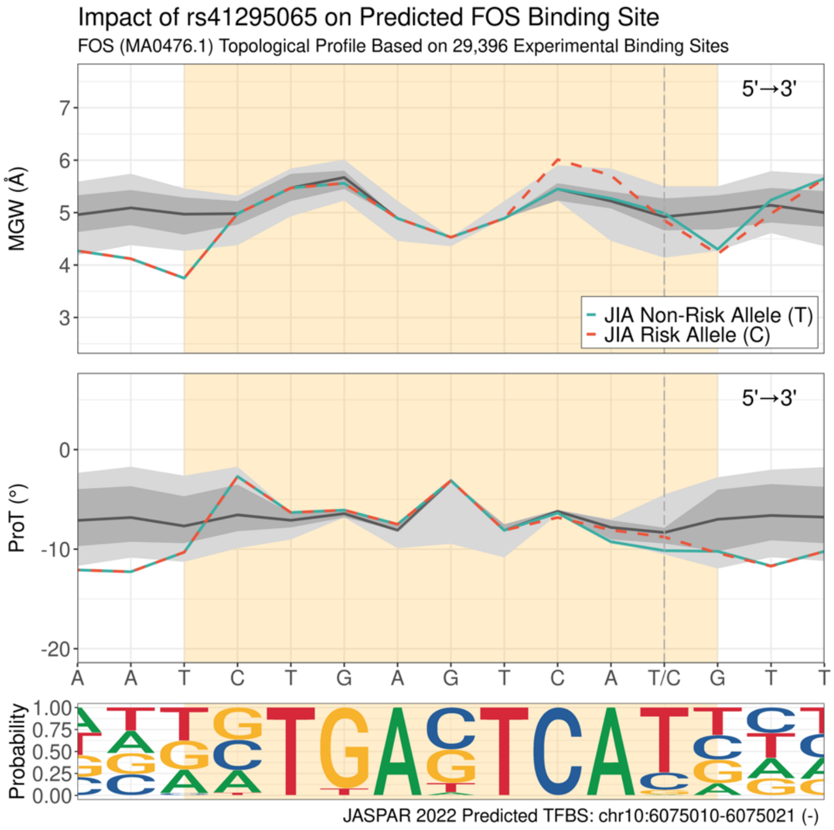

**(j) Impact of rs41295065 on predicted FOSL1 binding site (positive strand).**

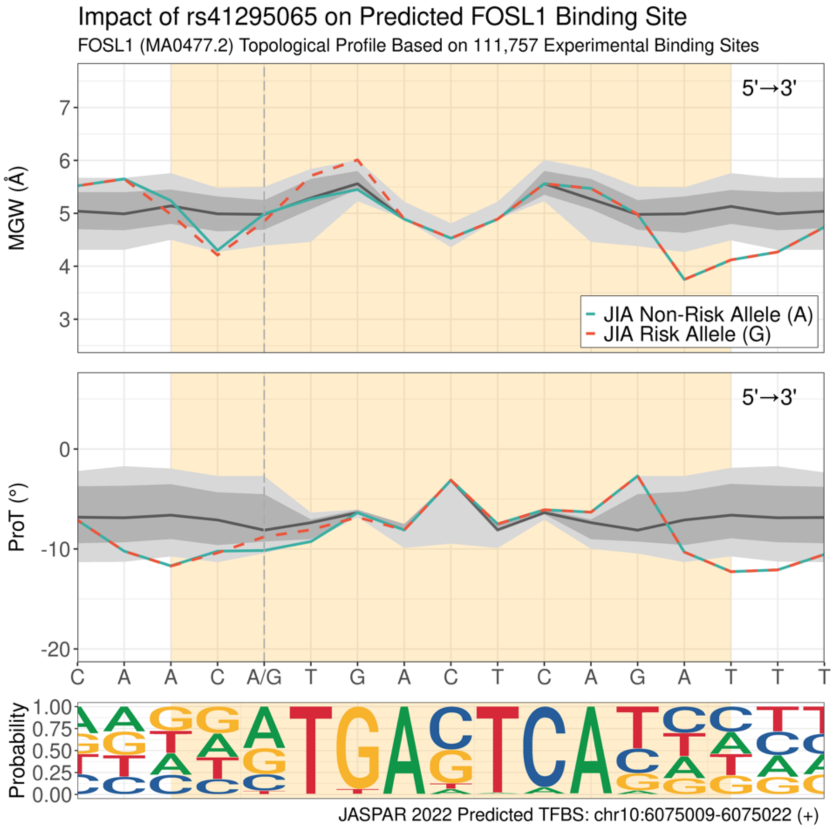

**(k) Impact of rs41295065 on predicted FOSL2 binding site (positive strand).**

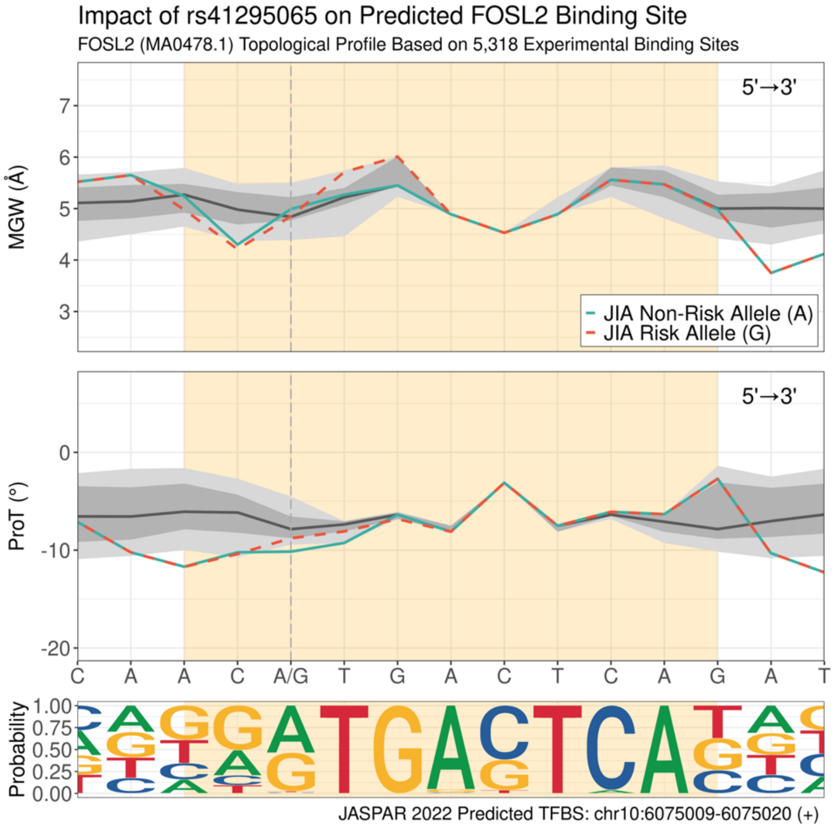

**(l) Impact of rs1217203 on predicted FOXA2 binding site (negative strand).**

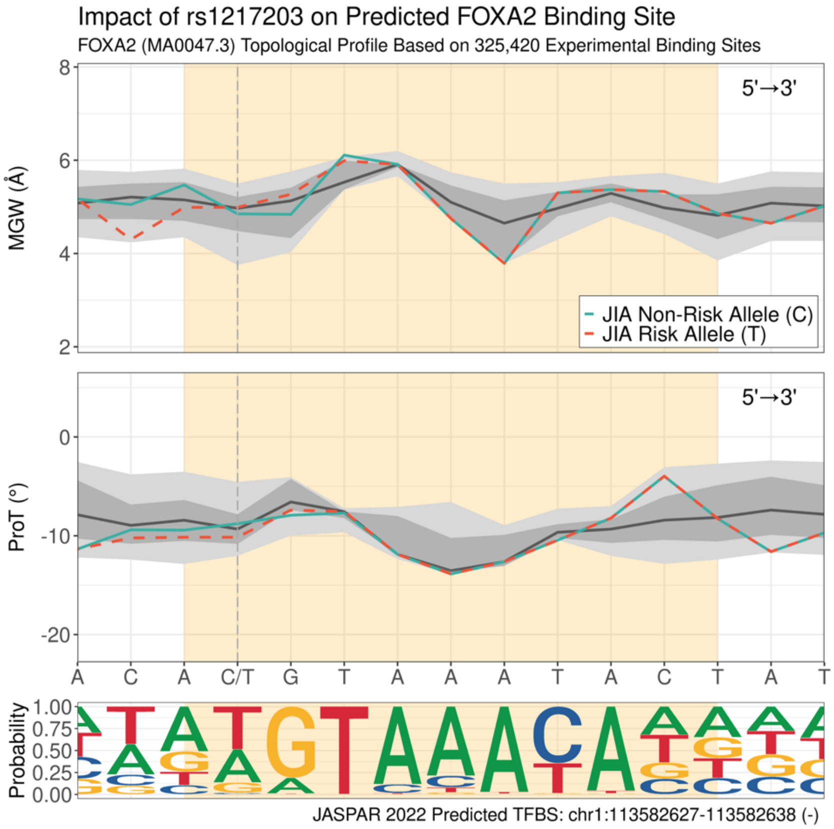

**(m) Impact of rs7234029 on predicted GATA1 binding site (negative strand).**

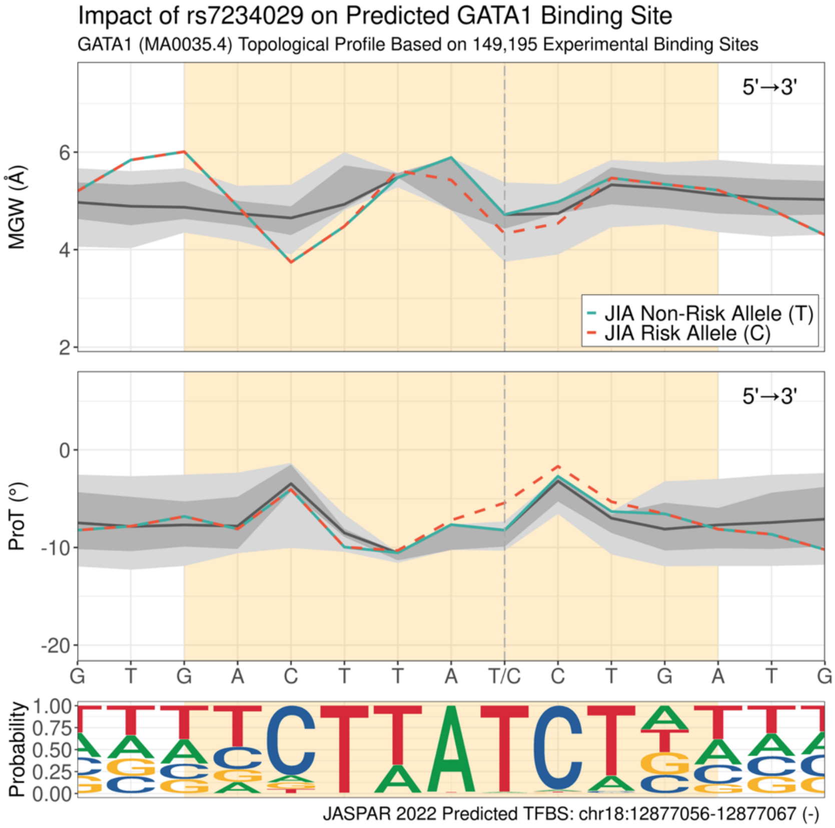

**(n) Impact of rs13032454 on predicted GATA2 binding site (negative strand).**

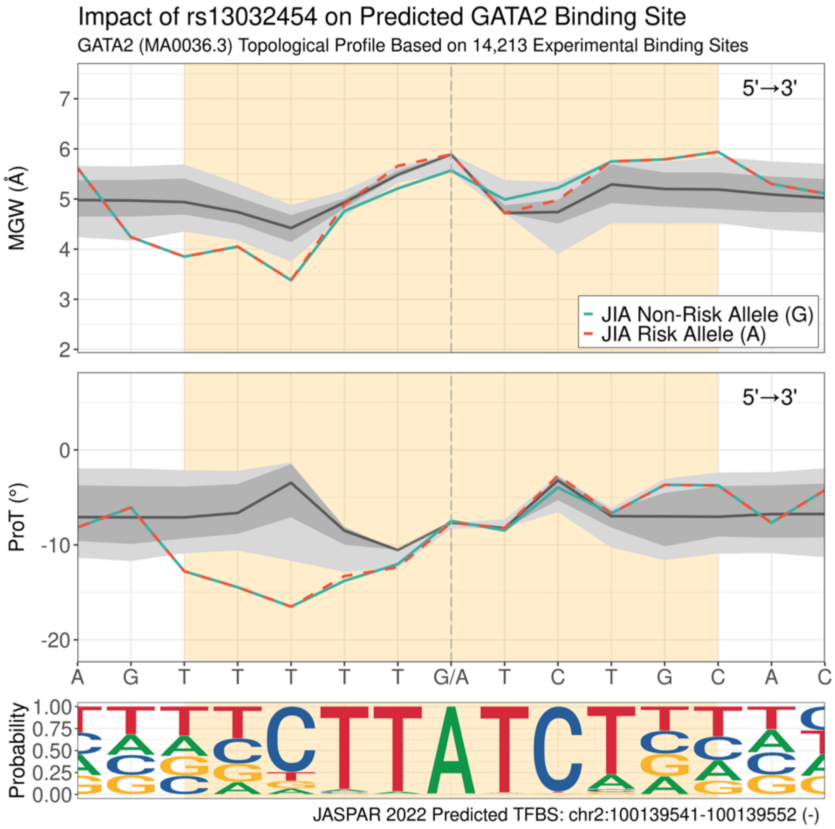

**(o) Impact of rs41295065 on predicted JUND binding site (positive strand).**

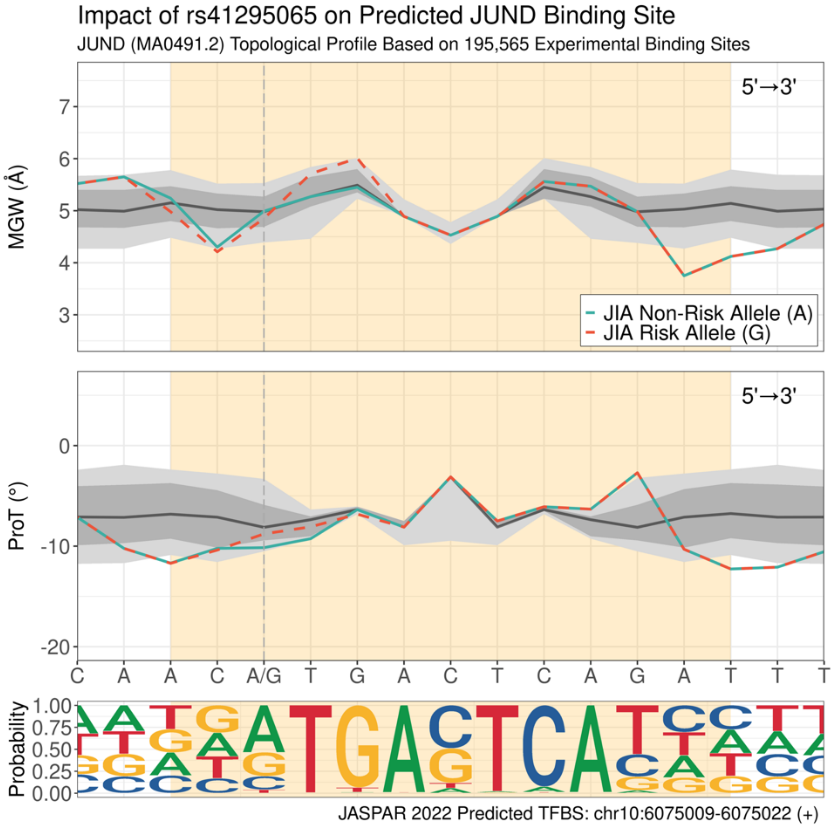

**(p) Impact of rs2569693 on predicted KLF4 binding site (positive strand).**

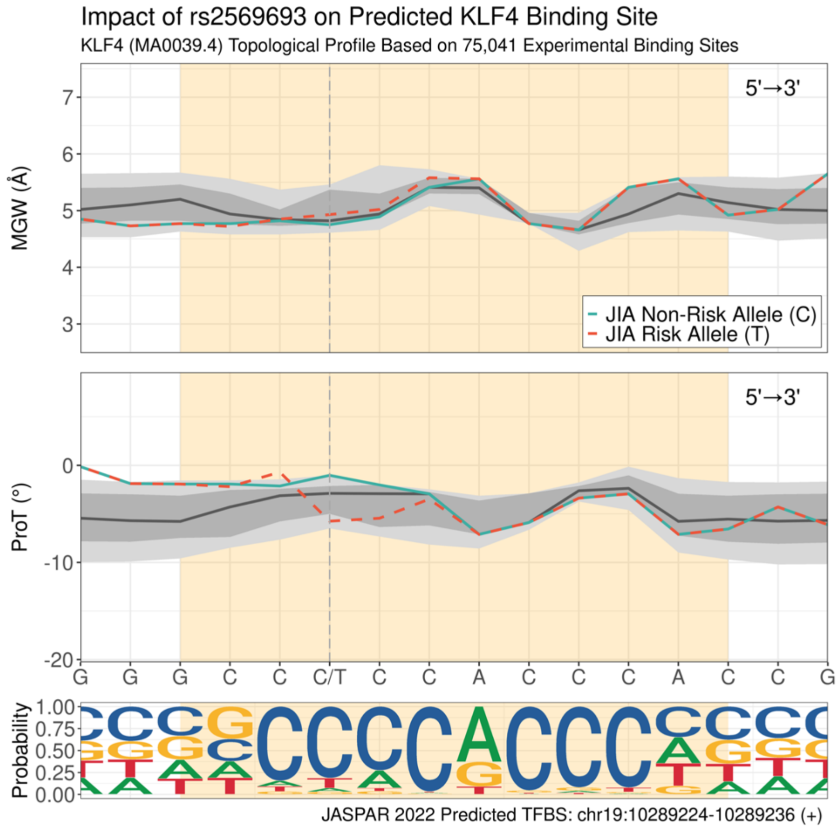

**(q) Impact of rs2569693 on predicted KLF9 binding site (positive strand).**

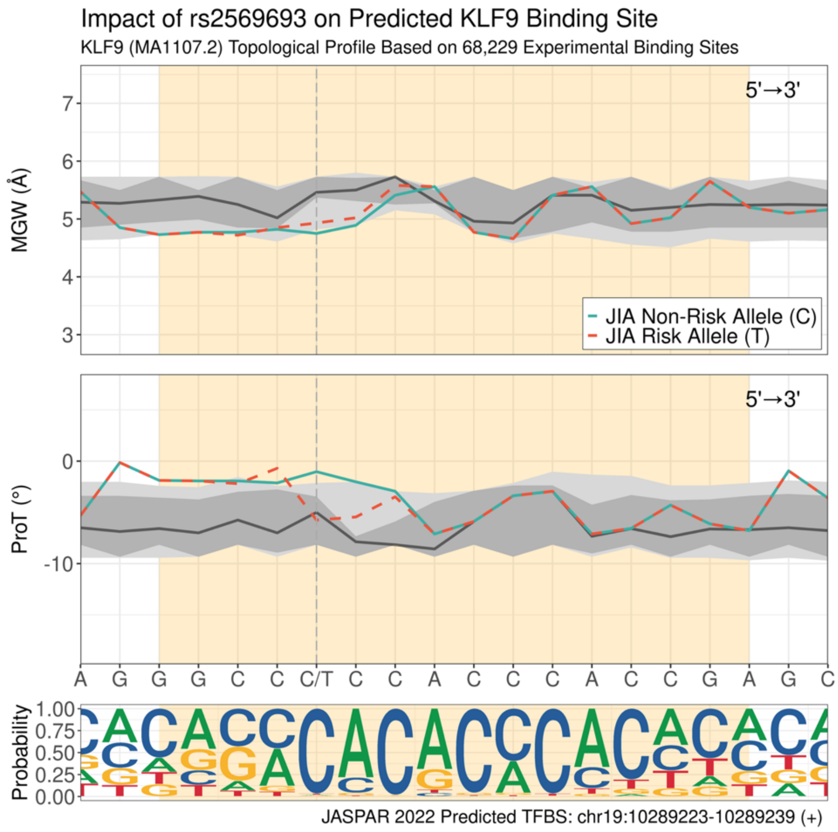

**(r) Impact of rs9483788 on predicted MEF2A binding site (positive strand).**

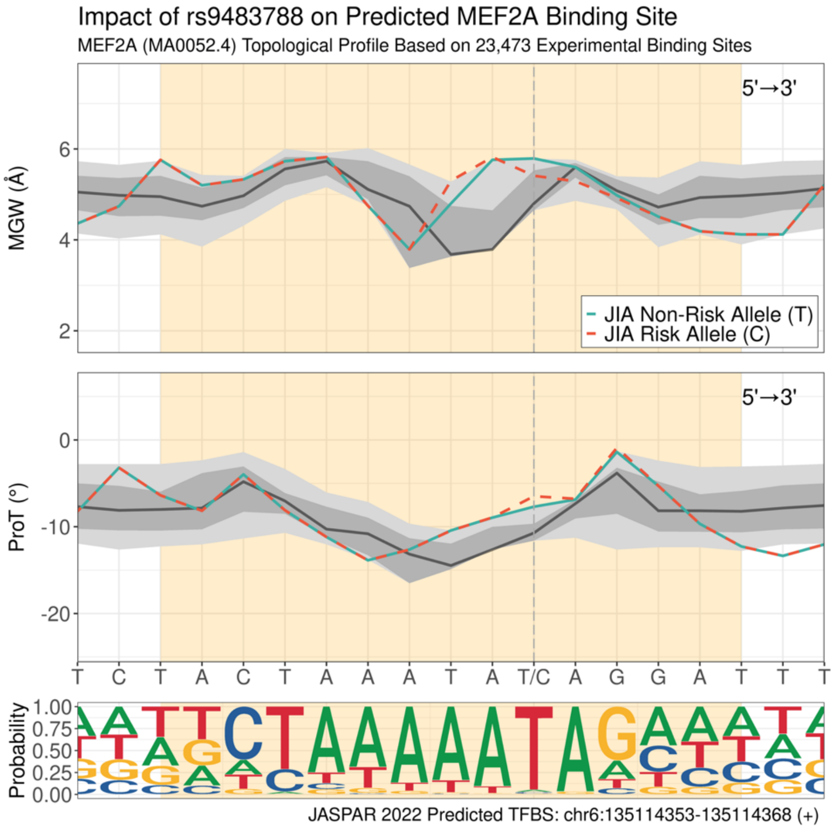

**(s) Impact of rs61839660 on predicted MEF2A binding site (negative strand).**

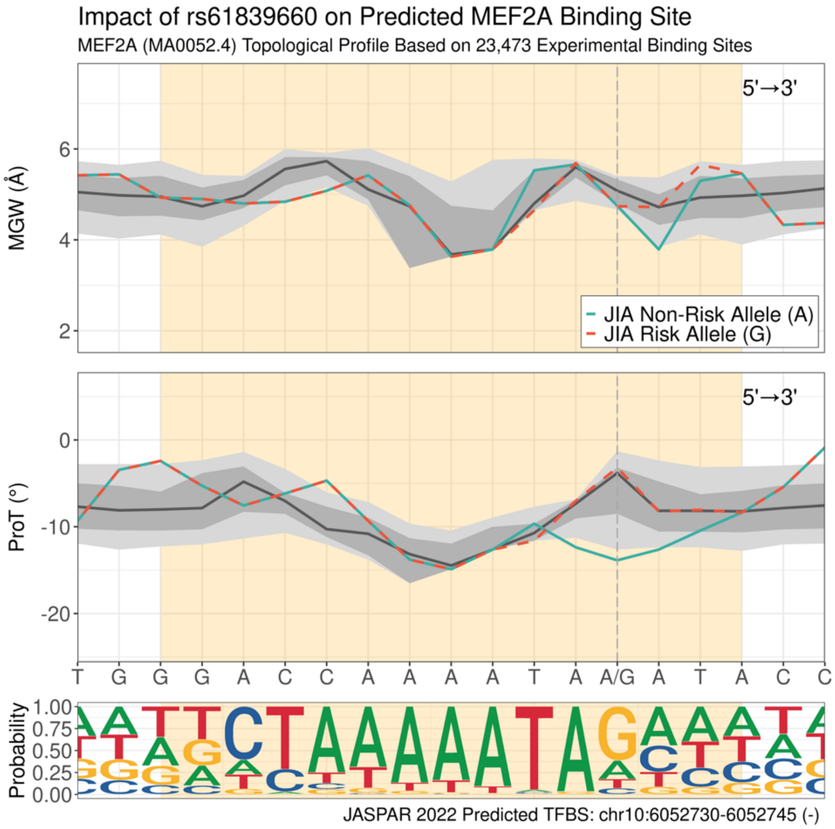

**(t) Impact of rs9483788 on predicted MEF2C binding site (positive strand).**

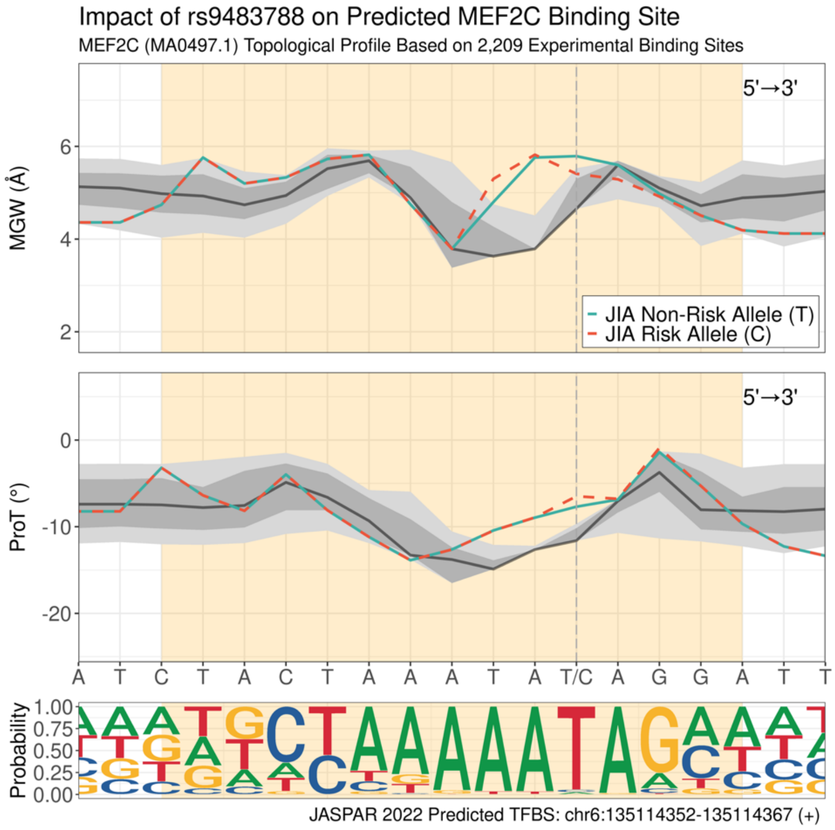

**(u) Impact of rs61839660 on predicted MEF2C binding site (negative strand).**

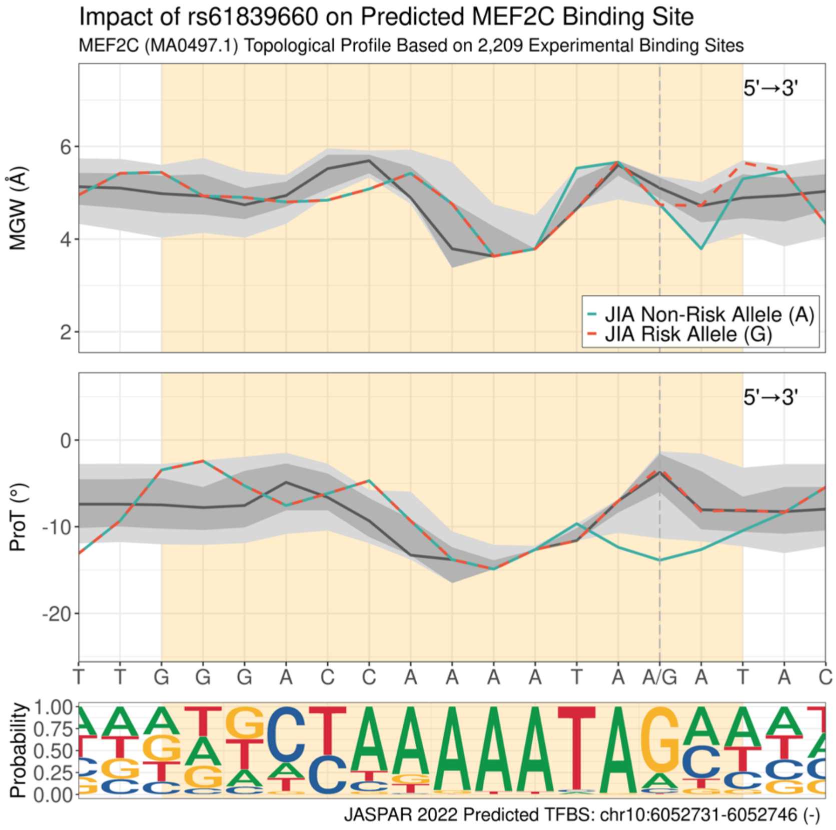

**(v) Impact of rs4796146 on predicted MITF binding site (negative strand).**

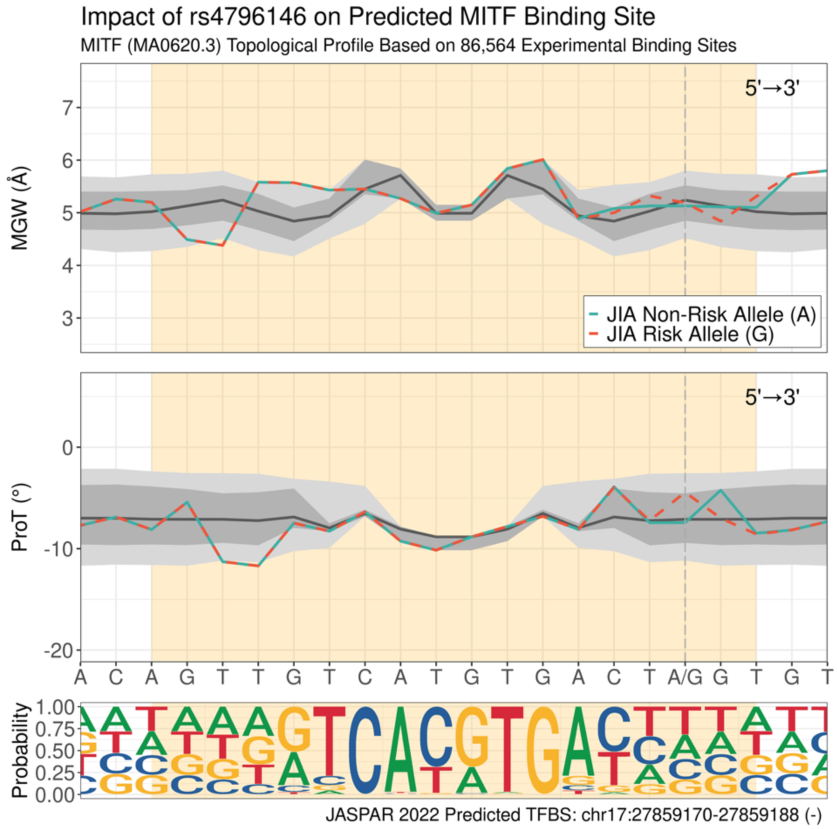

**w) Impact of rs4796146 on predicted MITF binding site (positive strand).**

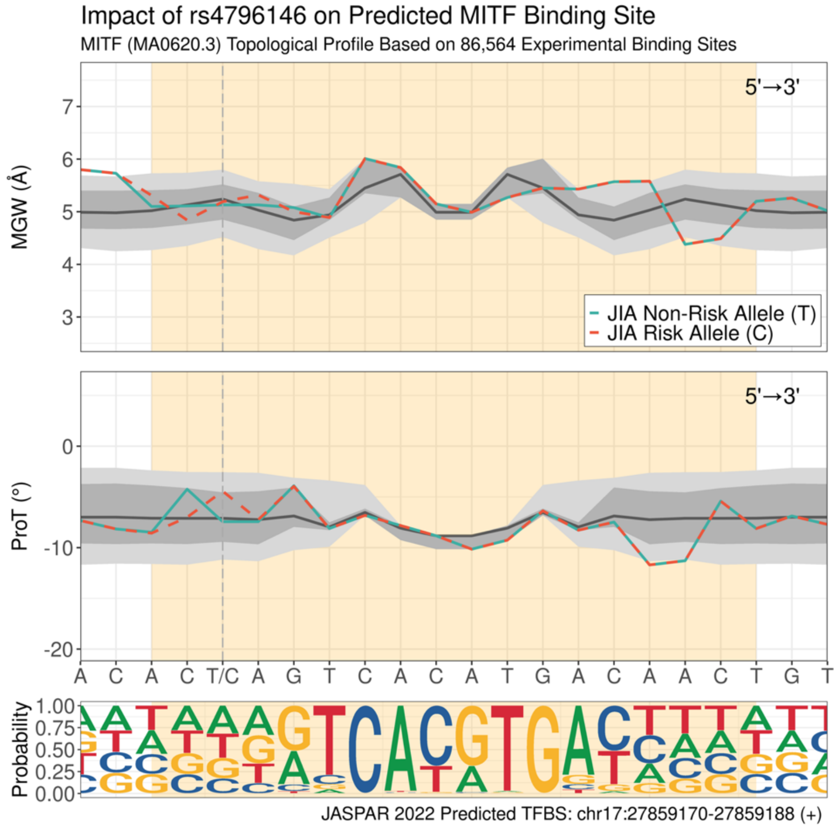

**(x) Impact of rs75867630 on predicted MYOD1 binding site (positive strand).**

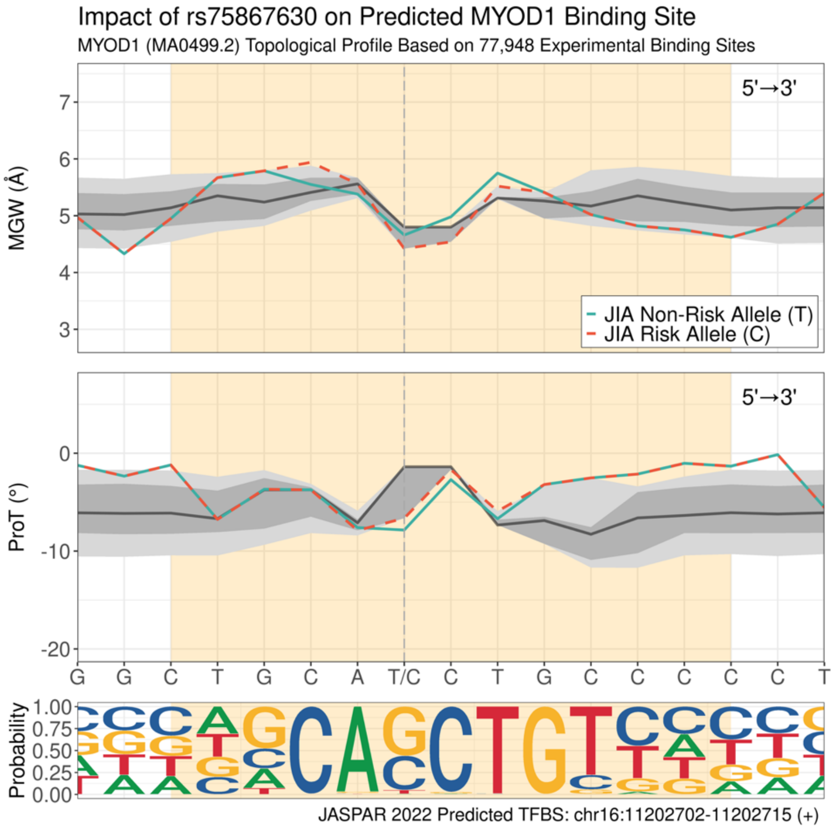

**(y) Impact of rs79699236 on predicted MZF1 binding site (positive strand).**

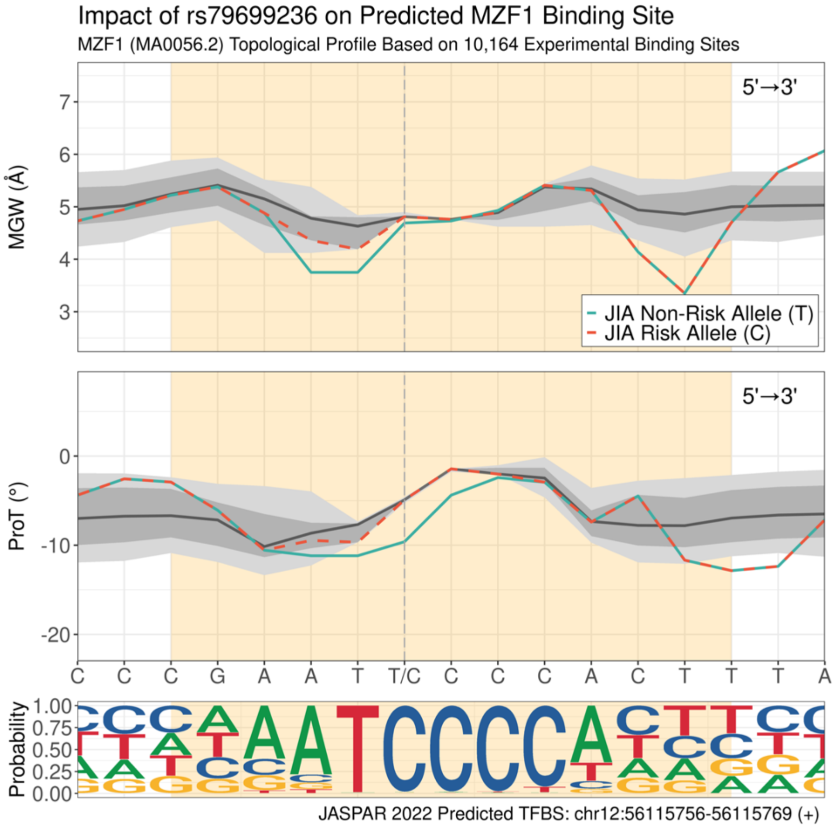

**(z) Impact of rs2188962 on predicted NR2C2 binding site (negative strand).**

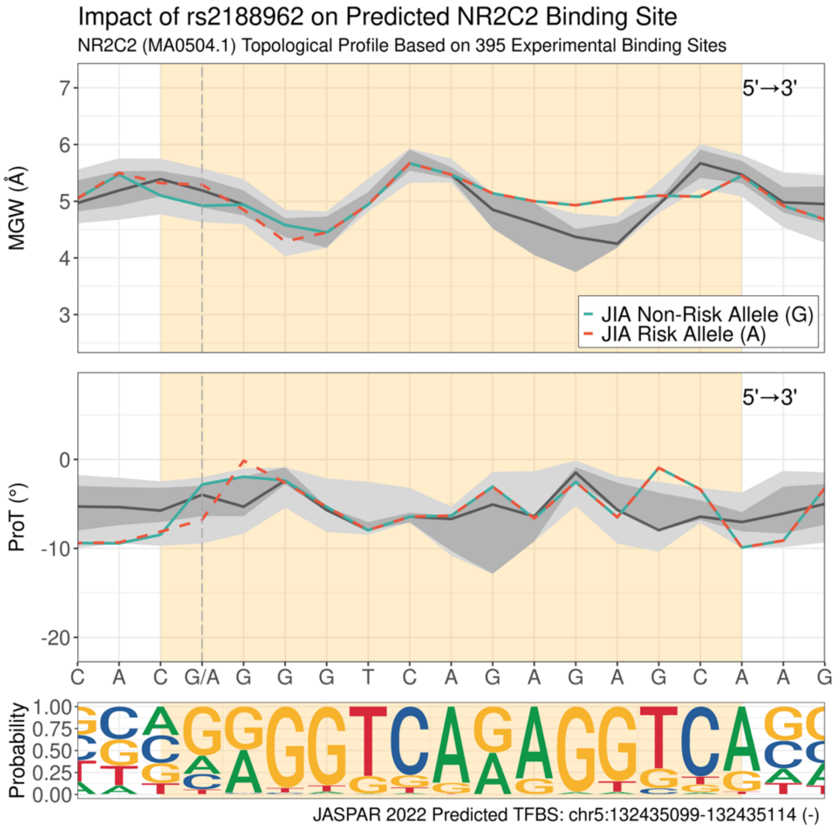

**(aa) Impact of rs113010081 on predicted ONECUT1 binding site (negative strand).**

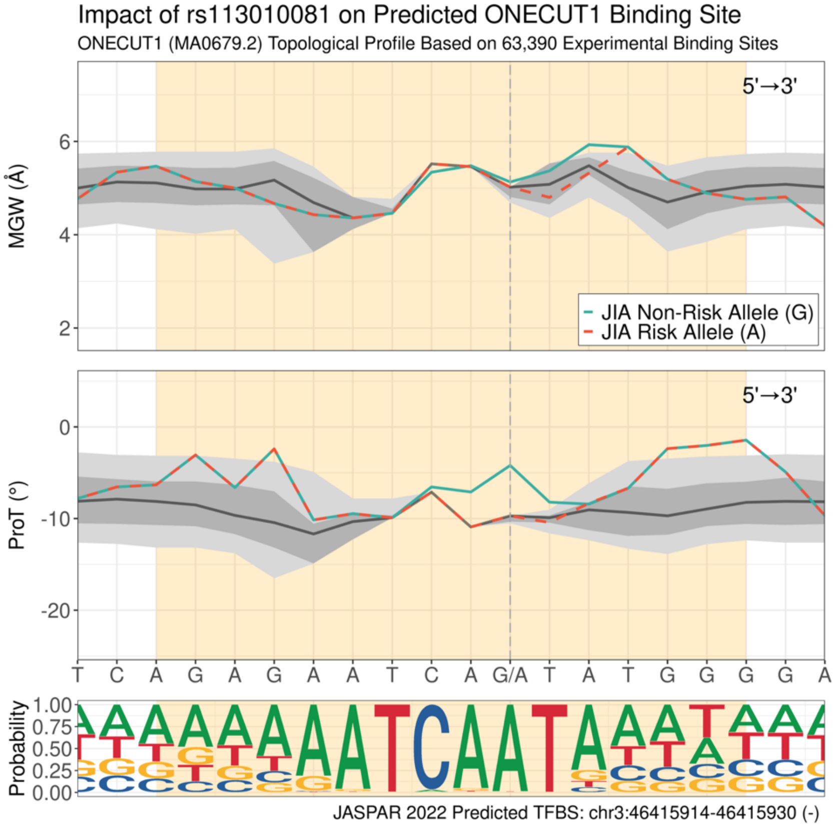

**(ab) Impact of rs34799913 on predicted PHOX2B binding site (negative strand).**

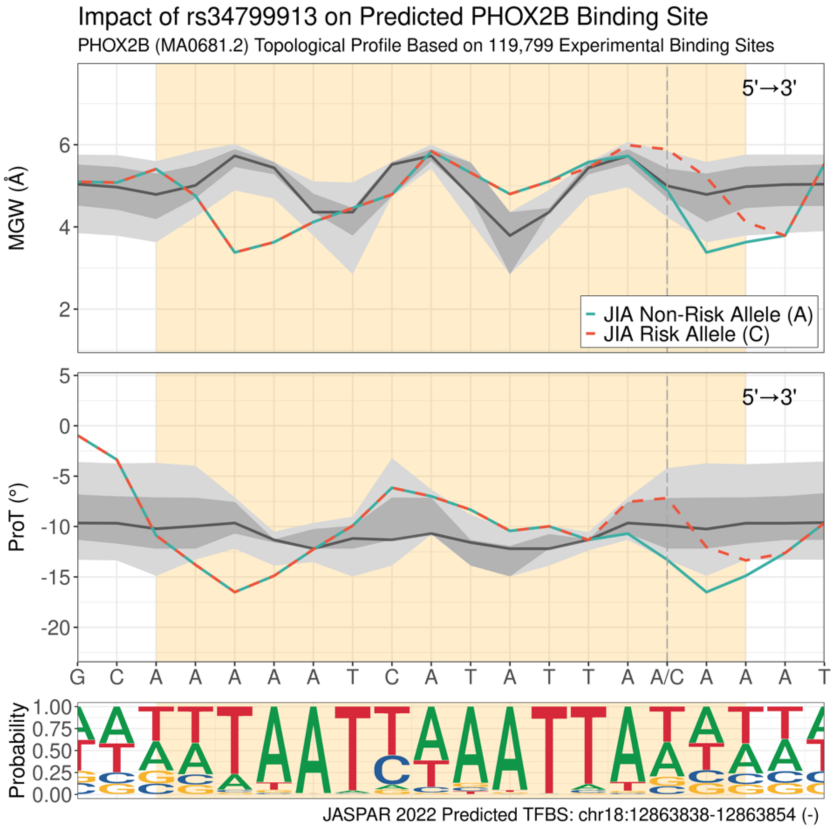

**(ac) Impact of rs10745339 on predicted POU2F3 binding site (negative strand).**

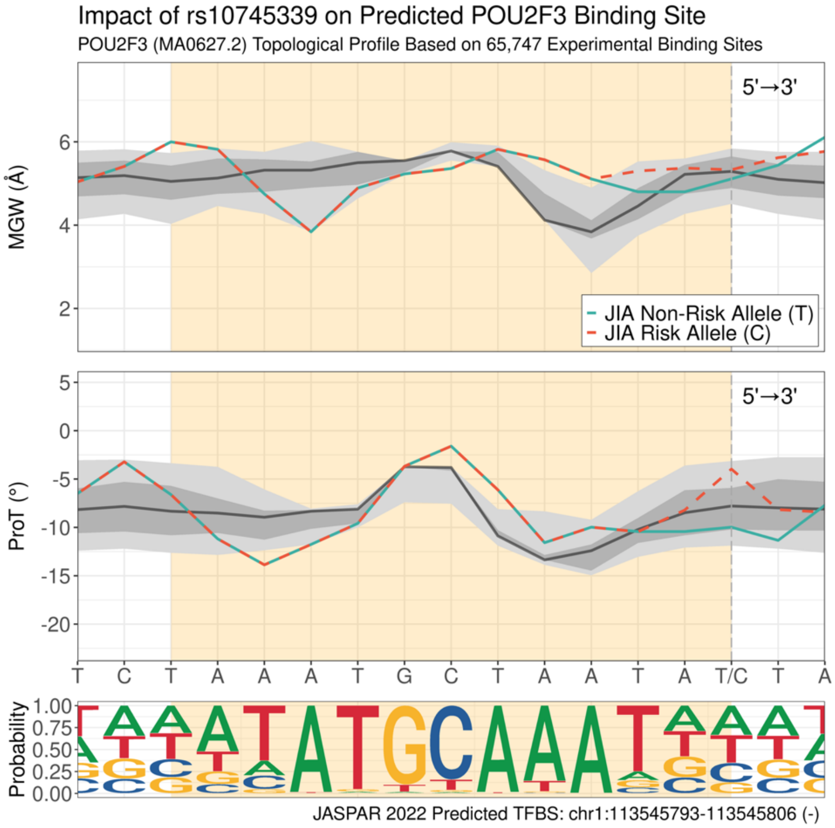

**(ad) Impact of rs2271893 on predicted SCRT1 binding site (negative strand).**

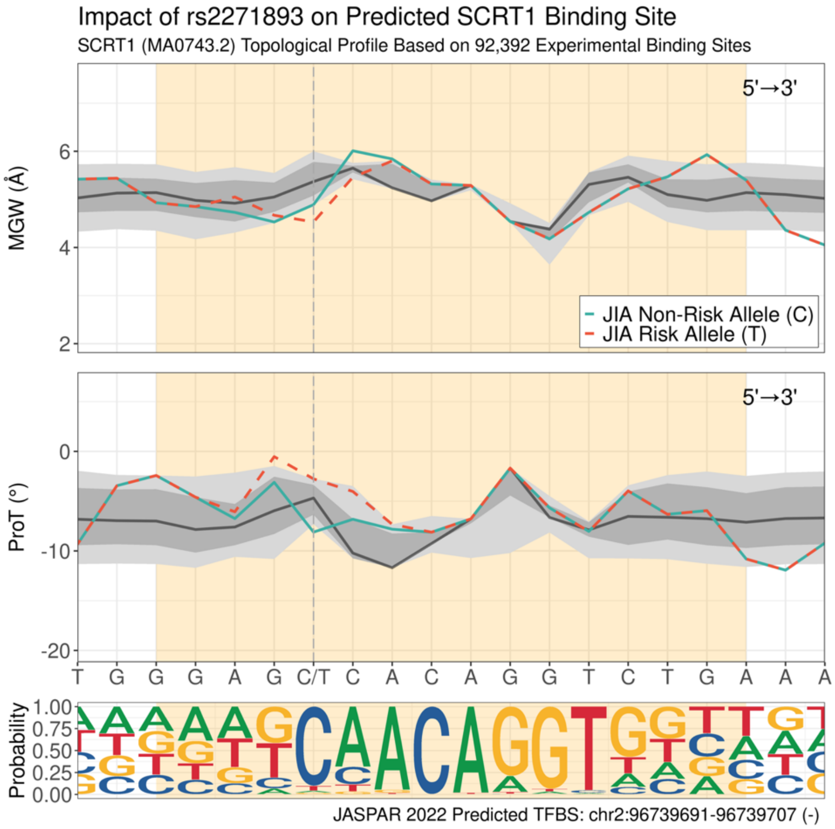

**(ae) Impact of rs2309837 on predicted SCRT1 binding site (negative strand).**

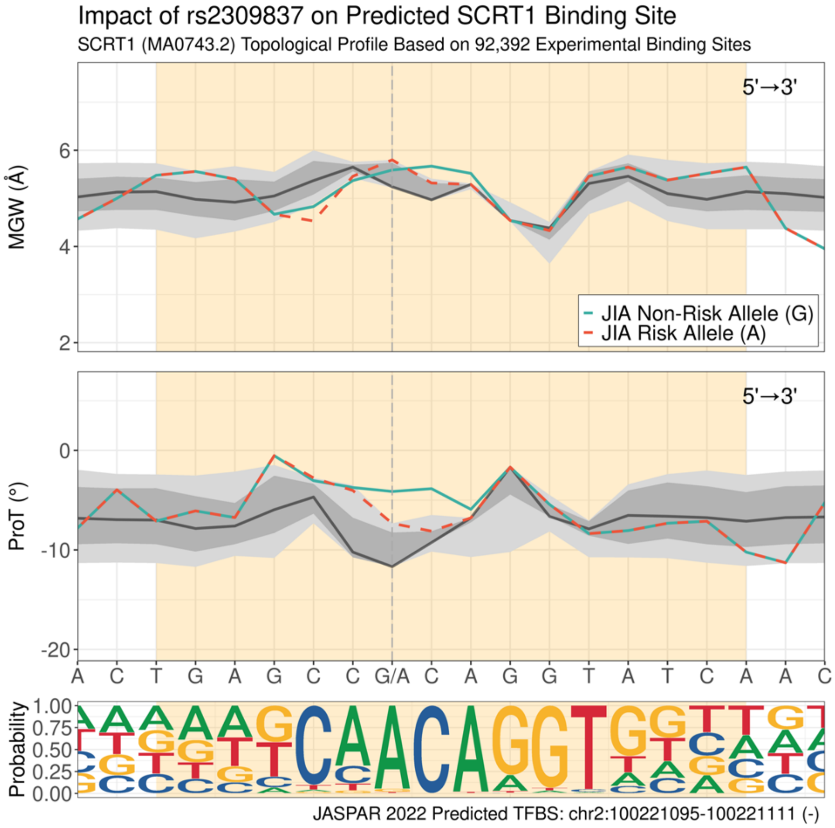

**(af) Impact of rs412861 on predicted SIX2 binding site (negative strand).**

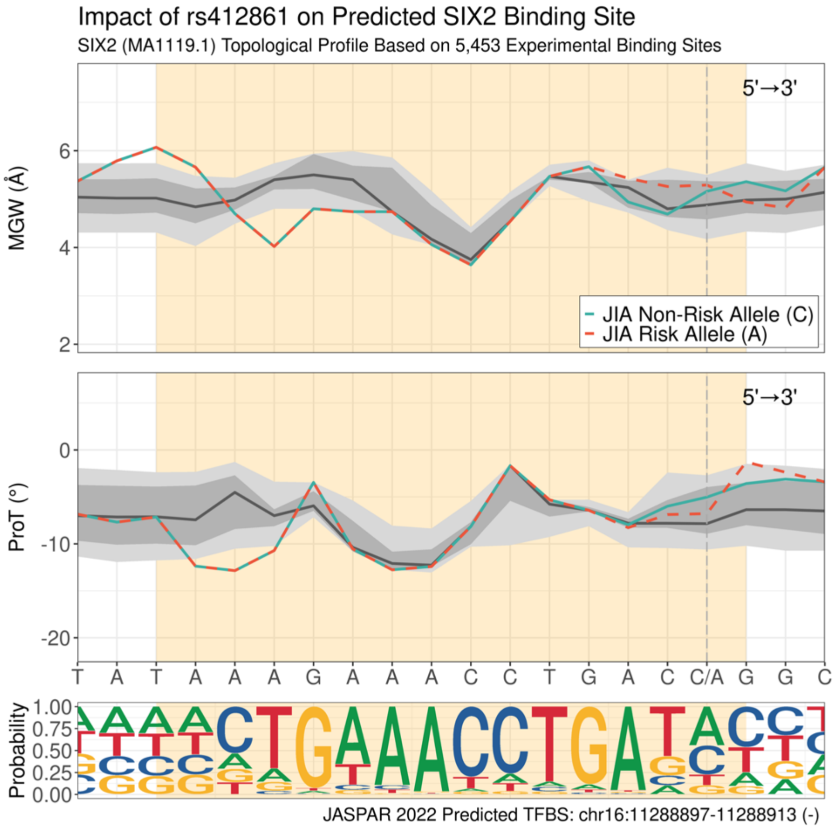

**(ag) Impact of rs75867630 on predicted SNAI2 binding site (positive strand).**

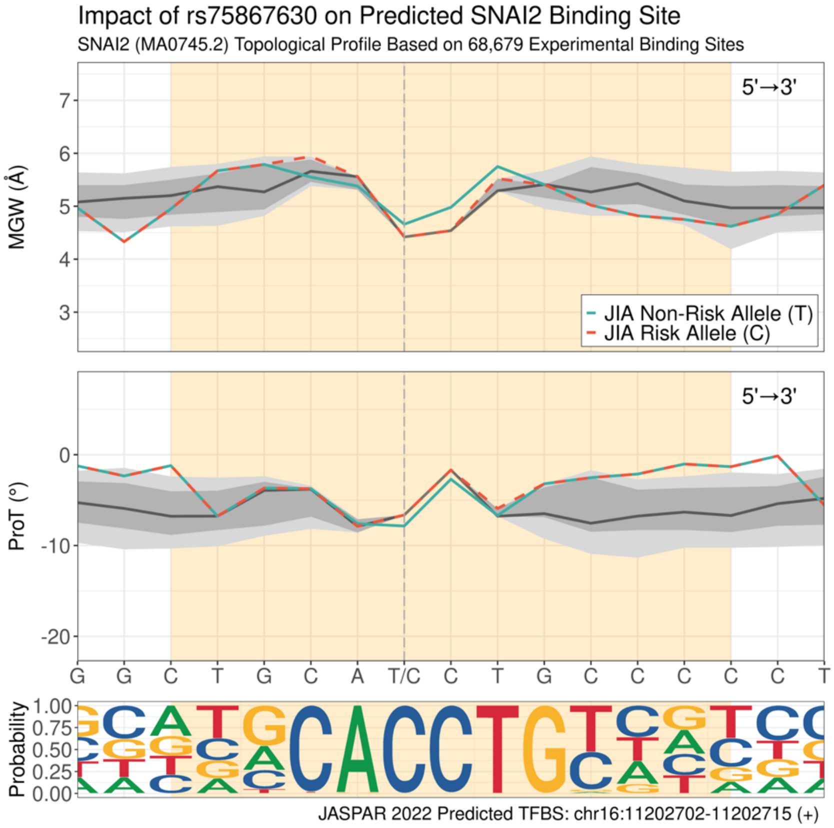

**(ah) Impact of rs10185510 on predicted SOX2 binding site (positive strand).**

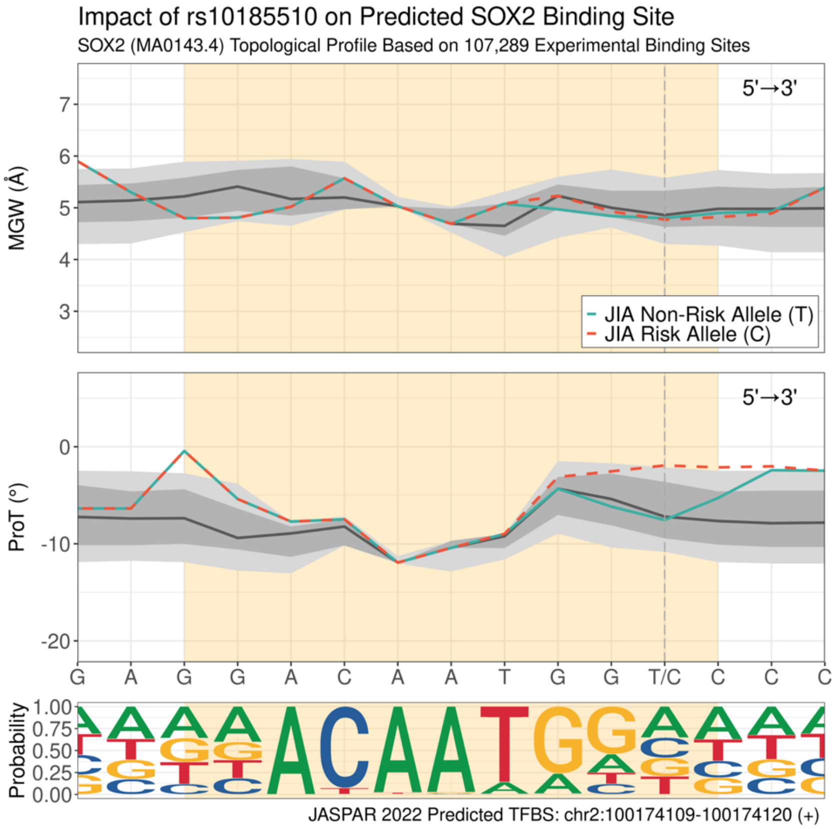

**(ai) Impact of rs13003982 on predicted SPIB binding site (negative strand).**

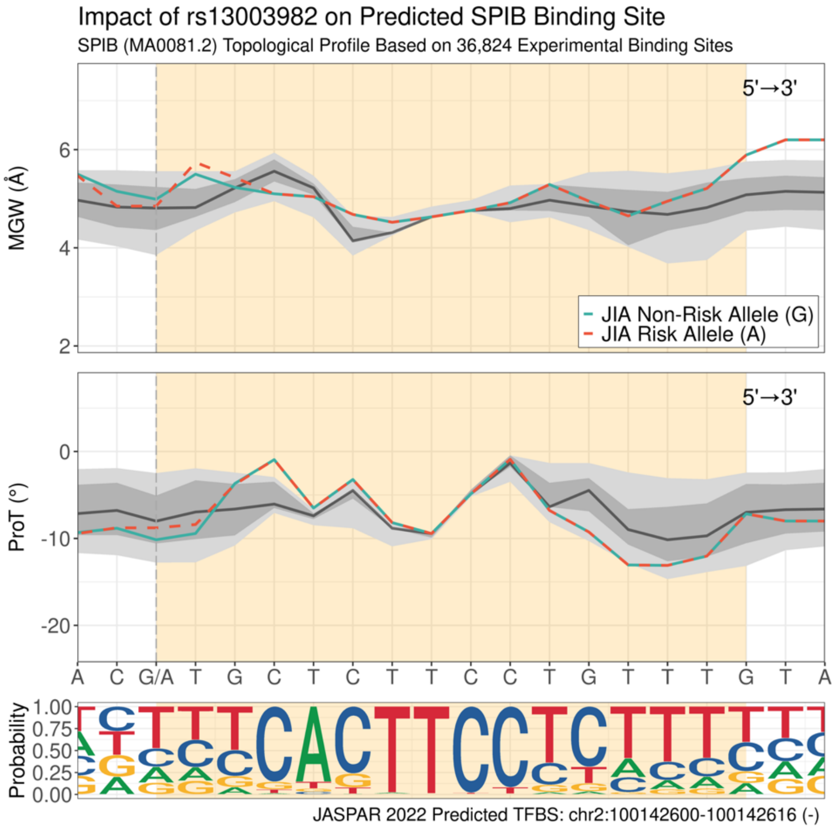

**(aj) Impact of rs13132308 on predicted SPIB binding site (positive strand).**

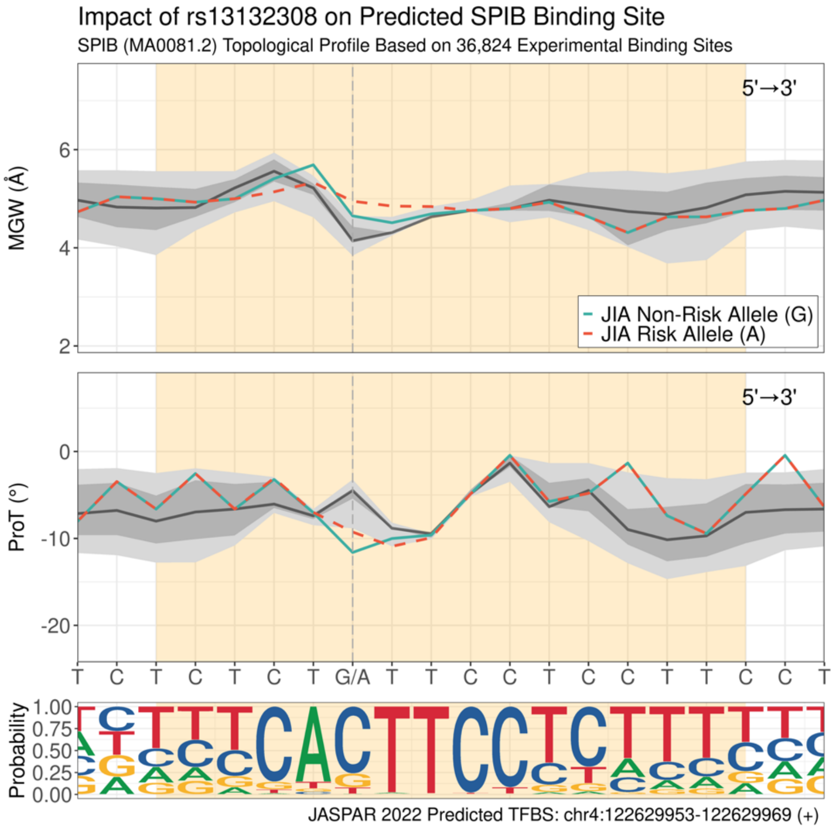

**(ak) Impact of rs62097857 on predicted SPIB binding site (negative strand).**

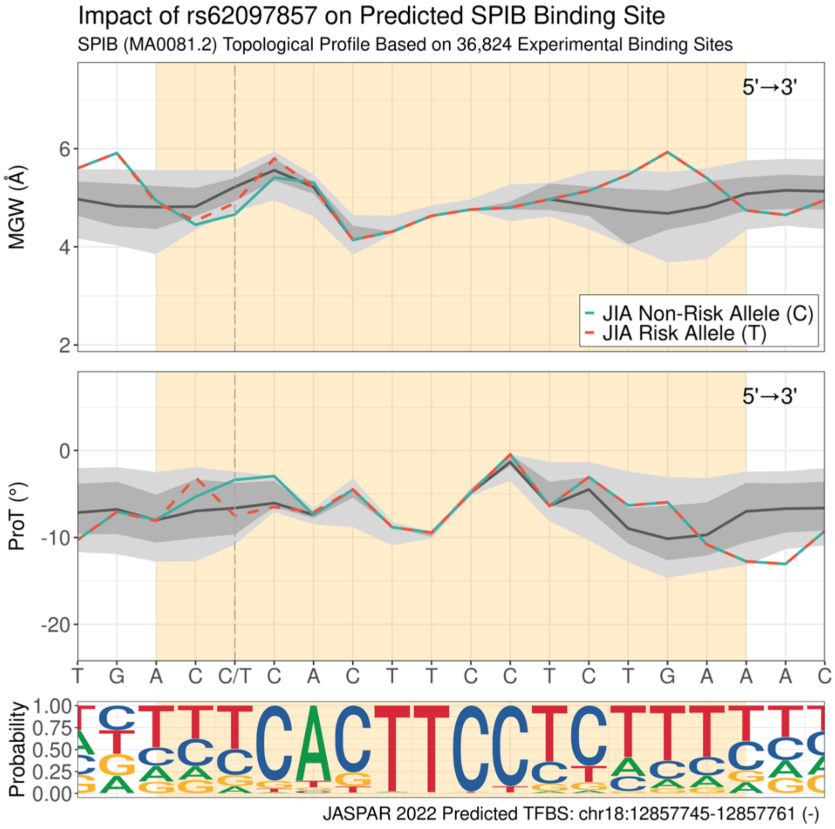

**(al) Impact of rs6807532 on predicted STAT3 binding site (negative strand).**

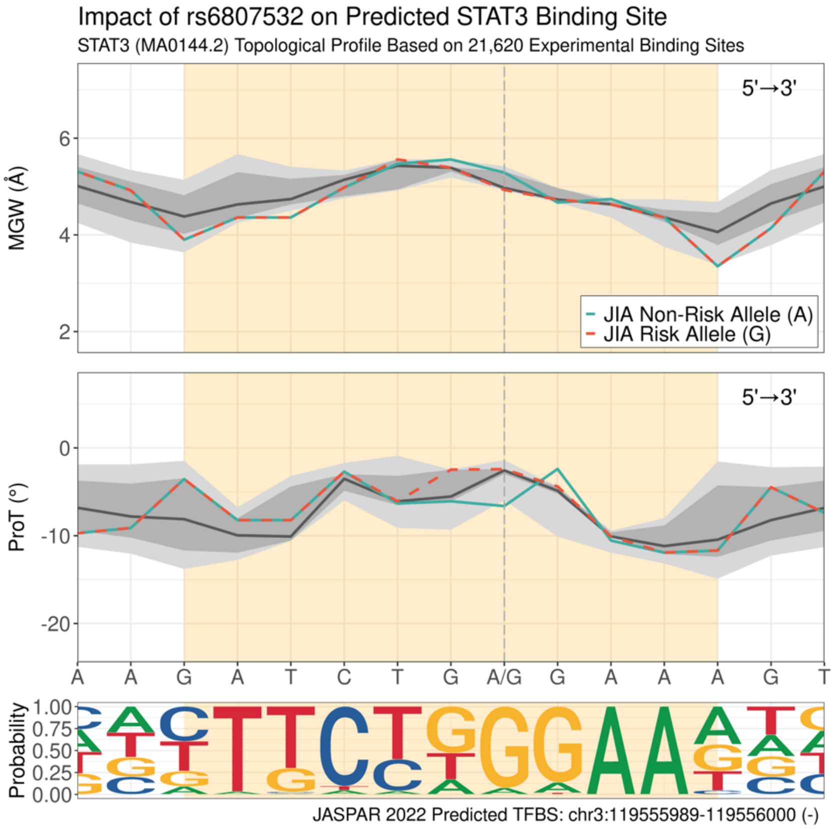

**(am) Impact of rs12484550 on predicted TCF3 binding site (negative strand).**

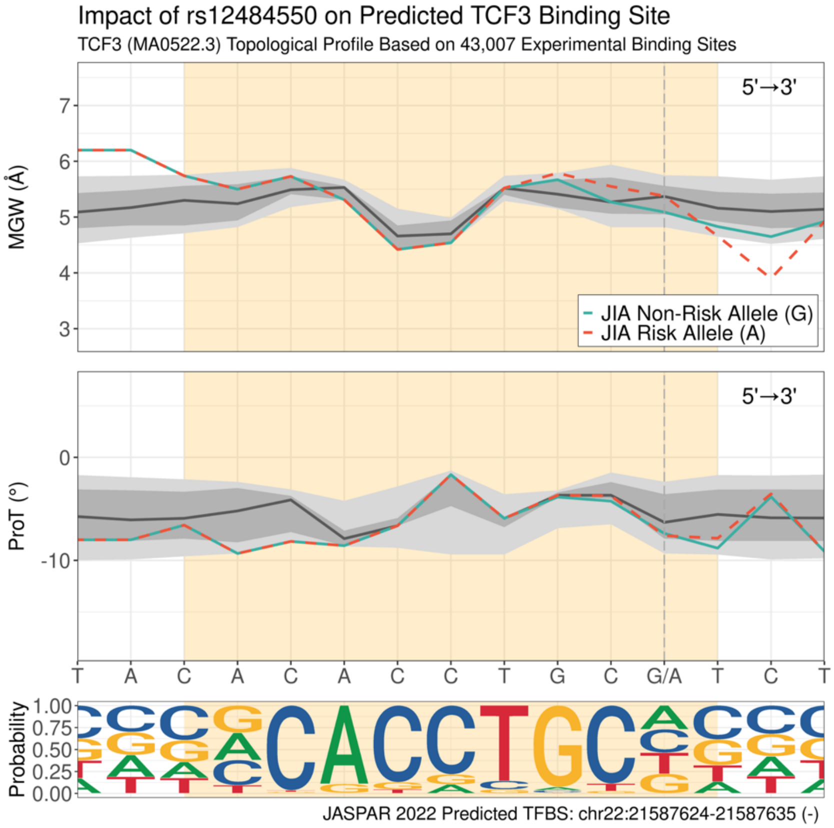

**(an) Impact of rs75867630 on predicted TCF3 binding site (positive strand).**

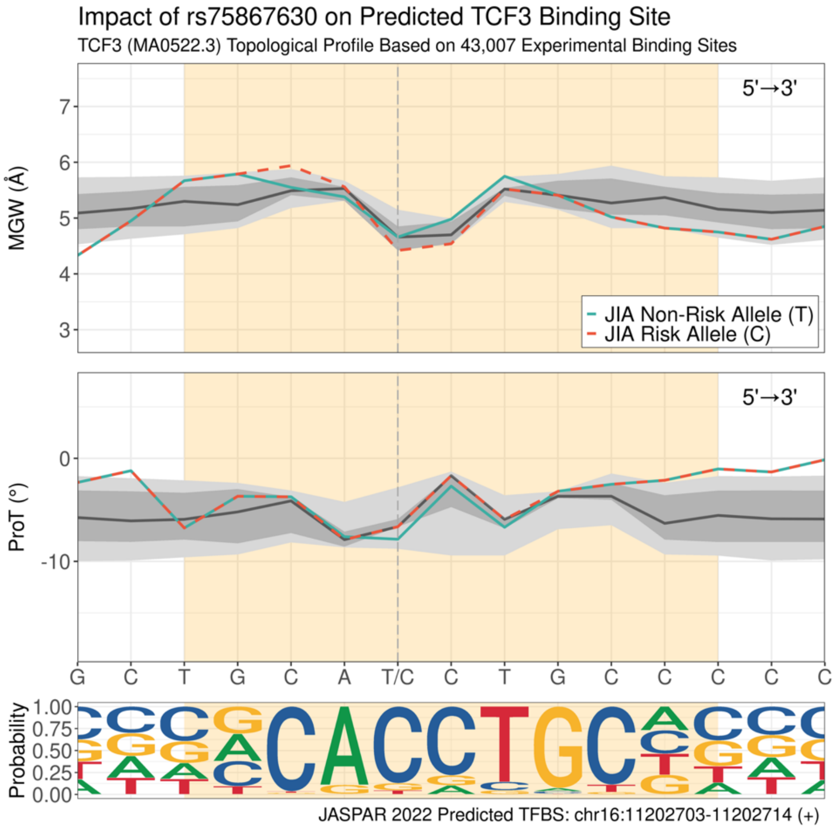

**(ao) Impact of rs12484550 on predicted TCF4 binding site (negative strand).**

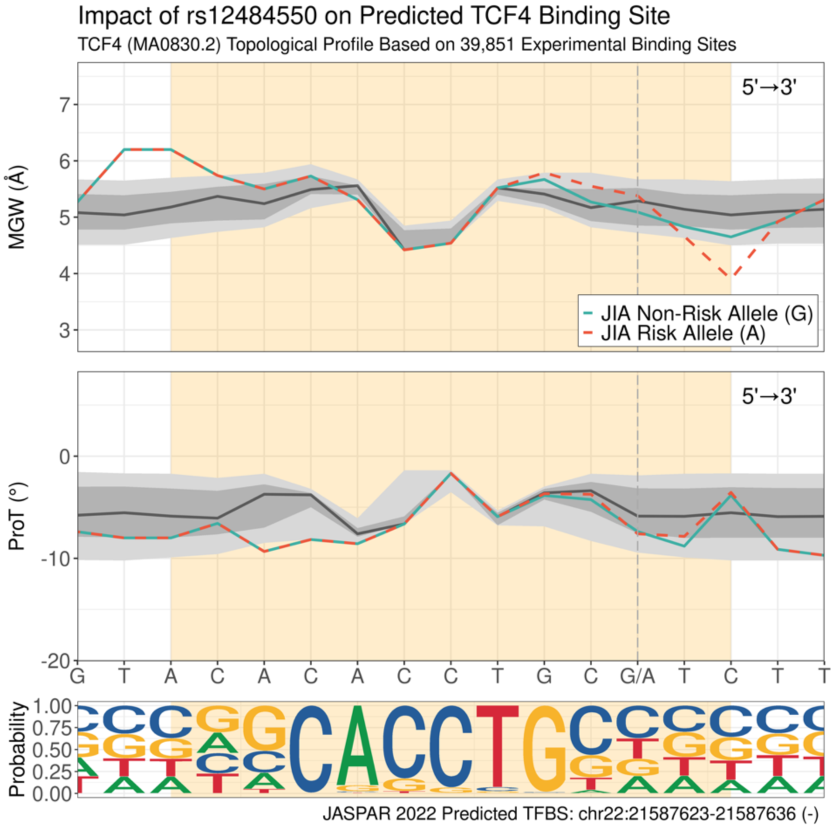

**(ap) Impact of rs75867630 on predicted TCF12 binding site (positive strand).**

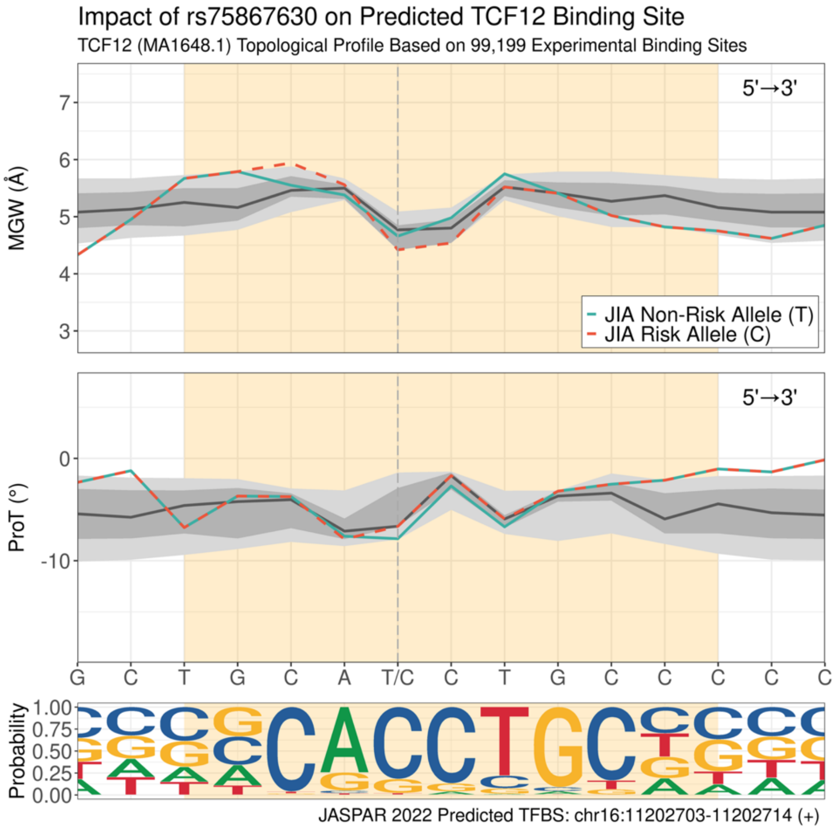

**(aq) Impact of rs11938795 on predicted TEAD4 binding site (positive strand).**

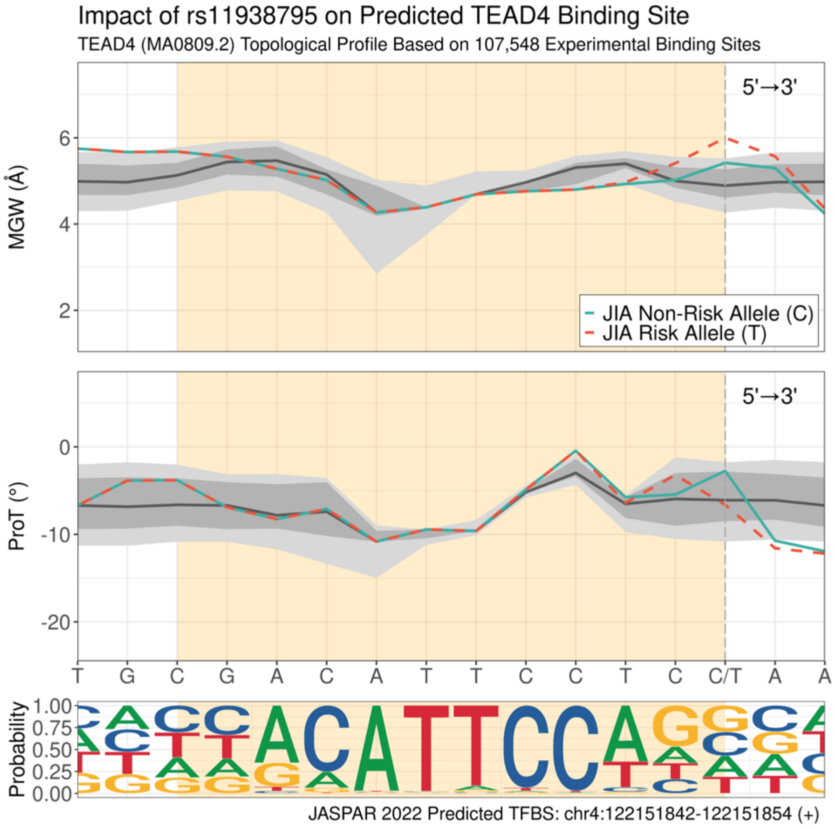

**(ar) Impact of rs75867630 on predicted ZEB1 binding site (positive strand).**

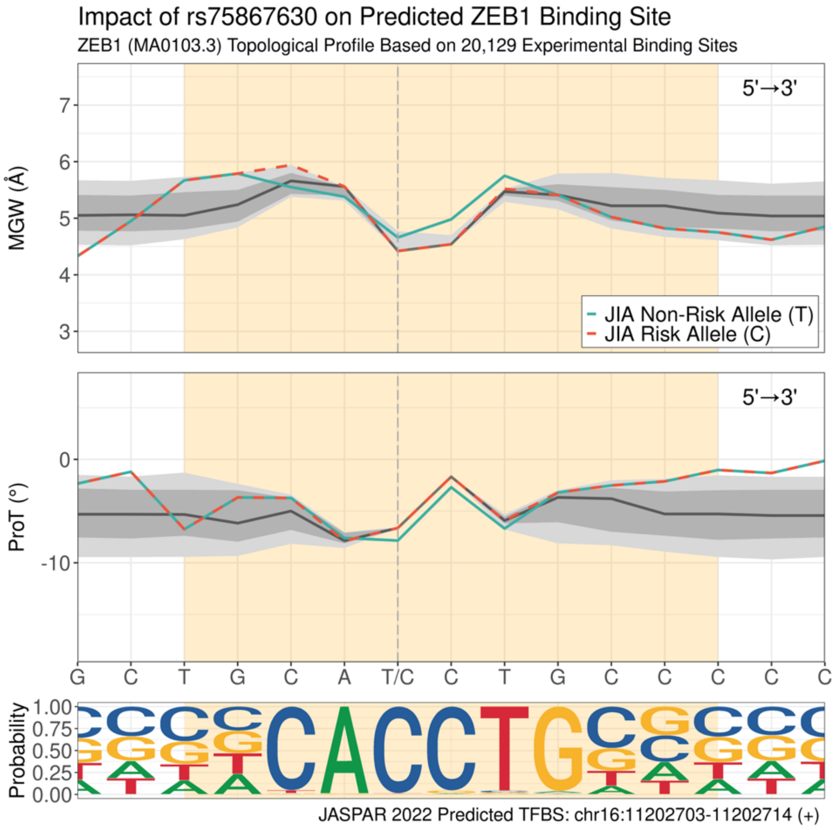

**(as) Impact of rs34437200 on predicted ZNF148 binding site (positive strand).**

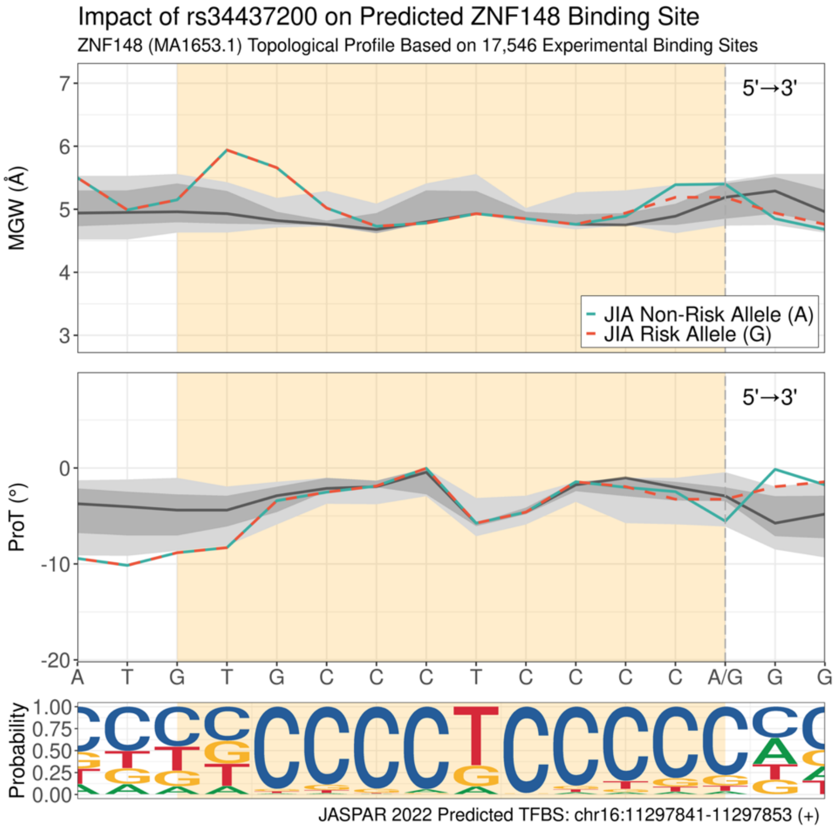

**(at) Impact of rs140489 on predicted ZNF652 binding site (positive strand).**

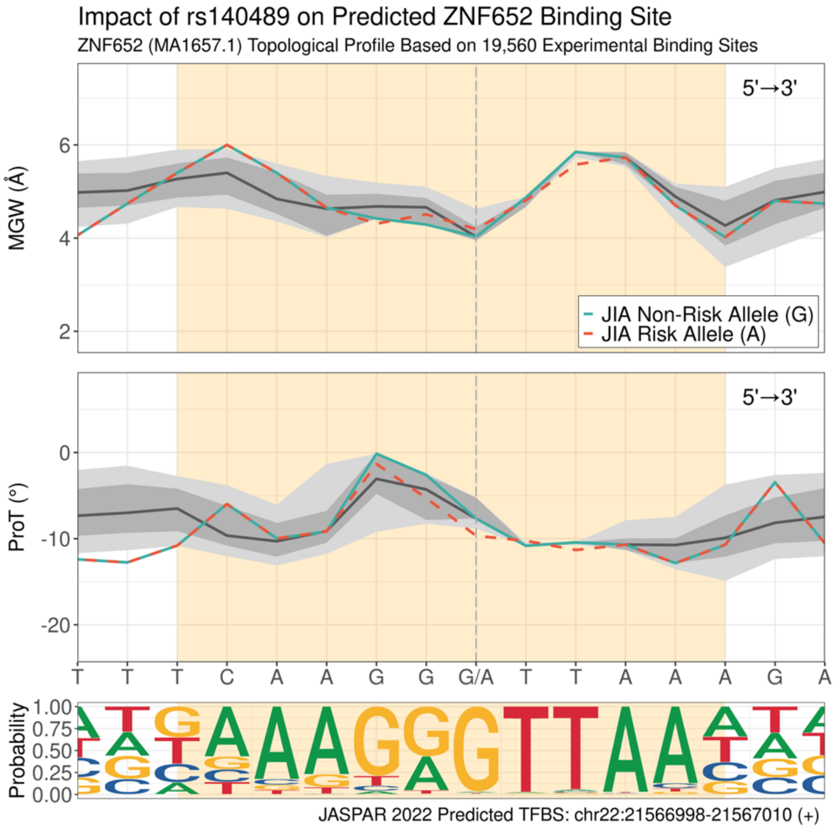

**Supplementary Figure 16: Annotated gel images**

**A.** Binding of TF CREB1 with rs4147359. **B.** Binding of TF GATA1 with rs7234029

A

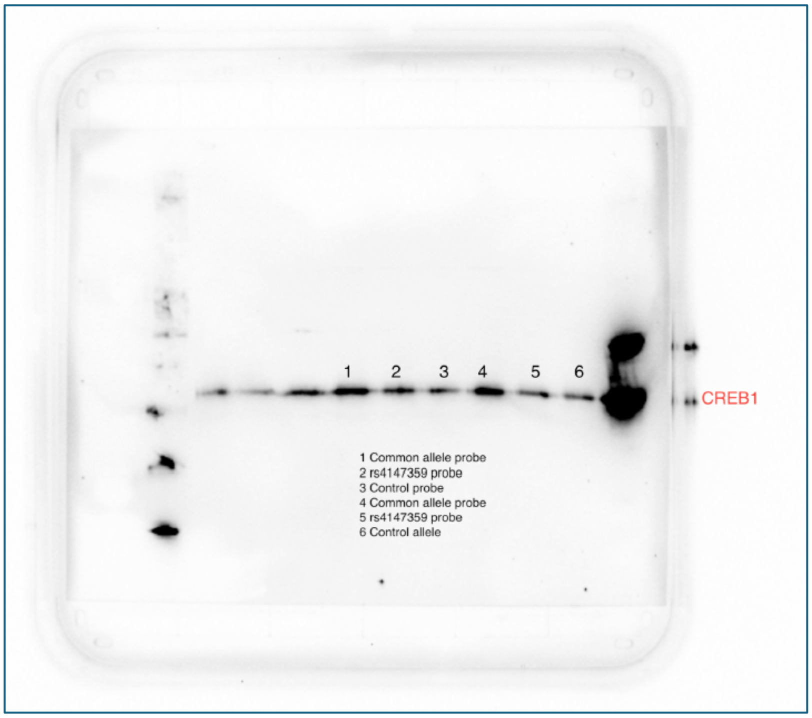

B

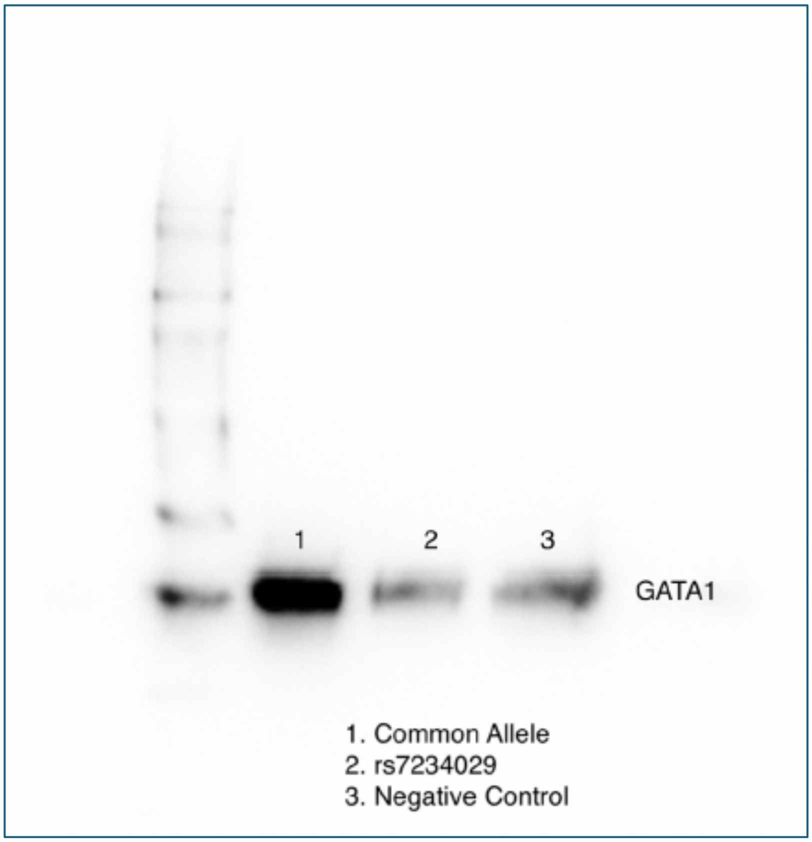

**Supplementary Figure 17: Distribution of autoimmune and rheumatic (A&R) relevance of identified drugs via drug-gene interactions.**

Drugs/small molecules evaluated for relevance to autoimmune and/or rheumatic (A&R) outcomes. Drugs were separately summarized if sourced from CYP* eGenes (n=278 drugs), non-CYP* eGenes (n=243 drugs) or identified by both non-CYP and CYP* eGenes (n= 61). Although CYP* related drugs accounted for the greatest total, A&R relevance was greatest among drugs identified by non-CYP eGenes (29% vs 11%).

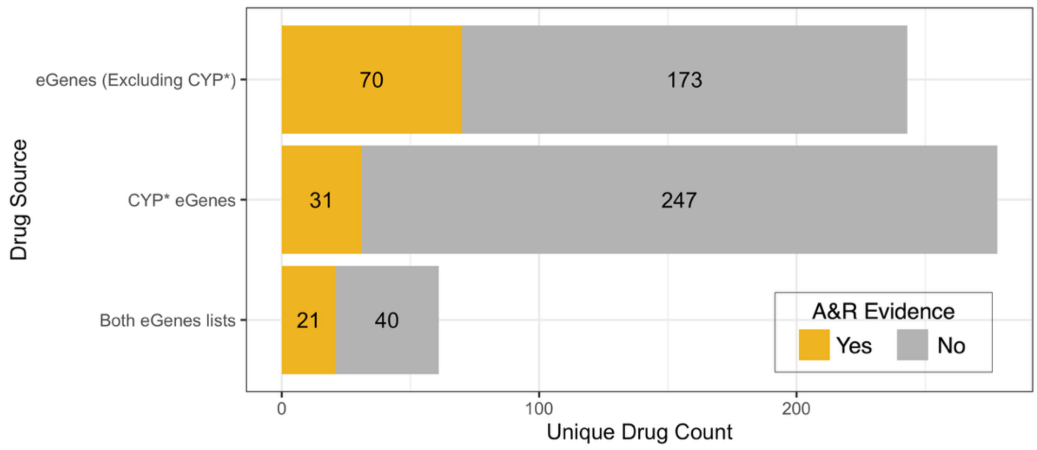

**Supplementary Figure 18: Protein clusters and Reactome annotations from pathway analysis.** Pathway analysis was computed for identified eGenes. Protein-protein interactions were identified through the web database STRING (v12.0), and MCODE clustering and visualization was executed using Cytoscape. Clusters that did not contain more than two proteins were removed. In total, 11 clusters were identified. Color corresponds to odds ratio of minor allele of the eQTL that was most strongly associated with JIA (red: OR<1; blue: OR>1). Size corresponds to strength of the association with JIA (larger size = smaller p-value).

**(a)-(k)** Each cluster is labeled by top responses via enrichment p-values in Reactome (v92).

**(a) MCODE Cluster 1**

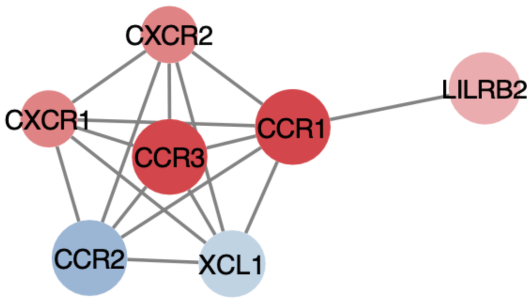

Color corresponds to odds ratio of minor allele of the eQTL that was most strongly associated with JIA (red: OR<1; blue: OR>1). Size corresponds to strength of the association with JIA (larger size = smaller p-value).

| Pathway Name | Genes Found | Genes Found List (Alphabetically) | Entities Found | Entities Total | Entities Ratio | Entities P-Value | Entities FDR |
|---|---|---|---|---|---|---|---|
| Chemokine receptors bind chemokines | 6 | CCR1, CCR2, CCR3, CXCR1, CXCR2, XCL1 | 7 | 57 | 0.004 | $2.55x10^{-15}$ | $5.11x10^{-14}$ |
| Peptide ligand-binding receptors | 6 | CCR1, CCR2, CCR3, CXCR1, CXCR2, XCL1 | 7 | 203 | 0.013 | $1.80x10^{-11}$ | $1.80x10^{-10}$ |
| Class A/1 (Rhodopsin-like receptors) | 6 | CCR1, CCR2, CCR3, CXCR1, CXCR2, XCL1 | 7 | 414 | 0.026 | $2.52x10^{-9}$ | $1.51x10^{-8}$ |
| GPCR downstream signaling | 6 | CCR1, CCR2, CCR3, CXCR1, CXCR2, XCL1 | 8 | 796 | 0.050 | $5.89x10^{-9}$ | $2.95x10^{-8}$ |
| Signaling by GPCR | 6 | CCR1, CCR2, CCR3, CXCR1, CXCR2, XCL1 | 8 | 877 | 0.055 | $1.26x10^{-8}$ | $5.05x10^{-8}$ |
| GPCR ligand binding | 6 | CCR1, CCR2, CCR3, CXCR1, CXCR2, XCL1 | 7 | 609 | 0.038 | $3.60x10^{-8}$ | $1.08x10^{-7}$ |
| G alpha (i) signaling events | 6 | CCR1, CCR2, CCR3, CXCR1, CXCR2, XCL1 | 6 | 428 | 0.027 | $1.61x10^{-7}$ | $3.22x10^{-7}$ |
| Interleukin-10 signaling | 2 | CCR1, CCR2 | 4 | 86 | 0.005 | $2.79x10^{-7}$ | $5.58x10^{-7}$ |
| Signal Transduction | 6 | CCR1, CCR2, CCR3, CXCR1, CXCR2, XCL1 | 8 | 3,049 | 0.193 | $1.79x10^{-4}$ | $3.57x10^{-4}$ |
| Immune System | 5 | CCR1, CCR2, CXCR1, CXCR2, LILRB2 | 7 | 2,664 | 0.168 | $6.70x10^{-4}$ | $7.27x10^{-4}$ |

### (b) MCODE Cluster 2

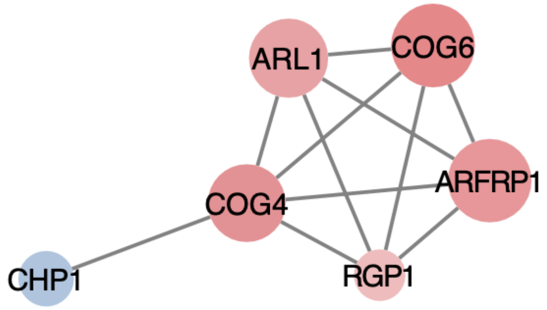

Color corresponds to odds ratio of minor allele of the eQTL that was most strongly associated with JIA (red: OR<1; blue: OR>1). Size corresponds to strength of the association with JIA (larger size = smaller p-value).

| Pathway Name | Genes Found | Genes Found List (Alphabetically) | Entities Found | Entities Total | Entities Ratio | Entities P-Value | Entities FDR |
|---|---|---|---|---|---|---|---|
| Retrograde transport at the Trans-Golgi-Network | 5 | ARFRP1, ARL1, COG4, COG6, RGP1 | 5 | 49 | 0.004 | $2.46 \times 10^{-11}$ | $7.87 \times 10^{-10}$ |
| Intra-Golgi and retrograde Golgi-to-ER traffic | 5 | ARFRP1, ARL1, COG4, COG6, RGP1 | 5 | 206 | 0.017 | $3.16 \times 10^{-8}$ | $5.06 \times 10^{-7}$ |
| Intra-Golgi traffic | 3 | COG4, COG6, RGP1 | 3 | 44 | 0.004 | $1.75 \times 10^{-6}$ | $1.75 \times 10^{-5}$ |
| Membrane Trafficking | 5 | ARFRP1, ARL1, COG4, COG6, RGP1 | 5 | 637 | 0.053 | $8.40 \times 10^{-6}$ | $6.72 \times 10^{-5}$ |
| Vesicle-mediated transport | 5 | ARFRP1, ARL1, COG4, COG6, RGP1 | 5 | 763 | 0.064 | $2.03 \times 10^{-5}$ | $1.22 \times 10^{-4}$ |
| COPI-mediated anterograde transport | 2 | COG4, COG6 | 2 | 102 | 0.009 | $1.50 \times 10^{-3}$ | $5.99 \times 10^{-3}$ |
| ER to Golgi Anterograde Transport | 2 | COG4, COG6 | 2 | 156 | 0.013 | $3.45 \times 10^{-3}$ | $1.04 \times 10^{-2}$ |
| Transport to the Golgi and subsequent modification | 2 | COG4, COG6 | 2 | 187 | 0.016 | $4.91 \times 10^{-3}$ | $1.41 \times 10^{-2}$ |
| Hyaluronan uptake and degradation | 1 | CHP1 | 1 | 12 | 0.001 | $7.03 \times 10^{-3}$ | $1.41 \times 10^{-2}$ |
| Hyaluronan metabolism | 1 | CHP1 | 1 | 17 | 0.001 | $9.95 \times 10^{-3}$ | $1.99 \times 10^{-2}$ |

### (c) MCODE Cluster 3

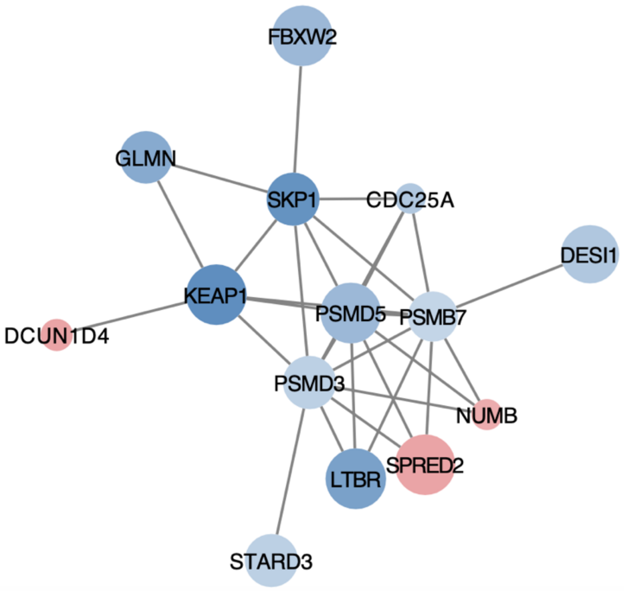

Color corresponds to odds ratio of minor allele of the eQTL that was most strongly associated with JIA (red: OR<1; blue: OR>1). Size corresponds to strength of the association with JIA (larger size = smaller p-value).

| Pathway Name | Genes Found | Genes Found List (Alphabetically) | Entities Found | Entities Total | Entities Ratio | Entities P-Value | Entities FDR |
|---|---|---|---|---|---|---|---|
| Cyclin E associated events during G1/S transition | 4 | CDC25A, PSMB7, PSMD3, SKP1 | 5 | 78 | 0.005 | $1.71x10^{-8}$ | $2.49x10^{-6}$ |
| Cyclin A:Cdk2-associated events at S phase entry | 4 | CDC25A, PSMB7, PSMD3, SKP1 | 5 | 80 | 0.005 | $1.94x10^{-8}$ | $2.49x10^{-6}$ |
| Class I MHC mediated antigen processing & presentation | 6 | FBXW2, GLMN, KEAP1, PSMB7, PSMD3, SKP1 | 7 | 404 | 0.026 | $1.10x10^{-7}$ | $8.55x10^{-6}$ |
| Neddylation | 6 | DCUN1D4, FBXW2, KEAP1, PSMB7, PSMD3, SKP1 | 6 | 241 | 0.015 | $1.34x10^{-7}$ | $8.55x10^{-6}$ |
| Degradation of GLI1 by the proteasome | 4 | NUMB, PSMB7, PSMD3, SKP1 | 4 | 49 | 0.003 | $2.12x10^{-7}$ | $1.00x10^{-5}$ |
| Nuclear events mediated by NFE2L2 | 4 | KEAP1, PSMB7, PSMD3, SKP1 | 5 | 134 | 0.008 | $2.47x10^{-7}$ | $1.00x10^{-5}$ |
| Proteasome assembly | 3 | PSMB7, PSMD3, PSMD5 | 4 | 52 | 0.003 | $2.68x10^{-7}$ | $1.00x10^{-5}$ |
| G1/S Transition | 4 | CDC25A, PSMB7, PSMD3, SKP1 | 5 | 140 | 0.009 | $3.07x10^{-7}$ | $1.00x10^{-5}$ |
| Antigen processing: Ubiquitination & Proteasome degradation | 6 | FBXW2, GLMN, KEAP1, PSMB7, PSMD3, SKP1 | 6 | 302 | 0.019 | $4.99x10^{-7}$ | $1.40x10^{-5}$ |
| Mitotic G1 phase and G1/S transition | 4 | CDC25A, PSMB7, PSMD3, SKP1 | 5 | 164 | 0.010 | $6.67x10^{-7}$ | $1.58x10^{-5}$ |

**(d) MCODE Cluster 4**

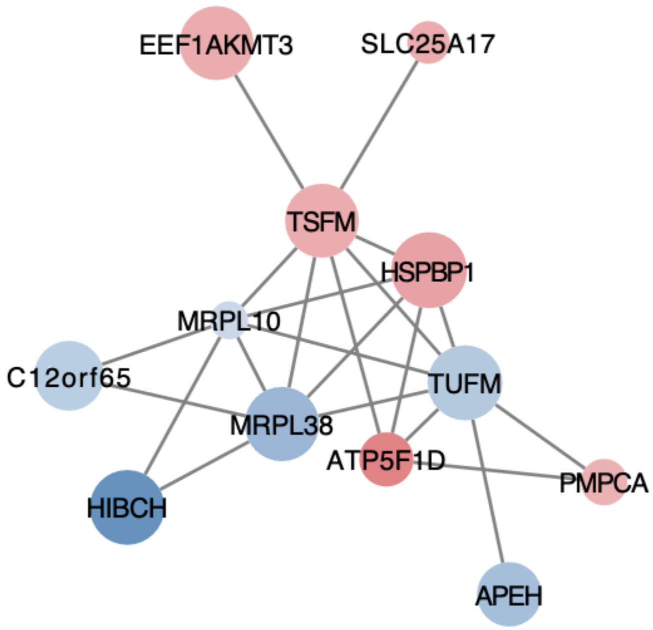

Color corresponds to odds ratio of minor allele of the eQTL that was most strongly associated with JIA (red: OR<1; blue: OR>1). Size corresponds to strength of the association with JIA (larger size = smaller p-value).

| Pathway Name | Genes Found | Genes Found List (Alphabetically) | Entities Found | Entities Total | Entities Ratio | Entities P-Value | Entities FDR |
|---|---|---|---|---|---|---|---|
| Mitochondrial translation elongation | 4 | MRPL10, MRPL38, TSFM, TUFM | 4 | 88 | 0.007 | $1.41 \times 10^{-6}$ | $3.47 \times 10^{-5}$ |
| Mitochondrial translation | 4 | MRPL10, MRPL38, TSFM, TUFM | 4 | 94 | 0.008 | $1.83 \times 10^{-6}$ | $3.47 \times 10^{-5}$ |
| Translation | 5 | APEH, MRPL10, MRPL38, TSFM, TUFM | 5 | 294 | 0.025 | $6.28 \times 10^{-6}$ | $8.17 \times 10^{-5}$ |
| 3-hydroxyisobutyryl-CoA hydrolase deficiency | 1 | HIBCH | 1 | 1 | 0 | $1.01 \times 10^{-3}$ | $9.07 \times 10^{-3}$ |
| Mitochondrial translation termination | 2 | MRPL10, MRPL38 | 2 | 88 | 0.007 | $3.43 \times 10^{-3}$ | $2.06 \times 10^{-2}$ |
| Mitochondrial translation initiation | 2 | MRPL10, MRPL38 | 2 | 88 | 0.007 | $3.43 \times 10^{-3}$ | $2.06 \times 10^{-2}$ |
| Alpha-oxidation of phytanate | 1 | SLC25A17 | 1 | 6 | 0.001 | $6.03 \times 10^{-3}$ | $3.01 \times 10^{-2}$ |
| Metabolism of proteins | 6 | APEH, MRPL10, MRPL38, PMPCA, TSFM, TUFM | 6 | 2,073 | 0.175 | $1.01 \times 10^{-2}$ | $3.91 \times 10^{-2}$ |
| Protein localization | 2 | PMPCA, SLC25A17 | 2 | 166 | 0.014 | $1.17 \times 10^{-2}$ | $3.91 \times 10^{-2}$ |
| Diseases of branched-chain amino acid catabolism | 1 | HIBCH | 1 | 13 | 0.001 | $1.30 \times 10^{-2}$ | $3.91 \times 10^{-2}$ |

### (e) MCODE Cluster 5

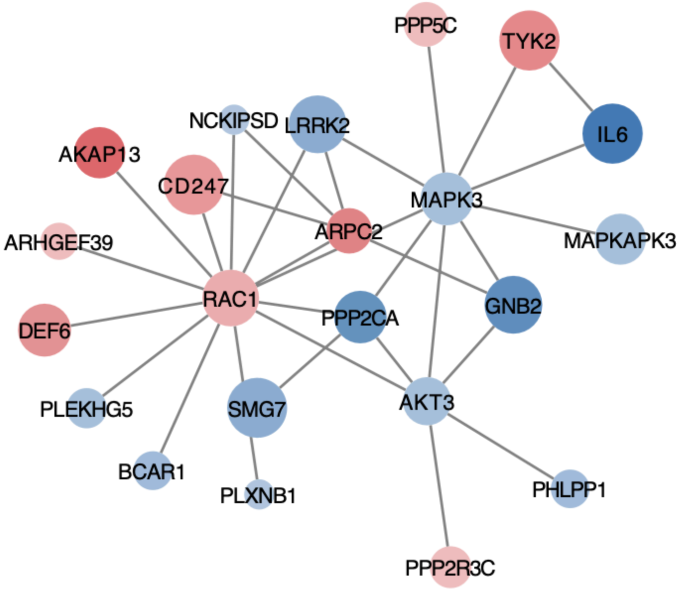

Color corresponds to odds ratio of minor allele of the eQTL that was most strongly associated with JIA (red: OR<1; blue: OR>1). Size corresponds to strength of the association with JIA (larger size = smaller p-value).

| Pathway Name | Genes Found | Genes Found List (Alphabetically) | Entities Found | Entities Total | Entities Ratio | Entities P-Value | Entities FDR |
|---|---|---|---|---|---|---|---|
| Signal Transduction | 19 | AKAP13, AKT3, ARHGEF39, ARPC2, BCAR1, CD247, DEF6, GNB2, IL6, LRRK2, MAPK3, MAPKAPK3, NCKIPSD, PHLPP1, PLEKHG5, PLXNB1, PPP2CA, PPP5C, RAC1, SMG7, TYK2 | 20 | 3,049 | 0.193 | $3.40x10^{-10}$ | $1.72x10^{-7}$ |
| Signaling by Rho GTPases | 10 | AKAP13, ARHGEF39, ARPC2, DEF6, MAPK3, NCKIPSD, PLEKHG5, PLXNB1, PPP2CA, RAC1 | 11 | 708 | 0.045 | $5.93x10^{-9}$ | $1.26x10^{-6}$ |
| Signaling by Rho GTPases, Miro GTPases and RHOBTB3 | 10 | AKAP13, ARHGEF39, ARPC2, DEF6, MAPK3, NCKIPSD, PLEKHG5, PLXNB1, PPP2CA, RAC1 | 11 | 724 | 0.046 | $7.48x10^{-9}$ | $1.26x10^{-6}$ |
| Parasitic Infection Pathways | 7 | ARPC2, CD247, GNB2, IL6, MAPK3, NCKIPSD, RAC1 | 8 | 301 | 0.019 | $1.97x10^{-9}$ | $1.99x10^{-6}$ |
| Leishmania infection | 7 | ARPC2, CD247, GNB2, IL6, MAPK3, NCKIPSD, RAC1 | 8 | 301 | 0.019 | $1.97x10^{-9}$ | $1.99x10^{-6}$ |
| G alpha (12/13) signaling events | 5 | AKAP13, ARHGEF39, GNB2, PLEKHG5, PLXNB1 | 5 | 86 | 0.005 | $2.83x10^{-7}$ | $2.38x10^{-5}$ |
| RHO GTPases Activate WASPs and WAVEs | 4 | ARPC2, MAPK3, NCKIPSD, RAC1 | 4 | 41 | 0.003 | $6.44x10^{-7}$ | $4.63x10^{-5}$ |
| MAPK3 (ERK1) activation | 3 | IL6, MAPK3, TYK2 | 3 | 13 | 0.001 | $1.42x10^{-6}$ | $9.00x10^{-5}$ |
| Negative regulation of the PI3K/AKT network | 5 | AKT3, MAPK3, PHLPP1, PPP2CA, RAC1 | 5 | 145 | 0.009 | $3.62x10^{-6}$ | $1.86x10^{-4}$ |
| NRAGE signals death through JNK | 4 | AKAP13, ARHGEF39, PLEKHG5, RAC1 | 4 | 64 | 0.004 | $3.72x10^{-6}$ | $1.86x10^{-4}$ |

### (g) MCODE Cluster 6

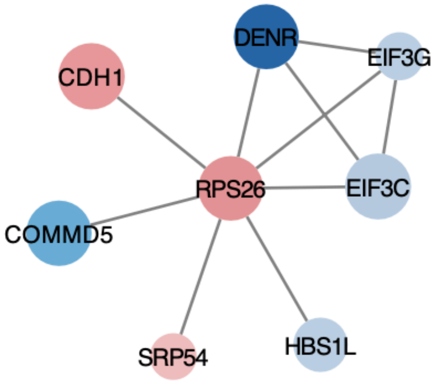

Color corresponds to odds ratio of minor allele of the eQTL that was most strongly associated with JIA (red: OR<1; blue: OR>1). Size corresponds to strength of the association with JIA (larger size = smaller p-value).

| Pathway Name | Genes Found | Genes Found List (Alphabetically) | Entities Found | Entities Total | Entities Ratio | Entities P-Value | Entities FDR |
|---|---|---|---|---|---|---|---|
| Formation of the ternary complex, and subsequently, the 43S complex | 3 | EIF3C, EIF3G, RPS26 | 3 | 54 | 0.003 | $6.42 \times 10^{-6}$ | $3.62 \times 10^{-4}$ |
| Translation initiation complex formation | 3 | EIF3C, EIF3G, RPS26 | 3 | 62 | 0.004 | $1.00 \times 10^{-5}$ | $3.62 \times 10^{-4}$ |
| Ribosomal scanning and start codon recognition | 3 | EIF3C, EIF3G, RPS26 | 3 | 64 | 0.004 | $1.07 \times 10^{-5}$ | $3.62 \times 10^{-4}$ |
| Activation of the mRNA upon binding of the cap-binding complex and eIFs, and subsequent binding to 43S | 3 | EIF3C, EIF3G, RPS26 | 3 | 66 | 0.004 | $1.17 \times 10^{-5}$ | $3.62 \times 10^{-4}$ |
| Formation of a pool of free 40S subunits | 3 | EIF3C, EIF3G, RPS26 | 3 | 106 | 0.007 | $4.76 \times 10^{-5}$ | $1.04 \times 10^{-3}$ |
| GTP hydrolysis and joining of the 60S ribosomal subunit | 3 | EIF3C, EIF3G, RPS26 | 3 | 120 | 0.008 | $6.87 \times 10^{-5}$ | $1.04 \times 10^{-3}$ |
| L13a-mediated translational silencing of Ceruloplasmin expression | 3 | EIF3C, EIF3G, RPS26 | 3 | 120 | 0.008 | $6.87 \times 10^{-5}$ | $1.04 \times 10^{-3}$ |
| Translation | 4 | EIF3C, EIF3G, RPS26, SRP54 | 4 | 352 | 0.022 | $7.13 \times 10^{-5}$ | $1.04 \times 10^{-3}$ |
| Cap-dependent Translation Initiation | 3 | EIF3C, EIF3G, RPS26 | 3 | 130 | 0.008 | $8.71 \times 10^{-5}$ | $1.04 \times 10^{-3}$ |
| Eukaryotic Translation Initiation | 3 | EIF3C, EIF3G, RPS26 | 3 | 130 | 0.008 | $8.71 \times 10^{-5}$ | $1.04 \times 10^{-3}$ |

**(e) MCODE Cluster 7**

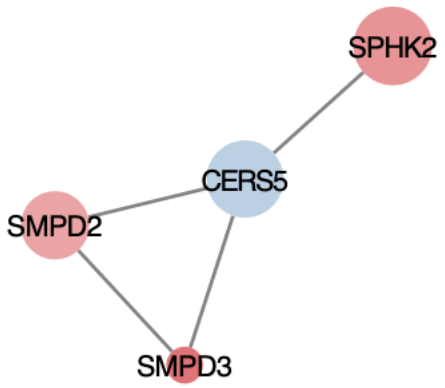

Color corresponds to odds ratio of minor allele of the eQTL that was most strongly associated with JIA (red: OR<1; blue: OR>1). Size corresponds to strength of the association with JIA (larger size = smaller p-value).

| Pathway Name | Genes Found | Genes Found List (Alphabetically) | Entities Found | Entities Total | Entities Ratio | Entities P-Value | Entities FDR |
|---|---|---|---|---|---|---|---|
| Sphingolipid metabolism | 4 | CERS5, SMPD2, SMPD3, SPHK2 | 4 | 107 | 0.009 | $6.52x10^{-9}$ | $7.82x10^{-8}$ |
| TNFR1-mediated ceramide production | 2 | SMPD2, SMPD3 | 2 | 6 | 0.001 | $1.52x10^{-6}$ | $1.00x10^{-5}$ |
| Metabolism of lipids | 4 | CERS5, SMPD2, SMPD3, SPHK2 | 4 | 765 | 0.064 | $1.70x10^{-5}$ | $6.81x10^{-5}$ |
| Sphingolipid de novo biosynthesis | 2 | CERS5, SPHK2 | 2 | 37 | 0.003 | $5.77x10^{-5}$ | $1.28x10^{-4}$ |
| Glycosphingolipid catabolism | 2 | SMPD2, SMPD3 | 2 | 39 | 0.003 | $6.41x10^{-5}$ | $1.28x10^{-4}$ |
| Glycosphingolipid metabolism | 2 | SMPD2, SMPD3 | 2 | 58 | 0.005 | $1.41x10^{-4}$ | $1.56x10^{-4}$ |
| TNF signaling | 2 | SMPD2, SMPD3 | 2 | 61 | 0.005 | $1.56x10^{-4}$ | $1.56x10^{-4}$ |
| Ceramide signalling | 1 | SMPD2 | 1 | 3 | 0 | $1.01x10^{-3}$ | $1.01x10^{-3}$ |
| Death Receptor Signaling | 2 | SMPD2, SMPD3 | 2 | 161 | 0.014 | $1.08x10^{-3}$ | $1.08x10^{-3}$ |
| Metabolism | 4 | CERS5, SMPD2, SMPD3, SPHK2 | 4 | 2,202 | 0.185 | $1.17x10^{-3}$ | $1.17x10^{-3}$ |

### (h) MCODE Cluster 8

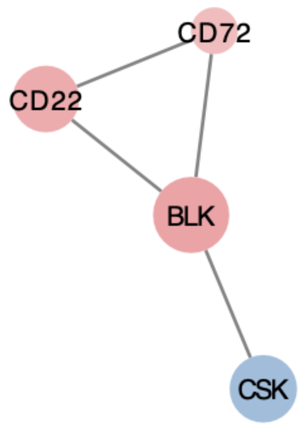

Color corresponds to odds ratio of minor allele of the eQTL that was most strongly associated with JIA (red: OR<1; blue: OR>1). Size corresponds to strength of the association with JIA (larger size = smaller p-value).

| Pathway Name | Genes Found | Genes Found List (Alphabetically) | Entities Found | Entities Total | Entities Ratio | Entities P-Value | Entities FDR |
|---|---|---|---|---|---|---|---|
| RUNX1 regulates transcription of genes involved in BCR signaling | 1 | BLK | 2 | 7 | 0 | $1.95x10^{-6}$ | $9.77x10^{-5}$ |
| Antigen activates B Cell Receptor (BCR) leading to generation of second messengers | 2 | BLK, CD22 | 2 | 103 | 0.007 | $4.18x10^{-4}$ | $1.05x10^{-2}$ |
| Signaling by the B Cell Receptor (BCR) | 2 | BLK, CD22 | 2 | 176 | 0.011 | $1.21x10^{-3}$ | $1.94x10^{-2}$ |
| Adaptive Immune System | 3 | BLK, CD22, CSK | 3 | 928 | 0.059 | $1.84x10^{-3}$ | $2.21x10^{-2}$ |
| Transcriptional regulation by RUNX1 | 1 | BLK | 2 | 246 | 0.016 | $2.34x10^{-3}$ | $2.29x10^{-2}$ |
| Other semaphorin interactions | 1 | CD72 | 1 | 19 | 0.001 | $5.99x10^{-3}$ | $2.29x10^{-2}$ |
| Negative regulation of FLT3 | 1 | CSK | 1 | 20 | 0.001 | $6.30x10^{-3}$ | $2.29x10^{-2}$ |
| GAB1 signalosome | 1 | CSK | 1 | 23 | 0.001 | $7.25x10^{-3}$ | $2.29x10^{-2}$ |
| Co-inhibition by PD-1 | 1 | CSK | 1 | 33 | 0.002 | $1.04x10^{-2}$ | $2.29x10^{-2}$ |
| Phosphorylation of CD3 and TCR zeta chains | 1 | CSK | 1 | 33 | 0.002 | $1.04x10^{-2}$ | $2.29x10^{-2}$ |

### (i) MCODE Cluster 9

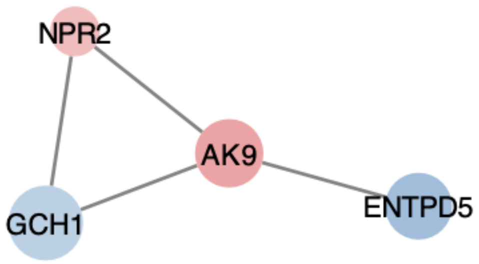

Color corresponds to odds ratio of minor allele of the eQTL that was most strongly associated with JIA (red: OR<1; blue: OR>1). Size corresponds to strength of the association with JIA (larger size = smaller p-value).

| Pathway Name | Genes Found | Genes Found List (Alphabetically) | Entities Found | Entities Total | Entities Ratio | Entities P-Value | Entities FDR |
|---|---|---|---|---|---|---|---|
| Metabolism of nucleotides | 2 | AK9, ENTPD5 | 2 | 101 | 0.008 | $4.27x10^{-4}$ | $7.26x10^{-3}$ |
| Phosphate bond hydrolysis by NTPDase proteins | 1 | ENTPD5 | 1 | 8 | 0.001 | $2.68x10^{-3}$ | $1.27x10^{-2}$ |
| Tetrahydrobiopterin (BH4) synthesis, recycling, salvage and regulation | 1 | GCH1 | 1 | 10 | 0.001 | $3.35x10^{-3}$ | $1.27x10^{-2}$ |
| Physiological factors | 1 | NPR2 | 1 | 14 | 0.001 | $4.69x10^{-3}$ | $1.27x10^{-2}$ |
| Metabolism of cofactors | 1 | GCH1 | 1 | 25 | 0.002 | $8.37x10^{-3}$ | $1.27x10^{-2}$ |
| Cell recruitment (pro-inflammatory response) | 1 | ENTPD5 | 1 | 27 | 0.002 | $9.04x10^{-3}$ | $1.27x10^{-2}$ |
| Purinergic signaling in leishmaniasis infection | 1 | ENTPD5 | 1 | 27 | 0.002 | $9.04x10^{-3}$ | $1.27x10^{-2}$ |
| Interconversion of nucleotide di- and triphosphates | 1 | AK9 | 1 | 30 | 0.003 | $1.00x10^{-2}$ | $1.27x10^{-2}$ |
| Nucleotide catabolism | 1 | ENTPD5 | 1 | 38 | 0.003 | $1.27x10^{-2}$ | $1.27x10^{-2}$ |
| Metabolism | 3 | AK9, ENTPD5, GCH1 | 3 | 2,202 | 0.185 | $2.18x10^{-2}$ | $2.18x10^{-2}$ |

**(j) MCODE Cluster 10**

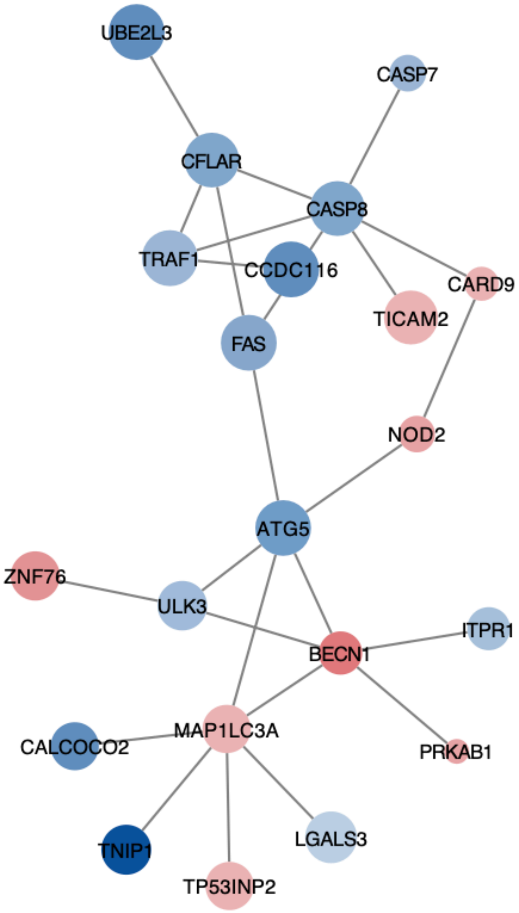

Color corresponds to odds ratio of minor allele of the eQTL that was most strongly associated with JIA (red: OR<1; blue: OR>1). Size corresponds to strength of the association with JIA (larger size = smaller p-value).

| Pathway Name | Genes Found | Genes Found List (Alphabetically) | Entities Found | Entities Total | Entities Ratio | Entities P-Value | Entities FDR |
|---|---|---|---|---|---|---|---|
| Caspase activation via Death Receptors in the presence of ligand | 4 | CASP8, CFLAR, FAS, TICAM2 | 5 | 20 | 0.001 | $2.54x10^{-10}$ | $6.33x10^{-8}$ |
| Caspase activation via extrinsic apoptotic signalling pathway | 4 | CASP8, CFLAR, FAS, TICAM2 | 5 | 32 | 0.002 | $2.63x10^{-9}$ | $3.36x10^{-7}$ |
| CASP8 activity is inhibited | 3 | CFLAR, FAS, TICAM2 | 4 | 12 | 0.001 | $5.72x10^{-9}$ | $4.35x10^{-7}$ |
| Regulation of necroptotic cell death | 4 | CASP8, CFLAR, FAS, UBE2L3 | 5 | 39 | 0.002 | $7.01x10^{-9}$ | $4.35 x10^{-7}$ |
| RIPK1-mediated regulated necrosis | 4 | CASP8, CFLAR, FAS, UBE2L3 | 5 | 45 | 0.003 | $1.42x10^{-8}$ | $6.98x10^{-7}$ |
| Programmed Cell Death | 6 | CASP7, CASP8, CFLAR, FAS, TICAM2, UBE2L3 | 7 | 225 | 0.014 | $8.12x10^{-8}$ | $3.33x10^{-6}$ |
| Regulated Necrosis | 4 | CASP8, CFLAR, FAS, UBE2L3 | 5 | 76 | 0.005 | $1.89x10^{-7}$ | $6.61x10^{-6}$ |
| Death Receptor Signaling | 5 | CASP8, CFLAR, FAS, TRAF1, UBE2L3 | 6 | 172 | 0.011 | $4.01x10^{-7}$ | $1.24x10^{-5}$ |
| Apoptosis | 5 | CASP7, CASP8, CFLAR, FAS, TICAM2 | 6 | 180 | 0.011 | $5.22x10^{-7}$ | $1.41x10^{-5}$ |
| Regulation by c-FLIP | 3 | CASP8, CFLAR, FAS, | 3 | 11 | 0.001 | $9.70x10^{-7}$ | $2.13x10^{-5}$ |

### (k) MCODE Cluster 11

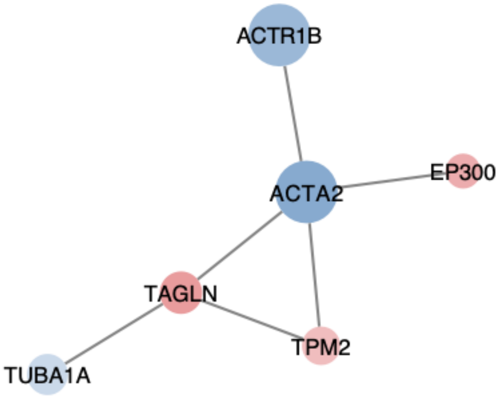

Color corresponds to odds ratio of minor allele of the eQTL that was most strongly associated with JIA (red: OR<1; blue: OR>1). Size corresponds to strength of the association with JIA (larger size = smaller p-value).

| Pathway Name | Genes Found | Genes Found List (Alphabetically) | Entities Found | Entities Total | Entities Ratio | Entities P-Value | Entities FDR |
|---|---|---|---|---|---|---|---|
| NOTCH4 Intracellular Domain Regulates Transcription | 2 | ACTA2, EP300 | 3 | 26 | 0.002 | $1.54 \times 10^{-7}$ | $3.55 \times 10^{-5}$ |
| Signaling by NOTCH4 | 2 | ACTA2, EP300 | 3 | 82 | 0.005 | $4.79 \times 10^{-6}$ | $5.51 \times 10^{-4}$ |
| Signaling by NOTCH | 2 | ACTA2, EP300 | 3 | 260 | 0.016 | $1.48 \times 10^{-4}$ | $1.12 \times 10^{-2}$ |
| Smooth Muscle Contraction | 2 | ACTA2, TPM2 | 2 | 61 | 0.004 | $3.08 \times 10^{-4}$ | $1.76 \times 10^{-2}$ |
| MHC class II antigen presentation | 2 | ACTR1B, TUBA1A | 2 | 137 | 0.009 | $1.53 \times 10^{-3}$ | $4.17 \times 10^{-2}$ |
| NFE2L2 regulating ER-stress associated genes | 1 | EP300 | 1 | 6 | 0 | $2.65 \times 10^{-3}$ | $4.17 \times 10^{-2}$ |
| NFE2L2 regulating inflammation associated genes | 1 | EP300 | 1 | 7 | 0 | $3.09 \times 10^{-3}$ | $4.17 \times 10^{-2}$ |
| LRR FLII-interacting protein 1 (LRRFIP1) activates type I IFN production | 1 | EP300 | 1 | 7 | 0 | $3.09 \times 10^{-3}$ | $4.17 \times 10^{-2}$ |
| G2/M Transition | 2 | EP300, TUBA1A | 2 | 205 | 0.013 | $3.37 \times 10^{-3}$ | $4.17 \times 10^{-2}$ |
| Mitotic G2-G2/M phases | 2 | EP300, TUBA1A | 2 | 207 | 0.013 | $3.44 \times 10^{-3}$ | $4.17 \times 10^{-2}$ |
